# Connectivity within the Hippocampus as a Neural Marker of Early Clinical Trajectories in the Psychosis Risk State

**DOI:** 10.64898/2026.03.10.26348090

**Authors:** Lukas Roell, Christoph Lindner, Ye Ella Tian, Sidhant Chopra, Isabel Maurus, Joanna Moussiopoulou, Vladislav Yakimov, Maxim Korman, Daniel Keeser, Andrea Schmitt, Peter Falkai, Maria A. Di Biase, Steffen Zitzmann, Andrew Zalesky

## Abstract

Psychotic disorders lack treatment-informative biomarkers, especially during the earliest illness stages when interventions are most effective. Integrating etiological theories on hippocampal pathology and whole-brain neural dysconnectivity, we studied connectivity changes within the hippocampus as a neuroimaging marker of emerging symptomatic and functional trajectories in the psychosis risk state. We analyzed multicenter longitudinal clinical and functional neuroimaging data across an eight-month period from 434 participants (356 individuals at clinical high risk for psychosis and 78 healthy controls) using latent variable regressions. Decreases of intra-hippocampal connectivity over time tracked worsening negative symptoms, depressive symptoms, and psychosocial functioning in at-risk subjects. This finding was not observed for attenuated positive symptoms and cognition, was specific to high-risk individuals relative to healthy controls, and was not obtained for connectivity within other brain areas. Unveiling the temporal sequence of these associations, we found that an early decrease in connectivity within the hippocampus preceded a subsequent worsening of negative symptoms, but not vice versa. These findings position intra-hippocampal connectivity changes as a neuroimaging marker of early affective-motivational and functional trajectories in the psychosis risk state. They further indicate that changes of connectivity within the hippocampus hold prognostic value specifically for emerging negative symptoms. This informs future risk stratification approaches and neurostimulation therapies in the psychosis risk state: Intra-hippocampal connectivity decline could be a valuable predictive marker to improve risk stratification. Ameliorating connectivity reduction within the hippocampus may represent a promising neurostimulation target to prevent unfavorable clinical trajectories.

**One Sentence Summary:** Decreasing connectivity within the hippocampus is a neural prognostic marker of worsening negative symptoms in the psychosis risk state

## INTRODUCTION

Psychosis has a typical onset in late adolescence or early adulthood and is often characterized by a prodromal phase *(1, 2)*. The concept of clinical high risk for psychosis (CHR-P) facilitates the early identification of individuals with an increased risk to develop psychosis, some of whom may already be experiencing a manifest prodromal phase *(3)*. CHR-P individuals typically show attenuated positive symptoms accompanied by negative and depressive symptoms, cognitive impairment, and reduced psychosocial functioning *(4–7)*. Up to 35% of CHR-P individuals transition to a psychotic disorder over a ten-year period *(8)*. Among non-converters, only 50% fully remit from subclinical symptoms *(9)*. To resolve this heterogeneity in clinical outcomes, a better neurobiological understanding of early emerging clinical changes during the psychosis risk state is warranted.

In this regard, converging evidence implicates hippocampal dysfunction as a key pathophysiological mechanism and early driver of psychotic illness *(10–12)*. Empirical MRI studies in CHR-P individuals demonstrate hippocampal hyperactivity *(13–15)*, particularly in those with poor functional outcomes *(14, 16)*. Beyond regional hyperactivity, altered hippocampal connectivity has also been found in the psychosis risk state *(17–19)*. Reported findings include reduced connectivity between the hippocampus and inferior frontal cortex *(17)*, altered coupling between the CA1 subfield and the ventromedial prefrontal cortex, nucleus accumbens, and amygdala *(18)*, as well as associations between hippocampus-to-whole-brain connectivity and attenuated positive symptoms *(19)*. However, these cross-sectional findings predominantly focus on inter-regional hippocampal connectivity and provide limited insight into early longitudinal changes of connectivity within the hippocampus itself.

Intra-hippocampal connectivity may be particularly informative in light of two influential pathophysiological theories of psychosis. First, models of hippocampal dysfunction propose that early neural pathology in psychosis is initially confined to the hippocampus and only later expands to distributed brain networks as the illness progresses *(12)*. In support of this view, the hippocampus has been consistently identified as a focal epicenter of gray matter volume loss across multiple stages of psychotic disorders *(20)*. Second, psychosis has been conceptualized as a disorder of multi-level neural dysconnectivity *(21)*, a phenomenon robustly observed using functional MRI *(22)*. Importantly, the hippocampus is proposed to significantly influence such broader dysconnectivity patterns in psychosis *(10)*. Both the hippocampal pathology model and the disconnection hypothesis jointly suggest that intra-hippocampal connectivity may constitute a promising neural marker of emerging clinical trajectories at an early illness stage.

Several approaches exist to estimate connectivity within the hippocampus. Microstructural studies subdivide the hippocampus along mediolateral or ventrodorsal axes into subfields such as CA1-CA4 and the dentate gyrus between which connectivity could be computed *(23)*. Anatomical and electrophysiological recordings in rodents and functional neuroimaging studies in humans instead suggest an organization of the hippocampus along its anterior-posterior axis *(23)*. This pattern has been confirmed across development from childhood to early adulthood *(24)*. The anterior-posterior hippocampal axis is assumed to be related to emotional, motivational, and self-referential systems, while the mediolateral and ventrodorsal axes contribute to a more complex multi-demand system *(23)*. To quantify intra-hippocampal connectivity in this study, we rely on functional subunits along the anterior-to-posterior axis of the hippocampus, given that emotional and motivational processes are key in early clinical trajectories in CHR-P individuals. We additionally consider interhemispheric connections between both hippocampi due to the important role of homotopic connectivity in general psychopathology *(25)*.

Using large-scale data from the multicenter North American Prodrome Longitudinal Study (NAPLS-3) *(26)*, we investigated connectivity within the hippocampus as a neural marker of emerging symptomatic and functional trajectories across an eight-month period in the psychosis risk state. We first studied the bidirectional associations between change in intra-hippocampal connectivity and change in attenuated positive symptoms, negative symptoms, depressive symptoms, cognition, and psychosocial functioning. Then, we probed the temporal sequence of these relationships in prospective association analyses, linking early intra-hippocampal connectivity change to subsequent clinical change and vice versa. Our work enhances the current understanding of hippocampal pathology in the psychosis risk state and provides an essential step toward early risk stratification and targeted neurostimulation treatments.

## RESULTS

Figure 1 provides an overview of the methodological approach of this study, illustrating the study design (A), strategy to compute intra-hippocampal connectivity (B), and both statistical analyses (C and D). The bidirectional association analysis (Figure 1C) between change in intra-hippocampal connectivity and clinical change was conducted in 434 subjects (356 CHR-P, 78 healthy controls). The prospective association analysis (Figure 1D) included 257 CHR-P subjects. Both were performed using latent variable regressions. The reduced sample size in the prospective analysis was due to the fact that 99 CHR-P subjects who had at least two clinical and imaging sessions (and were therefore included in the first analysis) did not have a data structure that allowed estimation of separate early and subsequent slopes for connectivity and clinical change. For example, if clinical and imaging data were available only at the first and second time points, a bidirectional association between the two change scores could be computed, but a prospective association could not.

**Figure 1.**
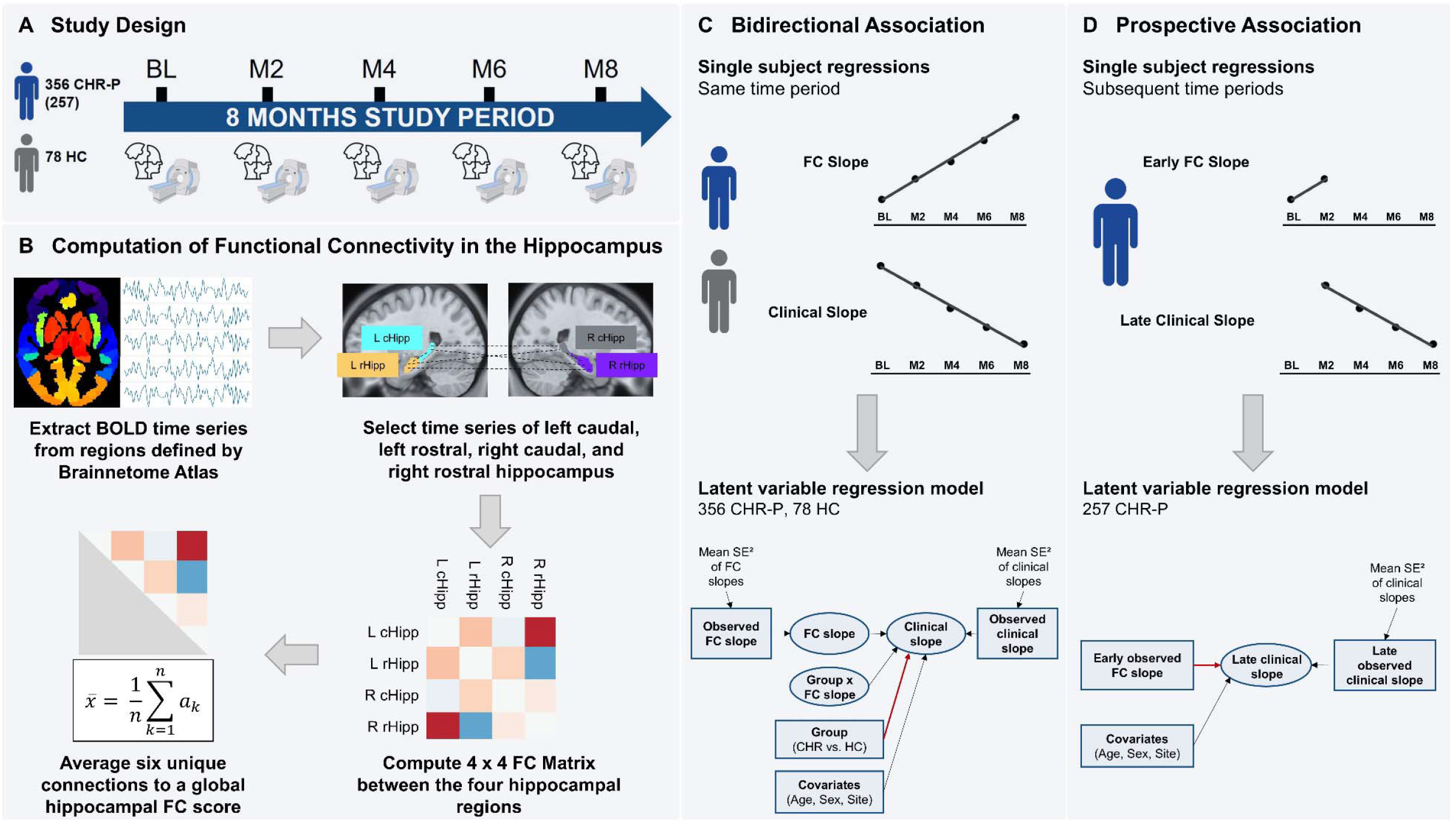
Study overview. A) Illustration of the initial phase of the NAPLS-3 study. B) Workflow to derive connectivity within the hippocampus after pre- and postprocessing of resting-state fMRI data. C) Bidirectional association analysis. The red bold arrow indicates the effect of interest. D) Prospective association analysis. The red bold arrow indicates the effect of interest. FC, functional connectivity; L rHipp, left rostral hippocampus; R rHipp, right rostral hippocampus; L cHipp, left caudal hippocampus; R cHipp, right caudal hippocampus; SE, standard error.

### Intra-hippocampal connectivity is reduced in CHR-P individuals

At baseline, there were no significant differences between individuals at CHR-P and healthy controls in demographic characteristics. However, the CHR-P group showed significantly higher antipsychotic medication use, greater symptom severity, lower psychosocial functioning, and lower symbol coding performance compared to controls (Table 1). CHR-P subjects also had significantly lower baseline connectivity within the hippocampus compared to controls (b = −0.03, p = 0.039; Figure S2).

**Table 1.**
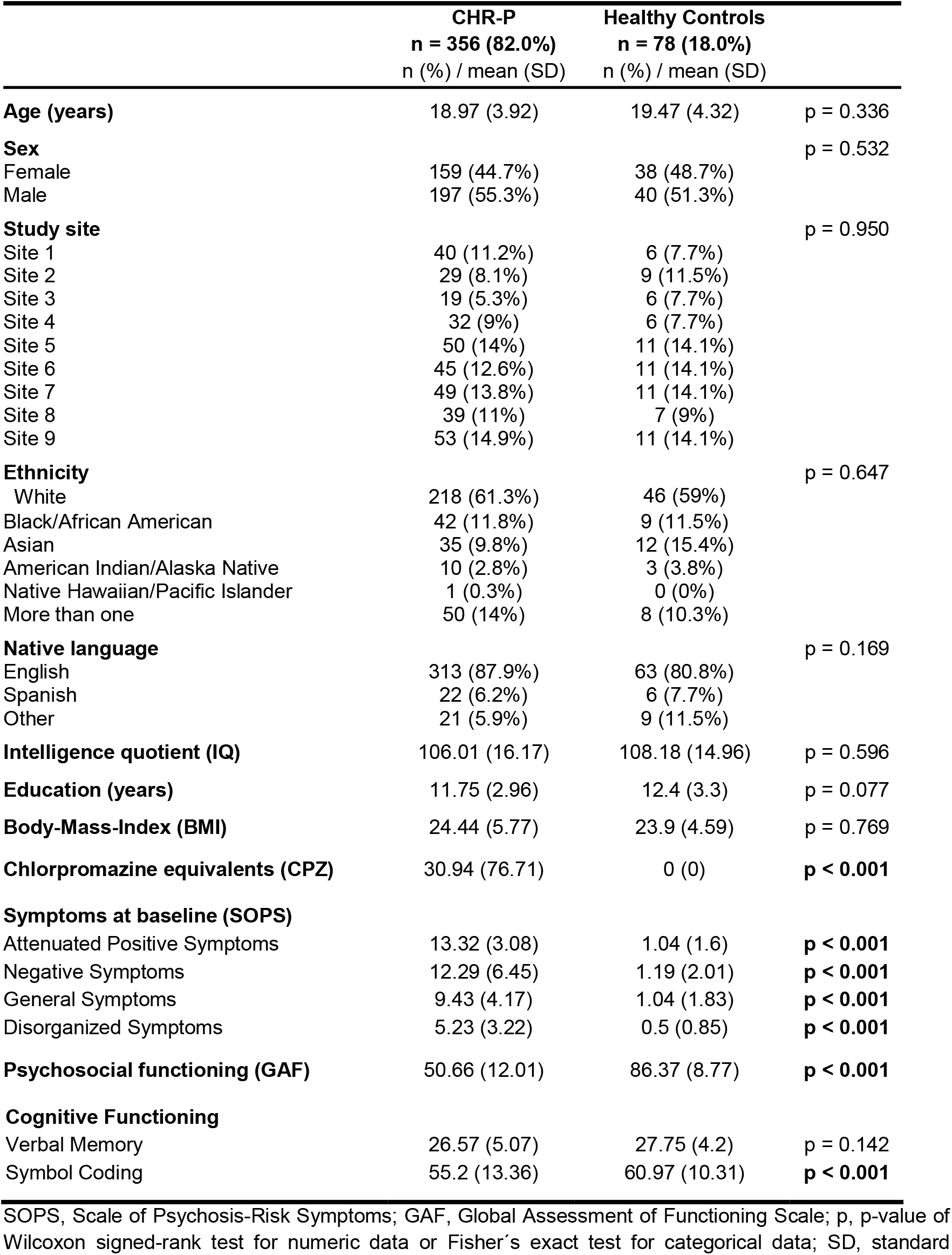
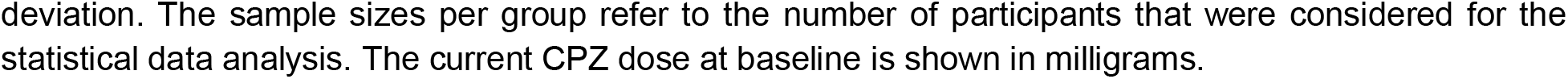
Sample characteristics.

|  | **CHR-P**<br>**n = 356 (82.0%)**<br>n (%) / mean (SD) | **Healthy Controls**<br>**n = 78 (18.0%)**<br>n (%) / mean (SD) |  |
| --- | --- | --- | --- |
| **Age (years)** | 18.97 (3.92) | 19.47 (4.32) | p = 0.336 |
| **Sex** |  |  | p = 0.532 |
| Female | 159 (44.7%) | 38 (48.7%) |  |
| Male | 197 (55.3%) | 40 (51.3%) |  |
| **Study site** |  |  | p = 0.950 |
| Site 1 | 40 (11.2%) | 6 (7.7%) |  |
| Site 2 | 29 (8.1%) | 9 (11.5%) |  |
| Site 3 | 19 (5.3%) | 6 (7.7%) |  |
| Site 4 | 32 (9%) | 6 (7.7%) |  |
| Site 5 | 50 (14%) | 11 (14.1%) |  |
| Site 6 | 45 (12.6%) | 11 (14.1%) |  |
| Site 7 | 49 (13.8%) | 11 (14.1%) |  |
| Site 8 | 39 (11%) | 7 (9%) |  |
| Site 9 | 53 (14.9%) | 11 (14.1%) |  |
| **Ethnicity** |  |  | p = 0.647 |
| White | 218 (61.3%) | 46 (59%) |  |
| Black/African American | 42 (11.8%) | 9 (11.5%) |  |
| Asian | 35 (9.8%) | 12 (15.4%) |  |
| American Indian/Alaska Native | 10 (2.8%) | 3 (3.8%) |  |
| Native Hawaiian/Pacific Islander | 1 (0.3%) | 0 (0%) |  |
| More than one | 50 (14%) | 8 (10.3%) |  |
| **Native language** |  |  | p = 0.169 |
| English | 313 (87.9%) | 63 (80.8%) |  |
| Spanish | 22 (6.2%) | 6 (7.7%) |  |
| Other | 21 (5.9%) | 9 (11.5%) |  |
| **Intelligence quotient (IQ)** | 106.01 (16.17) | 108.18 (14.96) | p = 0.596 |
| **Education (years)** | 11.75 (2.96) | 12.4 (3.3) | p = 0.077 |
| **Body-Mass-Index (BMI)** | 24.44 (5.77) | 23.9 (4.59) | p = 0.769 |
| **Chlorpromazine equivalents (CPZ)** | 30.94 (76.71) | 0 (0) | **p < 0.001** |
| **Symptoms at baseline (SOPS)** |  |  |  |
| Attenuated Positive Symptoms | 13.32 (3.08) | 1.04 (1.6) | **p < 0.001** |
| Negative Symptoms | 12.29 (6.45) | 1.19 (2.01) | **p < 0.001** |
| General Symptoms | 9.43 (4.17) | 1.04 (1.83) | **p < 0.001** |
| Disorganized Symptoms | 5.23 (3.22) | 0.5 (0.85) | **p < 0.001** |
| **Psychosocial functioning (GAF)** | 50.66 (12.01) | 86.37 (8.77) | **p < 0.001** |
| **Cognitive Functioning** |  |  |  |
| Verbal Memory | 26.57 (5.07) | 27.75 (4.2) | p = 0.142 |
| Symbol Coding | 55.2 (13.36) | 60.97 (10.31) | **p < 0.001** |

SOPS, Scale of Psychosis-Risk Symptoms; GAF, Global Assessment of Functioning Scale; p, p-value of Wilcoxon signed-rank test for numeric data or Fisher's exact test for categorical data; SD, standard
deviation. The sample sizes per group refer to the number of participants that were considered for the statistical data analysis. The current CPZ dose at baseline is shown in milligrams.

### Intra-hippocampal connectivity reduction links to worsening clinical changes

As shown in Figure 2A, compared to healthy controls, CHR-P individuals showed a stronger negative association between changes in intra-hippocampal connectivity and changes in negative symptoms (β = −0.18, p < 0.001, p_Bonf_ < 0.001) and depressive symptoms (β = −0.15, p = 0.005, p_Bonf_ = 0.028), as well as a stronger positive association with psychosocial functioning (β = 0.18, p < 0.001, p_Bonf_ = 0.004). No significant interaction effects were found for attenuated positive symptoms, verbal memory, or symbol coding. The full standardized test statistics are provided in the supplementary material. As illustrated descriptively in figures 2B, 2C, and 2D, within the CHR-P group, an average decrease of intra-hippocampal connectivity by 0.1 correlation units over one month was linked to an 1.5 point increase in the negative symptom subscale of the SOPS (p < 0.001), to an 1.1 point increase in the CDSS by (p = 0.018), and to a 3.0 point decrease in the GAF by (p = 0.001).

**Figure 2.**
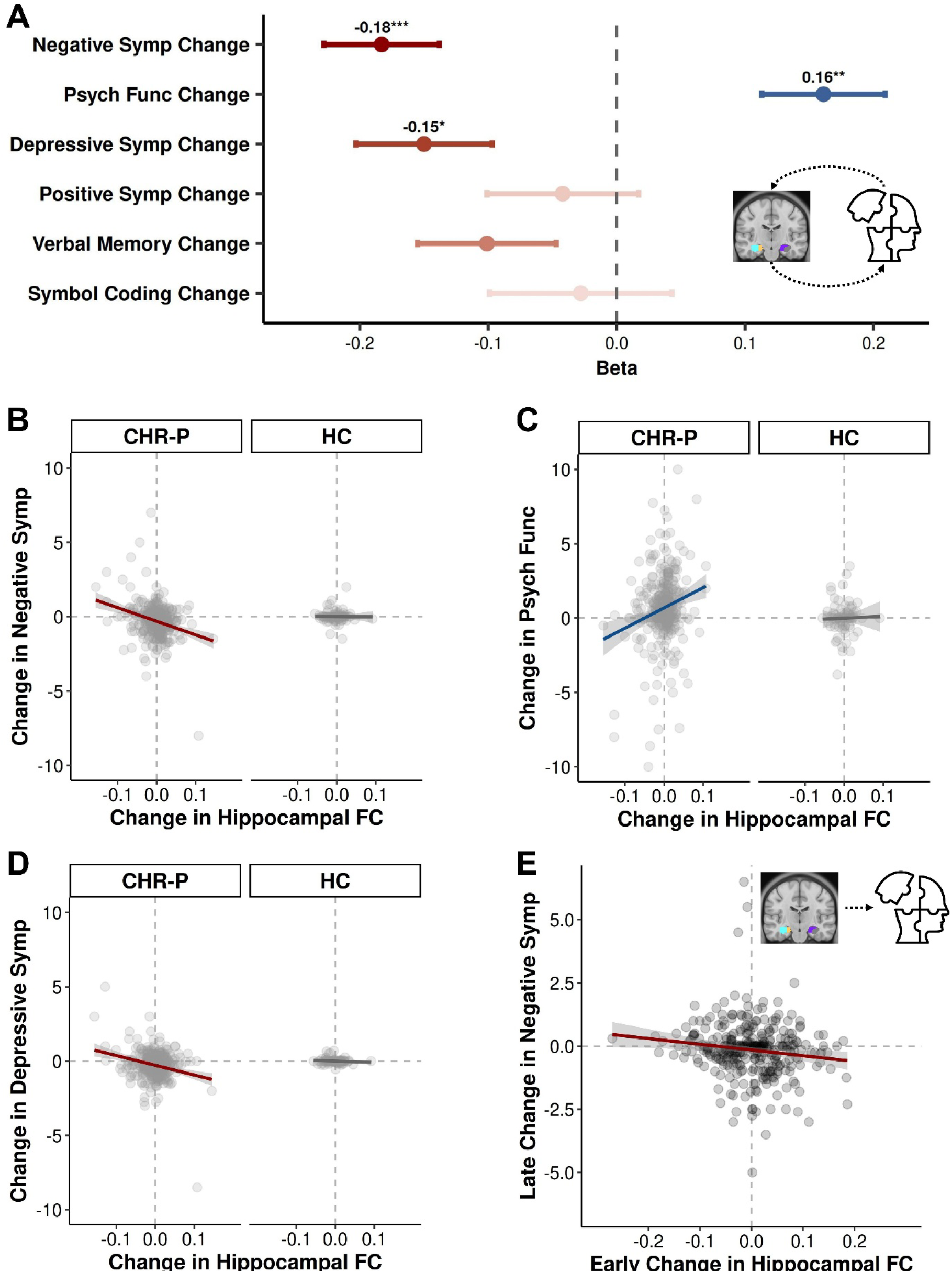
Longitudinal changes in intra-hippocampal connectivity tracks with clinical trajectories. A) The effect size beta of the intra-hippocampal connectivity change x group interaction from the latent variable regression is shown on the x-axis, the six clinical variables are plotted on the y-axis. The dots reflect the point estimates of the effect sizes and the error bars represent the standard errors of the effect sizes. Positive effect sizes are colored blue, negative effect sizes are colored red. The three significant effects are labelled with the exact effect size and the level of significance as asterisks (*, pbonf < 0.05, **, pbonf < 0.01, ***, pbonf < 0.001). A positive effect size indicates that the association between the change in intra-hippocampal connectivity and the clinical variable is more positive in CHR-P subjects compared to controls. A negative effect size indicates that this association is more negative in CHR-P subjects compared to controls. B) Association between change in intra-hippocampal connectivity and negative symptoms within each group. C) Association between change in intra-hippocampal connectivity and psychosocial functioning within each group. D) Association between change in intra-hippocampal connectivity and depressive symptoms within each group. E) Association between early change in intra-hippocampal connectivity and subsequent change in negative symptoms in CHR-P individuals. FC, functional connectivity; Symp, Symptoms; Psych Func, psychosocial functioning; CHR-P, subjects at clinical high risk for psychosis; HC, healthy controls.

As depicted in the supplemental material, our findings were most robust for negative symptoms compared to the other clinical domains (e.g. Monte Carlo simulations), were unaffected by age and changes in antipsychotic medication dose, were most consistent for average intra-hippocampal connectivity compared to left, right, or homotopic hippocampal connectivity, and were specific to the hippocampus in contrast to connectivity within other brain regions and networks (supplementary material, Figure S3).

### Intra-hippocampal connectivity reductions predate worsening negative symptoms

We found that early decreases in intra-hippocampal connectivity were associated with a subsequent worsening in negative symptoms (β = −0.16, p = 0.001, p_Bonf_ = 0.004; Figure 2E), but not in the other clinical domains. In contrast, early changes in negative symptoms were not linked to subsequent changes in intra-hippocampal connectivity. Further details are described in the supplementary material.

## DISCUSSION

Drawing on models of hippocampal pathology and multi-level neural dysconnectivity in psychosis, we examined whether longitudinal changes in intra-hippocampal connectivity track emerging clinical trajectories in the psychosis risk state. Declining intra-hippocampal connectivity marked worsening negative symptoms, depressive symptoms, and psychosocial functioning, with prognostic value specifically for negative symptom progression. At baseline, CHR-P individuals showed lower intra-hippocampal connectivity than controls.

This is the first time connectivity changes within the hippocampus have been shown to track emerging affective-motivational and functional trajectories in the psychosis risk state, with prognostic value for negative symptoms in particular. Our findings broadly align with previous studies in CHR-P individuals. They report hypoconnectivity between the hippocampus and inferior frontal cortex *(17)*, altered connectivity between the CA1 subregion and the ventromedial prefrontal cortex, nucleus accumbens, and amygdala *(18)*, as well as correlations between hippocampus-to-whole-brain connectivity and symptom severity *(19)*. Interventional studies in CHR-P individuals show that exercise and diazepam treatment can modulate connectivity of the hippocampus with multiple brain areas *(18, 27, 28)*, which occurs with symptom benefits across several clinical domains *(27, 28)*. In addition to connectivity, previous evidence indicates hippocampal hyperactivity, manifested as hyperperfusion, in CHR-P individuals *(13–15)*. Stronger hyperperfusion in several hippocampal subfields is particularly observed in those with worse overall functional outcomes *(14, 16)*. Together with our novel findings on intra-hippocampal connectivity, this points toward a broader hippocampal dysfunction in the psychosis risk state comprising subfield-specific hyperactivity, internal desynchronization, and inter-regional decoupling.

Such macroscale dysfunctional patterns observed via functional neuroimaging can be embedded into a broader biological framework that identifies the hippocampus as a central node in the early pathophysiology of psychosis *(10–12)*. The hippocampus is highly sensitive to environmental stressors – such as childhood adversity and poverty – which, when combined with genetic vulnerability, may trigger certain molecular disturbances. Proposed mechanisms are alterations in the redox cycle of the oxygen metabolism, N-methyl-D-aspartate receptor hypofunction, a damage of parvalbumin-positive interneurons, and a loss of synapses in the hippocampus *(11)*. This occurs with deteriorations in other hippocampal cell types and their properties *(29, 30)*. Together, these processes may deteriorate the balance between excitatory and inhibitory signaling within the hippocampus, manifesting in observable focal hyperactivity patterns. Hyperactivity in specific hippocampal subregions may disrupt intra-hippocampal connectivity. This can further extent to connectivity alterations between the hippocampus and other brain areas. Given the interaction of the hippocampus with the hypothalamic pituitary adrenal axis, hippocampal dysfunction is assumed to reinforce an increased sensitivity to environmental stressors, finally resulting in psychosis-related symptoms *(10, 11)*. This is supported by recent evidence suggesting that hippocampal connectivity constitutes a common neural substrate of psychotic illness in general and positive and negative symptoms in particular *(31)*. In the psychosis risk state, this may only apply to negative symptoms, as positive symptoms are only present in an attenuated form. The relationship between hippocampal dysfunction and negative symptoms may be explained by impaired positive reward learning *(32)*. The hippocampus contributes to the formation of internal representations of positive goals, and its dysfunction may disrupt this process, contributing to the experiential domain of negative symptoms, namely anhedonia, avolition and alogia *(32)*.

We explicitly focused on intra-hippocampal connectivity and demonstrated the specificity of this marker by considering connectivity within other brain networks and regions as control conditions. This supports the notion of the hippocampus as the epicenter of neural dysfunction emerging in the psychosis risk state *(12)*. However, we did not investigate longitudinal changes in inter-network connectivity. Meta-analytic evidence in CHR-P individuals suggests that negative symptom severity is linked to hyperconnectivity between the default-mode and salience networks, as well as between the default-mode and central executive networks *(33)*. Other studies also report associations between various network alterations and clinical domains in the psychosis risk state *(34–36)*. These findings may challenge the idea of the hippocampus as the epicenter of neural dysfunction in psychosis *(12)*. Nonetheless, as a crucial hub region, the hippocampus is assumed to exert a significant influence on widespread dysconnectivity in psychosis *(10)*. This potential interplay between the hippocampus and between-network dysconnectivity warrants further investigation in future large-scale longitudinal studies of the psychosis risk state.

Our findings have important clinical implications given that they are most consistent for negative symptoms. These symptoms are very difficult to treat, typically emerge prior to psychosis onset, and increase the likelihood of conversion *(37)*. They can thus be regarded as an early clinical indicator of an unfavorable clinical outcome. Intra-hippocampal connectivity changes may, in turn, represent an earlier occurring predictive neural marker of emerging negative symptoms in the psychosis risk state. This positions intra-hippocampal connectivity as a promising feature included in future predictive modelling approaches aiming to achieve robust risk stratification of individual CHR-P subjects. In addition to risk stratification, preventing the decrease of intra-hippocampal connectivity may represent a novel, mechanistically grounded treatment target to prevent an unfavorable trajectory of early negative symptoms in the psychosis risk state. Based on our longitudinal observational data, we cannot make definite causal claims, but this may be achieved using non-invasive neurostimulation approaches in CHR-P individuals. Recently, stimulation of a site within the right rostromedial prefrontal cortex using transcranial magnetic stimulation has been proposed to indirectly modulate hippocampal connectivity *(31)*. To directly target a subcortical structure such as the hippocampus, non-invasive transcranial ultrasonic stimulation may also be a promising approach *(38)*. Before conducting a randomized-controlled study, neurostimulation approaches require comprehensive evaluation to find an individualized strategy to impact intra-hippocampal connectivity most efficiently. Considerations comprise the exact stimulation target, dose, context, and timing *(39)*.

Beyond the aspect of lacking causality described previously, our work has further limitations that have important implications for future studies. First, while our results were robust across study sites and supported by Monte Carlo simulations, external validation remains necessary to establish generalizability across different population characteristics (e.g., increased ethnic diversity), technical specifications (e.g., MRI acquisition protocols), and other clinical target populations (e.g. manifested psychotic or depressive disorders). The latter is particularly important, as negative symptoms, depressive symptoms, and psychosocial functioning can occur across multiple psychiatric conditions. Hence, evaluating intra-hippocampal connectivity in other target populations may be promising. Second, the wide age range of our sample (12-30 years) covers different developmental phases in which intra-hippocampal connectivity changes may differ. We did not find an effect of age on change in intra-hippocampal connectivity, but future studies could still aim to disentangle the change dynamics of distinct developmental periods in this age span. Third, we estimated the linear change of connectivity and clinical domains over a restricted period of eight months. However, dynamic non-linear fluctuations over a longer time period and their contribution to more general psychiatric outcomes (e.g. transition to psychosis) may also be relevant. Longitudinal data in CHR-P individuals is typically collected over at least two years to cover clinical outcomes such as conversion to psychosis *(26, 40)*. Therefore, future research examining the link between connectivity and clinical trajectories should use deep-sampling approaches to capture non-linear dynamics across longer timeframes.

## MATERIALS AND METHODS

### Study Design

This study utilized demographic, clinical, and MRI data from NAPLS-3 – a large multicenter longitudinal observational study in people at CHR-P conducted in the United States of America *(26)*. Data access and permission was granted by the National Institute of Mental Health Data Archive (collection ID: 2275, DAR credits IDs: 23242 and 23815, dates of last approval: July 11th, 2025; and December 17^th^, 2025). Data were collected across nine US sites, comprising 560 individuals at CHR-P with enhanced risk, 150 CHR-P subjects with non-enhanced risk, and 96 healthy controls aged between 12 and 30 years. Due to MRI data availability, the current work focused on the first five timepoints (baseline, 2, 4, 6, and 8 months) acquired within the first eight months of the study (Figure 1A). We included CHR-P participants and healthy controls with at least two clinical assessments and MRI sessions. As the majority of CHR-P subjects with non-enhanced risk did not undergo two or more imaging sessions (> 95 %), we excluded them in this project. Criteria for enhanced risk and healthy control status are described in the supplementary material. Full details of ethical approval, informed consent, clinical diagnostics, recruitment procedures, inclusion and exclusion criteria, and other study characteristics are described elsewhere *(26)*.

### Clinical Assessments

We studied attenuated positive symptoms, negative symptoms (both using the SOPS: Scale of Psychosis-Risk Symptoms *(41)*), depressive symptoms (CDSS: Calgary Depression Scale for Schizophrenia *(42)*), cognitive functioning (BACS: Brief Assessment of Cognition in Schizophrenia, subscales for symbol coding and verbal memory *(43)*), and psychosocial functioning (GAF: Global Assessment of Functioning *(44)*). Chlorpromazine equivalents based on the Defined Daily Doses method *(45)* were used to measure current antipsychotic medication exposure at each timepoint. Further details are described in the supplementary material.

### MRI Data Acquisition and Processing

Identical imaging acquisition protocols were used across all nine study sites. Harmonization was supported by a structural phantom and nine traveling phantoms scanned twice at each site, as described elsewhere *(26)*. In this work, we utilized the T1-weighted structural sequences and the single-band resting-state functional MRI scans. We evaluated data quality and used fMRIPrep version 23.0.2 for preprocessing (supplementary material, Figure S1) *(46)*. We then smoothed, scaled, detrended, filtered, and denoised (white matter, cerebrospinal fluid, global signal, and ICA-AROMA motion components) the preprocessed functional images (supplementary material).

### Parcellation-based Connectivity Within the Hippocampus

The Human Brainnetome Atlas was used to parcellate the hippocampus along its anterior– posterior functional specialization into a rostral (anterior) and caudal (posterior) subregion *(47)* We extracted the mean regional time series of the Blood Oxygenation Level Dependent (BOLD) signal from the bilateral rostral and caudal hippocampus. We computed the connectivity between these four subregions of the hippocampus (left and right caudal hippocampus and left and right rostral hippocampus) using the Fisher’s r-to-z transformed Pearson correlation. This resulted in six unique hippocampal connectivity measures for each individual. We averaged these six connectivity measures to derive a global score for each subject, indicating the mean connectivity within the hippocampus across rostral and caudal subregions in both hemispheres (Figure 1B, supplementary material). We use the terms *connectivity within the hippocampus* and *intra-hippocampal connectivity* interchangeably to refer to this global score.

### Quantifying Change of Intra-Hippocampal Connectivity and Clinical Domains

We first estimated the within-person rate of change over time for intra-hippocampal connectivity and clinical variables (attenuated positive symptoms, negative symptoms, depressive symptoms, symbol coding performance, verbal memory performance, and psychosocial functioning). To this end, for each participant and each variable, a separate linear regression was fitted with time (baseline, two, four, six, and eight months) as the predictor and the variable value as the outcome, yielding an individual slope (Figure 1C). As the number of available timepoints per subject was limited and varied from two to five, we used latent variable regression, thereby correcting for average slope unreliability (supplementary material).

### Statistical Analysis

Significance level for all analyses was set at α = 0.05. To establish change of intra-hippocampal connectivity over time as a correlate of early clinical changes, six latent variable regressions were fit (one for each clinical domain). In each model, the latent slope of the clinical variable served as the dependent variable. Predictors were the latent slope of intra-hippocampal connectivity, group (CHR-P vs healthy controls), and the group x connectivity interaction. Covariates included age, sex (male and female), and site (dummy-coded with one reference site) (Figure 1C, supplementary material). We estimated the standardized coefficient of the effect size and significance of the group x connectivity interaction and corrected for multiple comparisons using the Bonferroni method across the six distinct interaction terms of the latent variable regressions. We included healthy control data and introduced an interaction term to evaluate whether the associations between connectivity and clinical changes are more pronounced in CHR-P subjects, rather than reflecting an effect in the general population. When the interaction effect was significant after Bonferroni correction, we also interpreted the within-group effects in raw units and their significance. We used Monte Carlo simulations to assess the robustness of the results and conducted several additional sensitivity analyses accounting for antipsychotic medication, differentiating between left, right and inter-hemispheric hippocampal connectivity, and considering potential effects of connectivity within other brain areas (see supplementary material).

Additionally, we probed the temporal sequence of the associations between change in intra-hippocampal connectivity and clinical domains significant in the first analysis, thereby exploring intra-hippocampal connectivity change as a candidate for future prognostic models in the psychosis risk state. We estimated the slope of intra-hippocampal connectivity based on the first two measurement timepoints available for each subject and used the available following timepoints (starting from the latest available timepoint for connectivity) to compute the slopes of the clinical variables (Figure 1D). We performed a latent variable regression similar to the main analysis, but only including CHR-P subjects and one latent change variable for the respective clinical domain. Given that the aim of this analysis was to inform future prognostic models which are typically applied in clinical populations and do not necessarily aim to show specificity for a certain population, we restricted this analysis to CHR-P individuals. Early changes in intra-hippocampal connectivity, age, sex, and site were used to predict subsequent change in the clinical domain (Figure 1D). Then, we used early clinical change as predictor, and subsequent connectivity change as outcome to test for opposite effects and undertook several additional control analyses (supplementary material).

## List of Supplementary Materials

Materials and Methods

Fig S1 to S3

Data files S1 to S9

References (*1*–*31*)

## Acknowledgments

Data and/or research tools used in the preparation of this manuscript were obtained from the National Institute of Mental Health (NIMH) Data Archive (NDA). NDA is a collaborative informatics system created by the National Institutes of Health to provide a national resource to support and accelerate research in mental health. Dataset identifier: 2275 (collection ID). This manuscript reflects the views of the authors and may not reflect the opinions or views of the NIH or of the submitters submitting original data to NDA. Access to the NIMH Data Archive permission group that contains the dataset 2275 was last approved on July 11th 2025 (OMB Control Number: 0925-0667).

## Author contributions

Conceptualization: LR, CL, YT, MD, SZ, AZ

Methodology: LR, CL, YT, SC, MD, SZ, AZ

Investigation: LR, MK

Visualization: LR

Funding acquisition: LR, AS, PF

Project administration: LR, DK, PF

Supervision: CL, IM, JM, VY, DK, MD, SZ, AZ

Writing – original draft: LR

Writing – review & editing: LR, CL, SC, IM, JM, VY, MK, DK, AS, PF, YT, MD, SZ, AZ

## Data and materials availability

The data supporting the findings of this study cannot be made publicly available as they require a data access permission provided by the NIMH Data Archive. All analysis scripts can be made available upon request to LR.

## References and Notes

1. S. Jauhar, M. Johnstone, P. J. McKenna, Schizophrenia. Lancet 399, 473–486 (2022).

2. D. Benrimoh, V. Dlugunovych, A. C. Wright, P. Phalen, M. C. Funaro, M. Ferrara, A. R. Powers, S. W. Woods, S. Guloksuz, A. R. Yung, V. Srihari, J. Shah, On the proportion of patients who experience a prodrome prior to psychosis onset: A systematic review and meta-analysis. Mol Psychiatry 29, 1361–1381 (2024).

3. M. Poletti, L. Pelizza, A. Preti, A. Raballo, Clinical High-Risk for Psychosis (CHR-P) circa 2024: Synoptic analysis and synthesis of contemporary treatment guidelines. Asian Journal of Psychiatry 100, 104142 (2024).

4. A. Catalan, A. Richter, G. Salazar de Pablo, J. Vaquerizo-Serrano, G. Mancebo, B. Pedruzo, C. Aymerich, M. Solmi, M. Á. González-Torres, P. Gil, P. McGuire, P. Fusar-Poli, Proportion and predictors of remission and recovery in first-episode psychosis: Systematic review and meta-analysis. Eur Psychiatry 64, e69 (2021).

5. S. W. Woods, B. C. Walsh, J. Addington, K. S. Cadenhead, T. D. Cannon, B. A. Cornblatt, R. Heinssen, D. O. Perkins, L. J. Seidman, S. I. Tarbox, M. T. Tsuang, E. F. Walker, T. H. McGlashan, Current status specifiers for patients at clinical high risk for psychosis. Schizophr Res 158, 69–75 (2014).

6. T. Y. Lee, S. N. Kim, C. U. Correll, M. S. Byun, E. Kim, J. H. Jang, D.-H. Kang, J.-Y. Yun, J. S. Kwon, Symptomatic and functional remission of subjects at clinical high risk for psychosis: a 2-year naturalistic observational study. Schizophr Res 156, 266–271 (2014).

7. J. Addington, J. Stowkowy, L. Liu, K. S. Cadenhead, T. D. Cannon, B. A. Cornblatt, T. H. McGlashan, D. O. Perkins, L. J. Seidman, M. T. Tsuang, E. F. Walker, C. E. Bearden, D. H. Mathalon, O. Santesteban-Echarri, S. W. Woods, Clinical and functional characteristics of youth at clinical high-risk for psychosis who do not transition to psychosis. Psychol Med 49, 1670–1677 (2019).

8. G. Salazar de Pablo, J. Radua, J. Pereira, I. Bonoldi, V. Arienti, F. Besana, L. Soardo, A. Cabras, L. Fortea, A. Catalan, J. Vaquerizo-Serrano, F. Coronelli, S. Kaur, J. Da Silva, J. I. Shin, M. Solmi, N. Brondino, P. Politi, P. McGuire, P. Fusar-Poli, Probability of Transition to Psychosis in Individuals at Clinical High Risk: An Updated Meta-analysis. JAMA Psychiatry 78, 970–978 (2021).

9. G. Salazar de Pablo, L. Soardo, A. Cabras, J. Pereira, S. Kaur, F. Besana, V. Arienti, F. Coronelli, J. I. Shin, M. Solmi, N. Petros, A. F. Carvalho, P. McGuire, P. Fusar-Poli, Clinical outcomes in individuals at clinical high risk of psychosis who do not transition to psychosis: a meta-analysis. Epidemiol Psychiatr Sci 31, e9 (2022).

10. K. S. F. Damme, The hippocampus: At the nexus of risk and resilience for schizophrenia. Schizophrenia Research 285, 141–148 (2025).

11. S. Knight, R. McCutcheon, D. Dwir, A. A. Grace, O. O’Daly, P. McGuire, G. Modinos, Hippocampal circuit dysfunction in psychosis. Transl Psychiatry 12 (2022), doi:10.1038/s41398-022-02115-5.

12. J. A. Lieberman, R. R. Girgis, G. Brucato, H. Moore, F. Provenzano, L. Kegeles, D. Javitt, J. Kantrowitz, M. M. Wall, C. M. Corcoran, S. A. Schobel, S. A. Small, Hippocampal dysfunction in the pathophysiology of schizophrenia: a selective review and hypothesis for early detection and intervention. Mol Psychiatry 23, 1764–1772 (2018).

13. F. A. Provenzano, J. Guo, M. M. Wall, X. Feng, H. C. Sigmon, G. Brucato, M. B. First, D. L. Rothman, R. R. Girgis, J. A. Lieberman, S. A. Small, Hippocampal Pathology in Clinical High-Risk Patients and the Onset of Schizophrenia. Biological Psychiatry 87, 234–242 (2020).

14. P. Allen, C. A. Chaddock, A. Egerton, O. D. Howes, I. Bonoldi, F. Zelaya, S. Bhattacharyya, R. Murray, P. McGuire, Resting Hyperperfusion of the Hippocampus, Midbrain, and Basal Ganglia in People at High Risk for Psychosis. AJP 173, 392–399 (2016).

15. P. Allen, M. Azis, G. Modinos, M. G. Bossong, I. Bonoldi, C. Samson, B. Quinn, M. J. Kempton, O. D. Howes, J. M. Stone, M. Calem, J. Perez, S. Bhattacharayya, M. R. Broome, A. A. Grace, F. Zelaya, P. McGuire, Increased Resting Hippocampal and Basal Ganglia Perfusion in People at Ultra High Risk for Psychosis: Replication in a Second Cohort. Schizophrenia Bulletin 44, 1323–1331 (2018).

16. G. Modinos, A. Richter, A. Egerton, I. Bonoldi, M. Azis, M. Antoniades, M. Bossong, N. Crossley, J. Perez, J. M. Stone, M. Veronese, F. Zelaya, A. A. Grace, O. D. Howes, P. Allen, P. McGuire, Interactions between hippocampal activity and striatal dopamine in people at clinical high risk for psychosis: relationship to adverse outcomes. Neuropsychopharmacol. 46, 1468–1474 (2021).

17. G. Brunner, R. Gajwani, J. Gross, A. Gumley, R. Krishnadas, S. M. Lawrie, M. Schwannauer, F. Schultze-Lutter, P. J. Uhlhaas, A. Fracasso, Altered functional connectivity of the hippocampus in cortico-subcortical networks in early-stage and emerging psychosis. Eur Arch Psychiatry Clin Neurosci (2025), doi:10.1007/s00406-025-02079-9.

18. N. R. Livingston, A. Kiemes, O. O’Daly, S. R. Knight, P. B. Lukow, L. A. Jelen, T. J. Reilly, A. Dima, M. A. Nettis, C. Casetta, G. A. Devenyi, T. Spencer, A. De Micheli, P. Fusar-Poli, A. A. Grace, S. C. R. Williams, P. McGuire, M. M. Chakravarty, A. Egerton, G. Modinos, Diazepam modulates hippocampal CA1 functional connectivity in people at clinical high-risk for psychosis (2024), doi:10.1101/2024.12.20.24319330.

19. T. Colibazzi, Z. Yang, G. Horga, C.-G. Yan, C. M. Corcoran, K. Klahr, G. Brucato, R. R. Girgis, A. Abi-Dargham, M. P. Milham, B. S. Peterson, Aberrant Temporal Connectivity in Persons at Clinical High Risk for Psychosis. Biological Psychiatry: Cognitive Neuroscience and Neuroimaging 2, 696–705 (2017).

20. S. Chopra, A. Segal, S. Oldham, A. Holmes, K. Sabaroedin, E. R. Orchard, S. M. Francey, B. O’Donoghue, V. Cropley, B. Nelson, J. Graham, L. Baldwin, J. Tiego, H. P. Yuen, K. Allott, M. Alvarez-Jimenez, S. Harrigan, B. D. Fulcher, K. Aquino, C. Pantelis, S. J. Wood, M. Bellgrove, P. D. McGorry, Fornito, Network-Based Spreading of Gray Matter Changes Across Different Stages of Psychosis. JAMA Psychiatry 80, 1246 (2023).

21. K. Friston, H. R. Brown, J. Siemerkus, K. E. Stephan, The dysconnection hypothesis (2016). Schizophrenia Research 176, 83–94 (2016).

22. F. Brandl, M. Avram, B. Weise, J. Shang, B. Simões, T. Bertram, D. Hoffmann Ayala, N. Penzel, D. Gürsel, J. Bäuml, A. M. Wohlschläger, Z. Vukadinovic, N. Koutsouleris, S. Leucht, C. Sorg, Specific Substantial Dysconnectivity in Schizophrenia: A Transdiagnostic Multimodal Meta-analysis of Resting-State Functional and Structural Magnetic Resonance Imaging Studies. Biological Psychiatry 85, 573–583 (2019).

23. S. Genon, B. C. Bernhardt, R. La Joie, K. Amunts, S. B. Eickhoff, The many dimensions of human hippocampal organization and (dys)function. Trends in Neurosciences 44, 977–989 (2021).

24. E. S. Nichols, A. Blumenthal, E. Kuenzel, J. K. Skinner, E. G. Duerden, Hippocampus long-axis specialization throughout development: A meta-analysis. Hum Brain Mapp 44, 4211–4224 (2023).

25. S. Yao, K. M. Kendrick, Reduced homotopic interhemispheric connectivity in psychiatric disorders: evidence for both transdiagnostic and disorder specific features. Psychoradiology 2, 129–145 (2022).

26. J. Addington, L. Liu, K. Brummitt, C. E. Bearden, K. S. Cadenhead, B. A. Cornblatt, M. Keshavan, D. H. Mathalon, T. H. McGlashan, D. O. Perkins, L. J. Seidman, W. Stone, M. T. Tsuang, E. F. Walker, S. W. Woods, T. D. Cannon, North American Prodrome Longitudinal Study (NAPLS 3): Methods and baseline description. Schizophrenia Research 243, 262–267 (2022).

27. K. S. F. Damme, T. Gupta, I. Ristanovic, D. Kimhy, A. D. Bryan, V. A. Mittal, Exercise Intervention in Individuals at Clinical High Risk for Psychosis: Benefits to Fitness, Symptoms, Hippocampal Volumes, and Functional Connectivity. Schizophrenia Bulletin 48, 1394–1405 (2022).

28. D. J. Dean, A. D. Bryan, R. Newberry, T. Gupta, E. Carol, V. A. Mittal, A Supervised Exercise Intervention for Youth at Risk for Psychosis: An Open-Label Pilot Study. J. Clin. Psychiatry 78, e1167–e1173 (2017).

29. M. J. Roeske, C. Konradi, S. Heckers, A. S. Lewis, Hippocampal volume and hippocampal neuron density, number and size in schizophrenia: a systematic review and meta-analysis of postmortem studies. Mol Psychiatry 26, 3524–3535 (2021).

30. A. Schmitt, L. Tatsch, A. Vollhardt, T. Schneider-Axmann, F. J. Raabe, L. Roell, H. Heinsen, P. R. Hof, P. Falkai, C. Schmitz, Decreased Oligodendrocyte Number in Hippocampal Subfield CA4 in Schizophrenia: A Replication Study. Cells 11, 3242 (2022).

31. A. R. Pines, S. B. Frandsen, W. Drew, G. M. Meyer, C. Howard, S. T. Palm, F. L. W. V. J. Schaper, C. Lin, K. Butenko, M. A. Ferguson, M. U. Friedrich, J. H. Grafman, A. D. Kappel, C. Neudorfer, N. S. Rost, L. L. Sanderson, J. J. Taylor, O. Wu, I. Kletenik, J. W. Vogel, A. L. Cohen, A. Horn, M. D. Fox, D. Silbersweig, S. H. Siddiqi, Mapping Lesions That Cause Psychosis to a Human Brain Circuit and Proposed Stimulation Target. JAMA Psychiatry (2025), doi:10.1001/jamapsychiatry.2024.4534.

32. S. R. Marder, D. Umbricht, Negative symptoms in schizophrenia: Newly emerging measurements, pathways, and treatments. Schizophrenia Research 258, 71–77 (2023).

33. L. Del Fabro, A. Schmidt, L. Fortea, G. Delvecchio, A. D’Agostino, J. Radua, S. Borgwardt, P. Brambilla, Functional brain network dysfunctions in subjects at high-risk for psychosis: A meta-analysis of resting-state functional connectivity. Neurosci Biobehav Rev 128, 90–101 (2021).

34. G. Collin, A. Nieto-Castanon, M. E. Shenton, O. Pasternak, S. Kelly, M. S. Keshavan, L. J. Seidman, R. W. McCarley, M. A. Niznikiewicz, H. Li, T. Zhang, Y. Tang, W. S. Stone, J. Wang, S. Whitfield-Gabrieli, Brain functional connectivity data enhance prediction of clinical outcome in youth at risk for psychosis. NeuroImage: Clinical 26, 102108 (2020).

35. J. Kindler, T. Ishida, C. Michel, A.-L. Klaassen, M. Stüble, N. Zimmermann, R. Wiest, M. Kaess, Y. Morishima, Aberrant Brain Dynamics in Individuals With Clinical High Risk of Psychosis. Schizophrenia Bulletin Open 5, sgae002 (2024).

36. L. Hoheisel, L. Kambeitz-Ilankovic, J. Wenzel, S. S. Haas, L. A. Antonucci, A. Ruef, N. Penzel, F. Schultze-Lutter, T. Lichtenstein, M. Rosen, D. B. Dwyer, R. K. R. Salokangas, R. Lencer, P. Brambilla, S. Borgwardt, S. J. Wood, R. Upthegrove, A. Bertolino, S. Ruhrmann, E. Meisenzahl, N. Koutsouleris, G. R. Fink, S. Daun, J. Kambeitz, L. Betz, A. Erkens, E. Gussmann, S. Haas, A. Hasan, C. Hoff, I. Khanyaree, A. Melo, S. Muckenhuber-Sternbauer, J. Köhler, Ö. Öztürk, N. Penzel, D. Popovic, A. Rangnick, S. Von Saldern, R. Sanfelici, M. Spangemacher, A. Tupac, M. F. Urquijo, J. Weiske, A. Wosgien, K. Blume, D. Gebhardt, N. Kaiser, R. Milz, A. Nikolaides, M. Seves, S. Vent, M. Wassen, C. Andreou, L. Egloff, F. Harrisberger, C. Lenz, L. Leanza, A. Mackintosh, R. Smieskova, E. Studerus, A. Walter, S. Widmayer, C. Day, M. Iqbal, M. Pelton, P. Mallikarjun, A. Stainton, A. Lin, A. Denissoff, A. Ellilä, T. From, M. Heinimaa, T. Ilonen, P. Jalo, H. Laurikainen, A. Luutonen, A. Mäkela, J. Paju, H. Pesonen, R.-L. Säilä, A. Toivonen, O. Turtonen, A. B. Solana, M. Abraham, N. Hehn, T. Schirmer, C. Altamura, M. Belleri, F. Bottinelli, A. Ferro, M. Re, E. Monzani, M. Sberna, A. D’Agostino, L. Del Fabro, G. Perna, M. Nobile, A. Alciati, M. Balestrieri, C. Bonivento, G. Cabras, F. Fabbro, M. Garzitto, S. Piccin, Alterations of Functional Connectivity Dynamics in Affective and Psychotic Disorders. Biological Psychiatry: Cognitive Neuroscience and Neuroimaging 9, 765–776 (2024).

37. C. U. Correll, N. R. Schooler, Negative Symptoms in Schizophrenia: A Review and Clinical Guide for Recognition, Assessment, and Treatment. Neuropsychiatr Dis Treat 16, 519–534 (2020).

38. H. Estrada, Y. Chen, T. Lemaire, N. Davoudi, A. Özbek, Q. Parduzi, S. Shoham, D. Razansky, Holographic transcranial ultrasound neuromodulation enhances stimulation efficacy by cooperatively recruiting distributed brain circuits. Nat. Biomed. Eng (2025), doi:10.1038/s41551-025-01449-x.

39. G. Soleimani, M. A. Nitsche, C. A. Hanlon, K. O. Lim, A. Opitz, H. Ekhtiari, Four dimensions of individualization in brain stimulation for psychiatric disorders: context, target, dose, and timing. Neuropsychopharmacol. 50, 857–870 (2025).

40. C. M. J. Wannan, B. Nelson, J. Addington, K. Allott, A. Anticevic, C. Arango, J. T. Baker, C. E. Bearden, T. Billah, S. Bouix, M. R. Broome, K. Buccilli, K. S. Cadenhead, M. E. Calkins, T. D. Cannon, G. Cecci, E. Y. H. Chen, K. I. K. Cho, J. Choi, S. R. Clark, M. J. Coleman, P. Conus, C. M. Corcoran, B. A. Cornblatt, C. M. Diaz-Caneja, D. Dwyer, B. H. Ebdrup, L. M. Ellman, P. Fusar-Poli, L. Galindo, P. A. Gaspar, C. Gerber, L. B. Glenthøj, R. Glynn, M. P. Harms, L. E. Horton, R. S. Kahn, J. Kambeitz, L. Kambeitz-Ilankovic, J. M. Kane, T. Kapur, M. S. Keshavan, S.-W. Kim, N. Koutsouleris, M. Kubicki, J. S. Kwon, K. Langbein, K. E. Lewandowski, G. A. Light, D. Mamah, P. J. Marcy, D. H. Mathalon, P. D. McGorry, V. A. Mittal, M. Nordentoft, A. Nunez, O. Pasternak, G. D. Pearlson, J. Perez, D. O. Perkins, A. R. Powers, D. R. Roalf, F. W. Sabb, J. Schiffman, J. L. Shah, S. Smesny, J. Spark, W. S. Stone, G. P. Strauss, Z. Tamayo, J. Torous, R. Upthegrove, M. Vangel, S. Verma, J. Wang, I. W. Rossum, D. H. Wolf, P. Wolff, S. J. Wood, A. R. Yung, C. Agurto, M. Alvarez-Jimenez, P. Amminger, M. Armando, A. Asgari-Targhi, J. Cahill, R. E. Carrión, E. Castro, S. Cetin-Karayumak, M. Mallar Chakravarty, Y. T. Cho, D. Cotter, S. D’Alfonso, M. Ennis, S. Fadnavis, C. Fonteneau, C. Gao, T. Gupta, R. E. Gur, R. C. Gur, H. K. Hamilton, G. D. Hoftman, G. R. Jacobs, J. Jarcho, J. L. Ji, C. G. Kohler, P. A. Lalousis, S. Lavoie, M. Lepage, E. Liebenthal, J. Mervis, V. Murty, S. C. Nicholas, L. Ning, N. Penzel, R. Poldrack, P. Polosecki, D. N. Pratt, R. Rabin, H. Rahimi Eichi, Y. Rathi, A. Reichenberg, J. Reinen, J. Rogers, B. Ruiz-Yu, I. Scott, J. Seitz-Holland, V. H. Srihari, A. Srivastava, A. Thompson, B. I. Turetsky, B. C. Walsh, T. Whitford, J. T. W. Wigman, B. Yao, H. P. Yuen, U. Ahmed, A. (Jin S. Byun, Y. Chung, K. Do, L. Hendricks, K. Huynh, C. Jeffries, E. Lane, C. Langholm, E. Lin, V. Mantua, G. Santorelli, K. Ruparel, E. Zoupou, T. Adasme, L. Addamo, L. Adery, M. Ali, A. Auther, S. Aversa, S.-H. Baek, K. Bates, A. Bathery, J. M. M. Bayer, R. Beedham, Z. Bilgrami, S. Birch, I. Bonoldi, O. Borders, R. Borgatti, L. Brown, A. Bruna, H. Carrington, R. I. Castillo-Passi, J. Chen, N. Cheng, A. E. Ching, C. Clifford, B.-L. Colton, P. Contreras, S. Corral, S. Damiani, M. Done, A. Estradé, B. A. Etuka, M. Formica, R. Furlan, M. Geljic, C. Germano, R. Getachew, M. Goncalves, A. Haidar, J. Hartmann, A. Jo, O. John, S. Kerins, M. Kerr, I. Kesselring, H. Kim, N. Kim, K. Kinney, M. Krcmar, E. Kotler, M. Lafanechere, C. Lee, J. Llerena, C. Markiewicz, P. Matnejl, A. Maturana, A. Mavambu, R. Mayol-Troncoso, A. McDonnell, A. McGowan, D. McLaughlin, R. McIlhenny, B. McQueen, Y. Mebrahtu, M. Mensi, C. L. M. Hui, Y. N. Suen, S. M. Y. Wong, N. Morrell, M. Omar, A. Partridge, C. Phassouliotis, A. Pichiecchio, P. Politi, C. Porter, U. Provenzani, N. Prunier, J. Raj, S. Ray, V. Rayner, M. Reyes, K. Reynolds, S. Rush, C. Salinas, J. Shetty, C. Snowball, S. Tod, G. Turra-Fariña, D. Valle, S. Veale, S. Whitson, A. Wickham, S. Youn, F. Zamorano, E. Zavaglia, J. Zinberg, S. W. Woods, M. E. Shenton, Accelerating Medicines Partnership® Schizophrenia (AMP® SCZ): Rationale and Study Design of the Largest Global Prospective Cohort Study of Clinical High Risk for Psychosis. Schizophrenia Bulletin 50, 496–512 (2024).

41. T. McGlashan, B. Walsh, S. Woods, The Psychosis-risk Syndrome: Handbook for Diagnosis and Follow-Up. (Oxford University Press, 2010).

42. D. Addington, J. Addington, E. Maticka-Tyndale, Assessing depression in schizophrenia: the Calgary Depression Scale. Br J Psychiatry Suppl, 39–44 (1993).

43. R. Keefe, The Brief Assessment of Cognition in Schizophrenia: reliability, sensitivity, and comparison with a standard neurocognitive battery. Schizophrenia Research 68, 283–297 (2004).

44. J. Endicott, R. L. Spitzer, J. L. Fleiss, J. Cohen, The global assessment scale. A procedure for measuring overall severity of psychiatric disturbance. Arch Gen Psychiatry 33, 766–771 (1976).

45. S. Leucht, M. Samara, S. Heres, J. M. Davis, Dose Equivalents for Antipsychotic Drugs: The DDD Method. Schizophr Bull 42 Suppl 1, S90–94 (2016).

46. O. Esteban, C. J. Markiewicz, R. W. Blair, C. A. Moodie, A. I. Isik, A. Erramuzpe, J. D. Kent, M. Goncalves, E. DuPre, M. Snyder, H. Oya, S. S. Ghosh, J. Wright, J. Durnez, R. A. Poldrack, K. J. Gorgolewski, fMRIPrep: a robust preprocessing pipeline for functional MRI. Nat Methods 16, 111–116 (2019).

47. L. Fan, H. Li, J. Zhuo, Y. Zhang, J. Wang, L. Chen, Z. Yang, C. Chu, S. Xie, A. R. Laird, P. T. Fox, S. B. Eickhoff, C. Yu, T. Jiang, The Human Brainnetome Atlas: A New Brain Atlas Based on Connectional Architecture. Cereb Cortex 26, 3508–3526 (2016).

