## Supplemental Material for "Connectivity within the Hippocampus as a Neural Marker of Early Clinical Trajectories in the Psychosis Risk State"

**-**

*Note: The tables containing the full test statistics (starting with the supplemental section *Latent Variable Regressions Conducted in Main Analysis*) are very large due to the complexity of the latent variable regression models computed. They are attached at the end of this supplemental information.

**Assessment and Processing of Clinical Data**

Enhanced criteria required participants to meet at least one of the following criteria derived from the Scale of Psychosis-Risk Symptoms (SOPS) (1): (i) a rating of ≥4 (moderately severe) on P1 (unusual thought content) or P2 (suspiciousness), (ii) a rating of 3 (moderate) on both P1 and P2, or (iii) impaired cognitive performance in symbol coding and verbal memory derived from the Brief Assessment of Cognition in Schizophrenia (BACS) (2), defined as a score at or below the 10th percentile based on age-adjusted norms for youth or adults. Exclusion criteria for CHR.P subjects were: (i) current or lifetime Axis I psychotic disorder (including affective psychosis), (ii) IQ < 70, (iii) history of a central nervous system disorder, or (iv) psychosis-risk symptoms attributable to another Axis I disorder. Controls could not meet criteria for a psychosis-risk syndrome or any current/past psychotic or Cluster A personality disorder, have a first-degree family history of psychosis or related disorders, or be using psychotropic medication.

Attenuated psychotic and negative symptoms were quantified with the SOPS (1). Depressive symptoms were measured with the Calgary Depression Scale for Schizophrenia (3). The symbol coding test and the Hopkins Verbal Learning Test Revised as part of the BACS battery were deployed to assess cognitive functioning including processing speed and verbal memory (2). Overall psychosocial functioning including symptom severity and daily-life functioning was covered with the Global Assessment of Functioning scale (4). Chlorpromazine equivalents (CPZ) for each subject at each measurement timepoint were computed using the Defined Daily Doses method (5).

We visualized the distributions of each variable at each timepoint and searched for implausible or erroneous data entries. In total, we only removed two values below 10 across all subjects and all sessions of the symbol coding test, assuming that this very low performance is more likely to be driven by lacking motivation than by cognitive impairment in such a young study sample. We further removed two CPZ values of 1500 and 3000 that were driven by a data entry of 25mg risperidone and 150mg aripiprazole, respectively. Both dosages are exceeding the normal dosage range of these drugs by manifold and are thus not realistic. No further clinical data was removed.

**Quality Control and Preprocessing of Resting-State fMRI Data**

The automated quality control software MRIQC version 24.0.0 was utilized to create one HTML report per participant including a comprehensive visual illustration of both the structural T1- and T2-weighted images and the resting state functional MRI sequence (6). These reports were visually inspected for severe artifacts. Furthermore, MRIQC provided a set of different image quality metrics to identify scans with lower image quality compared to others. We documented cases that required further inspection after preprocessing, but did not exclude any scans at this stage already

Next, we ran fMRIPrep version 23.0.2 to preprocess the T1- and T2-weighted images and the resting-state fMRI data (7). The following descriptions encompass all preprocessing steps. Note that not all outputs from all these intermediate steps were used for the subsequent analyses. For anatomical data preprocessing, a single T1-weighted (T1w) image was identified in the input BIDS dataset. The image was corrected for intensity non-uniformity (INU) using N4BiasFieldCorrection (8) as implemented in ANTs 2.3.3 (9) (RRID:SCR_004757), and served as the T1w reference throughout the pipeline. Skull-stripping was performed with the Nipype wrapper of the ANTs antsBrainExtraction.sh workflow, using OASIS30ANTs as the target template. Brain tissue was segmented into cerebrospinal fluid (CSF), white matter (WM), and gray matter (GM) on the skull-stripped T1w image using FAST (FSL 6.0.5.1, RRID:SCR_002823 (10)). Cortical surfaces were reconstructed with recon-all (FreeSurfer 7.3.2, RRID:SCR_001847 (11)). The brain mask was refined with a customized approach based on Mindboggle (RRID:SCR_002438 (12)), reconciling ANTs- and FreeSurfer-derived GM segmentations. Grayordinate “dscalar” files (91k sampleS8) were generated using the highest-resolution fsaverage as an intermediate standardized surface. Spatial normalization was performed to two standard spaces - MNI152NLin2009cAsym and MNI152NLin6Asym - via nonlinear registration with antsRegistration (ANTs 2.3.3), using skull-stripped T1w references and templates. Templates were accessed through TemplateFlow (13) (23.0.0): ICBM 152 Nonlinear Asymmetrical v2009c (14) (RRID:SCR_008796; TemplateFlow ID: MNI152NLin2009cAsym), FSL’s ICBM 152 Non-linear 6th Generation Asymmetric Average Brain (15) (RRID:SCR_002823; TemplateFlow ID: MNI152NLin6Asym).

For functional data preprocessing, each subject contributed one BOLD run per session. Preprocessing began with generating a reference volume and skull-stripped version using fMRIPrep’s custom method. Head-motion parameters (rigid-body transformations and six motion parameters) were estimated with MCFLIRT (16) (FSL 6.0.5.1) before any temporal filtering. BOLD runs were slice-time corrected to 1.21s (midpoint of the 0–2.42s slice acquisition range) using 3dTshift (AFNI (17), RRID:SCR_005927). Head-motion–corrected BOLD time series were resampled to their native space, hereafter referred to as preprocessed BOLD. The BOLD reference was co-registered to the T1w reference using bbregister (18) (FreeSurfer) with 12 degrees of freedom to account for residual distortions. Several confound regressors were derived, including Framewise displacement (FD) (16,19), DVARS, global signals from CSF, WM, and whole brain. CompCor components (20) were extracted following high-pass filtering (128s cut-off) with two variants: tCompCor from the top 2% most variable voxels and aCompCor from probabilistic CSF/WM masks in anatomical space (adjusted to exclude voxels with partial GM volume). Masks were resampled into BOLD space, binarized at 0.99, and PCA was performed. The number of components retained explained ≥50% of variance in each mask. Motion estimates were included in the confounds file, expanded with temporal derivatives and quadratic terms. Volumes exceeding 0.4 mm FD or 1.5 standardized DVARS were flagged as outliers. Additional nuisance regressors were derived from voxels along the brain edge (21). The BOLD series was then resampled to MNI152NLin2009cAsym space and to surface space (fsaverage). ICA-AROMA (22) was applied after excluding non-steady-state volumes and applying 6 mm FWHM spatial smoothing, producing “non-aggressively” denoised runs and aggressive noise regressors. Grayordinate files (91k sampleS8) were generated with fsaverage as intermediate space. All resamplings were executed in a single interpolation step, combining motion, distortion, co-registration, and normalization transforms. Volumetric resampling used antsApplyTransforms (ANTs; Lanczos interpolation (23)), while surface resampling used mri_vol2surf (FreeSurfer). Note that a distortion correction was not performed because fieldmaps were only available for a small subset of the sample. Internal operations relied on Nilearn 0.9.1 (24) (RRID:SCR_001362), primarily within the functional workflow. Detailed descriptions of each step are available in the fMRIPrep documentation.

From the fMRIPrep outputs, we extracted the framewise displacement (FD) time series for each scan, averaged it to derive the mean FD across the whole scanning period, marked all scans with mean FD > 0.4 mm, and visually inspected those again after preprocessing. We selected this rather liberal threshold of 0.4 mm because of the longitudinal character of our study. In other words: We accepted images at the lower end of the quality spectrum, if the image quality was constant across study visits. 87 (approx. 5%) scans had FD > 0.4 mm. All of them were finally excluded from further analyses after an additional visual inspection. Figure S1 depicts an example of three scans that were included (mean FD ≤ 0.4 and three that were not (mean FD > 0.4 and visual inspection) to illustrate the general principle of our data quality evaluation.

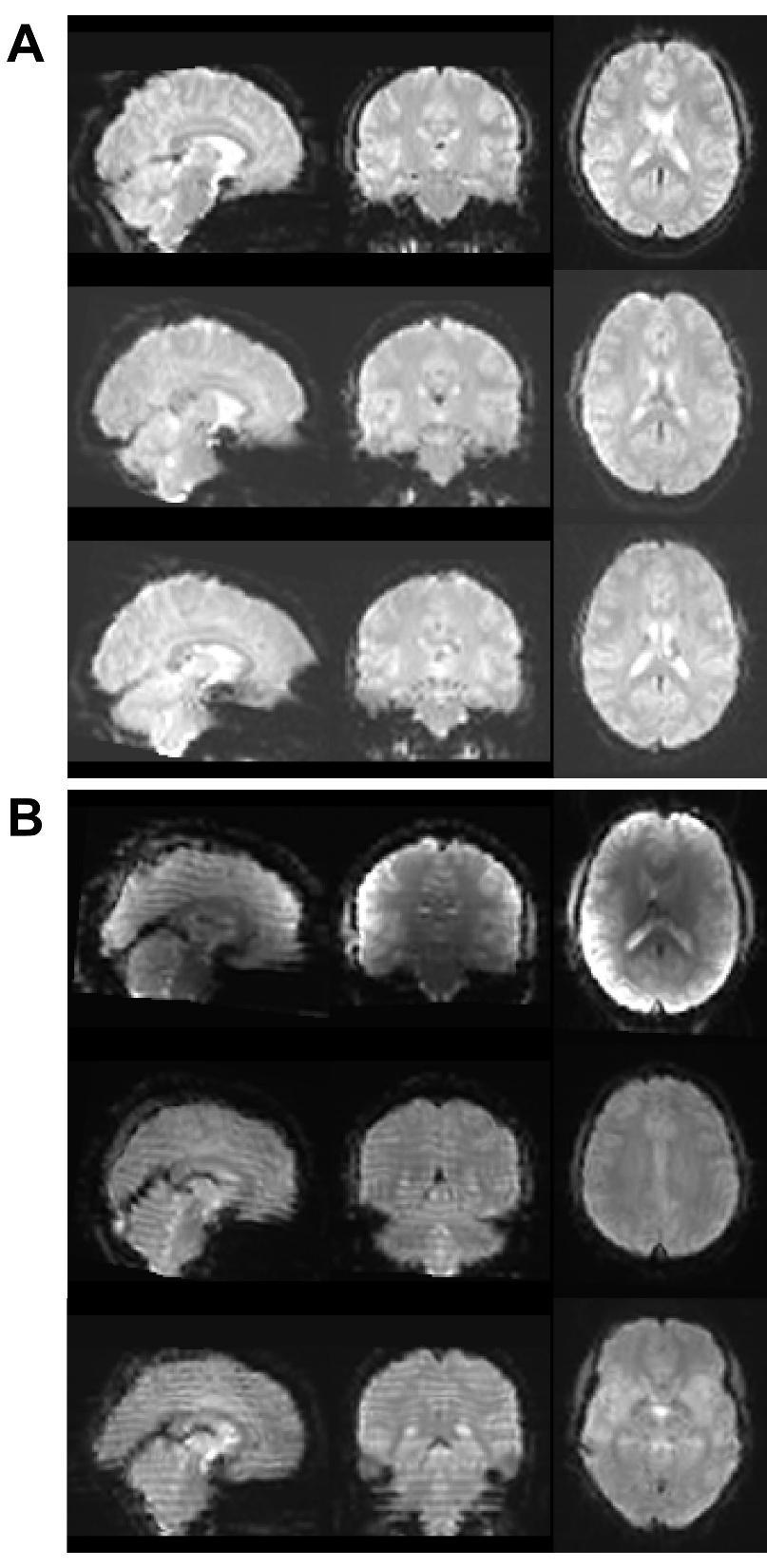

Figure S1: Examples of included and excluded fMRI scans. A) Three examples of scans with sufficient quality that were included. B) Three examples of scans that were excluded due to apparent artifacts corresponding to their mean FD values higher than 0.4mm.

**Postprocessing and Confound Regression of Resting-State fMRI Data**

Postprocessing included smoothing (FWHM = 6 mm), scaling, detrending, filtering (0.01-0.1 Hz), and denoising (white matter, cerebrospinal fluid, global signal, and ICA-AROMA motion components derived from fMRIPrep) using the *smooth_img* and *clean_img* functions as part of the *image* module in Nilearn. Note that *clean_img* performs scaling, detrending, filtering and denoising in one single step to avoid reintroducing artifacts into the fMRI data as outlined by Lindquist et al. (25) We selected ICA-AROMA combined with white matter, cerebrospinal fluid, and global signal as regressors based on a comprehensive comparison of different denoising strategies by Parkes et al., suggesting that this approach represents one of the most reliable strategies across different types of datasets (26). Since ICA-AROMA occasionally misclassifies biological signal as noise, we visually inspected a subset of ten percent of the sample at each study site to evaluate its performance. We did not encounter major errors and no systematic differences across study visits.

From the denoised functional MRI images, we extracted the Blood Oxygenation Level Dependent (BOLD) signal time series from the bilateral rostral and caudal hippocampus, as defined by the Brainnetome atlas. We then calculated the functional connectivity between these four hippocampal subregions (two per hemisphere) using Pearson correlations transformed via Fisher’s r-to-z. Finally, we averaged the individual connections to derive a global score, representing overall functional connectivity across the rostral and caudal hippocampus and between hemispheres. These analyses were performed using 3dNetCorr as part of the Functional And Tractographic Connectivity Analysis Toolbox (FATCAT)(27) and the Analysis of Functional NeuroImages (AFNI) toolbox (17,28).

**Latent Variable Regressions Conducted in Main Analysis**

As described in the main manuscript, we first estimated individual changes in all variables of interest, including hippocampal functional connectivity, attenuated positive and negative symptoms, depressive symptoms, symbol coding performance, verbal memory performance, and psychosocial functioning. To this end, we fitted separate linear regressions for each subject and variable, using time (baseline, 2, 4, 6, and 8 months) as the predictor and the respective variable as the outcome. The slope for time was extracted to reflect each subject’s linear change over time for that variable. As the number of available timepoints per subject was limited in each variable of interest (ranging from two to five) and differed between subjects, precision of individual slope estimation was not optimal. To address this issue, we used latent variable regression, thereby correcting for average slope unreliability. To this end, we extracted the standard errors from the single-subject linear regressions described above. We did that for all subjects that had available data in each variable including CHR-P subjects, non-enhanced CHR-P subjects, and healthy controls. Between 14.6 and 24.9 % of the sample only had two measurement time points available (see Table_S1_Summary_StandardErrorImputation). Thus, the respective single subject regressions did not provide any standard errors of the slope. Therefore, we trained a regression model based on all other subjects with three, four, or five timepoints available in the corresponding variable, using the slope from the single-subject regression, the number of available timepoints, the variance in the predictor time, and the variance in the values of the outcome variable and their interactions to predict the standard errors of the slope. These models had a high explained variance between 49 and 84 % (see Table_S1_Summary_StandardErrorImputation) and were used to estimate the standard errors of the slope for all subjects with only two measurement timepoints. For each outcome, we then computed the mean standard error of the slope of each variable across the final sample considered in the statistics. We then used this mean standard error to create latent slope variables. The goal of this “single-indicator approach” (e.g., Lai & Hsia et al. (2022) (29)) was to free the slope variables from the error that is due to estimating the slope from a limited number of measurement points, and thus to obtain error-free slope variables. These latent slope variables had one indicator each, namely the estimated slope; formally: estimated slope = latent slope + error. The error variance was fixed at the squared mean standard error. The latent slope variables were then employed as outcomes (LS_outcome) and predictor (LS_predictor) in latent variable regressions. These regressions also included a dummy for (non-enhanced) CHR-P subjects (CHRP), the product of this dummy and the predictor (LS_predictor*CHRP), as well as a number of covariates (age, sex, and site): LS_outcome = b0 + b1*LS_predictor + b2*CHRP + b3*LS_predictor*CHRP + b4*sex + b5*age + b5*site02 + b6*site03 + b7*site04 + b8*site05 + b9*site06 + b10*site07 + b11*site08 + b12*site09. The coefficient b3 was of most interest, because it describes the direction and extent to which the relation between LS_predictor and LS_outcome in (non-enhanced) CHR-P subjects differs from that observed in healthy controls.

**Baseline Comparison in Functional Connectivity between CHR-P and Controls**

To compare the functional connectivity within the hippocampus at baseline between CHR-P subjects and controls, we calculated a multiple linear regression model. Hippocampal functional connectivity was used as a dependent variable. Predictors were group (CHR-P vs controls), age, sex (male and female), and site (dummy coded with one reference site). Figure S2 visualizes the descriptive difference between CHR-P and controls.

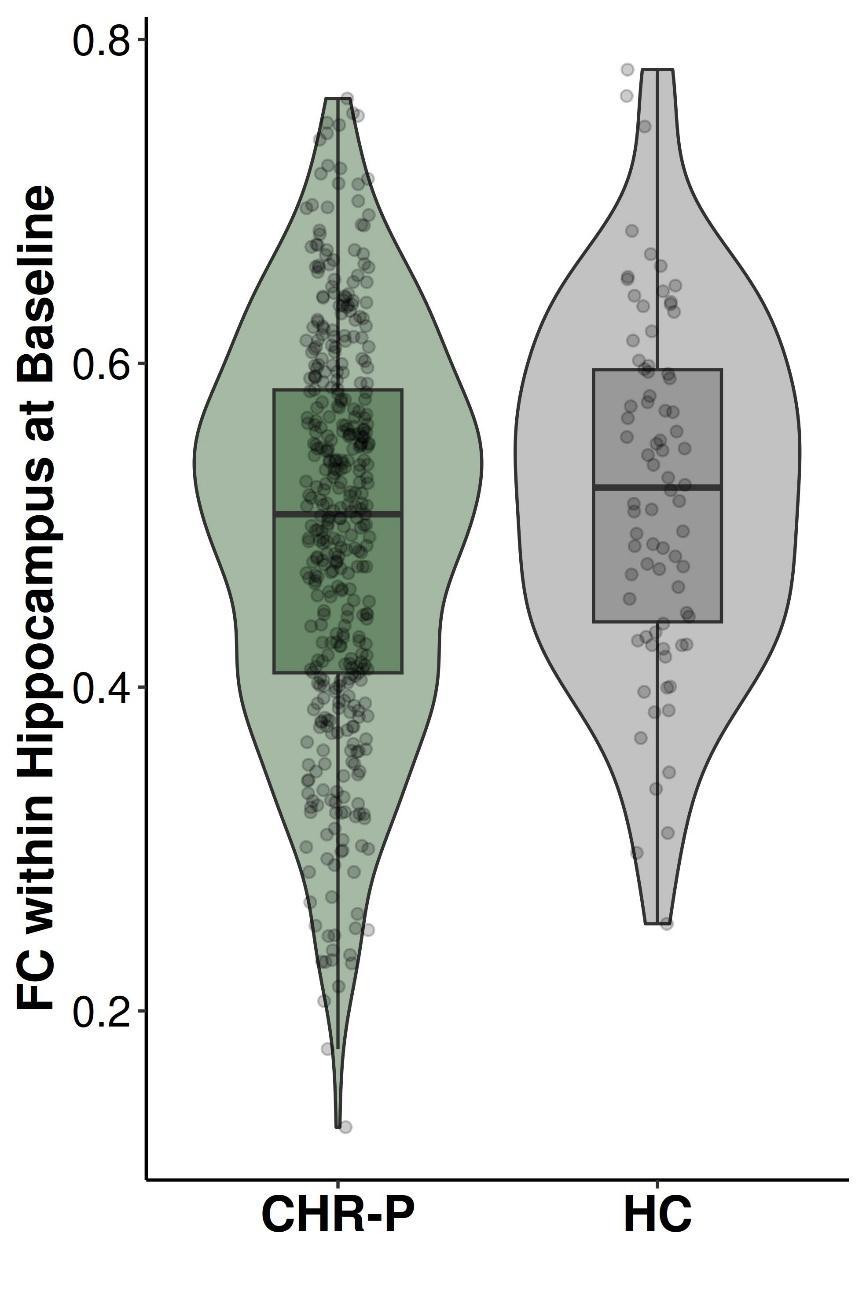

Figure S2: Case-control difference in hippocampal functional connectivity. The two groups (CHR-P = clinical high risk for psychosis, HC = healthy controls) are shown on the x-axis, the average functional connectivity within the hippocampus quantified as Fisher´s r-to-z transformed Pearson´s correlation is indicated on the y-axis.

**Full Standardized Test Statistics from Main Latent Variable Regression Models with Interaction Effects**

Tables S2a and S2b (see Table_S2a_MainAnalysisStandardized.xlsx, Table_S2b_MainAnalysisStandardized_InteractionsCorrected.xlsx) summarize the full standardized test statistics of the six latent variable regression models comparing the link between changes in hippocampal functional connectivity and the clinical changes between people at clinical high risk for psychosis and healthy controls. The models are described in the previous section *Latent Variable Regressions Conducted in Main Analysis*. As outlined in the main manuscript, the p-value from the interaction between groups (subjects at clinical high risk for psychosis and healthy controls) and change in hippocampus functional connectivity was corrected for multiple comparisons using the Bonferroni method across the six clinical scores. Note that Table S2a contains only the uncorrected p values for the main analysis, but includes all paths from the latent variable regression model. Table S2b comprises only the interaction effects of interest, but includes the corrected p values.

Table S2c includes the overall model summary (Table_S2c_MainAnalysis_ModelSummary.xlsx)

**Full Unstandardized Test Statistics from Main Latent Variable Regression Models with Interaction Effects and Within-Group effects**

Table S3 (see Table_S3_MainAnalysisUnstandardized.xlsx) summarize the full unstandardized test statistics of the six latent variable regression models comparing the link between changes in hippocampal functional connectivity and the clinical changes between people at clinical high risk for psychosis and healthy controls. The models are described in the section *Latent Variable Regressions Conducted in Main Analysis*. These tables also include the unstandardized associations between change in hippocampal functional connectivity and change in clinical variables within each group (colored yellow).

**Post-hoc Monte Carlo Simulation Studies for the Main Latent Variable Regression Models**

To evaluate the statistical power of detecting the estimates in our regression models with single-indicator variables, we conducted post-hoc power analyses using Monte Carlo simulation studies as implemented in Mplus 8.6. Table S7 (see Table_S4_MonteCarloSimulation) presents the parameter estimates, standard errors, coverage rates, and statistical power for associations between hippocampal functional connectivity, group (clinical high risk for psychosis = 1, healthy controls = 0), the interaction between hippocampal functional connectivity and group, age, sex (female = 1), site (sites 02–09 in reference to site 01), and the respective outcome variables that were significant in the main analysis (negative symptoms, depressive symptoms, and psychosocial functioning). Population values in all models were specified using the parameter estimates obtained from our primary data analyses. For each simulation study, the sample size was set to match the corresponding N from the empirical analyses. To ensure the stability of the simulation study outcomes, we opted for 10,000 replications with a fixed seed value of 10,000 for the random population draws. The statistical power was assessed by the percentage of replications that rejected the null hypothesis of a parameter being zero (p < .05), or equivalently, the percentage of 95% confidence intervals that excluded zero. Additionally, the accuracy of the interval estimates was evaluated by examining the coverage rates yielded by the simulations, defined as the percentage of outcomes where the intervals encompassed the actual population parameters.

All significant effects of interest demonstrated low to moderate statistical power ranging from 63.4% to 84.6% with highest power estimates for negative symptoms (colored yellow in Table_S4_ MonteCarloSimulation). Coverage rates ranged from 93.8% to 94.2%, indicating that the estimates of the 95% confidence intervals were reasonably accurate. The non-significant effects had negligible statistical power (8 – 26 %).

**Full Test Statistics of Latent Variable Regression Models to Account for Medication**

This latent variable regression model was similar to the one from the main analysis, except that no interaction term was included, as the analysis was only conducted in CHR-P subjects. After computing three latent slope variables for the clinical outcome, hippocampal connectivity, and chlorpromazine equivalents (CPZ), the latent variable regression was fit with the latent clinical slope as dependent variable and the latent hippocampal connectivity slope, age, sex, site and latent CPZ slope as predictors: LS_clinical = b0 + b1*LS_connectivity + b2*LS_cpz + b3*sex + b4*age + b5*site02 + b6*site03 + b7*site04 + b8*site05 + b9*site06 + b10*site07 + b11*site08 + b12*site09. The effect b1 of the latent slope variable of the connectivity within the hippocampus was of interest. Adding antipsychotic medication change was not possible in the main analysis, as healthy controls did not take any antipsychotic medication. The aim of this robustness check was to evaluate if introducing antipsychotic medication change in the model affects the significant associations between change in hippocampal functional connectivity and change in negative symptoms, depressive symptoms, and psychosocial functioning in CHR-P individuals derived from the main analysis. This was not the case. Tables S5a and S5b (see Table_S5a_SensitivityAnalysisStandardized_Medication.xlsx and Table_S5b_SensitivityAnalysisStandardized_Med_Corr.xlsx) summarize the full standardized test statistics of the six latent variable regression models studying the link between changes in hippocampal functional connectivity and the clinical changes only in people at clinical high risk for psychosis considering also changes in antipsychotic medication. The p-value from the change in hippocampal functional connectivity was corrected for multiple comparisons using the Bonferroni method across the six clinical scores. Note that Table S8a contains only the uncorrected p values, but includes all paths from the latent variable regression model. Table S8b comprises only the interaction effects of interest, but includes the corrected p values.

Table S5c includes the overall model summary (Table_S5c_SensitivityAnalysis_Medication_ModelSum.xlsx).

**Full Test Statistics of Latent Variable Regression Models to Predict Change in Hippocampal Functional Connectivity and Clinical Trajectories**

Only in CHR-P subjects, we first investigated whether early changes in hippocampal functional connectivity predict subsequent changes in the clinical variables that showed significant effects in the main analysis. Therefore, for each subject, we identified the first two time points at which hippocampal functional connectivity data was available and estimated the change of connectivity within this period using a single subject regression with the connectivity value as dependent variable and time in months as predictor. Note that in any case only two time points were used to estimate this slope. The average time period for which the connectivity slope was computed in this approach was 2.46 months. We then checked whether clinical data for negative symptoms, depressive symptoms, and psychosocial functioning was available after the period for which connectivity change was calculated. If this was the case, we estimated the slope of the respective clinical variable solely based on time points available after the time points used for the slope estimation of hippocampal connectivity. To perform a correction for potential measurement error, we again extracted the standard errors of the single subject regressions to correct for the average slope unreliability. Note that we did that only for the clinical variables in this analysis, because the slope estimation of hippocampal functioning connectivity was always based on two measurement timepoints, which does not allow us to extract standard errors. Hence a latent slope (LS_clinical_late) was computed for the clinical variables, while the slopes of hippocampal connectivity remained manifest (MS_connectivity_early). We then used the latent variable regression approach again to test whether early changes in hippocampal connectivity are linked to subsequent changes in the three significant clinical variables. The model was specified as follows: LS_clinical_late = b0 + b1*MS_connectivity_early + b2*sex + b3*age + b4*site02 + b5*site03 + b6*site04 + b7*site05 + b8*site06 + b9*site07 + b10*site08 + b11*site09. The effect b1 of the latent slope variable of the connectivity within the hippocampus was of interest.

The results of this analysis are summarized in the manuscript and in the supplemental tables: Table S6a contains the full test statistics of the latent variable regressions used to predict the late change in clinical variables based on early change in hippocampal functional connectivity (Table_S6a_PredAnalysisStand_FCtoPredictClin.xlsx). Table S6b includes the overall model summary of the symptom-by-functional connectivity prediction models (Table_S6b_PredAnalysisStand_FCtoPredictClin_MSum.xlsx). Note that we did not include a table with the Bonferroni-corrected main effects of early hippocampal connectivity change here. Corrected p-values are provided in the main manuscript, computed by multiplying the raw p values of the main effects reported in Table S6a by the number of latent variable regressions (in total: 3) we computed in this prospective association analysis (one for each clinical variable significant in main analysis, namely negative symptoms, depressive symptoms, and psychosocial functioning). We further assessed the statistical power of these effects through Monte Carlo Simulation (see section S7). We achieved a statistical power of 60.2% for negative symptoms, 15.8% for depressive symptoms, and 18.3% for psychosocial functioning. Coverage rates ranged from 94.2% to 94.5%, indicating that the estimates of the 95% confidence intervals were reasonably accurate.

We also examined these predictive effects in the opposite direction to evaluate whether early clinical changes predict subsequent changes of hippocampal connectivity. In these cases, the correction for the average slope unreliability was performed for connectivity changes, but not for clinical variables. The results are summarized in Tables S6c and S6d (Table_S6c_PredAnalysisStand_ClinToPredictFC.xlsx and Table_S6d_PredAnalysisStand_ClinToPredictFC_Msum.xlsx). No significant effects of early clinical change on later connectivity change were determined.

Given that we could not correct for potential measurement error in this analysis, we conducted several robustness checks to evaluate if the measurement error impacted the findings. Instead of always selecting the first two measurement timepoints to compute the hippocampal connectivity slope, we applied a more dynamic approach. For each subject, we first identified if three measurement timepoints are available for hippocampal connectivity and the clinical variable of interest and if these time points cover subsequent time periods (e.g. baseline, 2 months, 4 months for connectivity and 4 months, 6 months, and 8 months for clinical values). If this was the case, both slopes were computed based on all respective timepoints. If this was not the case, we searched for subsequent time periods using only two connectivity timepoints, but at least three clinical timepoints. If no match was found either, we tried to identify two consecutive time periods using only two clinical time points and at least three connectivity time points. If none of these approaches was successful, we searched for a match when only using two time points for both connectivity and the clinical variables. The hippocampal connectivity slope always referred to a time period prior to the clinical slope, while the last time point for connectivity had to match the first time point for the clinical variable. This approach allowed us to correct slopes for potential measurement error. The average time period for which the connectivity slope was computed in this approach was 3.25 months. The model was specified as follows: LS_clinical_late = b0 + b1*LS_connectivity_early + b2*sex + b3*age + b4*site02 + b5*site03 + b6*site04 + b7*site05 + b8*site06 + b9*site07 + b10*site08 + b11*site09. We again obtained a significant association between early functional connectivity changes and late negative symptom change (beta = -0.13, p = 0.032). To evaluate if measurement error plays a role, we ran this analysis again, but without performing the correction of both slopes (beta = -0.09, p = 0.034), the connectivity slope (beta = -0.11, p = 0.033), and the clinical slope (beta = -0.11, p = 0.032). The results remained unchanged. Thus, in our data, a lacking correction for potential measurement error did not induce any spurious significant effects. However, the effect is much stronger when using only two measurement time points to compute the early hippocampal connectivity slope. Given that the measurement error did not play a role, this difference must result from the different average time periods for which early hippocampal connectivity slope was computed in both approaches (2.46 vs 3.25 months). With regard to a potential future clinical translation, this finding is encouraging, as only two MRI measurements would be needed to monitor hippocampal connectivity, aiming to predict future negative symptoms. However, we acknowledge that clinical translation still requires several additional validation steps.

**Full Test Statistics of Latent Variable Regression Models Including Functional Connectivity Patterns within the Hippocampus**

These models were set-up equally to the main analysis, but with changes in specific functional connectivity patterns as predictors instead of the global hippocampal functional connectivity score.

Tables S7a and S7b (see Table_S7a_SensAnalysisStand_WithinHippo.xlsx and Table_S7b_SensAnalysisStand_WithinHippo_Corr.xlsx) summarize the full test statistics of the 18 latent variable regression models (six clinical scores x three hippocampal functional connectivity outcomes) comparing the link between changes in hippocampal functional connectivity and the clinical changes between people at clinical high risk for psychosis and healthy controls. In line with the main analysis, the p-value from the interaction between group (subjects at clinical high risk for psychosis and healthy controls) and change in hippocampal functional connectivity pattern was corrected for multiple comparisons using the Bonferroni method for each hippocampal functional connectivity score separately across the six clinical scores. Note that Table S7a contains only the uncorrected p values for this analysis, but includes all paths from the latent variable regression model. Table S7b comprises only the interaction effects of interest, but includes the corrected p values.

Table S7c includes the overall model summary (Table_S7c_ SensAnalysisStand_WithinHippo_ModelSum.xlsx).

**Computation of Functional Connectivity in Regions and Networks Other than the Hippocampus**

Table S8 (Table_S8_BrainnetomeRegionToNetworkAssigments.xlsx) provides an overview of which regions from the Brainnetome atlas (30) (https://atlas.brainnetome.org/bnatlas.html) were utilized to compute functional connectivity within certain regions and networks. Note that for the brain networks this mapping was performed based on the visual overlap of the respective brain region in the Brainnetome atlas and the main networks from the parcellation proposed by Laird et al. (31). Functional connectivity between all Brainnetome-based subregions assigned to one overall region or network was averaged across hemispheres to derive one mean functional connectivity score per region and network. This mean score was used for the downstream statistical analyses.

**Full Test Statistics of Latent Variable Regression Models Considering Brain Regions and Networks Other Than the Hippocampus**

These models were set-up equally to the main analysis, but with changes in functional connectivity patterns of other brain networks and regions as predictors instead of the global hippocampal functional connectivity score. Figure S3 illustrates that the effect was not significant for any connectivity within any other brain region or network.

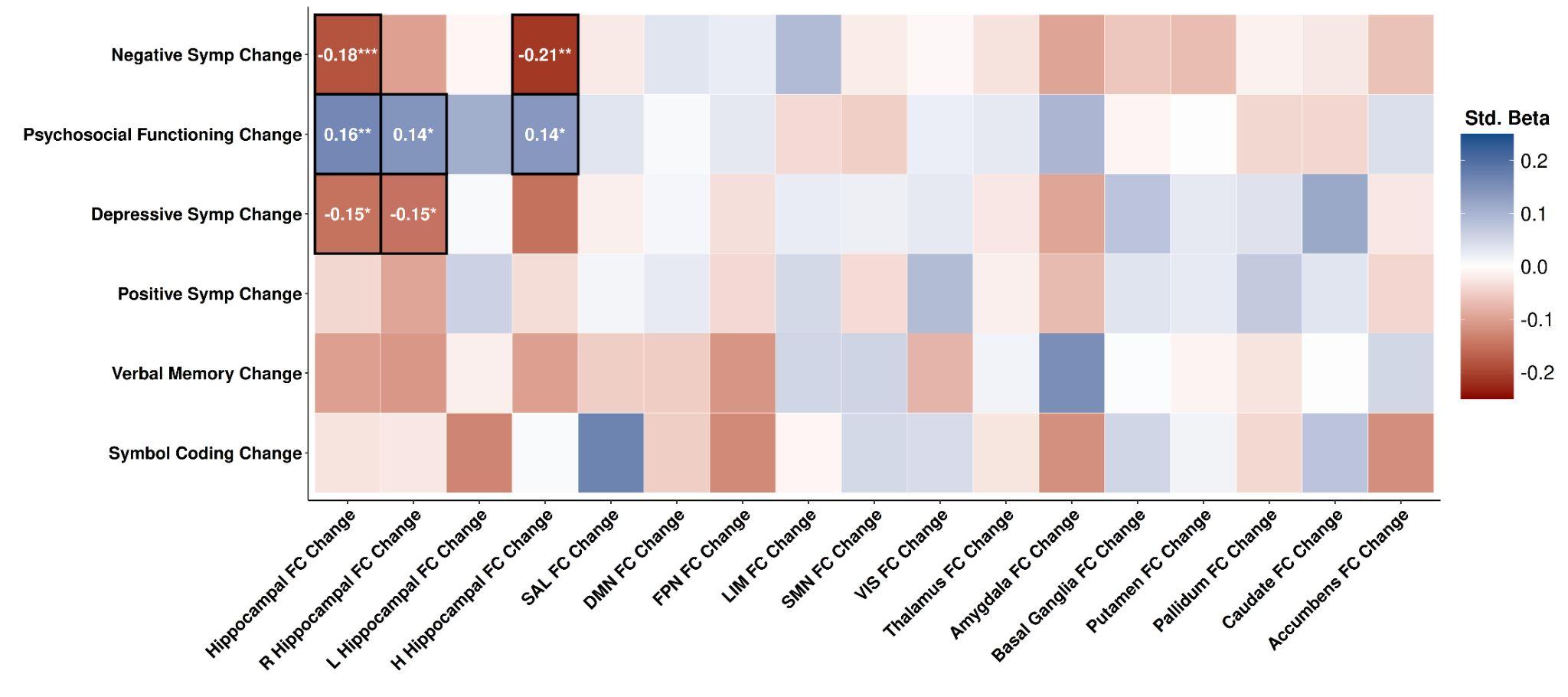

Figure S3. Effect sizes of the hippocampal connectivity change x group interaction for differential hippocampal connectivity patterns and other network- and region-based connectivity scores. The significant effects are marked with a black border, the exact effect size, and the level of significance as asterisks (*, p_bonf_ < 0.05, **, p_bonf_ < 0.01, ***, p_bonf_ < 0.001). Std., standardized; Symp, Symptoms; rHipp, rostral hippocampus; cHipp caudal hippocampus; CHR-P, subjects at clinical high risk for psychosis; HC, healthy controls; L, left; R, right; H, homotopic; VIS, visual network; SMN, somatosensory network, LIM, limbic network; FPN, fronto-parietal network; DMN, default-mode network; SAL, salience network.

Table S9a (Table_S9a_SensAnalysisStand_OtherNetw_Corr.xlsx) summarize the test statistics of the 78 latent variable regression models (six clinical scores x 13 functional connectivity outcomes) comparing the links between changes in functional connectivity in certain regions and networks and the clinical changes between people at clinical high risk for psychosis and healthy controls. Analogously to the main analysis, the p-value from the interaction between group (subjects at clinical high risk for psychosis and healthy controls) and change in network- or region-specific functional connectivity was corrected for multiple comparisons using the Bonferroni method for each functional connectivity score separately across the six clinical scores.

Table S9b includes the overall model summary (Table_S9b_SensAnalysisStand_OtherNetw_ModelSum.xlsx).

**Supplementary References**

1. McGlashan T, Walsh B, Woods S (2010): *The Psychosis-Risk Syndrome: Handbook for Diagnosis and Follow-Up.* Oxford University Press.

2. Keefe R (2004): The Brief Assessment of Cognition in Schizophrenia: reliability, sensitivity, and comparison with a standard neurocognitive battery. *Schizophrenia Research* 68: 283–297.

3. Addington D, Addington J, Maticka-Tyndale E (1993): Assessing depression in schizophrenia: the Calgary Depression Scale. *Br J Psychiatry Suppl* 39–44.

4. Endicott J, Spitzer RL, Fleiss JL, Cohen J (1976): The global assessment scale. A procedure for measuring overall severity of psychiatric disturbance. *Arch Gen Psychiatry* 33: 766–771.

5. Leucht S, Samara M, Heres S, Davis JM (2016): Dose Equivalents for Antipsychotic Drugs: The DDD Method. *Schizophr Bull* 42 Suppl 1: S80-94.

6. Esteban O, Birman D, Schaer M, Koyejo OO, Poldrack RA, Gorgolewski KJ (2017): MRIQC: Advancing the automatic prediction of image quality in MRI from unseen sites. *PLoS One* 12: e0184661.

7. Esteban O, Markiewicz CJ, Blair RW, Moodie CA, Isik AI, Erramuzpe A, *et al.* (2019): fMRIPrep: a robust preprocessing pipeline for functional MRI. *Nat Methods* 16: 111–116.

8. Tustison NJ, Avants BB, Cook PA, Zheng Y, Egan A, Yushkevich PA, Gee JC (2010): N4ITK: Improved N3 Bias Correction. *IEEE Transactions on Medical Imaging* 29: 1310–1320.

9. Avants B, Epstein C, Grossman M, Gee J (2008): Symmetric diffeomorphic image registration with cross-correlation: Evaluating automated labeling of elderly and neurodegenerative brain. *Medical Image Analysis* 12: 26–41.

10. Zhang Y, Brady M, Smith S (2001): Segmentation of brain MR images through a hidden Markov random field model and the expectation-maximization algorithm. *IEEE Transactions on Medical Imaging* 20: 45–57.

11. Dale A, Fischl B, Sereno MI (1999): Cortical Surface-Based Analysis: I. Segmentation and Surface Reconstruction. *NeuroImage* 9: 179–194.

12. Klein A, Ghosh SS, Bao FS, Giard J, Häme Y, Stavsky E, *et al.* (2017): Mindboggling morphometry of human brains ((D. Schneidman, editor)). *PLoS Comput Biol* 13: e1005350.

13. Ciric R, Thompson WH, Lorenz R, Goncalves M, MacNicol EE, Markiewicz CJ, *et al.* (2022): TemplateFlow: FAIR-sharing of multi-scale, multi-species brain models. *Nat Methods* 19: 1568–1571.

14. Fonov V, Evans A, McKinstry R, Almli C, Collins D (2009): Unbiased nonlinear average age-appropriate brain templates from birth to adulthood. *NeuroImage* 47: S62.

15. Evans AC, Janke AL, Collins DL, Baillet S (2012): Brain templates and atlases. *NeuroImage* 62: 911–922.

16. Jenkinson M, Bannister P, Brady M, Smith S (2002): Improved optimization for the robust and accurate linear registration and motion correction of brain images. *Neuroimage* 17: 825–841.

17. Cox RW, Hyde JS (1997): Software tools for analysis and visualization of fMRI data. *NMR Biomed* 10: 171–178.

18. Greve DN, Fischl B (2009): Accurate and robust brain image alignment using boundary-based registration. *NeuroImage* 48: 63–72.

19. Power JD, Mitra A, Laumann TO, Snyder AZ, Schlaggar BL, Petersen SE (2014): Methods to detect, characterize, and remove motion artifact in resting state fMRI. *NeuroImage* 84: 320–341.

20. Behzadi Y, Restom K, Liau J, Liu TT (2007): A component based noise correction method (CompCor) for BOLD and perfusion based fMRI. *NeuroImage* 37: 90–101.

21. Patriat R, Reynolds RC, Birn RM (2017): An improved model of motion-related signal changes in fMRI. *NeuroImage* 144: 74–82.

22. Pruim RHR, Mennes M, Buitelaar JK, Beckmann CF (2015): Evaluation of ICA-AROMA and alternative strategies for motion artifact removal in resting state fMRI. *Neuroimage* 112: 278–287.

23. Lanczos C (1964): Evaluation of Noisy Data. *Journal of the Society for Industrial and Applied Mathematics Series B Numerical Analysis* 1: 76–85.

24. Abraham A, Pedregosa F, Eickenberg M, Gervais P, Mueller A, Kossaifi J, *et al.* (2014): Machine learning for neuroimaging with scikit-learn. *Front Neuroinform* 8. https://doi.org/10.3389/fninf.2014.00014

25. Lindquist MA, Geuter S, Wager TD, Caffo BS (2019): Modular preprocessing pipelines can reintroduce artifacts into fMRI data. *Hum Brain Mapp* 40: 2358–2376.

26. Parkes L, Fulcher B, Yücel M, Fornito A (2018): An evaluation of the efficacy, reliability, and sensitivity of motion correction strategies for resting-state functional MRI. *NeuroImage* 171: 415–436.

27. Taylor PA, Saad ZS (2013): FATCAT: (an efficient) Functional and Tractographic Connectivity Analysis Toolbox. *Brain Connect* 3: 523–535.

28. Cox RW (1996): AFNI: software for analysis and visualization of functional magnetic resonance neuroimages. *Comput Biomed Res* 29: 162–173.

29. Lai MHC, Hsiao Y-Y (2022): Two-stage path analysis with definition variables: An alternative framework to account for measurement error. *Psychol Methods* 27: 568–588.

30. Fan L, Li H, Zhuo J, Zhang Y, Wang J, Chen L, *et al.* (2016): The Human Brainnetome Atlas: A New Brain Atlas Based on Connectional Architecture. *Cereb Cortex* 26: 3508–3526.

31. Laird AR, Fox PM, Eickhoff SB, Turner JA, Ray KL, McKay DR, *et al.* (2011): Behavioral interpretations of intrinsic connectivity networks. *J Cogn Neurosci* 23: 4022–4037.

Table_S1_Summary_StandardErrorImputation

| **Outcome** | **R2** | **Model_p_value** | **N_total_with_nt_larger2** | **N_imputed** | **Prop_imputed** |
| --- | --- | --- | --- | --- | --- |
| cpz | 0.841242 | 4.25E-215 | 683 | 100 | 0.146412884 |
| DepS | 0.686001 | 3.86E-104 | 560 | 89 | 0.158928571 |
| fcacu | 0.688419 | 3.42E-71 | 445 | 111 | 0.249438202 |
| fcamy | 0.625349 | 9.88E-59 | 445 | 111 | 0.249438202 |
| fcbg | 0.585137 | 7.07E-52 | 445 | 111 | 0.249438202 |
| fccau | 0.538209 | 1.04E-44 | 445 | 111 | 0.249438202 |
| fcdmn | 0.603793 | 5.75E-55 | 445 | 111 | 0.249438202 |
| fcfpn | 0.7304 | 5.09E-81 | 445 | 111 | 0.249438202 |
| fchi | 0.491982 | 2.26E-38 | 445 | 111 | 0.249438202 |
| fchih | 0.508623 | 1.40E-40 | 445 | 111 | 0.249438202 |
| fchil | 0.604532 | 4.31E-55 | 445 | 111 | 0.249438202 |
| fchir | 0.631526 | 7.48E-60 | 445 | 111 | 0.249438202 |
| fclim | 0.558894 | 9.05E-48 | 445 | 111 | 0.249438202 |
| fcpal | 0.598068 | 5.29E-54 | 445 | 111 | 0.249438202 |
| fcput | 0.532247 | 7.42E-44 | 445 | 111 | 0.249438202 |
| fcsal | 0.515176 | 1.80E-41 | 445 | 111 | 0.249438202 |
| fcsmn | 0.661072 | 1.70E-65 | 445 | 111 | 0.249438202 |
| fctha | 0.50072 | 1.60E-39 | 445 | 111 | 0.249438202 |
| fcvis | 0.605934 | 2.49E-55 | 445 | 111 | 0.249438202 |
| gaf | 0.796686 | 2.53E-146 | 560 | 90 | 0.160714286 |
| NegS | 0.702178 | 1.58E-108 | 553 | 85 | 0.153707052 |
| PosS | 0.713242 | 1.69E-142 | 683 | 101 | 0.147877013 |
| SyCod | 0.659815 | 6.48E-93 | 545 | 89 | 0.163302752 |
| VeMem | 0.666925 | 2.28E-95 | 544 | 86 | 0.158088235 |

Table_S2a_MainAnalysisStandardized

| **paramHeader** | **param** | **est** | **se** | **est_se** | **pval** | **zval** | **pval_exact** |
| --- | --- | --- | --- | --- | --- | --- | --- |
| LV_HI.BY | HI_CH | 0.812 | 0.025 | 32.792 | 0 | 32.48 | 0 |
| LV_DEPS.BY | DEPS_CH | 0.863 | 0.036 | 23.738 | 0 | 23.97222 | 0 |
| LV_DEPS.ON | LV_HI | 0.004 | 0.036 | 0.103 | 0.918 | 0.111111 | 0.911528 |
| LV_DEPS.ON | HIXCHR | -0.15 | 0.053 | -2.811 | 0.005 | -2.83019 | 0.004652 |
| LV_DEPS.ON | CHR | -0.149 | 0.032 | -4.595 | 0 | -4.65625 | 3.22E-06 |
| LV_DEPS.ON | AGE | -0.091 | 0.048 | -1.892 | 0.058 | -1.89583 | 0.057982 |
| LV_DEPS.ON | FEMALE | -0.029 | 0.051 | -0.571 | 0.568 | -0.56863 | 0.569609 |
| LV_DEPS.ON | SITE02 | -0.033 | 0.095 | -0.349 | 0.727 | -0.34737 | 0.728315 |
| LV_DEPS.ON | SITE03 | 0.112 | 0.064 | 1.751 | 0.08 | 1.75 | 0.080118 |
| LV_DEPS.ON | SITE04 | 0.038 | 0.066 | 0.574 | 0.566 | 0.575758 | 0.564779 |
| LV_DEPS.ON | SITE05 | 0.107 | 0.074 | 1.459 | 0.145 | 1.445946 | 0.148192 |
| LV_DEPS.ON | SITE06 | 0.045 | 0.076 | 0.595 | 0.552 | 0.592105 | 0.55378 |
| LV_DEPS.ON | SITE07 | 0.063 | 0.071 | 0.883 | 0.377 | 0.887324 | 0.374905 |
| LV_DEPS.ON | SITE08 | 0.03 | 0.075 | 0.394 | 0.694 | 0.4 | 0.689157 |
| LV_DEPS.ON | SITE09 | 0.159 | 0.081 | 1.953 | 0.051 | 1.962963 | 0.04965 |
| LV_HI.WITH | CHR | 0 | 0.047 | 0.005 | 0.996 | 0 | 1 |
| LV_HI.WITH | AGE | 0.022 | 0.056 | 0.386 | 0.7 | 0.392857 | 0.694425 |
| LV_HI.WITH | FEMALE | -0.085 | 0.06 | -1.416 | 0.157 | -1.41667 | 0.15658 |
| LV_HI.WITH | SITE02 | 0.025 | 0.06 | 0.411 | 0.681 | 0.416667 | 0.676922 |
| LV_HI.WITH | SITE03 | -0.011 | 0.085 | -0.132 | 0.895 | -0.12941 | 0.897032 |
| LV_HI.WITH | SITE04 | 0.065 | 0.047 | 1.391 | 0.164 | 1.382979 | 0.166671 |
| LV_HI.WITH | SITE05 | -0.062 | 0.062 | -1.009 | 0.313 | -1 | 0.317311 |
| LV_HI.WITH | SITE06 | 0.074 | 0.045 | 1.666 | 0.096 | 1.644444 | 0.100084 |
| LV_HI.WITH | SITE07 | -0.01 | 0.064 | -0.152 | 0.879 | -0.15625 | 0.875836 |
| LV_HI.WITH | SITE08 | -0.013 | 0.057 | -0.224 | 0.823 | -0.22807 | 0.819592 |
| LV_HI.WITH | SITE09 | -0.043 | 0.059 | -0.727 | 0.467 | -0.72881 | 0.466116 |
| CHR.WITH | AGE | -0.049 | 0.051 | -0.956 | 0.339 | -0.96078 | 0.336661 |
| CHR.WITH | FEMALE | -0.032 | 0.048 | -0.67 | 0.503 | -0.66667 | 0.504985 |
| CHR.WITH | SITE02 | -0.046 | 0.053 | -0.881 | 0.378 | -0.86792 | 0.385436 |
| CHR.WITH | SITE03 | -0.034 | 0.052 | -0.645 | 0.519 | -0.65385 | 0.513211 |
| CHR.WITH | SITE04 | 0.017 | 0.046 | 0.377 | 0.706 | 0.369565 | 0.711706 |
| CHR.WITH | SITE05 | -0.001 | 0.048 | -0.022 | 0.982 | -0.02083 | 0.983379 |
| CHR.WITH | SITE06 | -0.017 | 0.049 | -0.347 | 0.728 | -0.34694 | 0.728637 |
| CHR.WITH | SITE07 | -0.004 | 0.048 | -0.087 | 0.931 | -0.08333 | 0.933586 |
| CHR.WITH | SITE08 | 0.024 | 0.045 | 0.538 | 0.591 | 0.533333 | 0.593803 |
| CHR.WITH | SITE09 | 0.008 | 0.047 | 0.17 | 0.865 | 0.170213 | 0.864843 |
| AGE.WITH | FEMALE | -0.11 | 0.048 | -2.305 | 0.021 | -2.29167 | 0.021925 |
| AGE.WITH | SITE02 | 0.304 | 0.052 | 5.894 | 0 | 5.846154 | 5.03E-09 |
| AGE.WITH | SITE03 | 0 | 0.044 | 0.003 | 0.998 | 0 | 1 |
| AGE.WITH | SITE04 | -0.013 | 0.038 | -0.343 | 0.732 | -0.34211 | 0.732272 |
| AGE.WITH | SITE05 | 0.155 | 0.043 | 3.59 | 0 | 3.604651 | 0.000313 |
| AGE.WITH | SITE06 | -0.009 | 0.04 | -0.222 | 0.824 | -0.225 | 0.821979 |
| AGE.WITH | SITE07 | -0.143 | 0.046 | -3.123 | 0.002 | -3.1087 | 0.001879 |
| AGE.WITH | SITE08 | -0.17 | 0.049 | -3.474 | 0.001 | -3.46939 | 0.000522 |
| AGE.WITH | SITE09 | -0.03 | 0.044 | -0.676 | 0.499 | -0.68182 | 0.495354 |
| FEMALE.WITH | SITE02 | 0.029 | 0.048 | 0.608 | 0.543 | 0.604167 | 0.545733 |
| FEMALE.WITH | SITE03 | -0.015 | 0.048 | -0.317 | 0.751 | -0.3125 | 0.754661 |
| FEMALE.WITH | SITE04 | -0.003 | 0.048 | -0.071 | 0.943 | -0.0625 | 0.950165 |
| FEMALE.WITH | SITE05 | 0.005 | 0.048 | 0.104 | 0.917 | 0.104167 | 0.917037 |
| FEMALE.WITH | SITE06 | -0.06 | 0.047 | -1.282 | 0.2 | -1.2766 | 0.201745 |
| FEMALE.WITH | SITE07 | 0.065 | 0.048 | 1.343 | 0.179 | 1.354167 | 0.175683 |
| FEMALE.WITH | SITE08 | 0.003 | 0.048 | 0.053 | 0.958 | 0.0625 | 0.950165 |
| FEMALE.WITH | SITE09 | 0.013 | 0.048 | 0.276 | 0.783 | 0.270833 | 0.786519 |
| SITE02.WITH | SITE03 | -0.078 | 0.01 | -7.882 | 0 | -7.8 | 6.22E-15 |
| SITE02.WITH | SITE04 | -0.096 | 0.011 | -8.758 | 0 | -8.72727 | 0 |
| SITE02.WITH | SITE05 | -0.125 | 0.013 | -9.754 | 0 | -9.61538 | 0 |
| SITE02.WITH | SITE06 | -0.119 | 0.012 | -9.584 | 0 | -9.91667 | 0 |
| SITE02.WITH | SITE07 | -0.124 | 0.013 | -9.722 | 0 | -9.53846 | 0 |
| SITE02.WITH | SITE08 | -0.106 | 0.012 | -9.176 | 0 | -8.83333 | 0 |
| SITE02.WITH | SITE09 | -0.128 | 0.013 | -9.848 | 0 | -9.84615 | 0 |
| SITE03.WITH | SITE04 | -0.078 | 0.01 | -7.882 | 0 | -7.8 | 6.22E-15 |
| SITE03.WITH | SITE05 | -0.102 | 0.012 | -8.584 | 0 | -8.5 | 0 |
| SITE03.WITH | SITE06 | -0.097 | 0.011 | -8.467 | 0 | -8.81818 | 0 |
| SITE03.WITH | SITE07 | -0.101 | 0.012 | -8.562 | 0 | -8.41667 | 0 |
| SITE03.WITH | SITE08 | -0.087 | 0.011 | -8.182 | 0 | -7.90909 | 2.66E-15 |
| SITE03.WITH | SITE09 | -0.105 | 0.012 | -8.647 | 0 | -8.75 | 0 |
| SITE04.WITH | SITE05 | -0.125 | 0.013 | -9.754 | 0 | -9.61538 | 0 |
| SITE04.WITH | SITE06 | -0.119 | 0.012 | -9.584 | 0 | -9.91667 | 0 |
| SITE04.WITH | SITE07 | -0.124 | 0.013 | -9.722 | 0 | -9.53846 | 0 |
| SITE04.WITH | SITE08 | -0.106 | 0.012 | -9.176 | 0 | -8.83333 | 0 |
| SITE04.WITH | SITE09 | -0.128 | 0.013 | -9.848 | 0 | -9.84615 | 0 |
| SITE05.WITH | SITE06 | -0.155 | 0.014 | -10.939 | 0 | -11.0714 | 0 |
| SITE05.WITH | SITE07 | -0.162 | 0.014 | -11.146 | 0 | -11.5714 | 0 |
| SITE05.WITH | SITE08 | -0.139 | 0.013 | -10.342 | 0 | -10.6923 | 0 |
| SITE05.WITH | SITE09 | -0.168 | 0.015 | -11.338 | 0 | -11.2 | 0 |
| SITE06.WITH | SITE07 | -0.154 | 0.014 | -10.893 | 0 | -11 | 0 |
| SITE06.WITH | SITE08 | -0.132 | 0.013 | -10.14 | 0 | -10.1538 | 0 |
| SITE06.WITH | SITE09 | -0.16 | 0.014 | -11.072 | 0 | -11.4286 | 0 |
| SITE07.WITH | SITE08 | -0.138 | 0.013 | -10.303 | 0 | -10.6154 | 0 |
| SITE07.WITH | SITE09 | -0.166 | 0.015 | -11.287 | 0 | -11.0667 | 0 |
| SITE08.WITH | SITE09 | -0.143 | 0.014 | -10.454 | 0 | -10.2143 | 0 |
| Means | CHR | 2.139 | 0.134 | 16.002 | 0 | 15.96269 | 0 |
| Means | AGE | 4.784 | 0.15 | 31.887 | 0 | 31.89333 | 0 |
| Means | FEMALE | 0.91 | 0.044 | 20.763 | 0 | 20.68182 | 0 |
| Means | SITE02 | 0.309 | 0.026 | 11.778 | 0 | 11.88462 | 0 |
| Means | SITE03 | 0.252 | 0.025 | 9.889 | 0 | 10.08 | 0 |
| Means | SITE04 | 0.309 | 0.026 | 11.778 | 0 | 11.88462 | 0 |
| Means | SITE05 | 0.404 | 0.028 | 14.483 | 0 | 14.42857 | 0 |
| Means | SITE06 | 0.384 | 0.028 | 13.971 | 0 | 13.71429 | 0 |
| Means | SITE07 | 0.4 | 0.028 | 14.384 | 0 | 14.28571 | 0 |
| Means | SITE08 | 0.344 | 0.027 | 12.827 | 0 | 12.74074 | 0 |
| Means | SITE09 | 0.415 | 0.028 | 14.776 | 0 | 14.82143 | 0 |
| Intercepts | DEPS_CH | 0.243 | 0.27 | 0.9 | 0.368 | 0.9 | 0.36812 |
| Intercepts | HI_CH | -0.048 | 0.048 | -1.016 | 0.31 | -1 | 0.317311 |
| Variances | CHR | 1 | 0 |  |  |  |  |
| Variances | AGE | 1 | 0 |  |  |  |  |
| Variances | FEMALE | 1 | 0 |  |  |  |  |
| Variances | SITE02 | 1 | 0 |  |  |  |  |
| Variances | SITE03 | 1 | 0 |  |  |  |  |
| Variances | SITE04 | 1 | 0 |  |  |  |  |
| Variances | SITE05 | 1 | 0 |  |  |  |  |
| Variances | SITE06 | 1 | 0 |  |  |  |  |
| Variances | SITE07 | 1 | 0 |  |  |  |  |
| Variances | SITE08 | 1 | 0 |  |  |  |  |
| Variances | SITE09 | 1 | 0 |  |  |  |  |
| Variances | LV_HI | 1 | 0 |  |  |  |  |
| Residual.Variances | DEPS_CH | 0.255 | 0.063 | 4.053 | 0 | 4.047619 | 5.17E-05 |
| Residual.Variances | HI_CH | 0.341 | 0.04 | 8.473 | 0 | 8.525 | 0 |
| Residual.Variances | LV_DEPS | 0.809 | 0.082 | 9.826 | 0 | 9.865854 | 0 |
| LV_HI.BY | HI_CH | 0.812 | 0.025 | 32.754 | 0 | 32.48 | 0 |
| LV_GAF.BY | GAF_CH | 0.91 | 0.013 | 67.629 | 0 | 70 | 0 |
| LV_GAF.ON | LV_HI | -0.028 | 0.053 | -0.517 | 0.605 | -0.5283 | 0.59729 |
| LV_GAF.ON | HIXCHR | 0.161 | 0.048 | 3.325 | 0.001 | 3.354167 | 0.000796 |
| LV_GAF.ON | CHR | 0.125 | 0.035 | 3.563 | 0 | 3.571429 | 0.000355 |
| LV_GAF.ON | AGE | -0.106 | 0.053 | -1.993 | 0.046 | -2 | 0.0455 |
| LV_GAF.ON | FEMALE | -0.001 | 0.052 | -0.025 | 0.98 | -0.01923 | 0.984657 |
| LV_GAF.ON | SITE02 | -0.077 | 0.069 | -1.109 | 0.267 | -1.11594 | 0.264447 |
| LV_GAF.ON | SITE03 | -0.107 | 0.057 | -1.877 | 0.061 | -1.87719 | 0.060492 |
| LV_GAF.ON | SITE04 | -0.044 | 0.061 | -0.717 | 0.473 | -0.72131 | 0.470718 |
| LV_GAF.ON | SITE05 | -0.053 | 0.074 | -0.71 | 0.478 | -0.71622 | 0.473858 |
| LV_GAF.ON | SITE06 | -0.138 | 0.073 | -1.901 | 0.057 | -1.89041 | 0.058703 |
| LV_GAF.ON | SITE07 | -0.137 | 0.087 | -1.577 | 0.115 | -1.57471 | 0.115323 |
| LV_GAF.ON | SITE08 | -0.009 | 0.088 | -0.104 | 0.917 | -0.10227 | 0.91854 |
| LV_GAF.ON | SITE09 | -0.154 | 0.079 | -1.939 | 0.053 | -1.94937 | 0.051252 |
| LV_HI.WITH | CHR | 0.001 | 0.047 | 0.022 | 0.982 | 0.021277 | 0.983025 |
| LV_HI.WITH | AGE | 0.018 | 0.055 | 0.335 | 0.738 | 0.327273 | 0.743462 |
| LV_HI.WITH | FEMALE | -0.09 | 0.06 | -1.503 | 0.133 | -1.5 | 0.133614 |
| LV_HI.WITH | SITE02 | 0.023 | 0.058 | 0.389 | 0.697 | 0.396552 | 0.691698 |
| LV_HI.WITH | SITE03 | -0.011 | 0.085 | -0.129 | 0.897 | -0.12941 | 0.897032 |
| LV_HI.WITH | SITE04 | 0.064 | 0.047 | 1.372 | 0.17 | 1.361702 | 0.173292 |
| LV_HI.WITH | SITE05 | -0.064 | 0.061 | -1.034 | 0.301 | -1.04918 | 0.294095 |
| LV_HI.WITH | SITE06 | 0.068 | 0.045 | 1.509 | 0.131 | 1.511111 | 0.13076 |
| LV_HI.WITH | SITE07 | -0.007 | 0.065 | -0.104 | 0.917 | -0.10769 | 0.91424 |
| LV_HI.WITH | SITE08 | -0.012 | 0.058 | -0.207 | 0.836 | -0.2069 | 0.836091 |
| LV_HI.WITH | SITE09 | -0.043 | 0.059 | -0.732 | 0.464 | -0.72881 | 0.466116 |
| CHR.WITH | AGE | -0.048 | 0.051 | -0.942 | 0.346 | -0.94118 | 0.346614 |
| CHR.WITH | FEMALE | -0.031 | 0.048 | -0.649 | 0.516 | -0.64583 | 0.518387 |
| CHR.WITH | SITE02 | -0.046 | 0.053 | -0.875 | 0.382 | -0.86792 | 0.385436 |
| CHR.WITH | SITE03 | -0.034 | 0.052 | -0.64 | 0.522 | -0.65385 | 0.513211 |
| CHR.WITH | SITE04 | 0.018 | 0.046 | 0.384 | 0.701 | 0.391304 | 0.695572 |
| CHR.WITH | SITE05 | -0.001 | 0.048 | -0.013 | 0.989 | -0.02083 | 0.983379 |
| CHR.WITH | SITE06 | -0.017 | 0.049 | -0.339 | 0.735 | -0.34694 | 0.728637 |
| CHR.WITH | SITE07 | -0.004 | 0.048 | -0.078 | 0.938 | -0.08333 | 0.933586 |
| CHR.WITH | SITE08 | 0.025 | 0.045 | 0.546 | 0.585 | 0.555556 | 0.578515 |
| CHR.WITH | SITE09 | 0.009 | 0.047 | 0.18 | 0.857 | 0.191489 | 0.848142 |
| AGE.WITH | FEMALE | -0.111 | 0.048 | -2.336 | 0.02 | -2.3125 | 0.02075 |
| AGE.WITH | SITE02 | 0.304 | 0.052 | 5.886 | 0 | 5.846154 | 5.03E-09 |
| AGE.WITH | SITE03 | 0 | 0.044 | -0.006 | 0.995 | 0 | 1 |
| AGE.WITH | SITE04 | -0.014 | 0.038 | -0.355 | 0.722 | -0.36842 | 0.712559 |
| AGE.WITH | SITE05 | 0.155 | 0.043 | 3.577 | 0 | 3.604651 | 0.000313 |
| AGE.WITH | SITE06 | -0.01 | 0.04 | -0.237 | 0.813 | -0.25 | 0.802587 |
| AGE.WITH | SITE07 | -0.143 | 0.046 | -3.137 | 0.002 | -3.1087 | 0.001879 |
| AGE.WITH | SITE08 | -0.171 | 0.049 | -3.485 | 0 | -3.4898 | 0.000483 |
| AGE.WITH | SITE09 | -0.03 | 0.044 | -0.691 | 0.49 | -0.68182 | 0.495354 |
| FEMALE.WITH | SITE02 | 0.029 | 0.048 | 0.595 | 0.552 | 0.604167 | 0.545733 |
| FEMALE.WITH | SITE03 | -0.016 | 0.048 | -0.328 | 0.743 | -0.33333 | 0.738883 |
| FEMALE.WITH | SITE04 | -0.004 | 0.048 | -0.085 | 0.932 | -0.08333 | 0.933586 |
| FEMALE.WITH | SITE05 | 0.004 | 0.048 | 0.086 | 0.931 | 0.083333 | 0.933586 |
| FEMALE.WITH | SITE06 | -0.061 | 0.047 | -1.299 | 0.194 | -1.29787 | 0.194331 |
| FEMALE.WITH | SITE07 | 0.064 | 0.048 | 1.326 | 0.185 | 1.333333 | 0.182422 |
| FEMALE.WITH | SITE08 | 0.002 | 0.048 | 0.038 | 0.97 | 0.041667 | 0.966764 |
| FEMALE.WITH | SITE09 | 0.012 | 0.048 | 0.258 | 0.797 | 0.25 | 0.802587 |
| SITE02.WITH | SITE03 | -0.078 | 0.01 | -7.882 | 0 | -7.8 | 6.22E-15 |
| SITE02.WITH | SITE04 | -0.096 | 0.011 | -8.758 | 0 | -8.72727 | 0 |
| SITE02.WITH | SITE05 | -0.125 | 0.013 | -9.754 | 0 | -9.61538 | 0 |
| SITE02.WITH | SITE06 | -0.119 | 0.012 | -9.584 | 0 | -9.91667 | 0 |
| SITE02.WITH | SITE07 | -0.124 | 0.013 | -9.722 | 0 | -9.53846 | 0 |
| SITE02.WITH | SITE08 | -0.107 | 0.012 | -9.176 | 0 | -8.91667 | 0 |
| SITE02.WITH | SITE09 | -0.129 | 0.013 | -9.848 | 0 | -9.92308 | 0 |
| SITE03.WITH | SITE04 | -0.078 | 0.01 | -7.882 | 0 | -7.8 | 6.22E-15 |
| SITE03.WITH | SITE05 | -0.102 | 0.012 | -8.584 | 0 | -8.5 | 0 |
| SITE03.WITH | SITE06 | -0.097 | 0.011 | -8.468 | 0 | -8.81818 | 0 |
| SITE03.WITH | SITE07 | -0.101 | 0.012 | -8.562 | 0 | -8.41667 | 0 |
| SITE03.WITH | SITE08 | -0.087 | 0.011 | -8.182 | 0 | -7.90909 | 2.66E-15 |
| SITE03.WITH | SITE09 | -0.105 | 0.012 | -8.648 | 0 | -8.75 | 0 |
| SITE04.WITH | SITE05 | -0.125 | 0.013 | -9.754 | 0 | -9.61538 | 0 |
| SITE04.WITH | SITE06 | -0.119 | 0.012 | -9.584 | 0 | -9.91667 | 0 |
| SITE04.WITH | SITE07 | -0.124 | 0.013 | -9.722 | 0 | -9.53846 | 0 |
| SITE04.WITH | SITE08 | -0.107 | 0.012 | -9.176 | 0 | -8.91667 | 0 |
| SITE04.WITH | SITE09 | -0.129 | 0.013 | -9.848 | 0 | -9.92308 | 0 |
| SITE05.WITH | SITE06 | -0.156 | 0.014 | -10.94 | 0 | -11.1429 | 0 |
| SITE05.WITH | SITE07 | -0.162 | 0.015 | -11.147 | 0 | -10.8 | 0 |
| SITE05.WITH | SITE08 | -0.139 | 0.013 | -10.343 | 0 | -10.6923 | 0 |
| SITE05.WITH | SITE09 | -0.168 | 0.015 | -11.339 | 0 | -11.2 | 0 |
| SITE06.WITH | SITE07 | -0.154 | 0.014 | -10.894 | 0 | -11 | 0 |
| SITE06.WITH | SITE08 | -0.133 | 0.013 | -10.14 | 0 | -10.2308 | 0 |
| SITE06.WITH | SITE09 | -0.16 | 0.014 | -11.073 | 0 | -11.4286 | 0 |
| SITE07.WITH | SITE08 | -0.138 | 0.013 | -10.304 | 0 | -10.6154 | 0 |
| SITE07.WITH | SITE09 | -0.167 | 0.015 | -11.287 | 0 | -11.1333 | 0 |
| SITE08.WITH | SITE09 | -0.143 | 0.014 | -10.454 | 0 | -10.2143 | 0 |
| Means | CHR | 2.136 | 0.134 | 15.998 | 0 | 15.9403 | 0 |
| Means | AGE | 4.782 | 0.15 | 31.873 | 0 | 31.88 | 0 |
| Means | FEMALE | 0.912 | 0.044 | 20.747 | 0 | 20.72727 | 0 |
| Means | SITE02 | 0.31 | 0.026 | 11.776 | 0 | 11.92308 | 0 |
| Means | SITE03 | 0.252 | 0.026 | 9.888 | 0 | 9.692308 | 0 |
| Means | SITE04 | 0.31 | 0.026 | 11.776 | 0 | 11.92308 | 0 |
| Means | SITE05 | 0.405 | 0.028 | 14.482 | 0 | 14.46429 | 0 |
| Means | SITE06 | 0.385 | 0.028 | 13.967 | 0 | 13.75 | 0 |
| Means | SITE07 | 0.401 | 0.028 | 14.381 | 0 | 14.32143 | 0 |
| Means | SITE08 | 0.344 | 0.027 | 12.826 | 0 | 12.74074 | 0 |
| Means | SITE09 | 0.416 | 0.028 | 14.774 | 0 | 14.85714 | 0 |
| Intercepts | GAF_CH | 0.718 | 0.279 | 2.577 | 0.01 | 2.573477 | 0.010068 |
| Intercepts | HI_CH | -0.049 | 0.048 | -1.027 | 0.305 | -1.02083 | 0.307333 |
| Variances | CHR | 1 | 0 |  |  |  |  |
| Variances | AGE | 1 | 0 |  |  |  |  |
| Variances | FEMALE | 1 | 0 |  |  |  |  |
| Variances | SITE02 | 1 | 0 |  |  |  |  |
| Variances | SITE03 | 1 | 0 |  |  |  |  |
| Variances | SITE04 | 1 | 0 |  |  |  |  |
| Variances | SITE05 | 1 | 0 |  |  |  |  |
| Variances | SITE06 | 1 | 0 |  |  |  |  |
| Variances | SITE07 | 1 | 0 |  |  |  |  |
| Variances | SITE08 | 1 | 0 |  |  |  |  |
| Variances | SITE09 | 1 | 0 |  |  |  |  |
| Variances | LV_HI | 1 | 0 |  |  |  |  |
| Residual.Variances | GAF_CH | 0.173 | 0.024 | 7.054 | 0 | 7.208333 | 5.66E-13 |
| Residual.Variances | HI_CH | 0.341 | 0.04 | 8.456 | 0 | 8.525 | 0 |
| Residual.Variances | LV_GAF | 0.818 | 0.071 | 11.582 | 0 | 11.52113 | 0 |
| LV_HI.BY | HI_CH | 0.81 | 0.025 | 31.882 | 0 | 32.4 | 0 |
| LV_NEGS.BY | NEGS_CH | 0.909 | 0.019 | 47.245 | 0 | 47.84211 | 0 |
| LV_NEGS.ON | LV_HI | 0.067 | 0.055 | 1.218 | 0.223 | 1.218182 | 0.223155 |
| LV_NEGS.ON | HIXCHR | -0.183 | 0.045 | -4.037 | 0 | -4.06667 | 4.77E-05 |
| LV_NEGS.ON | CHR | -0.129 | 0.036 | -3.611 | 0 | -3.58333 | 0.000339 |
| LV_NEGS.ON | AGE | -0.013 | 0.056 | -0.239 | 0.811 | -0.23214 | 0.816427 |
| LV_NEGS.ON | FEMALE | -0.002 | 0.047 | -0.043 | 0.966 | -0.04255 | 0.966058 |
| LV_NEGS.ON | SITE02 | -0.154 | 0.08 | -1.927 | 0.054 | -1.925 | 0.054229 |
| LV_NEGS.ON | SITE03 | 0.062 | 0.067 | 0.935 | 0.35 | 0.925373 | 0.354772 |
| LV_NEGS.ON | SITE04 | -0.117 | 0.066 | -1.778 | 0.075 | -1.77273 | 0.076274 |
| LV_NEGS.ON | SITE05 | -0.04 | 0.08 | -0.503 | 0.615 | -0.5 | 0.617075 |
| LV_NEGS.ON | SITE06 | -0.103 | 0.08 | -1.283 | 0.2 | -1.2875 | 0.19792 |
| LV_NEGS.ON | SITE07 | -0.081 | 0.075 | -1.067 | 0.286 | -1.08 | 0.280142 |
| LV_NEGS.ON | SITE08 | -0.147 | 0.075 | -1.953 | 0.051 | -1.96 | 0.049996 |
| LV_NEGS.ON | SITE09 | 0.031 | 0.091 | 0.346 | 0.729 | 0.340659 | 0.73336 |
| LV_HI.WITH | CHR | -0.004 | 0.048 | -0.091 | 0.927 | -0.08333 | 0.933586 |
| LV_HI.WITH | AGE | 0.024 | 0.057 | 0.417 | 0.677 | 0.421053 | 0.673717 |
| LV_HI.WITH | FEMALE | -0.079 | 0.06 | -1.313 | 0.189 | -1.31667 | 0.18795 |
| LV_HI.WITH | SITE02 | 0.032 | 0.062 | 0.508 | 0.611 | 0.516129 | 0.605764 |
| LV_HI.WITH | SITE03 | -0.054 | 0.079 | -0.685 | 0.493 | -0.68354 | 0.494263 |
| LV_HI.WITH | SITE04 | 0.069 | 0.047 | 1.452 | 0.147 | 1.468085 | 0.142081 |
| LV_HI.WITH | SITE05 | -0.063 | 0.062 | -1.01 | 0.312 | -1.01613 | 0.309568 |
| LV_HI.WITH | SITE06 | 0.079 | 0.045 | 1.74 | 0.082 | 1.755556 | 0.079164 |
| LV_HI.WITH | SITE07 | -0.001 | 0.065 | -0.016 | 0.987 | -0.01538 | 0.987725 |
| LV_HI.WITH | SITE08 | -0.009 | 0.058 | -0.149 | 0.882 | -0.15517 | 0.876685 |
| LV_HI.WITH | SITE09 | -0.041 | 0.059 | -0.69 | 0.49 | -0.69492 | 0.487108 |
| CHR.WITH | AGE | -0.048 | 0.051 | -0.953 | 0.34 | -0.94118 | 0.346614 |
| CHR.WITH | FEMALE | -0.031 | 0.048 | -0.649 | 0.516 | -0.64583 | 0.518387 |
| CHR.WITH | SITE02 | -0.046 | 0.053 | -0.875 | 0.382 | -0.86792 | 0.385436 |
| CHR.WITH | SITE03 | -0.039 | 0.053 | -0.731 | 0.465 | -0.73585 | 0.461823 |
| CHR.WITH | SITE04 | 0.018 | 0.046 | 0.384 | 0.701 | 0.391304 | 0.695572 |
| CHR.WITH | SITE05 | -0.001 | 0.048 | -0.013 | 0.989 | -0.02083 | 0.983379 |
| CHR.WITH | SITE06 | -0.017 | 0.049 | -0.339 | 0.735 | -0.34694 | 0.728637 |
| CHR.WITH | SITE07 | -0.004 | 0.048 | -0.078 | 0.938 | -0.08333 | 0.933586 |
| CHR.WITH | SITE08 | 0.025 | 0.045 | 0.546 | 0.585 | 0.555556 | 0.578515 |
| CHR.WITH | SITE09 | 0.009 | 0.047 | 0.18 | 0.857 | 0.191489 | 0.848142 |
| AGE.WITH | FEMALE | -0.11 | 0.048 | -2.312 | 0.021 | -2.29167 | 0.021925 |
| AGE.WITH | SITE02 | 0.304 | 0.052 | 5.892 | 0 | 5.846154 | 5.03E-09 |
| AGE.WITH | SITE03 | 0.001 | 0.045 | 0.032 | 0.975 | 0.022222 | 0.982271 |
| AGE.WITH | SITE04 | -0.013 | 0.038 | -0.345 | 0.73 | -0.34211 | 0.732272 |
| AGE.WITH | SITE05 | 0.155 | 0.043 | 3.587 | 0 | 3.604651 | 0.000313 |
| AGE.WITH | SITE06 | -0.009 | 0.04 | -0.225 | 0.822 | -0.225 | 0.821979 |
| AGE.WITH | SITE07 | -0.143 | 0.046 | -3.126 | 0.002 | -3.1087 | 0.001879 |
| AGE.WITH | SITE08 | -0.17 | 0.049 | -3.477 | 0.001 | -3.46939 | 0.000522 |
| AGE.WITH | SITE09 | -0.03 | 0.044 | -0.679 | 0.497 | -0.68182 | 0.495354 |
| FEMALE.WITH | SITE02 | 0.029 | 0.048 | 0.595 | 0.552 | 0.604167 | 0.545733 |
| FEMALE.WITH | SITE03 | -0.007 | 0.048 | -0.144 | 0.885 | -0.14583 | 0.884053 |
| FEMALE.WITH | SITE04 | -0.004 | 0.048 | -0.085 | 0.932 | -0.08333 | 0.933586 |
| FEMALE.WITH | SITE05 | 0.004 | 0.048 | 0.086 | 0.931 | 0.083333 | 0.933586 |
| FEMALE.WITH | SITE06 | -0.061 | 0.047 | -1.299 | 0.194 | -1.29787 | 0.194331 |
| FEMALE.WITH | SITE07 | 0.064 | 0.048 | 1.326 | 0.185 | 1.333333 | 0.182422 |
| FEMALE.WITH | SITE08 | 0.002 | 0.048 | 0.038 | 0.97 | 0.041667 | 0.966764 |
| FEMALE.WITH | SITE09 | 0.012 | 0.048 | 0.258 | 0.797 | 0.25 | 0.802587 |
| SITE02.WITH | SITE03 | -0.077 | 0.01 | -7.789 | 0 | -7.7 | 1.35E-14 |
| SITE02.WITH | SITE04 | -0.096 | 0.011 | -8.758 | 0 | -8.72727 | 0 |
| SITE02.WITH | SITE05 | -0.125 | 0.013 | -9.754 | 0 | -9.61538 | 0 |
| SITE02.WITH | SITE06 | -0.119 | 0.012 | -9.584 | 0 | -9.91667 | 0 |
| SITE02.WITH | SITE07 | -0.124 | 0.013 | -9.722 | 0 | -9.53846 | 0 |
| SITE02.WITH | SITE08 | -0.107 | 0.012 | -9.176 | 0 | -8.91667 | 0 |
| SITE02.WITH | SITE09 | -0.129 | 0.013 | -9.848 | 0 | -9.92308 | 0 |
| SITE03.WITH | SITE04 | -0.077 | 0.01 | -7.789 | 0 | -7.7 | 1.35E-14 |
| SITE03.WITH | SITE05 | -0.1 | 0.012 | -8.464 | 0 | -8.33333 | 0 |
| SITE03.WITH | SITE06 | -0.095 | 0.011 | -8.353 | 0 | -8.63636 | 0 |
| SITE03.WITH | SITE07 | -0.099 | 0.012 | -8.443 | 0 | -8.25 | 2.22E-16 |
| SITE03.WITH | SITE08 | -0.085 | 0.011 | -8.078 | 0 | -7.72727 | 1.09E-14 |
| SITE03.WITH | SITE09 | -0.103 | 0.012 | -8.525 | 0 | -8.58333 | 0 |
| SITE04.WITH | SITE05 | -0.125 | 0.013 | -9.754 | 0 | -9.61538 | 0 |
| SITE04.WITH | SITE06 | -0.119 | 0.012 | -9.584 | 0 | -9.91667 | 0 |
| SITE04.WITH | SITE07 | -0.124 | 0.013 | -9.722 | 0 | -9.53846 | 0 |
| SITE04.WITH | SITE08 | -0.107 | 0.012 | -9.176 | 0 | -8.91667 | 0 |
| SITE04.WITH | SITE09 | -0.129 | 0.013 | -9.848 | 0 | -9.92308 | 0 |
| SITE05.WITH | SITE06 | -0.156 | 0.014 | -10.94 | 0 | -11.1429 | 0 |
| SITE05.WITH | SITE07 | -0.162 | 0.015 | -11.147 | 0 | -10.8 | 0 |
| SITE05.WITH | SITE08 | -0.139 | 0.013 | -10.343 | 0 | -10.6923 | 0 |
| SITE05.WITH | SITE09 | -0.168 | 0.015 | -11.339 | 0 | -11.2 | 0 |
| SITE06.WITH | SITE07 | -0.154 | 0.014 | -10.894 | 0 | -11 | 0 |
| SITE06.WITH | SITE08 | -0.133 | 0.013 | -10.14 | 0 | -10.2308 | 0 |
| SITE06.WITH | SITE09 | -0.16 | 0.014 | -11.073 | 0 | -11.4286 | 0 |
| SITE07.WITH | SITE08 | -0.138 | 0.013 | -10.304 | 0 | -10.6154 | 0 |
| SITE07.WITH | SITE09 | -0.167 | 0.015 | -11.287 | 0 | -11.1333 | 0 |
| SITE08.WITH | SITE09 | -0.143 | 0.014 | -10.454 | 0 | -10.2143 | 0 |
| Means | CHR | 2.136 | 0.134 | 15.998 | 0 | 15.9403 | 0 |
| Means | AGE | 4.779 | 0.15 | 31.911 | 0 | 31.86 | 0 |
| Means | FEMALE | 0.912 | 0.044 | 20.743 | 0 | 20.72727 | 0 |
| Means | SITE02 | 0.31 | 0.026 | 11.777 | 0 | 11.92308 | 0 |
| Means | SITE03 | 0.247 | 0.025 | 9.707 | 0 | 9.88 | 0 |
| Means | SITE04 | 0.31 | 0.026 | 11.777 | 0 | 11.92308 | 0 |
| Means | SITE05 | 0.404 | 0.028 | 14.481 | 0 | 14.42857 | 0 |
| Means | SITE06 | 0.385 | 0.028 | 13.968 | 0 | 13.75 | 0 |
| Means | SITE07 | 0.401 | 0.028 | 14.381 | 0 | 14.32143 | 0 |
| Means | SITE08 | 0.344 | 0.027 | 12.826 | 0 | 12.74074 | 0 |
| Means | SITE09 | 0.416 | 0.028 | 14.773 | 0 | 14.85714 | 0 |
| Intercepts | NEGS_CH | 0.251 | 0.303 | 0.83 | 0.407 | 0.828383 | 0.407454 |
| Intercepts | HI_CH | -0.057 | 0.047 | -1.197 | 0.231 | -1.21277 | 0.225219 |
| Variances | CHR | 1 | 0 |  |  |  |  |
| Variances | AGE | 1 | 0 |  |  |  |  |
| Variances | FEMALE | 1 | 0 |  |  |  |  |
| Variances | SITE02 | 1 | 0 |  |  |  |  |
| Variances | SITE03 | 1 | 0 |  |  |  |  |
| Variances | SITE04 | 1 | 0 |  |  |  |  |
| Variances | SITE05 | 1 | 0 |  |  |  |  |
| Variances | SITE06 | 1 | 0 |  |  |  |  |
| Variances | SITE07 | 1 | 0 |  |  |  |  |
| Variances | SITE08 | 1 | 0 |  |  |  |  |
| Variances | SITE09 | 1 | 0 |  |  |  |  |
| Variances | LV_HI | 1 | 0 |  |  |  |  |
| Residual.Variances | NEGS_CH | 0.174 | 0.035 | 4.983 | 0 | 4.971429 | 6.65E-07 |
| Residual.Variances | HI_CH | 0.345 | 0.041 | 8.38 | 0 | 8.414634 | 0 |
| Residual.Variances | LV_NEGS | 0.776 | 0.072 | 10.792 | 0 | 10.77778 | 0 |
| LV_HI.BY | HI_CH | 0.812 | 0.025 | 32.873 | 0 | 32.48 | 0 |
| LV_POSS.BY | POSS_CH | 0.926 | 0.016 | 56.85 | 0 | 57.875 | 0 |
| LV_POSS.ON | LV_HI | -0.08 | 0.048 | -1.67 | 0.095 | -1.66667 | 0.095581 |
| LV_POSS.ON | HIXCHR | -0.042 | 0.059 | -0.711 | 0.477 | -0.71186 | 0.476549 |
| LV_POSS.ON | CHR | -0.261 | 0.033 | -7.804 | 0 | -7.90909 | 2.66E-15 |
| LV_POSS.ON | AGE | 0.003 | 0.054 | 0.054 | 0.957 | 0.055556 | 0.955696 |
| LV_POSS.ON | FEMALE | 0.065 | 0.046 | 1.407 | 0.159 | 1.413043 | 0.157643 |
| LV_POSS.ON | SITE02 | -0.006 | 0.094 | -0.059 | 0.953 | -0.06383 | 0.949106 |
| LV_POSS.ON | SITE03 | 0.055 | 0.059 | 0.931 | 0.352 | 0.932203 | 0.351231 |
| LV_POSS.ON | SITE04 | 0.037 | 0.064 | 0.577 | 0.564 | 0.578125 | 0.56318 |
| LV_POSS.ON | SITE05 | 0.134 | 0.076 | 1.747 | 0.081 | 1.763158 | 0.077874 |
| LV_POSS.ON | SITE06 | 0 | 0.072 | -0.006 | 0.995 | 0 | 1 |
| LV_POSS.ON | SITE07 | 0.043 | 0.069 | 0.623 | 0.533 | 0.623188 | 0.533161 |
| LV_POSS.ON | SITE08 | 0.057 | 0.072 | 0.787 | 0.431 | 0.791667 | 0.428555 |
| LV_POSS.ON | SITE09 | 0.065 | 0.077 | 0.85 | 0.395 | 0.844156 | 0.398582 |
| LV_HI.WITH | CHR | 0 | 0.047 | 0.005 | 0.996 | 0 | 1 |
| LV_HI.WITH | AGE | 0.02 | 0.056 | 0.351 | 0.726 | 0.357143 | 0.720985 |
| LV_HI.WITH | FEMALE | -0.086 | 0.06 | -1.436 | 0.151 | -1.43333 | 0.151763 |
| LV_HI.WITH | SITE02 | 0.017 | 0.059 | 0.296 | 0.767 | 0.288136 | 0.773243 |
| LV_HI.WITH | SITE03 | -0.008 | 0.086 | -0.099 | 0.921 | -0.09302 | 0.925885 |
| LV_HI.WITH | SITE04 | 0.065 | 0.047 | 1.379 | 0.168 | 1.382979 | 0.166671 |
| LV_HI.WITH | SITE05 | -0.061 | 0.061 | -0.994 | 0.32 | -1 | 0.317311 |
| LV_HI.WITH | SITE06 | 0.075 | 0.045 | 1.664 | 0.096 | 1.666667 | 0.095581 |
| LV_HI.WITH | SITE07 | -0.009 | 0.064 | -0.137 | 0.891 | -0.14063 | 0.888166 |
| LV_HI.WITH | SITE08 | -0.014 | 0.058 | -0.246 | 0.805 | -0.24138 | 0.809261 |
| LV_HI.WITH | SITE09 | -0.039 | 0.059 | -0.661 | 0.508 | -0.66102 | 0.508601 |
| CHR.WITH | AGE | -0.049 | 0.051 | -0.956 | 0.339 | -0.96078 | 0.336661 |
| CHR.WITH | FEMALE | -0.032 | 0.048 | -0.67 | 0.503 | -0.66667 | 0.504985 |
| CHR.WITH | SITE02 | -0.046 | 0.053 | -0.881 | 0.378 | -0.86792 | 0.385436 |
| CHR.WITH | SITE03 | -0.034 | 0.052 | -0.645 | 0.519 | -0.65385 | 0.513211 |
| CHR.WITH | SITE04 | 0.017 | 0.046 | 0.377 | 0.706 | 0.369565 | 0.711706 |
| CHR.WITH | SITE05 | -0.001 | 0.048 | -0.022 | 0.982 | -0.02083 | 0.983379 |
| CHR.WITH | SITE06 | -0.017 | 0.049 | -0.347 | 0.728 | -0.34694 | 0.728637 |
| CHR.WITH | SITE07 | -0.004 | 0.048 | -0.087 | 0.931 | -0.08333 | 0.933586 |
| CHR.WITH | SITE08 | 0.024 | 0.045 | 0.538 | 0.591 | 0.533333 | 0.593803 |
| CHR.WITH | SITE09 | 0.008 | 0.047 | 0.17 | 0.865 | 0.170213 | 0.864843 |
| AGE.WITH | FEMALE | -0.11 | 0.048 | -2.305 | 0.021 | -2.29167 | 0.021925 |
| AGE.WITH | SITE02 | 0.304 | 0.052 | 5.894 | 0 | 5.846154 | 5.03E-09 |
| AGE.WITH | SITE03 | 0 | 0.044 | 0.003 | 0.998 | 0 | 1 |
| AGE.WITH | SITE04 | -0.013 | 0.038 | -0.343 | 0.732 | -0.34211 | 0.732272 |
| AGE.WITH | SITE05 | 0.155 | 0.043 | 3.59 | 0 | 3.604651 | 0.000313 |
| AGE.WITH | SITE06 | -0.009 | 0.04 | -0.222 | 0.824 | -0.225 | 0.821979 |
| AGE.WITH | SITE07 | -0.143 | 0.046 | -3.123 | 0.002 | -3.1087 | 0.001879 |
| AGE.WITH | SITE08 | -0.17 | 0.049 | -3.474 | 0.001 | -3.46939 | 0.000522 |
| AGE.WITH | SITE09 | -0.03 | 0.044 | -0.676 | 0.499 | -0.68182 | 0.495354 |
| FEMALE.WITH | SITE02 | 0.029 | 0.048 | 0.608 | 0.543 | 0.604167 | 0.545733 |
| FEMALE.WITH | SITE03 | -0.015 | 0.048 | -0.317 | 0.751 | -0.3125 | 0.754661 |
| FEMALE.WITH | SITE04 | -0.003 | 0.048 | -0.071 | 0.943 | -0.0625 | 0.950165 |
| FEMALE.WITH | SITE05 | 0.005 | 0.048 | 0.104 | 0.917 | 0.104167 | 0.917037 |
| FEMALE.WITH | SITE06 | -0.06 | 0.047 | -1.282 | 0.2 | -1.2766 | 0.201745 |
| FEMALE.WITH | SITE07 | 0.065 | 0.048 | 1.343 | 0.179 | 1.354167 | 0.175683 |
| FEMALE.WITH | SITE08 | 0.003 | 0.048 | 0.053 | 0.958 | 0.0625 | 0.950165 |
| FEMALE.WITH | SITE09 | 0.013 | 0.048 | 0.276 | 0.783 | 0.270833 | 0.786519 |
| SITE02.WITH | SITE03 | -0.078 | 0.01 | -7.882 | 0 | -7.8 | 6.22E-15 |
| SITE02.WITH | SITE04 | -0.096 | 0.011 | -8.758 | 0 | -8.72727 | 0 |
| SITE02.WITH | SITE05 | -0.125 | 0.013 | -9.754 | 0 | -9.61538 | 0 |
| SITE02.WITH | SITE06 | -0.119 | 0.012 | -9.584 | 0 | -9.91667 | 0 |
| SITE02.WITH | SITE07 | -0.124 | 0.013 | -9.722 | 0 | -9.53846 | 0 |
| SITE02.WITH | SITE08 | -0.106 | 0.012 | -9.176 | 0 | -8.83333 | 0 |
| SITE02.WITH | SITE09 | -0.128 | 0.013 | -9.848 | 0 | -9.84615 | 0 |
| SITE03.WITH | SITE04 | -0.078 | 0.01 | -7.882 | 0 | -7.8 | 6.22E-15 |
| SITE03.WITH | SITE05 | -0.102 | 0.012 | -8.584 | 0 | -8.5 | 0 |
| SITE03.WITH | SITE06 | -0.097 | 0.011 | -8.467 | 0 | -8.81818 | 0 |
| SITE03.WITH | SITE07 | -0.101 | 0.012 | -8.562 | 0 | -8.41667 | 0 |
| SITE03.WITH | SITE08 | -0.087 | 0.011 | -8.182 | 0 | -7.90909 | 2.66E-15 |
| SITE03.WITH | SITE09 | -0.105 | 0.012 | -8.647 | 0 | -8.75 | 0 |
| SITE04.WITH | SITE05 | -0.125 | 0.013 | -9.754 | 0 | -9.61538 | 0 |
| SITE04.WITH | SITE06 | -0.119 | 0.012 | -9.584 | 0 | -9.91667 | 0 |
| SITE04.WITH | SITE07 | -0.124 | 0.013 | -9.722 | 0 | -9.53846 | 0 |
| SITE04.WITH | SITE08 | -0.106 | 0.012 | -9.176 | 0 | -8.83333 | 0 |
| SITE04.WITH | SITE09 | -0.128 | 0.013 | -9.848 | 0 | -9.84615 | 0 |
| SITE05.WITH | SITE06 | -0.155 | 0.014 | -10.939 | 0 | -11.0714 | 0 |
| SITE05.WITH | SITE07 | -0.162 | 0.014 | -11.146 | 0 | -11.5714 | 0 |
| SITE05.WITH | SITE08 | -0.139 | 0.013 | -10.342 | 0 | -10.6923 | 0 |
| SITE05.WITH | SITE09 | -0.168 | 0.015 | -11.338 | 0 | -11.2 | 0 |
| SITE06.WITH | SITE07 | -0.154 | 0.014 | -10.893 | 0 | -11 | 0 |
| SITE06.WITH | SITE08 | -0.132 | 0.013 | -10.14 | 0 | -10.1538 | 0 |
| SITE06.WITH | SITE09 | -0.16 | 0.014 | -11.072 | 0 | -11.4286 | 0 |
| SITE07.WITH | SITE08 | -0.138 | 0.013 | -10.303 | 0 | -10.6154 | 0 |
| SITE07.WITH | SITE09 | -0.166 | 0.015 | -11.287 | 0 | -11.0667 | 0 |
| SITE08.WITH | SITE09 | -0.143 | 0.014 | -10.454 | 0 | -10.2143 | 0 |
| Means | CHR | 2.139 | 0.134 | 16.002 | 0 | 15.96269 | 0 |
| Means | AGE | 4.784 | 0.15 | 31.887 | 0 | 31.89333 | 0 |
| Means | FEMALE | 0.91 | 0.044 | 20.761 | 0 | 20.68182 | 0 |
| Means | SITE02 | 0.309 | 0.026 | 11.778 | 0 | 11.88462 | 0 |
| Means | SITE03 | 0.252 | 0.025 | 9.888 | 0 | 10.08 | 0 |
| Means | SITE04 | 0.309 | 0.026 | 11.779 | 0 | 11.88462 | 0 |
| Means | SITE05 | 0.404 | 0.028 | 14.483 | 0 | 14.42857 | 0 |
| Means | SITE06 | 0.385 | 0.028 | 13.971 | 0 | 13.75 | 0 |
| Means | SITE07 | 0.4 | 0.028 | 14.384 | 0 | 14.28571 | 0 |
| Means | SITE08 | 0.344 | 0.027 | 12.827 | 0 | 12.74074 | 0 |
| Means | SITE09 | 0.415 | 0.028 | 14.776 | 0 | 14.82143 | 0 |
| Intercepts | POSS_CH | -0.332 | 0.304 | -1.09 | 0.276 | -1.09211 | 0.274787 |
| Intercepts | HI_CH | -0.048 | 0.048 | -1.002 | 0.316 | -1 | 0.317311 |
| Variances | CHR | 1 | 0 |  |  |  |  |
| Variances | AGE | 1 | 0 |  |  |  |  |
| Variances | FEMALE | 1 | 0 |  |  |  |  |
| Variances | SITE02 | 1 | 0 |  |  |  |  |
| Variances | SITE03 | 1 | 0 |  |  |  |  |
| Variances | SITE04 | 1 | 0 |  |  |  |  |
| Variances | SITE05 | 1 | 0 |  |  |  |  |
| Variances | SITE06 | 1 | 0 |  |  |  |  |
| Variances | SITE07 | 1 | 0 |  |  |  |  |
| Variances | SITE08 | 1 | 0 |  |  |  |  |
| Variances | SITE09 | 1 | 0 |  |  |  |  |
| Variances | LV_HI | 1 | 0 |  |  |  |  |
| Residual.Variances | POSS_CH | 0.143 | 0.03 | 4.733 | 0 | 4.766667 | 1.87E-06 |
| Residual.Variances | HI_CH | 0.34 | 0.04 | 8.468 | 0 | 8.5 | 0 |
| Residual.Variances | LV_POSS | 0.874 | 0.042 | 20.814 | 0 | 20.80952 | 0 |
| LV_HI.BY | HI_CH | 0.813 | 0.025 | 32.98 | 0 | 32.52 | 0 |
| LV_SYCOD.BY | SYCOD_CH | 0.892 | 0.016 | 55.425 | 0 | 55.75 | 0 |
| LV_SYCOD.ON | LV_HI | 0.155 | 0.153 | 1.016 | 0.31 | 1.013072 | 0.311026 |
| LV_SYCOD.ON | HIXCHR | -0.028 | 0.071 | -0.396 | 0.692 | -0.39437 | 0.693311 |
| LV_SYCOD.ON | CHR | -0.055 | 0.049 | -1.122 | 0.262 | -1.12245 | 0.261672 |
| LV_SYCOD.ON | AGE | -0.027 | 0.053 | -0.513 | 0.608 | -0.50943 | 0.610448 |
| LV_SYCOD.ON | FEMALE | -0.078 | 0.057 | -1.383 | 0.167 | -1.36842 | 0.17118 |
| LV_SYCOD.ON | SITE02 | 0.002 | 0.09 | 0.018 | 0.986 | 0.022222 | 0.982271 |
| LV_SYCOD.ON | SITE03 | 0.061 | 0.086 | 0.717 | 0.473 | 0.709302 | 0.478137 |
| LV_SYCOD.ON | SITE04 | -0.146 | 0.059 | -2.476 | 0.013 | -2.47458 | 0.013339 |
| LV_SYCOD.ON | SITE05 | 0.112 | 0.078 | 1.427 | 0.154 | 1.435897 | 0.151032 |
| LV_SYCOD.ON | SITE06 | -0.071 | 0.069 | -1.028 | 0.304 | -1.02899 | 0.303486 |
| LV_SYCOD.ON | SITE07 | -0.07 | 0.068 | -1.026 | 0.305 | -1.02941 | 0.303286 |
| LV_SYCOD.ON | SITE08 | -0.17 | 0.069 | -2.457 | 0.014 | -2.46377 | 0.013749 |
| LV_SYCOD.ON | SITE09 | -0.002 | 0.083 | -0.021 | 0.984 | -0.0241 | 0.980776 |
| LV_HI.WITH | CHR | -0.006 | 0.047 | -0.121 | 0.904 | -0.12766 | 0.898418 |
| LV_HI.WITH | AGE | 0.026 | 0.055 | 0.47 | 0.638 | 0.472727 | 0.636408 |
| LV_HI.WITH | FEMALE | -0.083 | 0.061 | -1.358 | 0.174 | -1.36066 | 0.173623 |
| LV_HI.WITH | SITE02 | 0.025 | 0.059 | 0.424 | 0.672 | 0.423729 | 0.671764 |
| LV_HI.WITH | SITE03 | -0.008 | 0.086 | -0.089 | 0.929 | -0.09302 | 0.925885 |
| LV_HI.WITH | SITE04 | 0.063 | 0.047 | 1.334 | 0.182 | 1.340426 | 0.180107 |
| LV_HI.WITH | SITE05 | -0.059 | 0.062 | -0.951 | 0.342 | -0.95161 | 0.341293 |
| LV_HI.WITH | SITE06 | 0.077 | 0.045 | 1.692 | 0.091 | 1.711111 | 0.087061 |
| LV_HI.WITH | SITE07 | -0.007 | 0.065 | -0.108 | 0.914 | -0.10769 | 0.91424 |
| LV_HI.WITH | SITE08 | -0.012 | 0.058 | -0.205 | 0.838 | -0.2069 | 0.836091 |
| LV_HI.WITH | SITE09 | -0.037 | 0.059 | -0.628 | 0.53 | -0.62712 | 0.530581 |
| CHR.WITH | AGE | -0.049 | 0.051 | -0.961 | 0.337 | -0.96078 | 0.336661 |
| CHR.WITH | FEMALE | -0.033 | 0.048 | -0.685 | 0.493 | -0.6875 | 0.491768 |
| CHR.WITH | SITE02 | -0.03 | 0.051 | -0.578 | 0.563 | -0.58824 | 0.556374 |
| CHR.WITH | SITE03 | -0.034 | 0.053 | -0.649 | 0.517 | -0.64151 | 0.521192 |
| CHR.WITH | SITE04 | 0.014 | 0.047 | 0.291 | 0.771 | 0.297872 | 0.765801 |
| CHR.WITH | SITE05 | -0.004 | 0.049 | -0.092 | 0.927 | -0.08163 | 0.934939 |
| CHR.WITH | SITE06 | -0.018 | 0.05 | -0.352 | 0.724 | -0.36 | 0.718847 |
| CHR.WITH | SITE07 | -0.004 | 0.049 | -0.092 | 0.927 | -0.08163 | 0.934939 |
| CHR.WITH | SITE08 | 0.024 | 0.046 | 0.533 | 0.594 | 0.521739 | 0.601852 |
| CHR.WITH | SITE09 | 0.008 | 0.048 | 0.165 | 0.869 | 0.166667 | 0.867632 |
| AGE.WITH | FEMALE | -0.11 | 0.048 | -2.305 | 0.021 | -2.29167 | 0.021925 |
| AGE.WITH | SITE02 | 0.31 | 0.052 | 5.938 | 0 | 5.961538 | 2.5E-09 |
| AGE.WITH | SITE03 | 0 | 0.045 | 0.007 | 0.995 | 0 | 1 |
| AGE.WITH | SITE04 | -0.01 | 0.039 | -0.256 | 0.798 | -0.25641 | 0.797634 |
| AGE.WITH | SITE05 | 0.151 | 0.044 | 3.466 | 0.001 | 3.431818 | 0.0006 |
| AGE.WITH | SITE06 | -0.009 | 0.041 | -0.216 | 0.829 | -0.21951 | 0.826251 |
| AGE.WITH | SITE07 | -0.143 | 0.046 | -3.122 | 0.002 | -3.1087 | 0.001879 |
| AGE.WITH | SITE08 | -0.171 | 0.049 | -3.474 | 0.001 | -3.4898 | 0.000483 |
| AGE.WITH | SITE09 | -0.03 | 0.044 | -0.67 | 0.503 | -0.68182 | 0.495354 |
| FEMALE.WITH | SITE02 | 0.034 | 0.048 | 0.704 | 0.481 | 0.708333 | 0.478738 |
| FEMALE.WITH | SITE03 | -0.018 | 0.048 | -0.373 | 0.709 | -0.375 | 0.70766 |
| FEMALE.WITH | SITE04 | 0.001 | 0.048 | 0.017 | 0.987 | 0.020833 | 0.983379 |
| FEMALE.WITH | SITE05 | 0.007 | 0.048 | 0.143 | 0.886 | 0.145833 | 0.884053 |
| FEMALE.WITH | SITE06 | -0.065 | 0.047 | -1.369 | 0.171 | -1.38298 | 0.166671 |
| FEMALE.WITH | SITE07 | 0.061 | 0.048 | 1.257 | 0.209 | 1.270833 | 0.203788 |
| FEMALE.WITH | SITE08 | -0.001 | 0.048 | -0.023 | 0.981 | -0.02083 | 0.983379 |
| FEMALE.WITH | SITE09 | 0.009 | 0.048 | 0.184 | 0.854 | 0.1875 | 0.851269 |
| SITE02.WITH | SITE03 | -0.078 | 0.01 | -7.839 | 0 | -7.8 | 6.22E-15 |
| SITE02.WITH | SITE04 | -0.094 | 0.011 | -8.641 | 0 | -8.54545 | 0 |
| SITE02.WITH | SITE05 | -0.124 | 0.013 | -9.642 | 0 | -9.53846 | 0 |
| SITE02.WITH | SITE06 | -0.119 | 0.012 | -9.507 | 0 | -9.91667 | 0 |
| SITE02.WITH | SITE07 | -0.124 | 0.013 | -9.642 | 0 | -9.53846 | 0 |
| SITE02.WITH | SITE08 | -0.106 | 0.012 | -9.108 | 0 | -8.83333 | 0 |
| SITE02.WITH | SITE09 | -0.128 | 0.013 | -9.765 | 0 | -9.84615 | 0 |
| SITE03.WITH | SITE04 | -0.078 | 0.01 | -7.839 | 0 | -7.8 | 6.22E-15 |
| SITE03.WITH | SITE05 | -0.102 | 0.012 | -8.563 | 0 | -8.5 | 0 |
| SITE03.WITH | SITE06 | -0.098 | 0.012 | -8.468 | 0 | -8.16667 | 2.22E-16 |
| SITE03.WITH | SITE07 | -0.102 | 0.012 | -8.563 | 0 | -8.5 | 0 |
| SITE03.WITH | SITE08 | -0.088 | 0.011 | -8.183 | 0 | -8 | 1.33E-15 |
| SITE03.WITH | SITE09 | -0.106 | 0.012 | -8.649 | 0 | -8.83333 | 0 |
| SITE04.WITH | SITE05 | -0.124 | 0.013 | -9.642 | 0 | -9.53846 | 0 |
| SITE04.WITH | SITE06 | -0.119 | 0.012 | -9.507 | 0 | -9.91667 | 0 |
| SITE04.WITH | SITE07 | -0.124 | 0.013 | -9.642 | 0 | -9.53846 | 0 |
| SITE04.WITH | SITE08 | -0.106 | 0.012 | -9.108 | 0 | -8.83333 | 0 |
| SITE04.WITH | SITE09 | -0.128 | 0.013 | -9.765 | 0 | -9.84615 | 0 |
| SITE05.WITH | SITE06 | -0.156 | 0.014 | -10.897 | 0 | -11.1429 | 0 |
| SITE05.WITH | SITE07 | -0.162 | 0.015 | -11.101 | 0 | -10.8 | 0 |
| SITE05.WITH | SITE08 | -0.139 | 0.014 | -10.306 | 0 | -9.92857 | 0 |
| SITE05.WITH | SITE09 | -0.168 | 0.015 | -11.291 | 0 | -11.2 | 0 |
| SITE06.WITH | SITE07 | -0.156 | 0.014 | -10.897 | 0 | -11.1429 | 0 |
| SITE06.WITH | SITE08 | -0.134 | 0.013 | -10.142 | 0 | -10.3077 | 0 |
| SITE06.WITH | SITE09 | -0.162 | 0.015 | -11.076 | 0 | -10.8 | 0 |
| SITE07.WITH | SITE08 | -0.139 | 0.014 | -10.306 | 0 | -9.92857 | 0 |
| SITE07.WITH | SITE09 | -0.168 | 0.015 | -11.291 | 0 | -11.2 | 0 |
| SITE08.WITH | SITE09 | -0.145 | 0.014 | -10.457 | 0 | -10.3571 | 0 |
| Means | CHR | 2.141 | 0.135 | 15.901 | 0 | 15.85926 | 0 |
| Means | AGE | 4.786 | 0.151 | 31.627 | 0 | 31.69536 | 0 |
| Means | FEMALE | 0.919 | 0.044 | 20.662 | 0 | 20.88636 | 0 |
| Means | SITE02 | 0.307 | 0.026 | 11.63 | 0 | 11.80769 | 0 |
| Means | SITE03 | 0.254 | 0.026 | 9.885 | 0 | 9.769231 | 0 |
| Means | SITE04 | 0.307 | 0.026 | 11.631 | 0 | 11.80769 | 0 |
| Means | SITE05 | 0.403 | 0.028 | 14.37 | 0 | 14.39286 | 0 |
| Means | SITE06 | 0.387 | 0.028 | 13.959 | 0 | 13.82143 | 0 |
| Means | SITE07 | 0.403 | 0.028 | 14.37 | 0 | 14.39286 | 0 |
| Means | SITE08 | 0.346 | 0.027 | 12.818 | 0 | 12.81481 | 0 |
| Means | SITE09 | 0.418 | 0.028 | 14.761 | 0 | 14.92857 | 0 |
| Intercepts | SYCOD_CH | 1.113 | 0.32 | 3.475 | 0.001 | 3.478125 | 0.000505 |
| Intercepts | HI_CH | -0.052 | 0.048 | -1.096 | 0.273 | -1.08333 | 0.27866 |
| Variances | CHR | 1 | 0 |  |  |  |  |
| Variances | AGE | 1 | 0 |  |  |  |  |
| Variances | FEMALE | 1 | 0 |  |  |  |  |
| Variances | SITE02 | 1 | 0 |  |  |  |  |
| Variances | SITE03 | 1 | 0 |  |  |  |  |
| Variances | SITE04 | 1 | 0 |  |  |  |  |
| Variances | SITE05 | 1 | 0 |  |  |  |  |
| Variances | SITE06 | 1 | 0 |  |  |  |  |
| Variances | SITE07 | 1 | 0 |  |  |  |  |
| Variances | SITE08 | 1 | 0 |  |  |  |  |
| Variances | SITE09 | 1 | 0 |  |  |  |  |
| Variances | LV_HI | 1 | 0 |  |  |  |  |
| Residual.Variances | SYCOD_CH | 0.204 | 0.029 | 7.085 | 0 | 7.034483 | 2E-12 |
| Residual.Variances | HI_CH | 0.338 | 0.04 | 8.43 | 0 | 8.45 | 0 |
| Residual.Variances | LV_SYCOD | 0.91 | 0.028 | 32.897 | 0 | 32.5 | 0 |
| LV_HI.BY | HI_CH | 0.814 | 0.025 | 33.166 | 0 | 32.56 | 0 |
| LV_VEMEM.BY | VEMEM_CH | 0.74 | 0.043 | 17.317 | 0 | 17.2093 | 0 |
| LV_VEMEM.ON | LV_HI | 0.098 | 0.094 | 1.044 | 0.296 | 1.042553 | 0.297155 |
| LV_VEMEM.ON | HIXCHR | -0.101 | 0.054 | -1.868 | 0.062 | -1.87037 | 0.061432 |
| LV_VEMEM.ON | CHR | -0.05 | 0.049 | -1.033 | 0.302 | -1.02041 | 0.307535 |
| LV_VEMEM.ON | AGE | 0.036 | 0.062 | 0.578 | 0.563 | 0.580645 | 0.56148 |
| LV_VEMEM.ON | FEMALE | -0.103 | 0.061 | -1.674 | 0.094 | -1.68852 | 0.091311 |
| LV_VEMEM.ON | SITE02 | 0.039 | 0.089 | 0.441 | 0.659 | 0.438202 | 0.66124 |
| LV_VEMEM.ON | SITE03 | 0.004 | 0.07 | 0.052 | 0.959 | 0.057143 | 0.954431 |
| LV_VEMEM.ON | SITE04 | -0.171 | 0.098 | -1.75 | 0.08 | -1.7449 | 0.081003 |
| LV_VEMEM.ON | SITE05 | 0.036 | 0.081 | 0.449 | 0.654 | 0.444444 | 0.656721 |
| LV_VEMEM.ON | SITE06 | 0.122 | 0.089 | 1.375 | 0.169 | 1.370787 | 0.170442 |
| LV_VEMEM.ON | SITE07 | 0.075 | 0.077 | 0.972 | 0.331 | 0.974026 | 0.330044 |
| LV_VEMEM.ON | SITE08 | 0.058 | 0.088 | 0.659 | 0.51 | 0.659091 | 0.509837 |
| LV_VEMEM.ON | SITE09 | 0.138 | 0.092 | 1.499 | 0.134 | 1.5 | 0.133614 |
| LV_HI.WITH | CHR | -0.004 | 0.047 | -0.077 | 0.939 | -0.08511 | 0.932177 |
| LV_HI.WITH | AGE | 0.026 | 0.056 | 0.459 | 0.646 | 0.464286 | 0.642443 |
| LV_HI.WITH | FEMALE | -0.076 | 0.06 | -1.264 | 0.206 | -1.26667 | 0.205275 |
| LV_HI.WITH | SITE02 | 0.022 | 0.058 | 0.38 | 0.704 | 0.37931 | 0.704457 |
| LV_HI.WITH | SITE03 | -0.005 | 0.086 | -0.063 | 0.95 | -0.05814 | 0.953637 |
| LV_HI.WITH | SITE04 | 0.066 | 0.048 | 1.395 | 0.163 | 1.375 | 0.169131 |
| LV_HI.WITH | SITE05 | -0.059 | 0.062 | -0.949 | 0.343 | -0.95161 | 0.341293 |
| LV_HI.WITH | SITE06 | 0.077 | 0.045 | 1.706 | 0.088 | 1.711111 | 0.087061 |
| LV_HI.WITH | SITE07 | -0.006 | 0.065 | -0.088 | 0.93 | -0.09231 | 0.926454 |
| LV_HI.WITH | SITE08 | -0.012 | 0.058 | -0.213 | 0.831 | -0.2069 | 0.836091 |
| LV_HI.WITH | SITE09 | -0.042 | 0.059 | -0.708 | 0.479 | -0.71186 | 0.476549 |
| CHR.WITH | AGE | -0.049 | 0.051 | -0.954 | 0.34 | -0.96078 | 0.336661 |
| CHR.WITH | FEMALE | -0.027 | 0.048 | -0.567 | 0.57 | -0.5625 | 0.573775 |
| CHR.WITH | SITE02 | -0.045 | 0.053 | -0.849 | 0.396 | -0.84906 | 0.39585 |
| CHR.WITH | SITE03 | -0.033 | 0.052 | -0.619 | 0.536 | -0.63462 | 0.525679 |
| CHR.WITH | SITE04 | 0.015 | 0.046 | 0.331 | 0.741 | 0.326087 | 0.744359 |
| CHR.WITH | SITE05 | -0.002 | 0.048 | -0.042 | 0.967 | -0.04167 | 0.966764 |
| CHR.WITH | SITE06 | -0.015 | 0.049 | -0.305 | 0.76 | -0.30612 | 0.759511 |
| CHR.WITH | SITE07 | -0.002 | 0.048 | -0.042 | 0.967 | -0.04167 | 0.966764 |
| CHR.WITH | SITE08 | 0.026 | 0.045 | 0.579 | 0.563 | 0.577778 | 0.563414 |
| CHR.WITH | SITE09 | 0.007 | 0.048 | 0.153 | 0.878 | 0.145833 | 0.884053 |
| AGE.WITH | FEMALE | -0.111 | 0.048 | -2.328 | 0.02 | -2.3125 | 0.02075 |
| AGE.WITH | SITE02 | 0.306 | 0.052 | 5.903 | 0 | 5.884615 | 3.99E-09 |
| AGE.WITH | SITE03 | 0 | 0.045 | 0 | 1 | 0 | 1 |
| AGE.WITH | SITE04 | -0.01 | 0.039 | -0.265 | 0.791 | -0.25641 | 0.797634 |
| AGE.WITH | SITE05 | 0.151 | 0.044 | 3.456 | 0.001 | 3.431818 | 0.0006 |
| AGE.WITH | SITE06 | -0.009 | 0.041 | -0.227 | 0.82 | -0.21951 | 0.826251 |
| AGE.WITH | SITE07 | -0.144 | 0.046 | -3.132 | 0.002 | -3.13043 | 0.001745 |
| AGE.WITH | SITE08 | -0.172 | 0.049 | -3.482 | 0 | -3.5102 | 0.000448 |
| AGE.WITH | SITE09 | -0.027 | 0.045 | -0.613 | 0.54 | -0.6 | 0.548506 |
| FEMALE.WITH | SITE02 | 0.026 | 0.048 | 0.54 | 0.589 | 0.541667 | 0.588048 |
| FEMALE.WITH | SITE03 | -0.018 | 0.048 | -0.373 | 0.709 | -0.375 | 0.70766 |
| FEMALE.WITH | SITE04 | 0.001 | 0.048 | 0.017 | 0.987 | 0.020833 | 0.983379 |
| FEMALE.WITH | SITE05 | 0.007 | 0.048 | 0.143 | 0.886 | 0.145833 | 0.884053 |
| FEMALE.WITH | SITE06 | -0.065 | 0.047 | -1.369 | 0.171 | -1.38298 | 0.166671 |
| FEMALE.WITH | SITE07 | 0.061 | 0.048 | 1.257 | 0.209 | 1.270833 | 0.203788 |
| FEMALE.WITH | SITE08 | -0.001 | 0.048 | -0.023 | 0.981 | -0.02083 | 0.983379 |
| FEMALE.WITH | SITE09 | 0.015 | 0.048 | 0.311 | 0.756 | 0.3125 | 0.754661 |
| SITE02.WITH | SITE03 | -0.079 | 0.01 | -7.883 | 0 | -7.9 | 2.89E-15 |
| SITE02.WITH | SITE04 | -0.096 | 0.011 | -8.699 | 0 | -8.72727 | 0 |
| SITE02.WITH | SITE05 | -0.125 | 0.013 | -9.724 | 0 | -9.61538 | 0 |
| SITE02.WITH | SITE06 | -0.12 | 0.013 | -9.586 | 0 | -9.23077 | 0 |
| SITE02.WITH | SITE07 | -0.125 | 0.013 | -9.724 | 0 | -9.61538 | 0 |
| SITE02.WITH | SITE08 | -0.108 | 0.012 | -9.177 | 0 | -9 | 0 |
| SITE02.WITH | SITE09 | -0.129 | 0.013 | -9.819 | 0 | -9.92308 | 0 |
| SITE03.WITH | SITE04 | -0.078 | 0.01 | -7.839 | 0 | -7.8 | 6.22E-15 |
| SITE03.WITH | SITE05 | -0.102 | 0.012 | -8.563 | 0 | -8.5 | 0 |
| SITE03.WITH | SITE06 | -0.098 | 0.012 | -8.468 | 0 | -8.16667 | 2.22E-16 |
| SITE03.WITH | SITE07 | -0.102 | 0.012 | -8.563 | 0 | -8.5 | 0 |
| SITE03.WITH | SITE08 | -0.088 | 0.011 | -8.183 | 0 | -8 | 1.33E-15 |
| SITE03.WITH | SITE09 | -0.105 | 0.012 | -8.628 | 0 | -8.75 | 0 |
| SITE04.WITH | SITE05 | -0.124 | 0.013 | -9.642 | 0 | -9.53846 | 0 |
| SITE04.WITH | SITE06 | -0.119 | 0.012 | -9.507 | 0 | -9.91667 | 0 |
| SITE04.WITH | SITE07 | -0.124 | 0.013 | -9.642 | 0 | -9.53846 | 0 |
| SITE04.WITH | SITE08 | -0.106 | 0.012 | -9.108 | 0 | -8.83333 | 0 |
| SITE04.WITH | SITE09 | -0.127 | 0.013 | -9.735 | 0 | -9.76923 | 0 |
| SITE05.WITH | SITE06 | -0.156 | 0.014 | -10.897 | 0 | -11.1429 | 0 |
| SITE05.WITH | SITE07 | -0.162 | 0.015 | -11.101 | 0 | -10.8 | 0 |
| SITE05.WITH | SITE08 | -0.139 | 0.014 | -10.306 | 0 | -9.92857 | 0 |
| SITE05.WITH | SITE09 | -0.167 | 0.015 | -11.245 | 0 | -11.1333 | 0 |
| SITE06.WITH | SITE07 | -0.156 | 0.014 | -10.897 | 0 | -11.1429 | 0 |
| SITE06.WITH | SITE08 | -0.134 | 0.013 | -10.142 | 0 | -10.3077 | 0 |
| SITE06.WITH | SITE09 | -0.16 | 0.015 | -11.033 | 0 | -10.6667 | 0 |
| SITE07.WITH | SITE08 | -0.139 | 0.014 | -10.306 | 0 | -9.92857 | 0 |
| SITE07.WITH | SITE09 | -0.167 | 0.015 | -11.245 | 0 | -11.1333 | 0 |
| SITE08.WITH | SITE09 | -0.143 | 0.014 | -10.42 | 0 | -10.2143 | 0 |
| Means | CHR | 2.124 | 0.133 | 15.981 | 0 | 15.96992 | 0 |
| Means | AGE | 4.789 | 0.152 | 31.605 | 0 | 31.50658 | 0 |
| Means | FEMALE | 0.919 | 0.044 | 20.662 | 0 | 20.88636 | 0 |
| Means | SITE02 | 0.311 | 0.026 | 11.772 | 0 | 11.96154 | 0 |
| Means | SITE03 | 0.254 | 0.026 | 9.885 | 0 | 9.769231 | 0 |
| Means | SITE04 | 0.307 | 0.026 | 11.631 | 0 | 11.80769 | 0 |
| Means | SITE05 | 0.403 | 0.028 | 14.37 | 0 | 14.39286 | 0 |
| Means | SITE06 | 0.387 | 0.028 | 13.959 | 0 | 13.82143 | 0 |
| Means | SITE07 | 0.403 | 0.028 | 14.37 | 0 | 14.39286 | 0 |
| Means | SITE08 | 0.346 | 0.027 | 12.819 | 0 | 12.81481 | 0 |
| Means | SITE09 | 0.414 | 0.028 | 14.665 | 0 | 14.78571 | 0 |
| Intercepts | VEMEM_CH | 0.258 | 0.303 | 0.851 | 0.395 | 0.851485 | 0.3945 |
| Intercepts | HI_CH | -0.055 | 0.048 | -1.145 | 0.252 | -1.14583 | 0.251864 |
| Variances | CHR | 1 | 0 |  |  |  |  |
| Variances | AGE | 1 | 0 |  |  |  |  |
| Variances | FEMALE | 1 | 0 |  |  |  |  |
| Variances | SITE02 | 1 | 0 |  |  |  |  |
| Variances | SITE03 | 1 | 0 |  |  |  |  |
| Variances | SITE04 | 1 | 0 |  |  |  |  |
| Variances | SITE05 | 1 | 0 |  |  |  |  |
| Variances | SITE06 | 1 | 0 |  |  |  |  |
| Variances | SITE07 | 1 | 0 |  |  |  |  |
| Variances | SITE08 | 1 | 0 |  |  |  |  |
| Variances | SITE09 | 1 | 0 |  |  |  |  |
| Variances | LV_HI | 1 | 0 |  |  |  |  |
| Residual.Variances | VEMEM_CH | 0.453 | 0.063 | 7.169 | 0 | 7.190476 | 6.46E-13 |
| Residual.Variances | HI_CH | 0.337 | 0.04 | 8.43 | 0 | 8.425 | 0 |
| Residual.Variances | LV_VEMEM | 0.893 | 0.045 | 20.033 | 0 | 19.84444 | 0 |

Table_S2b_MainAnalysisStandardized_InteractionsCorrected

| **paramHeader** | **param** | **est** | **se** | **est_se** | **pval** | **zval** | **p_exact** | **p_bonf** |
| --- | --- | --- | --- | --- | --- | --- | --- | --- |
| LV_DEPS.ON | HIXCHR | -0.15 | 0.053 | -2.811 | 0.005 | -2.83019 | 0.004652 | 0.027912 |
| LV_GAF.ON | HIXCHR | 0.161 | 0.048 | 3.325 | 0.001 | 3.354167 | 0.000796 | 0.004776 |
| LV_NEGS.ON | HIXCHR | -0.183 | 0.045 | -4.037 | 0 | -4.06667 | 4.77E-05 | 0.000286 |
| LV_POSS.ON | HIXCHR | -0.042 | 0.059 | -0.711 | 0.477 | -0.71186 | 0.476549 | 1 |
| LV_SYCOD.ON | HIXCHR | -0.028 | 0.071 | -0.396 | 0.692 | -0.39437 | 0.693311 | 1 |
| LV_VEMEM.ON | HIXCHR | -0.101 | 0.054 | -1.868 | 0.062 | -1.87037 | 0.061432 | 0.368594 |

Table_S2c_MainAnalysis_ModelSummary

| **behav** | **neuro** | **estimator** | **n_obs** | **n_parameters** | **aic** | **bic** |
| --- | --- | --- | --- | --- | --- | --- |
| PosS | fchi | MLR | 435 | 105 | 3819.273 | 4247.184 |
| NegS | fchi | MLR | 434 | 105 | 4100.172 | 4527.841 |
| DepS | fchi | MLR | 435 | 105 | 3979.92 | 4407.832 |
| gaf | fchi | MLR | 434 | 105 | 4794.846 | 5222.516 |
| SyCod | fchi | MLR | 430 | 105 | 4734.482 | 5161.179 |
| VeMem | fchi | MLR | 430 | 105 | 3892.97 | 4319.667 |

Table_S3_MainAnalysisUnstandardized

| **paramHeader** | **param** | **est** | **se** | **est_se** | **pval** | **zval** | **pval_exact** |
| --- | --- | --- | --- | --- | --- | --- | --- |
| LV_HI.BY | HI_CH | 1 | 0 |  |  |  |  |
| LV_DEPS.BY | DEPS_CH | 1 | 0 |  |  |  |  |
| LV_DEPS.ON | LV_HI | 0.103 | 1.005 | 0.103 | 0.918 | 0.102488 | 0.91837 |
| LV_DEPS.ON | HIXCHR | -10.949 | 4.964 | -2.206 | 0.027 | -2.20568 | 0.027406 |
| LV_DEPS.ON | CHR | -0.281 | 0.058 | -4.874 | 0 | -4.84483 | 1.27E-06 |
| LV_DEPS.ON | AGE | -0.017 | 0.009 | -1.745 | 0.081 | -1.88889 | 0.058907 |
| LV_DEPS.ON | FEMALE | -0.042 | 0.074 | -0.574 | 0.566 | -0.56757 | 0.570329 |
| LV_DEPS.ON | SITE02 | -0.085 | 0.25 | -0.339 | 0.734 | -0.34 | 0.733857 |
| LV_DEPS.ON | SITE03 | 0.34 | 0.19 | 1.792 | 0.073 | 1.789474 | 0.073539 |
| LV_DEPS.ON | SITE04 | 0.097 | 0.17 | 0.569 | 0.57 | 0.570588 | 0.568279 |
| LV_DEPS.ON | SITE05 | 0.223 | 0.15 | 1.492 | 0.136 | 1.486667 | 0.137103 |
| LV_DEPS.ON | SITE06 | 0.097 | 0.165 | 0.59 | 0.555 | 0.587879 | 0.556614 |
| LV_DEPS.ON | SITE07 | 0.132 | 0.149 | 0.885 | 0.376 | 0.885906 | 0.375668 |
| LV_DEPS.ON | SITE08 | 0.07 | 0.176 | 0.395 | 0.693 | 0.397727 | 0.690831 |
| LV_DEPS.ON | SITE09 | 0.324 | 0.167 | 1.945 | 0.052 | 1.94012 | 0.052365 |
| LV_HI.WITH | CHR | 0 | 0 | 0.005 | 0.996 |  |  |
| LV_HI.WITH | AGE | 0.002 | 0.006 | 0.386 | 0.7 | 0.333333 | 0.738883 |
| LV_HI.WITH | FEMALE | -0.001 | 0.001 | -1.442 | 0.149 | -1 | 0.317311 |
| LV_HI.WITH | SITE02 | 0 | 0 | 0.41 | 0.682 |  |  |
| LV_HI.WITH | SITE03 | 0 | 0.001 | -0.132 | 0.895 | 0 | 1 |
| LV_HI.WITH | SITE04 | 0 | 0 | 1.372 | 0.17 |  |  |
| LV_HI.WITH | SITE05 | -0.001 | 0.001 | -0.983 | 0.326 | -1 | 0.317311 |
| LV_HI.WITH | SITE06 | 0.001 | 0 | 1.614 | 0.107 |  |  |
| LV_HI.WITH | SITE07 | 0 | 0.001 | -0.152 | 0.879 | 0 | 1 |
| LV_HI.WITH | SITE08 | 0 | 0 | -0.224 | 0.823 |  |  |
| LV_HI.WITH | SITE09 | 0 | 0.001 | -0.72 | 0.471 | 0 | 1 |
| CHR.WITH | AGE | -0.074 | 0.078 | -0.953 | 0.34 | -0.94872 | 0.342764 |
| CHR.WITH | FEMALE | -0.006 | 0.009 | -0.669 | 0.504 | -0.66667 | 0.504985 |
| CHR.WITH | SITE02 | -0.005 | 0.006 | -0.875 | 0.382 | -0.83333 | 0.404657 |
| CHR.WITH | SITE03 | -0.003 | 0.005 | -0.642 | 0.521 | -0.6 | 0.548506 |
| CHR.WITH | SITE04 | 0.002 | 0.005 | 0.377 | 0.707 | 0.4 | 0.689157 |
| CHR.WITH | SITE05 | 0 | 0.006 | -0.022 | 0.982 | 0 | 1 |
| CHR.WITH | SITE06 | -0.002 | 0.006 | -0.347 | 0.729 | -0.33333 | 0.738883 |
| CHR.WITH | SITE07 | -0.001 | 0.006 | -0.087 | 0.931 | -0.16667 | 0.867632 |
| CHR.WITH | SITE08 | 0.003 | 0.005 | 0.536 | 0.592 | 0.6 | 0.548506 |
| CHR.WITH | SITE09 | 0.001 | 0.006 | 0.17 | 0.865 | 0.166667 | 0.867632 |
| AGE.WITH | FEMALE | -0.218 | 0.094 | -2.309 | 0.021 | -2.31915 | 0.020387 |
| AGE.WITH | SITE02 | 0.342 | 0.076 | 4.51 | 0 | 4.5 | 6.8E-06 |
| AGE.WITH | SITE03 | 0 | 0.042 | 0.003 | 0.998 | 0 | 1 |
| AGE.WITH | SITE04 | -0.015 | 0.043 | -0.342 | 0.732 | -0.34884 | 0.727212 |
| AGE.WITH | SITE05 | 0.215 | 0.062 | 3.463 | 0.001 | 3.467742 | 0.000525 |
| AGE.WITH | SITE06 | -0.012 | 0.054 | -0.222 | 0.824 | -0.22222 | 0.824141 |
| AGE.WITH | SITE07 | -0.196 | 0.066 | -2.986 | 0.003 | -2.9697 | 0.002981 |
| AGE.WITH | SITE08 | -0.208 | 0.064 | -3.238 | 0.001 | -3.25 | 0.001154 |
| AGE.WITH | SITE09 | -0.042 | 0.062 | -0.676 | 0.499 | -0.67742 | 0.49814 |
| FEMALE.WITH | SITE02 | 0.004 | 0.007 | 0.606 | 0.544 | 0.571429 | 0.567709 |
| FEMALE.WITH | SITE03 | -0.002 | 0.006 | -0.316 | 0.752 | -0.33333 | 0.738883 |
| FEMALE.WITH | SITE04 | 0 | 0.007 | -0.071 | 0.943 | 0 | 1 |
| FEMALE.WITH | SITE05 | 0.001 | 0.008 | 0.104 | 0.917 | 0.125 | 0.900524 |
| FEMALE.WITH | SITE06 | -0.01 | 0.008 | -1.273 | 0.203 | -1.25 | 0.2113 |
| FEMALE.WITH | SITE07 | 0.011 | 0.008 | 1.334 | 0.182 | 1.375 | 0.169131 |
| FEMALE.WITH | SITE08 | 0 | 0.007 | 0.053 | 0.958 | 0 | 1 |
| FEMALE.WITH | SITE09 | 0.002 | 0.008 | 0.276 | 0.783 | 0.25 | 0.802587 |
| SITE02.WITH | SITE03 | -0.005 | 0.001 | -4.242 | 0 | -5 | 5.73E-07 |
| SITE02.WITH | SITE04 | -0.008 | 0.002 | -4.798 | 0 | -4 | 6.33E-05 |
| SITE02.WITH | SITE05 | -0.012 | 0.002 | -5.462 | 0 | -6 | 1.97E-09 |
| SITE02.WITH | SITE06 | -0.011 | 0.002 | -5.347 | 0 | -5.5 | 3.8E-08 |
| SITE02.WITH | SITE07 | -0.012 | 0.002 | -5.44 | 0 | -6 | 1.97E-09 |
| SITE02.WITH | SITE08 | -0.009 | 0.002 | -5.073 | 0 | -4.5 | 6.8E-06 |
| SITE02.WITH | SITE09 | -0.013 | 0.002 | -5.526 | 0 | -6.5 | 8.03E-11 |
| SITE03.WITH | SITE04 | -0.005 | 0.001 | -4.242 | 0 | -5 | 5.73E-07 |
| SITE03.WITH | SITE05 | -0.008 | 0.002 | -4.68 | 0 | -4 | 6.33E-05 |
| SITE03.WITH | SITE06 | -0.008 | 0.002 | -4.607 | 0 | -4 | 6.33E-05 |
| SITE03.WITH | SITE07 | -0.008 | 0.002 | -4.666 | 0 | -4 | 6.33E-05 |
| SITE03.WITH | SITE08 | -0.006 | 0.001 | -4.428 | 0 | -6 | 1.97E-09 |
| SITE03.WITH | SITE09 | -0.009 | 0.002 | -4.72 | 0 | -4.5 | 6.8E-06 |
| SITE04.WITH | SITE05 | -0.012 | 0.002 | -5.462 | 0 | -6 | 1.97E-09 |
| SITE04.WITH | SITE06 | -0.011 | 0.002 | -5.347 | 0 | -5.5 | 3.8E-08 |
| SITE04.WITH | SITE07 | -0.012 | 0.002 | -5.44 | 0 | -6 | 1.97E-09 |
| SITE04.WITH | SITE08 | -0.009 | 0.002 | -5.073 | 0 | -4.5 | 6.8E-06 |
| SITE04.WITH | SITE09 | -0.013 | 0.002 | -5.526 | 0 | -6.5 | 8.03E-11 |
| SITE05.WITH | SITE06 | -0.018 | 0.003 | -6.318 | 0 | -6 | 1.97E-09 |
| SITE05.WITH | SITE07 | -0.019 | 0.003 | -6.473 | 0 | -6.33333 | 2.4E-10 |
| SITE05.WITH | SITE08 | -0.015 | 0.003 | -5.879 | 0 | -5 | 5.73E-07 |
| SITE05.WITH | SITE09 | -0.021 | 0.003 | -6.619 | 0 | -7 | 2.56E-12 |
| SITE06.WITH | SITE07 | -0.018 | 0.003 | -6.283 | 0 | -6 | 1.97E-09 |
| SITE06.WITH | SITE08 | -0.014 | 0.002 | -5.735 | 0 | -7 | 2.56E-12 |
| SITE06.WITH | SITE09 | -0.019 | 0.003 | -6.417 | 0 | -6.33333 | 2.4E-10 |
| SITE07.WITH | SITE08 | -0.015 | 0.002 | -5.851 | 0 | -7.5 | 6.37E-14 |
| SITE07.WITH | SITE09 | -0.02 | 0.003 | -6.58 | 0 | -6.66667 | 2.62E-11 |
| SITE08.WITH | SITE09 | -0.016 | 0.003 | -5.958 | 0 | -5.33333 | 9.64E-08 |
| Means | CHR | 0.821 | 0.018 | 44.62 | 0 | 45.61111 | 0 |
| Means | AGE | 19.056 | 0.191 | 99.773 | 0 | 99.76963 | 0 |
| Means | FEMALE | 0.453 | 0.024 | 18.974 | 0 | 18.875 | 0 |
| Means | SITE02 | 0.087 | 0.014 | 6.453 | 0 | 6.214286 | 5.16E-10 |
| Means | SITE03 | 0.06 | 0.011 | 5.259 | 0 | 5.454545 | 4.91E-08 |
| Means | SITE04 | 0.087 | 0.014 | 6.453 | 0 | 6.214286 | 5.16E-10 |
| Means | SITE05 | 0.14 | 0.017 | 8.423 | 0 | 8.235294 | 2.22E-16 |
| Means | SITE06 | 0.129 | 0.016 | 8.017 | 0 | 8.0625 | 6.66E-16 |
| Means | SITE07 | 0.138 | 0.017 | 8.343 | 0 | 8.117647 | 4.44E-16 |
| Means | SITE08 | 0.106 | 0.015 | 7.172 | 0 | 7.066667 | 1.59E-12 |
| Means | SITE09 | 0.147 | 0.017 | 8.662 | 0 | 8.647059 | 0 |
| Intercepts | DEPS_CH | 0.204 | 0.234 | 0.871 | 0.384 | 0.871795 | 0.38332 |
| Intercepts | HI_CH | -0.002 | 0.002 | -1.007 | 0.314 | -1 | 0.317311 |
| Variances | CHR | 0.147 | 0.012 | 12.474 | 0 | 12.25 | 0 |
| Variances | AGE | 15.868 | 1.097 | 14.462 | 0 | 14.4649 | 0 |
| Variances | FEMALE | 0.248 | 0.002 | 110.149 | 0 | 124 | 0 |
| Variances | SITE02 | 0.08 | 0.011 | 7.136 | 0 | 7.272727 | 3.52E-13 |
| Variances | SITE03 | 0.056 | 0.01 | 5.616 | 0 | 5.6 | 2.14E-08 |
| Variances | SITE04 | 0.08 | 0.011 | 7.136 | 0 | 7.272727 | 3.52E-13 |
| Variances | SITE05 | 0.121 | 0.012 | 10.065 | 0 | 10.08333 | 0 |
| Variances | SITE06 | 0.112 | 0.012 | 9.407 | 0 | 9.333333 | 0 |
| Variances | SITE07 | 0.119 | 0.012 | 9.932 | 0 | 9.916667 | 0 |
| Variances | SITE08 | 0.095 | 0.012 | 8.134 | 0 | 7.916667 | 2.44E-15 |
| Variances | SITE09 | 0.125 | 0.012 | 10.468 | 0 | 10.41667 | 0 |
| Variances | LV_HI | 0.001 | 0 | 5.586 | 0 |  |  |
| Residual.Variances | DEPS_CH | 0.178 | 0 |  |  |  |  |
| Residual.Variances | HI_CH | 0 | 0 |  |  |  |  |
| Residual.Variances | LV_DEPS | 0.422 | 0.114 | 3.72 | 0 | 3.701754 | 0.000214 |
| New.Additional.Parameters | EFFECT_C | 0.103 | 1.005 | 0.103 | 0.918 | 0.102488 | 0.91837 |
| New.Additional.Parameters | EFFECT_P | -10.846 | 4.579 | -2.369 | 0.018 | -2.36864 | 0.017854 |
| LV_HI.BY | HI_CH | 1 | 0 |  |  |  |  |
| LV_GAF.BY | GAF_CH | 1 | 0 |  |  |  |  |
| LV_GAF.ON | LV_HI | -2.109 | 4.067 | -0.519 | 0.604 | -0.51856 | 0.604065 |
| LV_GAF.ON | HIXCHR | 32.035 | 10.223 | 3.134 | 0.002 | 3.13362 | 0.001727 |
| LV_GAF.ON | CHR | 0.64 | 0.169 | 3.787 | 0 | 3.786982 | 0.000152 |
| LV_GAF.ON | AGE | -0.052 | 0.026 | -2.019 | 0.043 | -2 | 0.0455 |
| LV_GAF.ON | FEMALE | -0.005 | 0.204 | -0.025 | 0.98 | -0.02451 | 0.980446 |
| LV_GAF.ON | SITE02 | -0.533 | 0.481 | -1.108 | 0.268 | -1.10811 | 0.267815 |
| LV_GAF.ON | SITE03 | -0.889 | 0.469 | -1.896 | 0.058 | -1.89552 | 0.058023 |
| LV_GAF.ON | SITE04 | -0.306 | 0.428 | -0.714 | 0.475 | -0.71495 | 0.474638 |
| LV_GAF.ON | SITE05 | -0.298 | 0.419 | -0.712 | 0.476 | -0.71122 | 0.47695 |
| LV_GAF.ON | SITE06 | -0.81 | 0.43 | -1.883 | 0.06 | -1.88372 | 0.059603 |
| LV_GAF.ON | SITE07 | -0.781 | 0.497 | -1.573 | 0.116 | -1.57143 | 0.116083 |
| LV_GAF.ON | SITE08 | -0.059 | 0.565 | -0.104 | 0.917 | -0.10442 | 0.916832 |
| LV_GAF.ON | SITE09 | -0.852 | 0.443 | -1.925 | 0.054 | -1.92325 | 0.054449 |
| LV_HI.WITH | CHR | 0 | 0 | 0.022 | 0.982 |  |  |
| LV_HI.WITH | AGE | 0.002 | 0.006 | 0.335 | 0.738 | 0.333333 | 0.738883 |
| LV_HI.WITH | FEMALE | -0.001 | 0.001 | -1.533 | 0.125 | -1 | 0.317311 |
| LV_HI.WITH | SITE02 | 0 | 0 | 0.388 | 0.698 |  |  |
| LV_HI.WITH | SITE03 | 0 | 0.001 | -0.129 | 0.897 | 0 | 1 |
| LV_HI.WITH | SITE04 | 0 | 0 | 1.354 | 0.176 |  |  |
| LV_HI.WITH | SITE05 | -0.001 | 0.001 | -1.007 | 0.314 | -1 | 0.317311 |
| LV_HI.WITH | SITE06 | 0.001 | 0 | 1.466 | 0.143 |  |  |
| LV_HI.WITH | SITE07 | 0 | 0.001 | -0.105 | 0.917 | 0 | 1 |
| LV_HI.WITH | SITE08 | 0 | 0 | -0.207 | 0.836 |  |  |
| LV_HI.WITH | SITE09 | 0 | 0.001 | -0.726 | 0.468 | 0 | 1 |
| CHR.WITH | AGE | -0.073 | 0.078 | -0.939 | 0.347 | -0.9359 | 0.349326 |
| CHR.WITH | FEMALE | -0.006 | 0.009 | -0.649 | 0.517 | -0.66667 | 0.504985 |
| CHR.WITH | SITE02 | -0.005 | 0.006 | -0.868 | 0.385 | -0.83333 | 0.404657 |
| CHR.WITH | SITE03 | -0.003 | 0.005 | -0.637 | 0.524 | -0.6 | 0.548506 |
| CHR.WITH | SITE04 | 0.002 | 0.005 | 0.384 | 0.701 | 0.4 | 0.689157 |
| CHR.WITH | SITE05 | 0 | 0.006 | -0.013 | 0.989 | 0 | 1 |
| CHR.WITH | SITE06 | -0.002 | 0.006 | -0.339 | 0.735 | -0.33333 | 0.738883 |
| CHR.WITH | SITE07 | 0 | 0.006 | -0.078 | 0.938 | 0 | 1 |
| CHR.WITH | SITE08 | 0.003 | 0.005 | 0.544 | 0.586 | 0.6 | 0.548506 |
| CHR.WITH | SITE09 | 0.001 | 0.006 | 0.18 | 0.857 | 0.166667 | 0.867632 |
| AGE.WITH | FEMALE | -0.221 | 0.094 | -2.339 | 0.019 | -2.35106 | 0.01872 |
| AGE.WITH | SITE02 | 0.342 | 0.076 | 4.507 | 0 | 4.5 | 6.8E-06 |
| AGE.WITH | SITE03 | 0 | 0.042 | -0.006 | 0.995 | 0 | 1 |
| AGE.WITH | SITE04 | -0.015 | 0.043 | -0.354 | 0.723 | -0.34884 | 0.727212 |
| AGE.WITH | SITE05 | 0.214 | 0.062 | 3.451 | 0.001 | 3.451613 | 0.000557 |
| AGE.WITH | SITE06 | -0.013 | 0.054 | -0.237 | 0.813 | -0.24074 | 0.809756 |
| AGE.WITH | SITE07 | -0.197 | 0.066 | -2.998 | 0.003 | -2.98485 | 0.002837 |
| AGE.WITH | SITE08 | -0.21 | 0.065 | -3.247 | 0.001 | -3.23077 | 0.001235 |
| AGE.WITH | SITE09 | -0.043 | 0.062 | -0.69 | 0.49 | -0.69355 | 0.487965 |
| FEMALE.WITH | SITE02 | 0.004 | 0.007 | 0.593 | 0.553 | 0.571429 | 0.567709 |
| FEMALE.WITH | SITE03 | -0.002 | 0.006 | -0.328 | 0.743 | -0.33333 | 0.738883 |
| FEMALE.WITH | SITE04 | -0.001 | 0.007 | -0.085 | 0.932 | -0.14286 | 0.886403 |
| FEMALE.WITH | SITE05 | 0.001 | 0.008 | 0.086 | 0.931 | 0.125 | 0.900524 |
| FEMALE.WITH | SITE06 | -0.01 | 0.008 | -1.29 | 0.197 | -1.25 | 0.2113 |
| FEMALE.WITH | SITE07 | 0.011 | 0.008 | 1.317 | 0.188 | 1.375 | 0.169131 |
| FEMALE.WITH | SITE08 | 0 | 0.007 | 0.038 | 0.97 | 0 | 1 |
| FEMALE.WITH | SITE09 | 0.002 | 0.008 | 0.258 | 0.797 | 0.25 | 0.802587 |
| SITE02.WITH | SITE03 | -0.005 | 0.001 | -4.242 | 0 | -5 | 5.73E-07 |
| SITE02.WITH | SITE04 | -0.008 | 0.002 | -4.799 | 0 | -4 | 6.33E-05 |
| SITE02.WITH | SITE05 | -0.012 | 0.002 | -5.464 | 0 | -6 | 1.97E-09 |
| SITE02.WITH | SITE06 | -0.011 | 0.002 | -5.349 | 0 | -5.5 | 3.8E-08 |
| SITE02.WITH | SITE07 | -0.012 | 0.002 | -5.442 | 0 | -6 | 1.97E-09 |
| SITE02.WITH | SITE08 | -0.009 | 0.002 | -5.074 | 0 | -4.5 | 6.8E-06 |
| SITE02.WITH | SITE09 | -0.013 | 0.002 | -5.528 | 0 | -6.5 | 8.03E-11 |
| SITE03.WITH | SITE04 | -0.005 | 0.001 | -4.242 | 0 | -5 | 5.73E-07 |
| SITE03.WITH | SITE05 | -0.008 | 0.002 | -4.681 | 0 | -4 | 6.33E-05 |
| SITE03.WITH | SITE06 | -0.008 | 0.002 | -4.608 | 0 | -4 | 6.33E-05 |
| SITE03.WITH | SITE07 | -0.008 | 0.002 | -4.667 | 0 | -4 | 6.33E-05 |
| SITE03.WITH | SITE08 | -0.006 | 0.001 | -4.429 | 0 | -6 | 1.97E-09 |
| SITE03.WITH | SITE09 | -0.009 | 0.002 | -4.721 | 0 | -4.5 | 6.8E-06 |
| SITE04.WITH | SITE05 | -0.012 | 0.002 | -5.464 | 0 | -6 | 1.97E-09 |
| SITE04.WITH | SITE06 | -0.011 | 0.002 | -5.349 | 0 | -5.5 | 3.8E-08 |
| SITE04.WITH | SITE07 | -0.012 | 0.002 | -5.442 | 0 | -6 | 1.97E-09 |
| SITE04.WITH | SITE08 | -0.009 | 0.002 | -5.074 | 0 | -4.5 | 6.8E-06 |
| SITE04.WITH | SITE09 | -0.013 | 0.002 | -5.528 | 0 | -6.5 | 8.03E-11 |
| SITE05.WITH | SITE06 | -0.018 | 0.003 | -6.32 | 0 | -6 | 1.97E-09 |
| SITE05.WITH | SITE07 | -0.019 | 0.003 | -6.476 | 0 | -6.33333 | 2.4E-10 |
| SITE05.WITH | SITE08 | -0.015 | 0.003 | -5.881 | 0 | -5 | 5.73E-07 |
| SITE05.WITH | SITE09 | -0.021 | 0.003 | -6.622 | 0 | -7 | 2.56E-12 |
| SITE06.WITH | SITE07 | -0.018 | 0.003 | -6.286 | 0 | -6 | 1.97E-09 |
| SITE06.WITH | SITE08 | -0.014 | 0.002 | -5.737 | 0 | -7 | 2.56E-12 |
| SITE06.WITH | SITE09 | -0.019 | 0.003 | -6.42 | 0 | -6.33333 | 2.4E-10 |
| SITE07.WITH | SITE08 | -0.015 | 0.003 | -5.853 | 0 | -5 | 5.73E-07 |
| SITE07.WITH | SITE09 | -0.02 | 0.003 | -6.583 | 0 | -6.66667 | 2.62E-11 |
| SITE08.WITH | SITE09 | -0.016 | 0.003 | -5.96 | 0 | -5.33333 | 9.64E-08 |
| Means | CHR | 0.82 | 0.018 | 44.506 | 0 | 45.55556 | 0 |
| Means | AGE | 19.062 | 0.191 | 99.626 | 0 | 99.80105 | 0 |
| Means | FEMALE | 0.454 | 0.024 | 18.996 | 0 | 18.91667 | 0 |
| Means | SITE02 | 0.088 | 0.014 | 6.453 | 0 | 6.285714 | 3.26E-10 |
| Means | SITE03 | 0.06 | 0.011 | 5.259 | 0 | 5.454545 | 4.91E-08 |
| Means | SITE04 | 0.088 | 0.014 | 6.453 | 0 | 6.285714 | 3.26E-10 |
| Means | SITE05 | 0.141 | 0.017 | 8.425 | 0 | 8.294118 | 0 |
| Means | SITE06 | 0.129 | 0.016 | 8.018 | 0 | 8.0625 | 6.66E-16 |
| Means | SITE07 | 0.138 | 0.017 | 8.344 | 0 | 8.117647 | 4.44E-16 |
| Means | SITE08 | 0.106 | 0.015 | 7.173 | 0 | 7.066667 | 1.59E-12 |
| Means | SITE09 | 0.147 | 0.017 | 8.665 | 0 | 8.647059 | 0 |
| Intercepts | GAF_CH | 1.553 | 0.597 | 2.599 | 0.009 | 2.60134 | 0.009286 |
| Intercepts | HI_CH | -0.002 | 0.002 | -1.017 | 0.309 | -1 | 0.317311 |
| Variances | CHR | 0.147 | 0.012 | 12.487 | 0 | 12.25 | 0 |
| Variances | AGE | 15.888 | 1.099 | 14.463 | 0 | 14.45678 | 0 |
| Variances | FEMALE | 0.248 | 0.002 | 112.536 | 0 | 124 | 0 |
| Variances | SITE02 | 0.08 | 0.011 | 7.138 | 0 | 7.272727 | 3.52E-13 |
| Variances | SITE03 | 0.056 | 0.01 | 5.617 | 0 | 5.6 | 2.14E-08 |
| Variances | SITE04 | 0.08 | 0.011 | 7.138 | 0 | 7.272727 | 3.52E-13 |
| Variances | SITE05 | 0.121 | 0.012 | 10.072 | 0 | 10.08333 | 0 |
| Variances | SITE06 | 0.112 | 0.012 | 9.413 | 0 | 9.333333 | 0 |
| Variances | SITE07 | 0.119 | 0.012 | 9.939 | 0 | 9.916667 | 0 |
| Variances | SITE08 | 0.095 | 0.012 | 8.138 | 0 | 7.916667 | 2.44E-15 |
| Variances | SITE09 | 0.126 | 0.012 | 10.476 | 0 | 10.5 | 0 |
| Variances | LV_HI | 0.001 | 0 | 5.577 | 0 |  |  |
| Residual.Variances | GAF_CH | 0.807 | 0 |  |  |  |  |
| Residual.Variances | HI_CH | 0 | 0 |  |  |  |  |
| Residual.Variances | LV_GAF | 3.163 | 0.513 | 6.164 | 0 | 6.165692 | 7.02E-10 |
| New.Additional.Parameters | EFFECT_C | -2.109 | 4.067 | -0.519 | 0.604 | -0.51856 | 0.604065 |
| New.Additional.Parameters | EFFECT_P | 29.926 | 9.418 | 3.178 | 0.001 | 3.177532 | 0.001485 |
| LV_HI.BY | HI_CH | 1 | 0 |  |  |  |  |
| LV_NEGS.BY | NEGS_CH | 1 | 0 |  |  |  |  |
| LV_NEGS.ON | LV_HI | 2.409 | 2.021 | 1.192 | 0.233 | 1.191984 | 0.233267 |
| LV_NEGS.ON | HIXCHR | -17.297 | 5.167 | -3.348 | 0.001 | -3.34759 | 0.000815 |
| LV_NEGS.ON | CHR | -0.308 | 0.077 | -3.974 | 0 | -4 | 6.33E-05 |
| LV_NEGS.ON | AGE | -0.003 | 0.013 | -0.238 | 0.812 | -0.23077 | 0.817494 |
| LV_NEGS.ON | FEMALE | -0.004 | 0.086 | -0.043 | 0.966 | -0.04651 | 0.962902 |
| LV_NEGS.ON | SITE02 | -0.499 | 0.279 | -1.79 | 0.074 | -1.78853 | 0.07369 |
| LV_NEGS.ON | SITE03 | 0.245 | 0.259 | 0.946 | 0.344 | 0.945946 | 0.344176 |
| LV_NEGS.ON | SITE04 | -0.381 | 0.211 | -1.803 | 0.071 | -1.80569 | 0.070967 |
| LV_NEGS.ON | SITE05 | -0.106 | 0.213 | -0.499 | 0.617 | -0.49765 | 0.618729 |
| LV_NEGS.ON | SITE06 | -0.281 | 0.22 | -1.278 | 0.201 | -1.27727 | 0.201506 |
| LV_NEGS.ON | SITE07 | -0.214 | 0.202 | -1.059 | 0.29 | -1.05941 | 0.289415 |
| LV_NEGS.ON | SITE08 | -0.438 | 0.227 | -1.932 | 0.053 | -1.92952 | 0.053667 |
| LV_NEGS.ON | SITE09 | 0.081 | 0.236 | 0.345 | 0.73 | 0.34322 | 0.731433 |
| LV_HI.WITH | CHR | 0 | 0 | -0.091 | 0.927 |  |  |
| LV_HI.WITH | AGE | 0.002 | 0.006 | 0.417 | 0.677 | 0.333333 | 0.738883 |
| LV_HI.WITH | FEMALE | -0.001 | 0.001 | -1.34 | 0.18 | -1 | 0.317311 |
| LV_HI.WITH | SITE02 | 0 | 0 | 0.506 | 0.613 |  |  |
| LV_HI.WITH | SITE03 | 0 | 0 | -0.676 | 0.499 |  |  |
| LV_HI.WITH | SITE04 | 0 | 0 | 1.429 | 0.153 |  |  |
| LV_HI.WITH | SITE05 | -0.001 | 0.001 | -0.985 | 0.324 | -1 | 0.317311 |
| LV_HI.WITH | SITE06 | 0.001 | 0 | 1.677 | 0.094 |  |  |
| LV_HI.WITH | SITE07 | 0 | 0.001 | -0.016 | 0.987 | 0 | 1 |
| LV_HI.WITH | SITE08 | 0 | 0 | -0.149 | 0.882 |  |  |
| LV_HI.WITH | SITE09 | 0 | 0.001 | -0.685 | 0.493 | 0 | 1 |
| CHR.WITH | AGE | -0.074 | 0.078 | -0.95 | 0.342 | -0.94872 | 0.342764 |
| CHR.WITH | FEMALE | -0.006 | 0.009 | -0.649 | 0.517 | -0.66667 | 0.504985 |
| CHR.WITH | SITE02 | -0.005 | 0.006 | -0.868 | 0.385 | -0.83333 | 0.404657 |
| CHR.WITH | SITE03 | -0.003 | 0.005 | -0.725 | 0.469 | -0.6 | 0.548506 |
| CHR.WITH | SITE04 | 0.002 | 0.005 | 0.384 | 0.701 | 0.4 | 0.689157 |
| CHR.WITH | SITE05 | 0 | 0.006 | -0.013 | 0.989 | 0 | 1 |
| CHR.WITH | SITE06 | -0.002 | 0.006 | -0.339 | 0.735 | -0.33333 | 0.738883 |
| CHR.WITH | SITE07 | 0 | 0.006 | -0.078 | 0.938 | 0 | 1 |
| CHR.WITH | SITE08 | 0.003 | 0.005 | 0.544 | 0.586 | 0.6 | 0.548506 |
| CHR.WITH | SITE09 | 0.001 | 0.006 | 0.18 | 0.857 | 0.166667 | 0.867632 |
| AGE.WITH | FEMALE | -0.219 | 0.094 | -2.315 | 0.021 | -2.32979 | 0.019817 |
| AGE.WITH | SITE02 | 0.343 | 0.076 | 4.51 | 0 | 4.513158 | 6.39E-06 |
| AGE.WITH | SITE03 | 0.001 | 0.042 | 0.032 | 0.975 | 0.02381 | 0.981005 |
| AGE.WITH | SITE04 | -0.015 | 0.043 | -0.345 | 0.73 | -0.34884 | 0.727212 |
| AGE.WITH | SITE05 | 0.215 | 0.062 | 3.46 | 0.001 | 3.467742 | 0.000525 |
| AGE.WITH | SITE06 | -0.012 | 0.054 | -0.225 | 0.822 | -0.22222 | 0.824141 |
| AGE.WITH | SITE07 | -0.197 | 0.066 | -2.988 | 0.003 | -2.98485 | 0.002837 |
| AGE.WITH | SITE08 | -0.209 | 0.065 | -3.24 | 0.001 | -3.21538 | 0.001303 |
| AGE.WITH | SITE09 | -0.042 | 0.062 | -0.679 | 0.497 | -0.67742 | 0.49814 |
| FEMALE.WITH | SITE02 | 0.004 | 0.007 | 0.593 | 0.553 | 0.571429 | 0.567709 |
| FEMALE.WITH | SITE03 | -0.001 | 0.006 | -0.144 | 0.885 | -0.16667 | 0.867632 |
| FEMALE.WITH | SITE04 | -0.001 | 0.007 | -0.085 | 0.932 | -0.14286 | 0.886403 |
| FEMALE.WITH | SITE05 | 0.001 | 0.008 | 0.086 | 0.931 | 0.125 | 0.900524 |
| FEMALE.WITH | SITE06 | -0.01 | 0.008 | -1.29 | 0.197 | -1.25 | 0.2113 |
| FEMALE.WITH | SITE07 | 0.011 | 0.008 | 1.317 | 0.188 | 1.375 | 0.169131 |
| FEMALE.WITH | SITE08 | 0 | 0.007 | 0.038 | 0.97 | 0 | 1 |
| FEMALE.WITH | SITE09 | 0.002 | 0.008 | 0.258 | 0.797 | 0.25 | 0.802587 |
| SITE02.WITH | SITE03 | -0.005 | 0.001 | -4.185 | 0 | -5 | 5.73E-07 |
| SITE02.WITH | SITE04 | -0.008 | 0.002 | -4.799 | 0 | -4 | 6.33E-05 |
| SITE02.WITH | SITE05 | -0.012 | 0.002 | -5.464 | 0 | -6 | 1.97E-09 |
| SITE02.WITH | SITE06 | -0.011 | 0.002 | -5.349 | 0 | -5.5 | 3.8E-08 |
| SITE02.WITH | SITE07 | -0.012 | 0.002 | -5.442 | 0 | -6 | 1.97E-09 |
| SITE02.WITH | SITE08 | -0.009 | 0.002 | -5.074 | 0 | -4.5 | 6.8E-06 |
| SITE02.WITH | SITE09 | -0.013 | 0.002 | -5.528 | 0 | -6.5 | 8.03E-11 |
| SITE03.WITH | SITE04 | -0.005 | 0.001 | -4.185 | 0 | -5 | 5.73E-07 |
| SITE03.WITH | SITE05 | -0.008 | 0.002 | -4.604 | 0 | -4 | 6.33E-05 |
| SITE03.WITH | SITE06 | -0.007 | 0.002 | -4.534 | 0 | -3.5 | 0.000465 |
| SITE03.WITH | SITE07 | -0.008 | 0.002 | -4.591 | 0 | -4 | 6.33E-05 |
| SITE03.WITH | SITE08 | -0.006 | 0.001 | -4.363 | 0 | -6 | 1.97E-09 |
| SITE03.WITH | SITE09 | -0.008 | 0.002 | -4.642 | 0 | -4 | 6.33E-05 |
| SITE04.WITH | SITE05 | -0.012 | 0.002 | -5.464 | 0 | -6 | 1.97E-09 |
| SITE04.WITH | SITE06 | -0.011 | 0.002 | -5.349 | 0 | -5.5 | 3.8E-08 |
| SITE04.WITH | SITE07 | -0.012 | 0.002 | -5.442 | 0 | -6 | 1.97E-09 |
| SITE04.WITH | SITE08 | -0.009 | 0.002 | -5.074 | 0 | -4.5 | 6.8E-06 |
| SITE04.WITH | SITE09 | -0.013 | 0.002 | -5.528 | 0 | -6.5 | 8.03E-11 |
| SITE05.WITH | SITE06 | -0.018 | 0.003 | -6.32 | 0 | -6 | 1.97E-09 |
| SITE05.WITH | SITE07 | -0.019 | 0.003 | -6.476 | 0 | -6.33333 | 2.4E-10 |
| SITE05.WITH | SITE08 | -0.015 | 0.003 | -5.881 | 0 | -5 | 5.73E-07 |
| SITE05.WITH | SITE09 | -0.021 | 0.003 | -6.622 | 0 | -7 | 2.56E-12 |
| SITE06.WITH | SITE07 | -0.018 | 0.003 | -6.286 | 0 | -6 | 1.97E-09 |
| SITE06.WITH | SITE08 | -0.014 | 0.002 | -5.737 | 0 | -7 | 2.56E-12 |
| SITE06.WITH | SITE09 | -0.019 | 0.003 | -6.42 | 0 | -6.33333 | 2.4E-10 |
| SITE07.WITH | SITE08 | -0.015 | 0.003 | -5.853 | 0 | -5 | 5.73E-07 |
| SITE07.WITH | SITE09 | -0.02 | 0.003 | -6.583 | 0 | -6.66667 | 2.62E-11 |
| SITE08.WITH | SITE09 | -0.016 | 0.003 | -5.96 | 0 | -5.33333 | 9.64E-08 |
| Means | CHR | 0.82 | 0.018 | 44.506 | 0 | 45.55556 | 0 |
| Means | AGE | 19.057 | 0.191 | 99.551 | 0 | 99.77487 | 0 |
| Means | FEMALE | 0.454 | 0.024 | 18.992 | 0 | 18.91667 | 0 |
| Means | SITE02 | 0.088 | 0.014 | 6.453 | 0 | 6.285714 | 3.26E-10 |
| Means | SITE03 | 0.058 | 0.011 | 5.15 | 0 | 5.272727 | 1.34E-07 |
| Means | SITE04 | 0.088 | 0.014 | 6.454 | 0 | 6.285714 | 3.26E-10 |
| Means | SITE05 | 0.141 | 0.017 | 8.425 | 0 | 8.294118 | 0 |
| Means | SITE06 | 0.129 | 0.016 | 8.019 | 0 | 8.0625 | 6.66E-16 |
| Means | SITE07 | 0.138 | 0.017 | 8.344 | 0 | 8.117647 | 4.44E-16 |
| Means | SITE08 | 0.106 | 0.015 | 7.173 | 0 | 7.066667 | 1.59E-12 |
| Means | SITE09 | 0.147 | 0.017 | 8.664 | 0 | 8.647059 | 0 |
| Intercepts | NEGS_CH | 0.254 | 0.31 | 0.818 | 0.413 | 0.819355 | 0.412584 |
| Intercepts | HI_CH | -0.002 | 0.002 | -1.182 | 0.237 | -1 | 0.317311 |
| Variances | CHR | 0.147 | 0.012 | 12.487 | 0 | 12.25 | 0 |
| Variances | AGE | 15.904 | 1.099 | 14.472 | 0 | 14.47134 | 0 |
| Variances | FEMALE | 0.248 | 0.002 | 112.536 | 0 | 124 | 0 |
| Variances | SITE02 | 0.08 | 0.011 | 7.138 | 0 | 7.272727 | 3.52E-13 |
| Variances | SITE03 | 0.054 | 0.01 | 5.486 | 0 | 5.4 | 6.66E-08 |
| Variances | SITE04 | 0.08 | 0.011 | 7.138 | 0 | 7.272727 | 3.52E-13 |
| Variances | SITE05 | 0.121 | 0.012 | 10.072 | 0 | 10.08333 | 0 |
| Variances | SITE06 | 0.112 | 0.012 | 9.413 | 0 | 9.333333 | 0 |
| Variances | SITE07 | 0.119 | 0.012 | 9.939 | 0 | 9.916667 | 0 |
| Variances | SITE08 | 0.095 | 0.012 | 8.138 | 0 | 7.916667 | 2.44E-15 |
| Variances | SITE09 | 0.126 | 0.012 | 10.476 | 0 | 10.5 | 0 |
| Variances | LV_HI | 0.001 | 0 | 5.493 | 0 |  |  |
| Residual.Variances | NEGS_CH | 0.178 | 0 |  |  |  |  |
| Residual.Variances | HI_CH | 0 | 0 |  |  |  |  |
| Residual.Variances | LV_NEGS | 0.654 | 0.152 | 4.294 | 0 | 4.302632 | 1.69E-05 |
| New.Additional.Parameters | EFFECT_C | 2.409 | 2.021 | 1.192 | 0.233 | 1.191984 | 0.233267 |
| New.Additional.Parameters | EFFECT_P | -14.888 | 4.369 | -3.407 | 0.001 | -3.40764 | 0.000655 |
| LV_HI.BY | HI_CH | 1 | 0 |  |  |  |  |
| LV_POSS.BY | POSS_CH | 1 | 0 |  |  |  |  |
| LV_POSS.ON | LV_HI | -2.002 | 1.142 | -1.753 | 0.08 | -1.75306 | 0.079591 |
| LV_POSS.ON | HIXCHR | -2.699 | 4.028 | -0.67 | 0.503 | -0.67006 | 0.50282 |
| LV_POSS.ON | CHR | -0.436 | 0.049 | -8.936 | 0 | -8.89796 | 0 |
| LV_POSS.ON | AGE | 0 | 0.009 | 0.055 | 0.956 | 0 | 1 |
| LV_POSS.ON | FEMALE | 0.083 | 0.06 | 1.384 | 0.166 | 1.383333 | 0.166563 |
| LV_POSS.ON | SITE02 | -0.013 | 0.215 | -0.059 | 0.953 | -0.06047 | 0.951785 |
| LV_POSS.ON | SITE03 | 0.149 | 0.159 | 0.934 | 0.35 | 0.937107 | 0.348704 |
| LV_POSS.ON | SITE04 | 0.084 | 0.145 | 0.575 | 0.565 | 0.57931 | 0.56238 |
| LV_POSS.ON | SITE05 | 0.247 | 0.137 | 1.796 | 0.072 | 1.80292 | 0.071401 |
| LV_POSS.ON | SITE06 | -0.001 | 0.138 | -0.006 | 0.995 | -0.00725 | 0.994218 |
| LV_POSS.ON | SITE07 | 0.08 | 0.128 | 0.625 | 0.532 | 0.625 | 0.531971 |
| LV_POSS.ON | SITE08 | 0.118 | 0.149 | 0.794 | 0.427 | 0.791946 | 0.428392 |
| LV_POSS.ON | SITE09 | 0.118 | 0.139 | 0.852 | 0.394 | 0.848921 | 0.395925 |
| LV_HI.WITH | CHR | 0 | 0 | 0.005 | 0.996 |  |  |
| LV_HI.WITH | AGE | 0.002 | 0.006 | 0.35 | 0.726 | 0.333333 | 0.738883 |
| LV_HI.WITH | FEMALE | -0.001 | 0.001 | -1.462 | 0.144 | -1 | 0.317311 |
| LV_HI.WITH | SITE02 | 0 | 0 | 0.295 | 0.768 |  |  |
| LV_HI.WITH | SITE03 | 0 | 0.001 | -0.099 | 0.921 | 0 | 1 |
| LV_HI.WITH | SITE04 | 0 | 0 | 1.36 | 0.174 |  |  |
| LV_HI.WITH | SITE05 | -0.001 | 0.001 | -0.969 | 0.333 | -1 | 0.317311 |
| LV_HI.WITH | SITE06 | 0.001 | 0 | 1.613 | 0.107 |  |  |
| LV_HI.WITH | SITE07 | 0 | 0.001 | -0.137 | 0.891 | 0 | 1 |
| LV_HI.WITH | SITE08 | 0 | 0 | -0.247 | 0.805 |  |  |
| LV_HI.WITH | SITE09 | 0 | 0.001 | -0.656 | 0.512 | 0 | 1 |
| CHR.WITH | AGE | -0.074 | 0.078 | -0.953 | 0.34 | -0.94872 | 0.342764 |
| CHR.WITH | FEMALE | -0.006 | 0.009 | -0.669 | 0.504 | -0.66667 | 0.504985 |
| CHR.WITH | SITE02 | -0.005 | 0.006 | -0.875 | 0.382 | -0.83333 | 0.404657 |
| CHR.WITH | SITE03 | -0.003 | 0.005 | -0.642 | 0.521 | -0.6 | 0.548506 |
| CHR.WITH | SITE04 | 0.002 | 0.005 | 0.377 | 0.707 | 0.4 | 0.689157 |
| CHR.WITH | SITE05 | 0 | 0.006 | -0.022 | 0.982 | 0 | 1 |
| CHR.WITH | SITE06 | -0.002 | 0.006 | -0.347 | 0.729 | -0.33333 | 0.738883 |
| CHR.WITH | SITE07 | -0.001 | 0.006 | -0.087 | 0.931 | -0.16667 | 0.867632 |
| CHR.WITH | SITE08 | 0.003 | 0.005 | 0.536 | 0.592 | 0.6 | 0.548506 |
| CHR.WITH | SITE09 | 0.001 | 0.006 | 0.17 | 0.865 | 0.166667 | 0.867632 |
| AGE.WITH | FEMALE | -0.218 | 0.094 | -2.309 | 0.021 | -2.31915 | 0.020387 |
| AGE.WITH | SITE02 | 0.342 | 0.076 | 4.51 | 0 | 4.5 | 6.8E-06 |
| AGE.WITH | SITE03 | 0 | 0.042 | 0.003 | 0.998 | 0 | 1 |
| AGE.WITH | SITE04 | -0.015 | 0.043 | -0.342 | 0.732 | -0.34884 | 0.727212 |
| AGE.WITH | SITE05 | 0.215 | 0.062 | 3.463 | 0.001 | 3.467742 | 0.000525 |
| AGE.WITH | SITE06 | -0.012 | 0.054 | -0.222 | 0.824 | -0.22222 | 0.824141 |
| AGE.WITH | SITE07 | -0.196 | 0.066 | -2.986 | 0.003 | -2.9697 | 0.002981 |
| AGE.WITH | SITE08 | -0.208 | 0.064 | -3.238 | 0.001 | -3.25 | 0.001154 |
| AGE.WITH | SITE09 | -0.042 | 0.062 | -0.676 | 0.499 | -0.67742 | 0.49814 |
| FEMALE.WITH | SITE02 | 0.004 | 0.007 | 0.606 | 0.544 | 0.571429 | 0.567709 |
| FEMALE.WITH | SITE03 | -0.002 | 0.006 | -0.316 | 0.752 | -0.33333 | 0.738883 |
| FEMALE.WITH | SITE04 | 0 | 0.007 | -0.071 | 0.943 | 0 | 1 |
| FEMALE.WITH | SITE05 | 0.001 | 0.008 | 0.104 | 0.917 | 0.125 | 0.900524 |
| FEMALE.WITH | SITE06 | -0.01 | 0.008 | -1.273 | 0.203 | -1.25 | 0.2113 |
| FEMALE.WITH | SITE07 | 0.011 | 0.008 | 1.334 | 0.182 | 1.375 | 0.169131 |
| FEMALE.WITH | SITE08 | 0 | 0.007 | 0.053 | 0.958 | 0 | 1 |
| FEMALE.WITH | SITE09 | 0.002 | 0.008 | 0.276 | 0.783 | 0.25 | 0.802587 |
| SITE02.WITH | SITE03 | -0.005 | 0.001 | -4.242 | 0 | -5 | 5.73E-07 |
| SITE02.WITH | SITE04 | -0.008 | 0.002 | -4.798 | 0 | -4 | 6.33E-05 |
| SITE02.WITH | SITE05 | -0.012 | 0.002 | -5.462 | 0 | -6 | 1.97E-09 |
| SITE02.WITH | SITE06 | -0.011 | 0.002 | -5.347 | 0 | -5.5 | 3.8E-08 |
| SITE02.WITH | SITE07 | -0.012 | 0.002 | -5.44 | 0 | -6 | 1.97E-09 |
| SITE02.WITH | SITE08 | -0.009 | 0.002 | -5.073 | 0 | -4.5 | 6.8E-06 |
| SITE02.WITH | SITE09 | -0.013 | 0.002 | -5.526 | 0 | -6.5 | 8.03E-11 |
| SITE03.WITH | SITE04 | -0.005 | 0.001 | -4.242 | 0 | -5 | 5.73E-07 |
| SITE03.WITH | SITE05 | -0.008 | 0.002 | -4.68 | 0 | -4 | 6.33E-05 |
| SITE03.WITH | SITE06 | -0.008 | 0.002 | -4.607 | 0 | -4 | 6.33E-05 |
| SITE03.WITH | SITE07 | -0.008 | 0.002 | -4.666 | 0 | -4 | 6.33E-05 |
| SITE03.WITH | SITE08 | -0.006 | 0.001 | -4.428 | 0 | -6 | 1.97E-09 |
| SITE03.WITH | SITE09 | -0.009 | 0.002 | -4.72 | 0 | -4.5 | 6.8E-06 |
| SITE04.WITH | SITE05 | -0.012 | 0.002 | -5.462 | 0 | -6 | 1.97E-09 |
| SITE04.WITH | SITE06 | -0.011 | 0.002 | -5.347 | 0 | -5.5 | 3.8E-08 |
| SITE04.WITH | SITE07 | -0.012 | 0.002 | -5.44 | 0 | -6 | 1.97E-09 |
| SITE04.WITH | SITE08 | -0.009 | 0.002 | -5.073 | 0 | -4.5 | 6.8E-06 |
| SITE04.WITH | SITE09 | -0.013 | 0.002 | -5.526 | 0 | -6.5 | 8.03E-11 |
| SITE05.WITH | SITE06 | -0.018 | 0.003 | -6.318 | 0 | -6 | 1.97E-09 |
| SITE05.WITH | SITE07 | -0.019 | 0.003 | -6.473 | 0 | -6.33333 | 2.4E-10 |
| SITE05.WITH | SITE08 | -0.015 | 0.003 | -5.879 | 0 | -5 | 5.73E-07 |
| SITE05.WITH | SITE09 | -0.021 | 0.003 | -6.619 | 0 | -7 | 2.56E-12 |
| SITE06.WITH | SITE07 | -0.018 | 0.003 | -6.283 | 0 | -6 | 1.97E-09 |
| SITE06.WITH | SITE08 | -0.014 | 0.002 | -5.735 | 0 | -7 | 2.56E-12 |
| SITE06.WITH | SITE09 | -0.019 | 0.003 | -6.417 | 0 | -6.33333 | 2.4E-10 |
| SITE07.WITH | SITE08 | -0.015 | 0.002 | -5.851 | 0 | -7.5 | 6.37E-14 |
| SITE07.WITH | SITE09 | -0.02 | 0.003 | -6.58 | 0 | -6.66667 | 2.62E-11 |
| SITE08.WITH | SITE09 | -0.016 | 0.003 | -5.958 | 0 | -5.33333 | 9.64E-08 |
| Means | CHR | 0.821 | 0.018 | 44.62 | 0 | 45.61111 | 0 |
| Means | AGE | 19.056 | 0.191 | 99.773 | 0 | 99.76963 | 0 |
| Means | FEMALE | 0.453 | 0.024 | 18.973 | 0 | 18.875 | 0 |
| Means | SITE02 | 0.087 | 0.014 | 6.453 | 0 | 6.214286 | 5.16E-10 |
| Means | SITE03 | 0.06 | 0.011 | 5.259 | 0 | 5.454545 | 4.91E-08 |
| Means | SITE04 | 0.087 | 0.014 | 6.453 | 0 | 6.214286 | 5.16E-10 |
| Means | SITE05 | 0.14 | 0.017 | 8.423 | 0 | 8.235294 | 2.22E-16 |
| Means | SITE06 | 0.129 | 0.016 | 8.017 | 0 | 8.0625 | 6.66E-16 |
| Means | SITE07 | 0.138 | 0.017 | 8.343 | 0 | 8.117647 | 4.44E-16 |
| Means | SITE08 | 0.106 | 0.015 | 7.172 | 0 | 7.066667 | 1.59E-12 |
| Means | SITE09 | 0.147 | 0.017 | 8.662 | 0 | 8.647059 | 0 |
| Intercepts | POSS_CH | -0.23 | 0.199 | -1.152 | 0.249 | -1.15578 | 0.247772 |
| Intercepts | HI_CH | -0.002 | 0.002 | -0.993 | 0.321 | -1 | 0.317311 |
| Variances | CHR | 0.147 | 0.012 | 12.474 | 0 | 12.25 | 0 |
| Variances | AGE | 15.868 | 1.097 | 14.462 | 0 | 14.4649 | 0 |
| Variances | FEMALE | 0.248 | 0.002 | 110.149 | 0 | 124 | 0 |
| Variances | SITE02 | 0.08 | 0.011 | 7.136 | 0 | 7.272727 | 3.52E-13 |
| Variances | SITE03 | 0.056 | 0.01 | 5.616 | 0 | 5.6 | 2.14E-08 |
| Variances | SITE04 | 0.08 | 0.011 | 7.136 | 0 | 7.272727 | 3.52E-13 |
| Variances | SITE05 | 0.121 | 0.012 | 10.065 | 0 | 10.08333 | 0 |
| Variances | SITE06 | 0.112 | 0.012 | 9.407 | 0 | 9.333333 | 0 |
| Variances | SITE07 | 0.119 | 0.012 | 9.932 | 0 | 9.916667 | 0 |
| Variances | SITE08 | 0.095 | 0.012 | 8.134 | 0 | 7.916667 | 2.44E-15 |
| Variances | SITE09 | 0.125 | 0.012 | 10.468 | 0 | 10.41667 | 0 |
| Variances | LV_HI | 0.001 | 0 | 5.589 | 0 |  |  |
| Residual.Variances | POSS_CH | 0.069 | 0 |  |  |  |  |
| Residual.Variances | HI_CH | 0 | 0 |  |  |  |  |
| Residual.Variances | LV_POSS | 0.36 | 0.083 | 4.318 | 0 | 4.337349 | 1.44E-05 |
| New.Additional.Parameters | EFFECT_C | -2.002 | 1.142 | -1.753 | 0.08 | -1.75306 | 0.079591 |
| New.Additional.Parameters | EFFECT_P | -4.701 | 3.711 | -1.267 | 0.205 | -1.26677 | 0.205236 |
| LV_HI.BY | HI_CH | 1 | 0 |  |  |  |  |
| LV_SYCOD.BY | SYCOD_CH | 1 | 0 |  |  |  |  |
| LV_SYCOD.ON | LV_HI | 11.152 | 11.208 | 0.995 | 0.32 | 0.995004 | 0.319735 |
| LV_SYCOD.ON | HIXCHR | -5.265 | 13.396 | -0.393 | 0.694 | -0.39303 | 0.694299 |
| LV_SYCOD.ON | CHR | -0.269 | 0.239 | -1.127 | 0.26 | -1.12552 | 0.260367 |
| LV_SYCOD.ON | AGE | -0.013 | 0.024 | -0.519 | 0.604 | -0.54167 | 0.588048 |
| LV_SYCOD.ON | FEMALE | -0.292 | 0.211 | -1.388 | 0.165 | -1.38389 | 0.166393 |
| LV_SYCOD.ON | SITE02 | 0.011 | 0.594 | 0.018 | 0.986 | 0.018519 | 0.985225 |
| LV_SYCOD.ON | SITE03 | 0.479 | 0.681 | 0.705 | 0.481 | 0.703377 | 0.481821 |
| LV_SYCOD.ON | SITE04 | -0.965 | 0.389 | -2.484 | 0.013 | -2.48072 | 0.013112 |
| LV_SYCOD.ON | SITE05 | 0.599 | 0.415 | 1.444 | 0.149 | 1.443373 | 0.148915 |
| LV_SYCOD.ON | SITE06 | -0.394 | 0.386 | -1.02 | 0.308 | -1.02073 | 0.307385 |
| LV_SYCOD.ON | SITE07 | -0.375 | 0.367 | -1.022 | 0.307 | -1.0218 | 0.306876 |
| LV_SYCOD.ON | SITE08 | -1.022 | 0.438 | -2.333 | 0.02 | -2.33333 | 0.019631 |
| LV_SYCOD.ON | SITE09 | -0.009 | 0.435 | -0.021 | 0.984 | -0.02069 | 0.983493 |
| LV_HI.WITH | CHR | 0 | 0 | -0.121 | 0.904 |  |  |
| LV_HI.WITH | AGE | 0.003 | 0.006 | 0.47 | 0.638 | 0.5 | 0.617075 |
| LV_HI.WITH | FEMALE | -0.001 | 0.001 | -1.383 | 0.167 | -1 | 0.317311 |
| LV_HI.WITH | SITE02 | 0 | 0 | 0.422 | 0.673 |  |  |
| LV_HI.WITH | SITE03 | 0 | 0.001 | -0.089 | 0.929 | 0 | 1 |
| LV_HI.WITH | SITE04 | 0 | 0 | 1.316 | 0.188 |  |  |
| LV_HI.WITH | SITE05 | -0.001 | 0.001 | -0.928 | 0.353 | -1 | 0.317311 |
| LV_HI.WITH | SITE06 | 0.001 | 0 | 1.639 | 0.101 |  |  |
| LV_HI.WITH | SITE07 | 0 | 0.001 | -0.108 | 0.914 | 0 | 1 |
| LV_HI.WITH | SITE08 | 0 | 0 | -0.205 | 0.837 |  |  |
| LV_HI.WITH | SITE09 | 0 | 0.001 | -0.623 | 0.533 | 0 | 1 |
| CHR.WITH | AGE | -0.075 | 0.079 | -0.958 | 0.338 | -0.94937 | 0.342434 |
| CHR.WITH | FEMALE | -0.006 | 0.009 | -0.685 | 0.494 | -0.66667 | 0.504985 |
| CHR.WITH | SITE02 | -0.003 | 0.006 | -0.576 | 0.565 | -0.5 | 0.617075 |
| CHR.WITH | SITE03 | -0.003 | 0.005 | -0.645 | 0.519 | -0.6 | 0.548506 |
| CHR.WITH | SITE04 | 0.001 | 0.005 | 0.29 | 0.771 | 0.2 | 0.841481 |
| CHR.WITH | SITE05 | -0.001 | 0.006 | -0.092 | 0.927 | -0.16667 | 0.867632 |
| CHR.WITH | SITE06 | -0.002 | 0.006 | -0.352 | 0.725 | -0.33333 | 0.738883 |
| CHR.WITH | SITE07 | -0.001 | 0.006 | -0.092 | 0.927 | -0.16667 | 0.867632 |
| CHR.WITH | SITE08 | 0.003 | 0.005 | 0.532 | 0.595 | 0.6 | 0.548506 |
| CHR.WITH | SITE09 | 0.001 | 0.006 | 0.165 | 0.869 | 0.166667 | 0.867632 |
| AGE.WITH | FEMALE | -0.219 | 0.095 | -2.307 | 0.021 | -2.30526 | 0.021152 |
| AGE.WITH | SITE02 | 0.346 | 0.077 | 4.508 | 0 | 4.493506 | 7.01E-06 |
| AGE.WITH | SITE03 | 0 | 0.042 | 0.007 | 0.995 | 0 | 1 |
| AGE.WITH | SITE04 | -0.011 | 0.044 | -0.255 | 0.799 | -0.25 | 0.802587 |
| AGE.WITH | SITE05 | 0.209 | 0.062 | 3.349 | 0.001 | 3.370968 | 0.000749 |
| AGE.WITH | SITE06 | -0.012 | 0.054 | -0.215 | 0.829 | -0.22222 | 0.824141 |
| AGE.WITH | SITE07 | -0.198 | 0.066 | -2.984 | 0.003 | -3 | 0.0027 |
| AGE.WITH | SITE08 | -0.211 | 0.065 | -3.237 | 0.001 | -3.24615 | 0.00117 |
| AGE.WITH | SITE09 | -0.042 | 0.063 | -0.67 | 0.503 | -0.66667 | 0.504985 |
| FEMALE.WITH | SITE02 | 0.005 | 0.007 | 0.702 | 0.483 | 0.714286 | 0.475051 |
| FEMALE.WITH | SITE03 | -0.002 | 0.006 | -0.372 | 0.71 | -0.33333 | 0.738883 |
| FEMALE.WITH | SITE04 | 0 | 0.007 | 0.017 | 0.987 | 0 | 1 |
| FEMALE.WITH | SITE05 | 0.001 | 0.008 | 0.143 | 0.886 | 0.125 | 0.900524 |
| FEMALE.WITH | SITE06 | -0.011 | 0.008 | -1.358 | 0.174 | -1.375 | 0.169131 |
| FEMALE.WITH | SITE07 | 0.01 | 0.008 | 1.249 | 0.212 | 1.25 | 0.2113 |
| FEMALE.WITH | SITE08 | 0 | 0.007 | -0.023 | 0.981 | 0 | 1 |
| FEMALE.WITH | SITE09 | 0.002 | 0.009 | 0.184 | 0.854 | 0.222222 | 0.824141 |
| SITE02.WITH | SITE03 | -0.005 | 0.001 | -4.219 | 0 | -5 | 5.73E-07 |
| SITE02.WITH | SITE04 | -0.007 | 0.002 | -4.727 | 0 | -3.5 | 0.000465 |
| SITE02.WITH | SITE05 | -0.012 | 0.002 | -5.392 | 0 | -6 | 1.97E-09 |
| SITE02.WITH | SITE06 | -0.011 | 0.002 | -5.301 | 0 | -5.5 | 3.8E-08 |
| SITE02.WITH | SITE07 | -0.012 | 0.002 | -5.392 | 0 | -6 | 1.97E-09 |
| SITE02.WITH | SITE08 | -0.009 | 0.002 | -5.034 | 0 | -4.5 | 6.8E-06 |
| SITE02.WITH | SITE09 | -0.013 | 0.002 | -5.476 | 0 | -6.5 | 8.03E-11 |
| SITE03.WITH | SITE04 | -0.005 | 0.001 | -4.219 | 0 | -5 | 5.73E-07 |
| SITE03.WITH | SITE05 | -0.008 | 0.002 | -4.671 | 0 | -4 | 6.33E-05 |
| SITE03.WITH | SITE06 | -0.008 | 0.002 | -4.612 | 0 | -4 | 6.33E-05 |
| SITE03.WITH | SITE07 | -0.008 | 0.002 | -4.671 | 0 | -4 | 6.33E-05 |
| SITE03.WITH | SITE08 | -0.006 | 0.001 | -4.432 | 0 | -6 | 1.97E-09 |
| SITE03.WITH | SITE09 | -0.009 | 0.002 | -4.725 | 0 | -4.5 | 6.8E-06 |
| SITE04.WITH | SITE05 | -0.012 | 0.002 | -5.392 | 0 | -6 | 1.97E-09 |
| SITE04.WITH | SITE06 | -0.011 | 0.002 | -5.301 | 0 | -5.5 | 3.8E-08 |
| SITE04.WITH | SITE07 | -0.012 | 0.002 | -5.392 | 0 | -6 | 1.97E-09 |
| SITE04.WITH | SITE08 | -0.009 | 0.002 | -5.034 | 0 | -4.5 | 6.8E-06 |
| SITE04.WITH | SITE09 | -0.013 | 0.002 | -5.476 | 0 | -6.5 | 8.03E-11 |
| SITE05.WITH | SITE06 | -0.018 | 0.003 | -6.297 | 0 | -6 | 1.97E-09 |
| SITE05.WITH | SITE07 | -0.019 | 0.003 | -6.451 | 0 | -6.33333 | 2.4E-10 |
| SITE05.WITH | SITE08 | -0.015 | 0.003 | -5.862 | 0 | -5 | 5.73E-07 |
| SITE05.WITH | SITE09 | -0.021 | 0.003 | -6.595 | 0 | -7 | 2.56E-12 |
| SITE06.WITH | SITE07 | -0.018 | 0.003 | -6.297 | 0 | -6 | 1.97E-09 |
| SITE06.WITH | SITE08 | -0.014 | 0.002 | -5.745 | 0 | -7 | 2.56E-12 |
| SITE06.WITH | SITE09 | -0.019 | 0.003 | -6.431 | 0 | -6.33333 | 2.4E-10 |
| SITE07.WITH | SITE08 | -0.015 | 0.003 | -5.862 | 0 | -5 | 5.73E-07 |
| SITE07.WITH | SITE09 | -0.021 | 0.003 | -6.595 | 0 | -7 | 2.56E-12 |
| SITE08.WITH | SITE09 | -0.016 | 0.003 | -5.97 | 0 | -5.33333 | 9.64E-08 |
| Means | CHR | 0.821 | 0.018 | 44.399 | 0 | 45.61111 | 0 |
| Means | AGE | 19.053 | 0.192 | 99.253 | 0 | 99.23438 | 0 |
| Means | FEMALE | 0.458 | 0.024 | 19.066 | 0 | 19.08333 | 0 |
| Means | SITE02 | 0.086 | 0.014 | 6.363 | 0 | 6.142857 | 8.11E-10 |
| Means | SITE03 | 0.06 | 0.011 | 5.261 | 0 | 5.454545 | 4.91E-08 |
| Means | SITE04 | 0.086 | 0.014 | 6.363 | 0 | 6.142857 | 8.11E-10 |
| Means | SITE05 | 0.14 | 0.017 | 8.35 | 0 | 8.235294 | 2.22E-16 |
| Means | SITE06 | 0.13 | 0.016 | 8.024 | 0 | 8.125 | 4.44E-16 |
| Means | SITE07 | 0.14 | 0.017 | 8.35 | 0 | 8.235294 | 2.22E-16 |
| Means | SITE08 | 0.107 | 0.015 | 7.177 | 0 | 7.133333 | 9.8E-13 |
| Means | SITE09 | 0.149 | 0.017 | 8.671 | 0 | 8.764706 | 0 |
| Intercepts | SYCOD_CH | 2.32 | 0.647 | 3.584 | 0 | 3.585781 | 0.000336 |
| Intercepts | HI_CH | -0.002 | 0.002 | -1.086 | 0.278 | -1 | 0.317311 |
| Variances | CHR | 0.147 | 0.012 | 12.387 | 0 | 12.25 | 0 |
| Variances | AGE | 15.846 | 1.107 | 14.31 | 0 | 14.31436 | 0 |
| Variances | FEMALE | 0.248 | 0.002 | 123.408 | 0 | 124 | 0 |
| Variances | SITE02 | 0.079 | 0.011 | 7.024 | 0 | 7.181818 | 6.88E-13 |
| Variances | SITE03 | 0.057 | 0.01 | 5.622 | 0 | 5.7 | 1.2E-08 |
| Variances | SITE04 | 0.079 | 0.011 | 7.024 | 0 | 7.181818 | 6.88E-13 |
| Variances | SITE05 | 0.12 | 0.012 | 9.967 | 0 | 10 | 0 |
| Variances | SITE06 | 0.113 | 0.012 | 9.437 | 0 | 9.416667 | 0 |
| Variances | SITE07 | 0.12 | 0.012 | 9.967 | 0 | 10 | 0 |
| Variances | SITE08 | 0.096 | 0.012 | 8.154 | 0 | 8 | 1.33E-15 |
| Variances | SITE09 | 0.127 | 0.012 | 10.509 | 0 | 10.58333 | 0 |
| Variances | LV_HI | 0.001 | 0 | 5.578 | 0 |  |  |
| Residual.Variances | SYCOD_CH | 0.885 | 0 |  |  |  |  |
| Residual.Variances | HI_CH | 0 | 0 |  |  |  |  |
| Residual.Variances | LV_SYCOD | 3.15 | 0.546 | 5.772 | 0 | 5.769231 | 7.96E-09 |
| New.Additional.Parameters | EFFECT_C | 11.152 | 11.208 | 0.995 | 0.32 | 0.995004 | 0.319735 |
| New.Additional.Parameters | EFFECT_P | 5.888 | 7.327 | 0.804 | 0.422 | 0.803603 | 0.421626 |
| LV_HI.BY | HI_CH | 1 | 0 |  |  |  |  |
| LV_VEMEM.BY | VEMEM_CH | 1 | 0 |  |  |  |  |
| LV_VEMEM.ON | LV_HI | 2.148 | 2.039 | 1.053 | 0.292 | 1.053458 | 0.292131 |
| LV_VEMEM.ON | HIXCHR | -5.772 | 2.957 | -1.952 | 0.051 | -1.95198 | 0.050941 |
| LV_VEMEM.ON | CHR | -0.074 | 0.071 | -1.042 | 0.297 | -1.04225 | 0.297294 |
| LV_VEMEM.ON | AGE | 0.005 | 0.009 | 0.584 | 0.559 | 0.555556 | 0.578515 |
| LV_VEMEM.ON | FEMALE | -0.117 | 0.074 | -1.578 | 0.115 | -1.58108 | 0.113859 |
| LV_VEMEM.ON | SITE02 | 0.078 | 0.178 | 0.441 | 0.659 | 0.438202 | 0.66124 |
| LV_VEMEM.ON | SITE03 | 0.009 | 0.166 | 0.052 | 0.959 | 0.054217 | 0.956762 |
| LV_VEMEM.ON | SITE04 | -0.346 | 0.209 | -1.656 | 0.098 | -1.6555 | 0.097823 |
| LV_VEMEM.ON | SITE05 | 0.059 | 0.133 | 0.448 | 0.654 | 0.443609 | 0.657325 |
| LV_VEMEM.ON | SITE06 | 0.206 | 0.146 | 1.41 | 0.159 | 1.410959 | 0.158257 |
| LV_VEMEM.ON | SITE07 | 0.123 | 0.126 | 0.977 | 0.329 | 0.97619 | 0.32897 |
| LV_VEMEM.ON | SITE08 | 0.107 | 0.162 | 0.658 | 0.51 | 0.660494 | 0.508937 |
| LV_VEMEM.ON | SITE09 | 0.222 | 0.154 | 1.441 | 0.15 | 1.441558 | 0.149427 |
| LV_HI.WITH | CHR | 0 | 0 | -0.077 | 0.939 |  |  |
| LV_HI.WITH | AGE | 0.003 | 0.006 | 0.459 | 0.646 | 0.5 | 0.617075 |
| LV_HI.WITH | FEMALE | -0.001 | 0.001 | -1.283 | 0.2 | -1 | 0.317311 |
| LV_HI.WITH | SITE02 | 0 | 0 | 0.379 | 0.705 |  |  |
| LV_HI.WITH | SITE03 | 0 | 0.001 | -0.063 | 0.95 | 0 | 1 |
| LV_HI.WITH | SITE04 | 0 | 0 | 1.376 | 0.169 |  |  |
| LV_HI.WITH | SITE05 | -0.001 | 0.001 | -0.926 | 0.354 | -1 | 0.317311 |
| LV_HI.WITH | SITE06 | 0.001 | 0 | 1.652 | 0.099 |  |  |
| LV_HI.WITH | SITE07 | 0 | 0.001 | -0.088 | 0.93 | 0 | 1 |
| LV_HI.WITH | SITE08 | 0 | 0 | -0.213 | 0.831 |  |  |
| LV_HI.WITH | SITE09 | 0 | 0.001 | -0.702 | 0.483 | 0 | 1 |
| CHR.WITH | AGE | -0.075 | 0.079 | -0.951 | 0.342 | -0.94937 | 0.342434 |
| CHR.WITH | FEMALE | -0.005 | 0.009 | -0.567 | 0.571 | -0.55556 | 0.578515 |
| CHR.WITH | SITE02 | -0.005 | 0.006 | -0.844 | 0.399 | -0.83333 | 0.404657 |
| CHR.WITH | SITE03 | -0.003 | 0.005 | -0.616 | 0.538 | -0.6 | 0.548506 |
| CHR.WITH | SITE04 | 0.002 | 0.005 | 0.33 | 0.741 | 0.4 | 0.689157 |
| CHR.WITH | SITE05 | 0 | 0.006 | -0.042 | 0.967 | 0 | 1 |
| CHR.WITH | SITE06 | -0.002 | 0.006 | -0.305 | 0.761 | -0.33333 | 0.738883 |
| CHR.WITH | SITE07 | 0 | 0.006 | -0.042 | 0.967 | 0 | 1 |
| CHR.WITH | SITE08 | 0.003 | 0.005 | 0.577 | 0.564 | 0.6 | 0.548506 |
| CHR.WITH | SITE09 | 0.001 | 0.006 | 0.153 | 0.878 | 0.166667 | 0.867632 |
| AGE.WITH | FEMALE | -0.221 | 0.095 | -2.331 | 0.02 | -2.32632 | 0.020002 |
| AGE.WITH | SITE02 | 0.346 | 0.077 | 4.515 | 0 | 4.493506 | 7.01E-06 |
| AGE.WITH | SITE03 | 0 | 0.042 | 0 | 1 | 0 | 1 |
| AGE.WITH | SITE04 | -0.012 | 0.044 | -0.264 | 0.792 | -0.27273 | 0.785063 |
| AGE.WITH | SITE05 | 0.208 | 0.062 | 3.34 | 0.001 | 3.354839 | 0.000794 |
| AGE.WITH | SITE06 | -0.012 | 0.054 | -0.227 | 0.821 | -0.22222 | 0.824141 |
| AGE.WITH | SITE07 | -0.199 | 0.066 | -2.993 | 0.003 | -3.01515 | 0.002569 |
| AGE.WITH | SITE08 | -0.211 | 0.065 | -3.244 | 0.001 | -3.24615 | 0.00117 |
| AGE.WITH | SITE09 | -0.038 | 0.063 | -0.612 | 0.54 | -0.60317 | 0.546393 |
| FEMALE.WITH | SITE02 | 0.004 | 0.007 | 0.539 | 0.59 | 0.571429 | 0.567709 |
| FEMALE.WITH | SITE03 | -0.002 | 0.006 | -0.372 | 0.71 | -0.33333 | 0.738883 |
| FEMALE.WITH | SITE04 | 0 | 0.007 | 0.017 | 0.987 | 0 | 1 |
| FEMALE.WITH | SITE05 | 0.001 | 0.008 | 0.143 | 0.886 | 0.125 | 0.900524 |
| FEMALE.WITH | SITE06 | -0.011 | 0.008 | -1.358 | 0.174 | -1.375 | 0.169131 |
| FEMALE.WITH | SITE07 | 0.01 | 0.008 | 1.249 | 0.212 | 1.25 | 0.2113 |
| FEMALE.WITH | SITE08 | 0 | 0.007 | -0.023 | 0.981 | 0 | 1 |
| FEMALE.WITH | SITE09 | 0.003 | 0.009 | 0.311 | 0.756 | 0.333333 | 0.738883 |
| SITE02.WITH | SITE03 | -0.005 | 0.001 | -4.246 | 0 | -5 | 5.73E-07 |
| SITE02.WITH | SITE04 | -0.008 | 0.002 | -4.765 | 0 | -4 | 6.33E-05 |
| SITE02.WITH | SITE05 | -0.012 | 0.002 | -5.449 | 0 | -6 | 1.97E-09 |
| SITE02.WITH | SITE06 | -0.012 | 0.002 | -5.355 | 0 | -6 | 1.97E-09 |
| SITE02.WITH | SITE07 | -0.012 | 0.002 | -5.449 | 0 | -6 | 1.97E-09 |
| SITE02.WITH | SITE08 | -0.009 | 0.002 | -5.08 | 0 | -4.5 | 6.8E-06 |
| SITE02.WITH | SITE09 | -0.013 | 0.002 | -5.514 | 0 | -6.5 | 8.03E-11 |
| SITE03.WITH | SITE04 | -0.005 | 0.001 | -4.219 | 0 | -5 | 5.73E-07 |
| SITE03.WITH | SITE05 | -0.008 | 0.002 | -4.671 | 0 | -4 | 6.33E-05 |
| SITE03.WITH | SITE06 | -0.008 | 0.002 | -4.612 | 0 | -4 | 6.33E-05 |
| SITE03.WITH | SITE07 | -0.008 | 0.002 | -4.671 | 0 | -4 | 6.33E-05 |
| SITE03.WITH | SITE08 | -0.006 | 0.001 | -4.432 | 0 | -6 | 1.97E-09 |
| SITE03.WITH | SITE09 | -0.009 | 0.002 | -4.712 | 0 | -4.5 | 6.8E-06 |
| SITE04.WITH | SITE05 | -0.012 | 0.002 | -5.392 | 0 | -6 | 1.97E-09 |
| SITE04.WITH | SITE06 | -0.011 | 0.002 | -5.301 | 0 | -5.5 | 3.8E-08 |
| SITE04.WITH | SITE07 | -0.012 | 0.002 | -5.392 | 0 | -6 | 1.97E-09 |
| SITE04.WITH | SITE08 | -0.009 | 0.002 | -5.034 | 0 | -4.5 | 6.8E-06 |
| SITE04.WITH | SITE09 | -0.013 | 0.002 | -5.456 | 0 | -6.5 | 8.03E-11 |
| SITE05.WITH | SITE06 | -0.018 | 0.003 | -6.297 | 0 | -6 | 1.97E-09 |
| SITE05.WITH | SITE07 | -0.019 | 0.003 | -6.451 | 0 | -6.33333 | 2.4E-10 |
| SITE05.WITH | SITE08 | -0.015 | 0.003 | -5.862 | 0 | -5 | 5.73E-07 |
| SITE05.WITH | SITE09 | -0.02 | 0.003 | -6.56 | 0 | -6.66667 | 2.62E-11 |
| SITE06.WITH | SITE07 | -0.018 | 0.003 | -6.297 | 0 | -6 | 1.97E-09 |
| SITE06.WITH | SITE08 | -0.014 | 0.002 | -5.745 | 0 | -7 | 2.56E-12 |
| SITE06.WITH | SITE09 | -0.019 | 0.003 | -6.398 | 0 | -6.33333 | 2.4E-10 |
| SITE07.WITH | SITE08 | -0.015 | 0.003 | -5.862 | 0 | -5 | 5.73E-07 |
| SITE07.WITH | SITE09 | -0.02 | 0.003 | -6.56 | 0 | -6.66667 | 2.62E-11 |
| SITE08.WITH | SITE09 | -0.016 | 0.003 | -5.943 | 0 | -5.33333 | 9.64E-08 |
| Means | CHR | 0.819 | 0.019 | 44.051 | 0 | 43.10526 | 0 |
| Means | AGE | 19.058 | 0.192 | 99.303 | 0 | 99.26042 | 0 |
| Means | FEMALE | 0.458 | 0.024 | 19.066 | 0 | 19.08333 | 0 |
| Means | SITE02 | 0.088 | 0.014 | 6.456 | 0 | 6.285714 | 3.26E-10 |
| Means | SITE03 | 0.06 | 0.011 | 5.261 | 0 | 5.454545 | 4.91E-08 |
| Means | SITE04 | 0.086 | 0.014 | 6.363 | 0 | 6.142857 | 8.11E-10 |
| Means | SITE05 | 0.14 | 0.017 | 8.35 | 0 | 8.235294 | 2.22E-16 |
| Means | SITE06 | 0.13 | 0.016 | 8.024 | 0 | 8.125 | 4.44E-16 |
| Means | SITE07 | 0.14 | 0.017 | 8.35 | 0 | 8.235294 | 2.22E-16 |
| Means | SITE08 | 0.107 | 0.015 | 7.177 | 0 | 7.133333 | 9.8E-13 |
| Means | SITE09 | 0.146 | 0.017 | 8.591 | 0 | 8.588235 | 0 |
| Intercepts | VEMEM_CH | 0.198 | 0.234 | 0.847 | 0.397 | 0.846154 | 0.397467 |
| Intercepts | HI_CH | -0.002 | 0.002 | -1.134 | 0.257 | -1 | 0.317311 |
| Variances | CHR | 0.148 | 0.012 | 12.54 | 0 | 12.33333 | 0 |
| Variances | AGE | 15.837 | 1.107 | 14.31 | 0 | 14.30623 | 0 |
| Variances | FEMALE | 0.248 | 0.002 | 123.408 | 0 | 124 | 0 |
| Variances | SITE02 | 0.081 | 0.011 | 7.149 | 0 | 7.363636 | 1.79E-13 |
| Variances | SITE03 | 0.057 | 0.01 | 5.622 | 0 | 5.7 | 1.2E-08 |
| Variances | SITE04 | 0.079 | 0.011 | 7.024 | 0 | 7.181818 | 6.88E-13 |
| Variances | SITE05 | 0.12 | 0.012 | 9.967 | 0 | 10 | 0 |
| Variances | SITE06 | 0.113 | 0.012 | 9.437 | 0 | 9.416667 | 0 |
| Variances | SITE07 | 0.12 | 0.012 | 9.967 | 0 | 10 | 0 |
| Variances | SITE08 | 0.096 | 0.012 | 8.154 | 0 | 8 | 1.33E-15 |
| Variances | SITE09 | 0.125 | 0.012 | 10.372 | 0 | 10.41667 | 0 |
| Variances | LV_HI | 0.001 | 0 | 5.589 | 0 |  |  |
| Residual.Variances | VEMEM_CH | 0.267 | 0 |  |  |  |  |
| Residual.Variances | HI_CH | 0 | 0 |  |  |  |  |
| Residual.Variances | LV_VEMEM | 0.288 | 0.075 | 3.849 | 0 | 3.84 | 0.000123 |
| New.Additional.Parameters | EFFECT_C | 2.148 | 2.039 | 1.053 | 0.292 | 1.053458 | 0.292131 |
| New.Additional.Parameters | EFFECT_P | -3.624 | 2.183 | -1.66 | 0.097 | -1.6601 | 0.096894 |

Table_S4_MonteCarloSimulation

(next page)

| **paramHeader** | **param** | **population** | **average** | **population_sd** | **average_se** | **mse** | **cover_95** | **pct_sig_coef** |
| --- | --- | --- | --- | --- | --- | --- | --- | --- |
| LV_HI.BY | HI_CH | 1 | 1 | 0 | 0 | 0 | 1 | 0 |
| LV_DEPS.BY | DEPS_CH | 1 | 1 | 0 | 0 | 0 | 1 | 0 |
| LV_DEPS.ON | LV_HI | 0.004 | -0.0051 | 0.167 | 0.1632 | 0.028 | 0.942 | 0.059 |
| LV_DEPS.ON | HIXCHR | -0.417 | -0.4153 | 0.185 | 0.1809 | 0.0342 | 0.942 | 0.634 |
| LV_DEPS.ON | CHR | -0.334 | -0.3331 | 0.1214 | 0.1201 | 0.0147 | 0.945 | 0.79 |
| LV_DEPS.ON | AGE | -0.02 | -0.0197 | 0.0127 | 0.0125 | 0.0002 | 0.944 | 0.359 |
| LV_DEPS.ON | FEMALE | -0.051 | -0.0516 | 0.0946 | 0.0925 | 0.0089 | 0.942 | 0.091 |
| LV_DEPS.ON | SITE02 | -0.101 | -0.0986 | 0.22 | 0.2143 | 0.0484 | 0.943 | 0.085 |
| LV_DEPS.ON | SITE03 | 0.405 | 0.4068 | 0.2352 | 0.2312 | 0.0553 | 0.943 | 0.422 |
| LV_DEPS.ON | SITE04 | 0.116 | 0.1206 | 0.2124 | 0.2064 | 0.0451 | 0.943 | 0.101 |
| LV_DEPS.ON | SITE05 | 0.265 | 0.2652 | 0.1896 | 0.1853 | 0.0359 | 0.944 | 0.309 |
| LV_DEPS.ON | SITE06 | 0.116 | 0.1189 | 0.193 | 0.1888 | 0.0373 | 0.944 | 0.104 |
| LV_DEPS.ON | SITE07 | 0.157 | 0.1596 | 0.188 | 0.1847 | 0.0354 | 0.947 | 0.144 |
| LV_DEPS.ON | SITE08 | 0.083 | 0.0829 | 0.2 | 0.197 | 0.04 | 0.946 | 0.075 |
| LV_DEPS.ON | SITE09 | 0.386 | 0.3861 | 0.1862 | 0.1836 | 0.0347 | 0.946 | 0.557 |
| LV_HI.WITH | CHR | 0 | 0.0001 | 0.0184 | 0.0182 | 0.0003 | 0.951 | 0.049 |
| LV_HI.WITH | AGE | 0.07 | 0.0686 | 0.1857 | 0.1888 | 0.0345 | 0.952 | 0.06 |
| LV_HI.WITH | FEMALE | -0.034 | -0.0339 | 0.0238 | 0.0237 | 0.0006 | 0.95 | 0.296 |
| LV_HI.WITH | SITE02 | 0.006 | 0.0061 | 0.0134 | 0.0134 | 0.0002 | 0.95 | 0.072 |
| LV_HI.WITH | SITE03 | -0.002 | -0.0019 | 0.0112 | 0.0112 | 0.0001 | 0.951 | 0.05 |
| LV_HI.WITH | SITE04 | 0.015 | 0.0151 | 0.0134 | 0.0134 | 0.0002 | 0.95 | 0.2 |
| LV_HI.WITH | SITE05 | -0.017 | -0.0167 | 0.0164 | 0.0165 | 0.0003 | 0.952 | 0.172 |
| LV_HI.WITH | SITE06 | 0.02 | 0.0197 | 0.0159 | 0.0159 | 0.0003 | 0.951 | 0.236 |
| LV_HI.WITH | SITE07 | -0.003 | -0.0033 | 0.0166 | 0.0163 | 0.0003 | 0.947 | 0.057 |
| LV_HI.WITH | SITE08 | -0.003 | -0.0029 | 0.0147 | 0.0146 | 0.0002 | 0.946 | 0.058 |
| LV_HI.WITH | SITE09 | -0.012 | -0.012 | 0.0167 | 0.0168 | 0.0003 | 0.95 | 0.108 |
| CHR.WITH | AGE | -0.074 | -0.0741 | 0.0728 | 0.0729 | 0.0053 | 0.951 | 0.171 |
| CHR.WITH | FEMALE | -0.006 | -0.0059 | 0.0091 | 0.0091 | 0.0001 | 0.951 | 0.102 |
| CHR.WITH | SITE02 | -0.005 | -0.005 | 0.0052 | 0.0052 | 0 | 0.947 | 0.166 |
| CHR.WITH | SITE03 | -0.003 | -0.0029 | 0.0043 | 0.0043 | 0 | 0.951 | 0.106 |
| CHR.WITH | SITE04 | 0.002 | 0.002 | 0.0052 | 0.0052 | 0 | 0.951 | 0.071 |
| CHR.WITH | SITE05 | 0 | 0 | 0.0064 | 0.0064 | 0 | 0.95 | 0.051 |
| CHR.WITH | SITE06 | -0.002 | -0.002 | 0.0061 | 0.0061 | 0 | 0.949 | 0.06 |
| CHR.WITH | SITE07 | -0.001 | -0.001 | 0.0064 | 0.0063 | 0 | 0.948 | 0.056 |
| CHR.WITH | SITE08 | 0.003 | 0.003 | 0.0057 | 0.0056 | 0 | 0.946 | 0.083 |
| CHR.WITH | SITE09 | 0.001 | 0.001 | 0.0065 | 0.0065 | 0 | 0.946 | 0.057 |
| AGE.WITH | FEMALE | -0.218 | -0.216 | 0.0964 | 0.0952 | 0.0093 | 0.947 | 0.62 |
| AGE.WITH | SITE02 | 0.342 | 0.3407 | 0.0565 | 0.0561 | 0.0032 | 0.947 | 1 |
| AGE.WITH | SITE03 | 0 | -0.0005 | 0.0451 | 0.0449 | 0.002 | 0.95 | 0.05 |
| AGE.WITH | SITE04 | -0.015 | -0.0142 | 0.0543 | 0.0537 | 0.0029 | 0.947 | 0.061 |
| AGE.WITH | SITE05 | 0.215 | 0.2137 | 0.0675 | 0.0668 | 0.0046 | 0.949 | 0.898 |
| AGE.WITH | SITE06 | -0.012 | -0.0128 | 0.064 | 0.0635 | 0.0041 | 0.948 | 0.054 |
| AGE.WITH | SITE07 | -0.196 | -0.1956 | 0.0671 | 0.0662 | 0.0045 | 0.946 | 0.849 |
| AGE.WITH | SITE08 | -0.208 | -0.2065 | 0.0598 | 0.0594 | 0.0036 | 0.949 | 0.941 |
| AGE.WITH | SITE09 | -0.042 | -0.0404 | 0.0681 | 0.0671 | 0.0046 | 0.951 | 0.095 |
| FEMALE.WITH | SITE02 | 0.004 | 0.004 | 0.0067 | 0.0067 | 0 | 0.954 | 0.094 |
| FEMALE.WITH | SITE03 | -0.002 | -0.002 | 0.0056 | 0.0056 | 0 | 0.951 | 0.064 |
| FEMALE.WITH | SITE04 | 0 | 0 | 0.0068 | 0.0067 | 0 | 0.948 | 0.052 |
| FEMALE.WITH | SITE05 | 0.001 | 0.001 | 0.0083 | 0.0083 | 0.0001 | 0.948 | 0.054 |
| FEMALE.WITH | SITE06 | -0.01 | -0.01 | 0.008 | 0.008 | 0.0001 | 0.947 | 0.244 |
| FEMALE.WITH | SITE07 | 0.011 | 0.0111 | 0.0082 | 0.0082 | 0.0001 | 0.95 | 0.267 |
| FEMALE.WITH | SITE08 | 0 | -0.0001 | 0.0074 | 0.0073 | 0.0001 | 0.949 | 0.051 |
| FEMALE.WITH | SITE09 | 0.002 | 0.002 | 0.0085 | 0.0084 | 0.0001 | 0.948 | 0.058 |
| SITE02.WITH | SITE03 | -0.005 | -0.0051 | 0.0033 | 0.0032 | 0 | 0.945 | 0.354 |
| SITE02.WITH | SITE04 | -0.008 | -0.008 | 0.0038 | 0.0038 | 0 | 0.949 | 0.547 |
| SITE02.WITH | SITE05 | -0.012 | -0.012 | 0.0047 | 0.0047 | 0 | 0.949 | 0.725 |
| SITE02.WITH | SITE06 | -0.011 | -0.011 | 0.0046 | 0.0045 | 0 | 0.947 | 0.681 |
| SITE02.WITH | SITE07 | -0.012 | -0.012 | 0.0047 | 0.0047 | 0 | 0.943 | 0.729 |
| SITE02.WITH | SITE08 | -0.009 | -0.0089 | 0.0042 | 0.0042 | 0 | 0.949 | 0.575 |
| SITE02.WITH | SITE09 | -0.013 | -0.0129 | 0.0048 | 0.0048 | 0 | 0.949 | 0.773 |
| SITE03.WITH | SITE04 | -0.005 | -0.005 | 0.0032 | 0.0032 | 0 | 0.949 | 0.347 |
| SITE03.WITH | SITE05 | -0.008 | -0.008 | 0.0039 | 0.0039 | 0 | 0.949 | 0.531 |
| SITE03.WITH | SITE06 | -0.008 | -0.008 | 0.0038 | 0.0038 | 0 | 0.948 | 0.558 |
| SITE03.WITH | SITE07 | -0.008 | -0.0079 | 0.004 | 0.0039 | 0 | 0.946 | 0.527 |
| SITE03.WITH | SITE08 | -0.006 | -0.006 | 0.0035 | 0.0035 | 0 | 0.951 | 0.405 |
| SITE03.WITH | SITE09 | -0.009 | -0.009 | 0.0041 | 0.004 | 0 | 0.944 | 0.617 |
| SITE04.WITH | SITE05 | -0.012 | -0.012 | 0.0048 | 0.0047 | 0 | 0.946 | 0.726 |
| SITE04.WITH | SITE06 | -0.011 | -0.011 | 0.0045 | 0.0045 | 0 | 0.947 | 0.679 |
| SITE04.WITH | SITE07 | -0.012 | -0.0121 | 0.0047 | 0.0047 | 0 | 0.948 | 0.734 |
| SITE04.WITH | SITE08 | -0.009 | -0.009 | 0.0042 | 0.0042 | 0 | 0.947 | 0.581 |
| SITE04.WITH | SITE09 | -0.013 | -0.0129 | 0.0048 | 0.0048 | 0 | 0.95 | 0.776 |
| SITE05.WITH | SITE06 | -0.018 | -0.0179 | 0.0057 | 0.0056 | 0 | 0.946 | 0.896 |
| SITE05.WITH | SITE07 | -0.019 | -0.019 | 0.0058 | 0.0058 | 0 | 0.945 | 0.913 |
| SITE05.WITH | SITE08 | -0.015 | -0.015 | 0.0052 | 0.0052 | 0 | 0.946 | 0.834 |
| SITE05.WITH | SITE09 | -0.021 | -0.0209 | 0.0059 | 0.0059 | 0 | 0.951 | 0.948 |
| SITE06.WITH | SITE07 | -0.018 | -0.0179 | 0.0056 | 0.0056 | 0 | 0.947 | 0.9 |
| SITE06.WITH | SITE08 | -0.014 | -0.0139 | 0.005 | 0.005 | 0 | 0.947 | 0.799 |
| SITE06.WITH | SITE09 | -0.019 | -0.0191 | 0.0057 | 0.0057 | 0 | 0.952 | 0.928 |
| SITE07.WITH | SITE08 | -0.015 | -0.015 | 0.0052 | 0.0051 | 0 | 0.949 | 0.841 |
| SITE07.WITH | SITE09 | -0.02 | -0.0199 | 0.0059 | 0.0059 | 0 | 0.947 | 0.928 |
| SITE08.WITH | SITE09 | -0.016 | -0.016 | 0.0053 | 0.0052 | 0 | 0.948 | 0.869 |
| Means | CHR | 0.821 | 0.8212 | 0.0185 | 0.0184 | 0.0003 | 0.947 | 1 |
| Means | AGE | 19.056 | 19.0574 | 0.1916 | 0.1906 | 0.0367 | 0.95 | 1 |
| Means | FEMALE | 0.453 | 0.4529 | 0.0237 | 0.0238 | 0.0006 | 0.949 | 1 |
| Means | SITE02 | 0.087 | 0.0872 | 0.0135 | 0.0135 | 0.0002 | 0.95 | 1 |
| Means | SITE03 | 0.06 | 0.0602 | 0.0113 | 0.0113 | 0.0001 | 0.948 | 1 |
| Means | SITE04 | 0.087 | 0.0868 | 0.0135 | 0.0135 | 0.0002 | 0.951 | 1 |
| Means | SITE05 | 0.14 | 0.1398 | 0.0166 | 0.0166 | 0.0003 | 0.948 | 1 |
| Means | SITE06 | 0.129 | 0.1289 | 0.0164 | 0.016 | 0.0003 | 0.946 | 1 |
| Means | SITE07 | 0.138 | 0.1379 | 0.0165 | 0.0165 | 0.0003 | 0.949 | 1 |
| Means | SITE08 | 0.106 | 0.106 | 0.0149 | 0.0147 | 0.0002 | 0.949 | 1 |
| Means | SITE09 | 0.147 | 0.1472 | 0.017 | 0.0169 | 0.0003 | 0.95 | 1 |
| Intercepts | DEPS_CH | 0.508 | 0.5005 | 0.3 | 0.2952 | 0.0901 | 0.942 | 0.401 |
| Intercepts | HI_CH | 0 | -0.0002 | 0.0475 | 0.0477 | 0.0023 | 0.948 | 0.052 |
| Variances | CHR | 0.147 | 0.1468 | 0.01 | 0.0099 | 0.0001 | 0.941 | 1 |
| Variances | AGE | 15.868 | 15.8285 | 1.0755 | 1.0672 | 1.1581 | 0.942 | 1 |
| Variances | FEMALE | 0.248 | 0.2477 | 0.0168 | 0.0167 | 0.0003 | 0.943 | 1 |
| Variances | SITE02 | 0.08 | 0.0798 | 0.0054 | 0.0054 | 0 | 0.943 | 1 |
| Variances | SITE03 | 0.056 | 0.0559 | 0.0038 | 0.0038 | 0 | 0.942 | 1 |
| Variances | SITE04 | 0.08 | 0.0799 | 0.0054 | 0.0054 | 0 | 0.946 | 1 |
| Variances | SITE05 | 0.121 | 0.1207 | 0.0082 | 0.0081 | 0.0001 | 0.943 | 1 |
| Variances | SITE06 | 0.112 | 0.1117 | 0.0075 | 0.0075 | 0.0001 | 0.946 | 1 |
| Variances | SITE07 | 0.119 | 0.1188 | 0.0081 | 0.008 | 0.0001 | 0.941 | 1 |
| Variances | SITE08 | 0.095 | 0.0947 | 0.0063 | 0.0064 | 0 | 0.945 | 1 |
| Variances | SITE09 | 0.125 | 0.1246 | 0.0085 | 0.0084 | 0.0001 | 0.945 | 1 |
| Variances | LV_HI | 0.653 | 0.6486 | 0.0668 | 0.0665 | 0.0045 | 0.942 | 1 |
| Residual.Variances | DEPS_CH | 0.253 | 0.2526 | 0 | 0 | 0 | 1 | 0 |
| Residual.Variances | HI_CH | 0.339 | 0.339 | 0 | 0 | 0 | 1 | 0 |
| Residual.Variances | LV_DEPS | 0.598 | 0.5665 | 0.0599 | 0.0586 | 0.0046 | 0.889 | 1 |
| LV_HI.BY | HI_CH | 1 | 1 | 0 | 0 | 0 | 1 | 0 |
| LV_GAF.BY | GAF_CH | 1 | 1 | 0 | 0 | 0 | 1 | 0 |
| LV_GAF.ON | LV_HI | -0.031 | -0.0247 | 0.1649 | 0.162 | 0.0272 | 0.941 | 0.06 |
| LV_GAF.ON | HIXCHR | 0.471 | 0.4722 | 0.1836 | 0.1803 | 0.0337 | 0.941 | 0.74 |
| LV_GAF.ON | CHR | 0.296 | 0.295 | 0.1226 | 0.1205 | 0.015 | 0.944 | 0.688 |
| LV_GAF.ON | AGE | -0.024 | -0.0238 | 0.0129 | 0.0125 | 0.0002 | 0.941 | 0.488 |
| LV_GAF.ON | FEMALE | -0.002 | -0.0009 | 0.0949 | 0.0926 | 0.009 | 0.944 | 0.057 |
| LV_GAF.ON | SITE02 | -0.246 | -0.2475 | 0.2185 | 0.2136 | 0.0477 | 0.944 | 0.221 |
| LV_GAF.ON | SITE03 | -0.41 | -0.4096 | 0.234 | 0.2307 | 0.0548 | 0.944 | 0.427 |
| LV_GAF.ON | SITE04 | -0.141 | -0.1413 | 0.2105 | 0.2058 | 0.0443 | 0.942 | 0.114 |
| LV_GAF.ON | SITE05 | -0.137 | -0.1373 | 0.1886 | 0.1849 | 0.0356 | 0.946 | 0.12 |
| LV_GAF.ON | SITE06 | -0.374 | -0.3754 | 0.1931 | 0.1882 | 0.0373 | 0.942 | 0.516 |
| LV_GAF.ON | SITE07 | -0.36 | -0.3572 | 0.1871 | 0.1843 | 0.035 | 0.943 | 0.492 |
| LV_GAF.ON | SITE08 | -0.027 | -0.0289 | 0.2 | 0.1967 | 0.04 | 0.946 | 0.06 |
| LV_GAF.ON | SITE09 | -0.393 | -0.3936 | 0.1844 | 0.1821 | 0.034 | 0.945 | 0.578 |
| LV_HI.WITH | CHR | 0 | 0.0001 | 0.0184 | 0.0182 | 0.0003 | 0.95 | 0.05 |
| LV_HI.WITH | AGE | 0.06 | 0.0585 | 0.186 | 0.1891 | 0.0346 | 0.956 | 0.055 |
| LV_HI.WITH | FEMALE | -0.036 | -0.0359 | 0.0238 | 0.0237 | 0.0006 | 0.95 | 0.327 |
| LV_HI.WITH | SITE02 | 0.005 | 0.005 | 0.0135 | 0.0134 | 0.0002 | 0.95 | 0.07 |
| LV_HI.WITH | SITE03 | -0.002 | -0.0019 | 0.0112 | 0.0112 | 0.0001 | 0.95 | 0.054 |
| LV_HI.WITH | SITE04 | 0.015 | 0.0151 | 0.0134 | 0.0134 | 0.0002 | 0.951 | 0.194 |
| LV_HI.WITH | SITE05 | -0.018 | -0.0177 | 0.0164 | 0.0165 | 0.0003 | 0.95 | 0.183 |
| LV_HI.WITH | SITE06 | 0.019 | 0.0187 | 0.016 | 0.0159 | 0.0003 | 0.949 | 0.223 |
| LV_HI.WITH | SITE07 | -0.002 | -0.0023 | 0.0166 | 0.0164 | 0.0003 | 0.947 | 0.054 |
| LV_HI.WITH | SITE08 | -0.003 | -0.0029 | 0.0148 | 0.0146 | 0.0002 | 0.946 | 0.058 |
| LV_HI.WITH | SITE09 | -0.012 | -0.012 | 0.0168 | 0.0169 | 0.0003 | 0.951 | 0.107 |
| CHR.WITH | AGE | -0.073 | -0.0727 | 0.0734 | 0.073 | 0.0054 | 0.948 | 0.171 |
| CHR.WITH | FEMALE | -0.006 | -0.0059 | 0.0091 | 0.0091 | 0.0001 | 0.95 | 0.098 |
| CHR.WITH | SITE02 | -0.005 | -0.005 | 0.0053 | 0.0052 | 0 | 0.944 | 0.159 |
| CHR.WITH | SITE03 | -0.003 | -0.0029 | 0.0043 | 0.0043 | 0 | 0.95 | 0.102 |
| CHR.WITH | SITE04 | 0.002 | 0.002 | 0.0052 | 0.0052 | 0 | 0.951 | 0.068 |
| CHR.WITH | SITE05 | 0 | 0 | 0.0064 | 0.0064 | 0 | 0.949 | 0.051 |
| CHR.WITH | SITE06 | -0.002 | -0.002 | 0.0061 | 0.0061 | 0 | 0.949 | 0.061 |
| CHR.WITH | SITE07 | 0 | 0 | 0.0063 | 0.0063 | 0 | 0.949 | 0.051 |
| CHR.WITH | SITE08 | 0.003 | 0.003 | 0.0057 | 0.0056 | 0 | 0.951 | 0.085 |
| CHR.WITH | SITE09 | 0.001 | 0.001 | 0.0065 | 0.0065 | 0 | 0.944 | 0.057 |
| AGE.WITH | FEMALE | -0.221 | -0.2189 | 0.0963 | 0.0954 | 0.0093 | 0.949 | 0.635 |
| AGE.WITH | SITE02 | 0.342 | 0.3407 | 0.0562 | 0.0562 | 0.0032 | 0.947 | 1 |
| AGE.WITH | SITE03 | 0 | -0.0007 | 0.0451 | 0.045 | 0.002 | 0.951 | 0.049 |
| AGE.WITH | SITE04 | -0.015 | -0.0142 | 0.0546 | 0.0538 | 0.003 | 0.948 | 0.061 |
| AGE.WITH | SITE05 | 0.214 | 0.2127 | 0.0671 | 0.0669 | 0.0045 | 0.95 | 0.898 |
| AGE.WITH | SITE06 | -0.013 | -0.0138 | 0.0639 | 0.0637 | 0.0041 | 0.949 | 0.056 |
| AGE.WITH | SITE07 | -0.197 | -0.1967 | 0.0674 | 0.0663 | 0.0045 | 0.944 | 0.848 |
| AGE.WITH | SITE08 | -0.21 | -0.2083 | 0.0595 | 0.0595 | 0.0035 | 0.95 | 0.947 |
| AGE.WITH | SITE09 | -0.043 | -0.0416 | 0.069 | 0.0675 | 0.0048 | 0.944 | 0.098 |
| FEMALE.WITH | SITE02 | 0.004 | 0.004 | 0.0067 | 0.0067 | 0 | 0.947 | 0.093 |
| FEMALE.WITH | SITE03 | -0.002 | -0.002 | 0.0056 | 0.0056 | 0 | 0.949 | 0.065 |
| FEMALE.WITH | SITE04 | -0.001 | -0.001 | 0.0068 | 0.0067 | 0 | 0.946 | 0.055 |
| FEMALE.WITH | SITE05 | 0.001 | 0.001 | 0.0084 | 0.0083 | 0.0001 | 0.949 | 0.054 |
| FEMALE.WITH | SITE06 | -0.01 | -0.01 | 0.0081 | 0.008 | 0.0001 | 0.948 | 0.243 |
| FEMALE.WITH | SITE07 | 0.011 | 0.0111 | 0.0082 | 0.0082 | 0.0001 | 0.948 | 0.269 |
| FEMALE.WITH | SITE08 | 0 | -0.0001 | 0.0074 | 0.0073 | 0.0001 | 0.949 | 0.051 |
| FEMALE.WITH | SITE09 | 0.002 | 0.002 | 0.0084 | 0.0084 | 0.0001 | 0.949 | 0.058 |
| SITE02.WITH | SITE03 | -0.005 | -0.005 | 0.0033 | 0.0032 | 0 | 0.944 | 0.354 |
| SITE02.WITH | SITE04 | -0.008 | -0.008 | 0.0038 | 0.0038 | 0 | 0.954 | 0.553 |
| SITE02.WITH | SITE05 | -0.012 | -0.012 | 0.0047 | 0.0047 | 0 | 0.948 | 0.718 |
| SITE02.WITH | SITE06 | -0.011 | -0.011 | 0.0045 | 0.0045 | 0 | 0.954 | 0.681 |
| SITE02.WITH | SITE07 | -0.012 | -0.012 | 0.0048 | 0.0047 | 0 | 0.945 | 0.728 |
| SITE02.WITH | SITE08 | -0.009 | -0.0089 | 0.0042 | 0.0042 | 0 | 0.946 | 0.573 |
| SITE02.WITH | SITE09 | -0.013 | -0.0129 | 0.0049 | 0.0048 | 0 | 0.951 | 0.764 |
| SITE03.WITH | SITE04 | -0.005 | -0.005 | 0.0032 | 0.0032 | 0 | 0.948 | 0.345 |
| SITE03.WITH | SITE05 | -0.008 | -0.008 | 0.0039 | 0.0039 | 0 | 0.952 | 0.527 |
| SITE03.WITH | SITE06 | -0.008 | -0.008 | 0.0038 | 0.0038 | 0 | 0.948 | 0.558 |
| SITE03.WITH | SITE07 | -0.008 | -0.0079 | 0.004 | 0.0039 | 0 | 0.947 | 0.53 |
| SITE03.WITH | SITE08 | -0.006 | -0.006 | 0.0035 | 0.0035 | 0 | 0.951 | 0.399 |
| SITE03.WITH | SITE09 | -0.009 | -0.009 | 0.0041 | 0.004 | 0 | 0.945 | 0.613 |
| SITE04.WITH | SITE05 | -0.012 | -0.012 | 0.0048 | 0.0047 | 0 | 0.944 | 0.724 |
| SITE04.WITH | SITE06 | -0.011 | -0.011 | 0.0045 | 0.0046 | 0 | 0.948 | 0.681 |
| SITE04.WITH | SITE07 | -0.012 | -0.012 | 0.0047 | 0.0047 | 0 | 0.947 | 0.727 |
| SITE04.WITH | SITE08 | -0.009 | -0.009 | 0.0042 | 0.0042 | 0 | 0.949 | 0.576 |
| SITE04.WITH | SITE09 | -0.013 | -0.0129 | 0.0048 | 0.0048 | 0 | 0.95 | 0.77 |
| SITE05.WITH | SITE06 | -0.018 | -0.0179 | 0.0057 | 0.0056 | 0 | 0.948 | 0.894 |
| SITE05.WITH | SITE07 | -0.019 | -0.0191 | 0.0059 | 0.0058 | 0 | 0.944 | 0.913 |
| SITE05.WITH | SITE08 | -0.015 | -0.015 | 0.0052 | 0.0052 | 0 | 0.952 | 0.834 |
| SITE05.WITH | SITE09 | -0.021 | -0.0209 | 0.006 | 0.006 | 0 | 0.948 | 0.942 |
| SITE06.WITH | SITE07 | -0.018 | -0.0179 | 0.0056 | 0.0056 | 0 | 0.945 | 0.9 |
| SITE06.WITH | SITE08 | -0.014 | -0.0139 | 0.005 | 0.005 | 0 | 0.947 | 0.802 |
| SITE06.WITH | SITE09 | -0.019 | -0.0191 | 0.0057 | 0.0057 | 0 | 0.95 | 0.924 |
| SITE07.WITH | SITE08 | -0.015 | -0.0149 | 0.0052 | 0.0051 | 0 | 0.946 | 0.833 |
| SITE07.WITH | SITE09 | -0.02 | -0.0199 | 0.0059 | 0.0059 | 0 | 0.948 | 0.924 |
| SITE08.WITH | SITE09 | -0.016 | -0.016 | 0.0054 | 0.0053 | 0 | 0.945 | 0.859 |
| Means | CHR | 0.82 | 0.8202 | 0.0185 | 0.0184 | 0.0003 | 0.95 | 1 |
| Means | AGE | 19.062 | 19.0634 | 0.192 | 0.191 | 0.0369 | 0.948 | 1 |
| Means | FEMALE | 0.454 | 0.4539 | 0.0235 | 0.0239 | 0.0006 | 0.952 | 1 |
| Means | SITE02 | 0.088 | 0.0882 | 0.0136 | 0.0136 | 0.0002 | 0.951 | 1 |
| Means | SITE03 | 0.06 | 0.0602 | 0.0115 | 0.0113 | 0.0001 | 0.947 | 0.999 |
| Means | SITE04 | 0.088 | 0.0878 | 0.0136 | 0.0136 | 0.0002 | 0.948 | 1 |
| Means | SITE05 | 0.141 | 0.1408 | 0.0168 | 0.0167 | 0.0003 | 0.946 | 1 |
| Means | SITE06 | 0.129 | 0.1289 | 0.0162 | 0.016 | 0.0003 | 0.948 | 1 |
| Means | SITE07 | 0.138 | 0.1379 | 0.0166 | 0.0165 | 0.0003 | 0.948 | 1 |
| Means | SITE08 | 0.106 | 0.106 | 0.015 | 0.0148 | 0.0002 | 0.946 | 1 |
| Means | SITE09 | 0.147 | 0.1472 | 0.0172 | 0.017 | 0.0003 | 0.946 | 1 |
| Intercepts | GAF_CH | 0.453 | 0.4489 | 0.3043 | 0.2951 | 0.0926 | 0.942 | 0.335 |
| Intercepts | HI_CH | 0 | -0.0002 | 0.0479 | 0.0477 | 0.0023 | 0.948 | 0.052 |
| Variances | CHR | 0.147 | 0.1468 | 0.01 | 0.0099 | 0.0001 | 0.946 | 1 |
| Variances | AGE | 15.888 | 15.8469 | 1.0788 | 1.0699 | 1.1653 | 0.942 | 1 |
| Variances | FEMALE | 0.248 | 0.2477 | 0.0168 | 0.0167 | 0.0003 | 0.943 | 1 |
| Variances | SITE02 | 0.08 | 0.0798 | 0.0054 | 0.0054 | 0 | 0.946 | 1 |
| Variances | SITE03 | 0.056 | 0.0559 | 0.0038 | 0.0038 | 0 | 0.942 | 1 |
| Variances | SITE04 | 0.08 | 0.0799 | 0.0054 | 0.0054 | 0 | 0.944 | 1 |
| Variances | SITE05 | 0.121 | 0.1206 | 0.0082 | 0.0081 | 0.0001 | 0.943 | 1 |
| Variances | SITE06 | 0.112 | 0.1117 | 0.0076 | 0.0076 | 0.0001 | 0.944 | 1 |
| Variances | SITE07 | 0.119 | 0.1188 | 0.008 | 0.008 | 0.0001 | 0.943 | 1 |
| Variances | SITE08 | 0.095 | 0.0947 | 0.0064 | 0.0064 | 0 | 0.942 | 1 |
| Variances | SITE09 | 0.126 | 0.1256 | 0.0086 | 0.0085 | 0.0001 | 0.942 | 1 |
| Variances | LV_HI | 0.654 | 0.6499 | 0.0663 | 0.0667 | 0.0044 | 0.945 | 1 |
| Residual.Variances | GAF_CH | 0.172 | 0.1715 | 0 | 0 | 0 | 1 | 0 |
| Residual.Variances | HI_CH | 0.339 | 0.3387 | 0 | 0 | 0 | 1 | 0 |
| Residual.Variances | LV_GAF | 0.672 | 0.6411 | 0.0599 | 0.0586 | 0.0045 | 0.891 | 1 |
| LV_HI.BY | HI_CH | 1 | 1 | 0 | 0 | 0 | 1 | 0 |
| LV_NEGS.BY | NEGS_CH | 1 | 1 | 0 | 0 | 0 | 1 | 0 |
| LV_NEGS.ON | LV_HI | 0.075 | 0.0656 | 0.1645 | 0.16 | 0.0271 | 0.941 | 0.08 |
| LV_NEGS.ON | HIXCHR | -0.539 | -0.5376 | 0.1827 | 0.1783 | 0.0334 | 0.938 | 0.846 |
| LV_NEGS.ON | CHR | -0.304 | -0.3029 | 0.1198 | 0.119 | 0.0144 | 0.944 | 0.722 |
| LV_NEGS.ON | AGE | -0.003 | -0.0027 | 0.0126 | 0.0122 | 0.0002 | 0.942 | 0.063 |
| LV_NEGS.ON | FEMALE | -0.004 | -0.0047 | 0.0929 | 0.0909 | 0.0086 | 0.944 | 0.058 |
| LV_NEGS.ON | SITE02 | -0.492 | -0.4892 | 0.2147 | 0.2094 | 0.0461 | 0.945 | 0.641 |
| LV_NEGS.ON | SITE03 | 0.242 | 0.2428 | 0.2311 | 0.229 | 0.0534 | 0.945 | 0.186 |
| LV_NEGS.ON | SITE04 | -0.375 | -0.3702 | 0.209 | 0.2016 | 0.0437 | 0.938 | 0.455 |
| LV_NEGS.ON | SITE05 | -0.105 | -0.1049 | 0.1858 | 0.1809 | 0.0345 | 0.945 | 0.096 |
| LV_NEGS.ON | SITE06 | -0.277 | -0.2734 | 0.1886 | 0.1834 | 0.0356 | 0.942 | 0.33 |
| LV_NEGS.ON | SITE07 | -0.211 | -0.208 | 0.185 | 0.1802 | 0.0342 | 0.941 | 0.216 |
| LV_NEGS.ON | SITE08 | -0.432 | -0.4327 | 0.1971 | 0.1924 | 0.0389 | 0.945 | 0.608 |
| LV_NEGS.ON | SITE09 | 0.08 | 0.0799 | 0.1803 | 0.1775 | 0.0325 | 0.943 | 0.078 |
| LV_HI.WITH | CHR | -0.001 | -0.0009 | 0.0184 | 0.0182 | 0.0003 | 0.948 | 0.052 |
| LV_HI.WITH | AGE | 0.076 | 0.0747 | 0.1857 | 0.189 | 0.0345 | 0.955 | 0.064 |
| LV_HI.WITH | FEMALE | -0.032 | -0.0319 | 0.0237 | 0.0237 | 0.0006 | 0.95 | 0.269 |
| LV_HI.WITH | SITE02 | 0.007 | 0.0071 | 0.0135 | 0.0134 | 0.0002 | 0.95 | 0.086 |
| LV_HI.WITH | SITE03 | -0.01 | -0.0099 | 0.011 | 0.011 | 0.0001 | 0.95 | 0.143 |
| LV_HI.WITH | SITE04 | 0.016 | 0.0161 | 0.0134 | 0.0134 | 0.0002 | 0.952 | 0.219 |
| LV_HI.WITH | SITE05 | -0.018 | -0.0177 | 0.0164 | 0.0165 | 0.0003 | 0.951 | 0.185 |
| LV_HI.WITH | SITE06 | 0.021 | 0.0207 | 0.016 | 0.0159 | 0.0003 | 0.95 | 0.258 |
| LV_HI.WITH | SITE07 | 0 | -0.0002 | 0.0166 | 0.0163 | 0.0003 | 0.949 | 0.051 |
| LV_HI.WITH | SITE08 | -0.002 | -0.0019 | 0.0148 | 0.0146 | 0.0002 | 0.945 | 0.058 |
| LV_HI.WITH | SITE09 | -0.012 | -0.012 | 0.0169 | 0.0168 | 0.0003 | 0.951 | 0.109 |
| CHR.WITH | AGE | -0.074 | -0.0741 | 0.0734 | 0.0731 | 0.0054 | 0.95 | 0.177 |
| CHR.WITH | FEMALE | -0.006 | -0.0059 | 0.0091 | 0.0091 | 0.0001 | 0.95 | 0.099 |
| CHR.WITH | SITE02 | -0.005 | -0.005 | 0.0052 | 0.0052 | 0 | 0.946 | 0.157 |
| CHR.WITH | SITE03 | -0.003 | -0.0029 | 0.0042 | 0.0043 | 0 | 0.951 | 0.106 |
| CHR.WITH | SITE04 | 0.002 | 0.002 | 0.0052 | 0.0052 | 0 | 0.952 | 0.069 |
| CHR.WITH | SITE05 | 0 | 0 | 0.0064 | 0.0064 | 0 | 0.948 | 0.052 |
| CHR.WITH | SITE06 | -0.002 | -0.002 | 0.0061 | 0.0061 | 0 | 0.951 | 0.06 |
| CHR.WITH | SITE07 | 0 | 0 | 0.0063 | 0.0063 | 0 | 0.951 | 0.049 |
| CHR.WITH | SITE08 | 0.003 | 0.003 | 0.0057 | 0.0056 | 0 | 0.947 | 0.085 |
| CHR.WITH | SITE09 | 0.001 | 0.001 | 0.0065 | 0.0065 | 0 | 0.943 | 0.056 |
| AGE.WITH | FEMALE | -0.219 | -0.2169 | 0.0962 | 0.0954 | 0.0093 | 0.95 | 0.624 |
| AGE.WITH | SITE02 | 0.343 | 0.3416 | 0.0562 | 0.0562 | 0.0032 | 0.947 | 1 |
| AGE.WITH | SITE03 | 0.001 | 0.0005 | 0.0443 | 0.0442 | 0.002 | 0.952 | 0.048 |
| AGE.WITH | SITE04 | -0.015 | -0.0142 | 0.0547 | 0.0539 | 0.003 | 0.948 | 0.061 |
| AGE.WITH | SITE05 | 0.215 | 0.2137 | 0.0671 | 0.067 | 0.0045 | 0.95 | 0.897 |
| AGE.WITH | SITE06 | -0.012 | -0.0128 | 0.0641 | 0.0637 | 0.0041 | 0.949 | 0.054 |
| AGE.WITH | SITE07 | -0.197 | -0.1966 | 0.0675 | 0.0663 | 0.0046 | 0.945 | 0.848 |
| AGE.WITH | SITE08 | -0.209 | -0.2076 | 0.0595 | 0.0595 | 0.0035 | 0.95 | 0.945 |
| AGE.WITH | SITE09 | -0.042 | -0.0404 | 0.0692 | 0.0675 | 0.0048 | 0.945 | 0.095 |
| FEMALE.WITH | SITE02 | 0.004 | 0.004 | 0.0067 | 0.0067 | 0 | 0.949 | 0.093 |
| FEMALE.WITH | SITE03 | -0.001 | -0.001 | 0.0055 | 0.0055 | 0 | 0.949 | 0.054 |
| FEMALE.WITH | SITE04 | -0.001 | -0.001 | 0.0068 | 0.0067 | 0 | 0.947 | 0.056 |
| FEMALE.WITH | SITE05 | 0.001 | 0.001 | 0.0084 | 0.0083 | 0.0001 | 0.949 | 0.054 |
| FEMALE.WITH | SITE06 | -0.01 | -0.01 | 0.0081 | 0.008 | 0.0001 | 0.949 | 0.242 |
| FEMALE.WITH | SITE07 | 0.011 | 0.0111 | 0.0082 | 0.0082 | 0.0001 | 0.948 | 0.268 |
| FEMALE.WITH | SITE08 | 0 | -0.0001 | 0.0074 | 0.0073 | 0.0001 | 0.946 | 0.054 |
| FEMALE.WITH | SITE09 | 0.002 | 0.002 | 0.0085 | 0.0084 | 0.0001 | 0.947 | 0.057 |
| SITE02.WITH | SITE03 | -0.005 | -0.005 | 0.0032 | 0.0031 | 0 | 0.945 | 0.364 |
| SITE02.WITH | SITE04 | -0.008 | -0.008 | 0.0038 | 0.0038 | 0 | 0.956 | 0.551 |
| SITE02.WITH | SITE05 | -0.012 | -0.012 | 0.0047 | 0.0047 | 0 | 0.949 | 0.719 |
| SITE02.WITH | SITE06 | -0.011 | -0.011 | 0.0045 | 0.0045 | 0 | 0.953 | 0.68 |
| SITE02.WITH | SITE07 | -0.012 | -0.012 | 0.0048 | 0.0047 | 0 | 0.943 | 0.73 |
| SITE02.WITH | SITE08 | -0.009 | -0.0089 | 0.0042 | 0.0042 | 0 | 0.947 | 0.576 |
| SITE02.WITH | SITE09 | -0.013 | -0.0129 | 0.0049 | 0.0048 | 0 | 0.951 | 0.762 |
| SITE03.WITH | SITE04 | -0.005 | -0.005 | 0.0032 | 0.0031 | 0 | 0.951 | 0.355 |
| SITE03.WITH | SITE05 | -0.008 | -0.008 | 0.0038 | 0.0039 | 0 | 0.951 | 0.543 |
| SITE03.WITH | SITE06 | -0.007 | -0.007 | 0.0038 | 0.0037 | 0 | 0.947 | 0.468 |
| SITE03.WITH | SITE07 | -0.008 | -0.0079 | 0.0039 | 0.0038 | 0 | 0.946 | 0.543 |
| SITE03.WITH | SITE08 | -0.006 | -0.006 | 0.0034 | 0.0034 | 0 | 0.951 | 0.415 |
| SITE03.WITH | SITE09 | -0.008 | -0.008 | 0.004 | 0.004 | 0 | 0.943 | 0.527 |
| SITE04.WITH | SITE05 | -0.012 | -0.012 | 0.0048 | 0.0047 | 0 | 0.944 | 0.724 |
| SITE04.WITH | SITE06 | -0.011 | -0.011 | 0.0045 | 0.0046 | 0 | 0.949 | 0.682 |
| SITE04.WITH | SITE07 | -0.012 | -0.0121 | 0.0047 | 0.0047 | 0 | 0.945 | 0.727 |
| SITE04.WITH | SITE08 | -0.009 | -0.009 | 0.0042 | 0.0042 | 0 | 0.95 | 0.577 |
| SITE04.WITH | SITE09 | -0.013 | -0.0129 | 0.0048 | 0.0048 | 0 | 0.949 | 0.771 |
| SITE05.WITH | SITE06 | -0.018 | -0.0179 | 0.0057 | 0.0056 | 0 | 0.948 | 0.893 |
| SITE05.WITH | SITE07 | -0.019 | -0.0191 | 0.0059 | 0.0058 | 0 | 0.944 | 0.913 |
| SITE05.WITH | SITE08 | -0.015 | -0.015 | 0.0052 | 0.0052 | 0 | 0.951 | 0.833 |
| SITE05.WITH | SITE09 | -0.021 | -0.0209 | 0.006 | 0.006 | 0 | 0.947 | 0.942 |
| SITE06.WITH | SITE07 | -0.018 | -0.0179 | 0.0056 | 0.0056 | 0 | 0.946 | 0.9 |
| SITE06.WITH | SITE08 | -0.014 | -0.0139 | 0.005 | 0.005 | 0 | 0.946 | 0.804 |
| SITE06.WITH | SITE09 | -0.019 | -0.0191 | 0.0057 | 0.0057 | 0 | 0.95 | 0.927 |
| SITE07.WITH | SITE08 | -0.015 | -0.015 | 0.0052 | 0.0051 | 0 | 0.946 | 0.835 |
| SITE07.WITH | SITE09 | -0.02 | -0.0198 | 0.0059 | 0.0059 | 0 | 0.951 | 0.926 |
| SITE08.WITH | SITE09 | -0.016 | -0.016 | 0.0053 | 0.0053 | 0 | 0.947 | 0.861 |
| Means | CHR | 0.82 | 0.8202 | 0.0185 | 0.0184 | 0.0003 | 0.951 | 1 |
| Means | AGE | 19.057 | 19.0581 | 0.1922 | 0.1911 | 0.037 | 0.948 | 1 |
| Means | FEMALE | 0.454 | 0.4539 | 0.0235 | 0.0239 | 0.0006 | 0.952 | 1 |
| Means | SITE02 | 0.088 | 0.0882 | 0.0136 | 0.0136 | 0.0002 | 0.95 | 1 |
| Means | SITE03 | 0.058 | 0.0582 | 0.0112 | 0.0111 | 0.0001 | 0.947 | 0.999 |
| Means | SITE04 | 0.088 | 0.0878 | 0.0136 | 0.0136 | 0.0002 | 0.948 | 1 |
| Means | SITE05 | 0.141 | 0.1408 | 0.0168 | 0.0167 | 0.0003 | 0.946 | 1 |
| Means | SITE06 | 0.129 | 0.1289 | 0.0162 | 0.016 | 0.0003 | 0.948 | 1 |
| Means | SITE07 | 0.138 | 0.1379 | 0.0166 | 0.0165 | 0.0003 | 0.948 | 1 |
| Means | SITE08 | 0.106 | 0.1061 | 0.0149 | 0.0148 | 0.0002 | 0.948 | 1 |
| Means | SITE09 | 0.147 | 0.1472 | 0.0172 | 0.017 | 0.0003 | 0.945 | 1 |
| Intercepts | NEGS_CH | 0.483 | 0.4754 | 0.2953 | 0.2887 | 0.0873 | 0.943 | 0.385 |
| Intercepts | HI_CH | 0 | -0.0002 | 0.0479 | 0.0477 | 0.0023 | 0.947 | 0.053 |
| Variances | CHR | 0.147 | 0.1468 | 0.01 | 0.0099 | 0.0001 | 0.945 | 1 |
| Variances | AGE | 15.904 | 15.8651 | 1.0763 | 1.071 | 1.1598 | 0.944 | 1 |
| Variances | FEMALE | 0.248 | 0.2477 | 0.0168 | 0.0167 | 0.0003 | 0.943 | 1 |
| Variances | SITE02 | 0.08 | 0.0798 | 0.0054 | 0.0054 | 0 | 0.946 | 1 |
| Variances | SITE03 | 0.054 | 0.0539 | 0.0037 | 0.0036 | 0 | 0.943 | 1 |
| Variances | SITE04 | 0.08 | 0.0799 | 0.0054 | 0.0054 | 0 | 0.945 | 1 |
| Variances | SITE05 | 0.121 | 0.1206 | 0.0082 | 0.0081 | 0.0001 | 0.943 | 1 |
| Variances | SITE06 | 0.112 | 0.1117 | 0.0076 | 0.0076 | 0.0001 | 0.943 | 1 |
| Variances | SITE07 | 0.119 | 0.1188 | 0.008 | 0.008 | 0.0001 | 0.944 | 1 |
| Variances | SITE08 | 0.095 | 0.0947 | 0.0064 | 0.0064 | 0 | 0.942 | 1 |
| Variances | SITE09 | 0.126 | 0.1256 | 0.0086 | 0.0085 | 0.0001 | 0.941 | 1 |
| Variances | LV_HI | 0.649 | 0.645 | 0.0662 | 0.0666 | 0.0044 | 0.947 | 1 |
| Residual.Variances | NEGS_CH | 0.173 | 0.1731 | 0 | 0 | 0 | 1 | 0 |
| Residual.Variances | HI_CH | 0.343 | 0.3425 | 0 | 0 | 0 | 1 | 0 |
| Residual.Variances | LV_NEGS | 0.636 | 0.6049 | 0.0581 | 0.0567 | 0.0043 | 0.888 | 1 |
| LV_HI.BY | HI_CH | 1 | 1 | 0 | 0 | 0 | 1 | 0 |
| LV_POSS.BY | POSS_CH | 1 | 1 | 0 | 0 | 0 | 1 | 0 |
| LV_POSS.ON | LV_HI | -0.092 | -0.0978 | 0.1697 | 0.166 | 0.0288 | 0.943 | 0.103 |
| LV_POSS.ON | HIXCHR | -0.124 | -0.1223 | 0.1874 | 0.1829 | 0.0351 | 0.941 | 0.115 |
| LV_POSS.ON | CHR | -0.628 | -0.6268 | 0.1209 | 0.1192 | 0.0146 | 0.944 | 0.999 |
| LV_POSS.ON | AGE | 0.001 | 0.0012 | 0.0128 | 0.0125 | 0.0002 | 0.944 | 0.057 |
| LV_POSS.ON | FEMALE | 0.12 | 0.1194 | 0.0952 | 0.0928 | 0.0091 | 0.939 | 0.256 |
| LV_POSS.ON | SITE02 | -0.018 | -0.0158 | 0.2209 | 0.2149 | 0.0488 | 0.943 | 0.056 |
| LV_POSS.ON | SITE03 | 0.214 | 0.2168 | 0.2356 | 0.2319 | 0.0555 | 0.944 | 0.163 |
| LV_POSS.ON | SITE04 | 0.121 | 0.1252 | 0.2125 | 0.2069 | 0.0452 | 0.943 | 0.1 |
| LV_POSS.ON | SITE05 | 0.356 | 0.3568 | 0.1899 | 0.1859 | 0.0361 | 0.944 | 0.484 |
| LV_POSS.ON | SITE06 | -0.001 | 0.0014 | 0.1933 | 0.1893 | 0.0374 | 0.945 | 0.055 |
| LV_POSS.ON | SITE07 | 0.115 | 0.1181 | 0.1885 | 0.1853 | 0.0355 | 0.947 | 0.101 |
| LV_POSS.ON | SITE08 | 0.17 | 0.17 | 0.2006 | 0.1976 | 0.0402 | 0.946 | 0.144 |
| LV_POSS.ON | SITE09 | 0.17 | 0.1703 | 0.1859 | 0.1842 | 0.0345 | 0.947 | 0.156 |
| LV_HI.WITH | CHR | 0 | 0.0001 | 0.0185 | 0.0183 | 0.0003 | 0.951 | 0.049 |
| LV_HI.WITH | AGE | 0.063 | 0.0614 | 0.1863 | 0.1894 | 0.0347 | 0.953 | 0.058 |
| LV_HI.WITH | FEMALE | -0.035 | -0.0349 | 0.0238 | 0.0238 | 0.0006 | 0.952 | 0.314 |
| LV_HI.WITH | SITE02 | 0.004 | 0.0041 | 0.0135 | 0.0134 | 0.0002 | 0.949 | 0.06 |
| LV_HI.WITH | SITE03 | -0.002 | -0.0019 | 0.0112 | 0.0113 | 0.0001 | 0.95 | 0.051 |
| LV_HI.WITH | SITE04 | 0.015 | 0.0151 | 0.0135 | 0.0135 | 0.0002 | 0.95 | 0.2 |
| LV_HI.WITH | SITE05 | -0.017 | -0.0167 | 0.0165 | 0.0166 | 0.0003 | 0.952 | 0.172 |
| LV_HI.WITH | SITE06 | 0.02 | 0.0197 | 0.016 | 0.016 | 0.0003 | 0.951 | 0.234 |
| LV_HI.WITH | SITE07 | -0.002 | -0.0023 | 0.0166 | 0.0164 | 0.0003 | 0.947 | 0.054 |
| LV_HI.WITH | SITE08 | -0.004 | -0.0038 | 0.0147 | 0.0147 | 0.0002 | 0.947 | 0.061 |
| LV_HI.WITH | SITE09 | -0.011 | -0.011 | 0.0168 | 0.0168 | 0.0003 | 0.949 | 0.098 |
| CHR.WITH | AGE | -0.074 | -0.0742 | 0.0727 | 0.0729 | 0.0053 | 0.95 | 0.174 |
| CHR.WITH | FEMALE | -0.006 | -0.0059 | 0.0091 | 0.0091 | 0.0001 | 0.95 | 0.1 |
| CHR.WITH | SITE02 | -0.005 | -0.005 | 0.0052 | 0.0052 | 0 | 0.946 | 0.163 |
| CHR.WITH | SITE03 | -0.003 | -0.0029 | 0.0043 | 0.0043 | 0 | 0.951 | 0.104 |
| CHR.WITH | SITE04 | 0.002 | 0.002 | 0.0051 | 0.0052 | 0 | 0.951 | 0.07 |
| CHR.WITH | SITE05 | 0 | 0 | 0.0064 | 0.0064 | 0 | 0.949 | 0.051 |
| CHR.WITH | SITE06 | -0.002 | -0.002 | 0.0061 | 0.0061 | 0 | 0.95 | 0.059 |
| CHR.WITH | SITE07 | -0.001 | -0.001 | 0.0064 | 0.0063 | 0 | 0.946 | 0.056 |
| CHR.WITH | SITE08 | 0.003 | 0.003 | 0.0057 | 0.0056 | 0 | 0.946 | 0.086 |
| CHR.WITH | SITE09 | 0.001 | 0.001 | 0.0065 | 0.0065 | 0 | 0.945 | 0.056 |
| AGE.WITH | FEMALE | -0.218 | -0.2159 | 0.0965 | 0.0952 | 0.0093 | 0.946 | 0.622 |
| AGE.WITH | SITE02 | 0.342 | 0.3408 | 0.0565 | 0.0561 | 0.0032 | 0.947 | 1 |
| AGE.WITH | SITE03 | 0 | -0.0005 | 0.0451 | 0.045 | 0.002 | 0.95 | 0.05 |
| AGE.WITH | SITE04 | -0.015 | -0.0142 | 0.0543 | 0.0538 | 0.0029 | 0.947 | 0.06 |
| AGE.WITH | SITE05 | 0.215 | 0.2138 | 0.0674 | 0.0668 | 0.0045 | 0.949 | 0.898 |
| AGE.WITH | SITE06 | -0.012 | -0.0128 | 0.064 | 0.0636 | 0.0041 | 0.95 | 0.054 |
| AGE.WITH | SITE07 | -0.196 | -0.1956 | 0.067 | 0.0662 | 0.0045 | 0.946 | 0.849 |
| AGE.WITH | SITE08 | -0.208 | -0.2066 | 0.0598 | 0.0594 | 0.0036 | 0.949 | 0.939 |
| AGE.WITH | SITE09 | -0.042 | -0.0406 | 0.0681 | 0.0671 | 0.0046 | 0.949 | 0.093 |
| FEMALE.WITH | SITE02 | 0.004 | 0.004 | 0.0067 | 0.0067 | 0 | 0.952 | 0.094 |
| FEMALE.WITH | SITE03 | -0.002 | -0.002 | 0.0056 | 0.0056 | 0 | 0.951 | 0.065 |
| FEMALE.WITH | SITE04 | 0 | 0 | 0.0068 | 0.0067 | 0 | 0.948 | 0.052 |
| FEMALE.WITH | SITE05 | 0.001 | 0.001 | 0.0083 | 0.0083 | 0.0001 | 0.948 | 0.052 |
| FEMALE.WITH | SITE06 | -0.01 | -0.01 | 0.008 | 0.008 | 0.0001 | 0.947 | 0.242 |
| FEMALE.WITH | SITE07 | 0.011 | 0.0111 | 0.0082 | 0.0082 | 0.0001 | 0.951 | 0.268 |
| FEMALE.WITH | SITE08 | 0 | -0.0001 | 0.0074 | 0.0073 | 0.0001 | 0.949 | 0.051 |
| FEMALE.WITH | SITE09 | 0.002 | 0.002 | 0.0084 | 0.0084 | 0.0001 | 0.948 | 0.057 |
| SITE02.WITH | SITE03 | -0.005 | -0.0051 | 0.0033 | 0.0032 | 0 | 0.945 | 0.354 |
| SITE02.WITH | SITE04 | -0.008 | -0.008 | 0.0038 | 0.0038 | 0 | 0.948 | 0.549 |
| SITE02.WITH | SITE05 | -0.012 | -0.012 | 0.0047 | 0.0047 | 0 | 0.949 | 0.726 |
| SITE02.WITH | SITE06 | -0.011 | -0.011 | 0.0046 | 0.0045 | 0 | 0.946 | 0.683 |
| SITE02.WITH | SITE07 | -0.012 | -0.012 | 0.0047 | 0.0047 | 0 | 0.945 | 0.726 |
| SITE02.WITH | SITE08 | -0.009 | -0.0089 | 0.0042 | 0.0042 | 0 | 0.949 | 0.575 |
| SITE02.WITH | SITE09 | -0.013 | -0.0129 | 0.0048 | 0.0048 | 0 | 0.95 | 0.771 |
| SITE03.WITH | SITE04 | -0.005 | -0.005 | 0.0032 | 0.0032 | 0 | 0.948 | 0.348 |
| SITE03.WITH | SITE05 | -0.008 | -0.008 | 0.0039 | 0.0039 | 0 | 0.949 | 0.532 |
| SITE03.WITH | SITE06 | -0.008 | -0.008 | 0.0038 | 0.0038 | 0 | 0.948 | 0.558 |
| SITE03.WITH | SITE07 | -0.008 | -0.0079 | 0.004 | 0.0039 | 0 | 0.945 | 0.524 |
| SITE03.WITH | SITE08 | -0.006 | -0.006 | 0.0035 | 0.0035 | 0 | 0.951 | 0.404 |
| SITE03.WITH | SITE09 | -0.009 | -0.009 | 0.0041 | 0.004 | 0 | 0.944 | 0.616 |
| SITE04.WITH | SITE05 | -0.012 | -0.012 | 0.0048 | 0.0047 | 0 | 0.947 | 0.724 |
| SITE04.WITH | SITE06 | -0.011 | -0.011 | 0.0045 | 0.0045 | 0 | 0.947 | 0.678 |
| SITE04.WITH | SITE07 | -0.012 | -0.0121 | 0.0047 | 0.0047 | 0 | 0.948 | 0.734 |
| SITE04.WITH | SITE08 | -0.009 | -0.009 | 0.0042 | 0.0042 | 0 | 0.947 | 0.58 |
| SITE04.WITH | SITE09 | -0.013 | -0.0129 | 0.0048 | 0.0048 | 0 | 0.951 | 0.773 |
| SITE05.WITH | SITE06 | -0.018 | -0.0179 | 0.0057 | 0.0056 | 0 | 0.943 | 0.899 |
| SITE05.WITH | SITE07 | -0.019 | -0.019 | 0.0058 | 0.0058 | 0 | 0.947 | 0.913 |
| SITE05.WITH | SITE08 | -0.015 | -0.015 | 0.0052 | 0.0052 | 0 | 0.947 | 0.832 |
| SITE05.WITH | SITE09 | -0.021 | -0.0209 | 0.0059 | 0.0059 | 0 | 0.953 | 0.949 |
| SITE06.WITH | SITE07 | -0.018 | -0.0179 | 0.0056 | 0.0056 | 0 | 0.947 | 0.901 |
| SITE06.WITH | SITE08 | -0.014 | -0.0139 | 0.005 | 0.005 | 0 | 0.945 | 0.798 |
| SITE06.WITH | SITE09 | -0.019 | -0.0191 | 0.0057 | 0.0057 | 0 | 0.951 | 0.927 |
| SITE07.WITH | SITE08 | -0.015 | -0.015 | 0.0052 | 0.0051 | 0 | 0.949 | 0.84 |
| SITE07.WITH | SITE09 | -0.02 | -0.0199 | 0.0059 | 0.0059 | 0 | 0.947 | 0.928 |
| SITE08.WITH | SITE09 | -0.016 | -0.016 | 0.0053 | 0.0052 | 0 | 0.949 | 0.869 |
| Means | CHR | 0.821 | 0.8213 | 0.0185 | 0.0184 | 0.0003 | 0.947 | 1 |
| Means | AGE | 19.056 | 19.0572 | 0.1915 | 0.1907 | 0.0367 | 0.95 | 1 |
| Means | FEMALE | 0.453 | 0.4529 | 0.0238 | 0.0238 | 0.0006 | 0.949 | 1 |
| Means | SITE02 | 0.087 | 0.0872 | 0.0135 | 0.0135 | 0.0002 | 0.95 | 1 |
| Means | SITE03 | 0.06 | 0.0602 | 0.0113 | 0.0113 | 0.0001 | 0.947 | 1 |
| Means | SITE04 | 0.087 | 0.0868 | 0.0135 | 0.0135 | 0.0002 | 0.951 | 1 |
| Means | SITE05 | 0.14 | 0.1398 | 0.0166 | 0.0166 | 0.0003 | 0.949 | 1 |
| Means | SITE06 | 0.129 | 0.1289 | 0.0164 | 0.016 | 0.0003 | 0.946 | 1 |
| Means | SITE07 | 0.138 | 0.1379 | 0.0165 | 0.0165 | 0.0003 | 0.948 | 1 |
| Means | SITE08 | 0.106 | 0.106 | 0.0149 | 0.0147 | 0.0002 | 0.949 | 1 |
| Means | SITE09 | 0.147 | 0.1472 | 0.017 | 0.0169 | 0.0003 | 0.95 | 1 |
| Intercepts | POSS_CH | 0.318 | 0.3115 | 0.3017 | 0.2963 | 0.0911 | 0.943 | 0.188 |
| Intercepts | HI_CH | 0 | -0.0002 | 0.0476 | 0.0478 | 0.0023 | 0.947 | 0.053 |
| Variances | CHR | 0.147 | 0.1468 | 0.01 | 0.0099 | 0.0001 | 0.942 | 1 |
| Variances | AGE | 15.868 | 15.8308 | 1.0737 | 1.0673 | 1.1542 | 0.943 | 1 |
| Variances | FEMALE | 0.248 | 0.2477 | 0.0169 | 0.0167 | 0.0003 | 0.942 | 1 |
| Variances | SITE02 | 0.08 | 0.0798 | 0.0054 | 0.0054 | 0 | 0.942 | 1 |
| Variances | SITE03 | 0.056 | 0.0559 | 0.0038 | 0.0038 | 0 | 0.94 | 1 |
| Variances | SITE04 | 0.08 | 0.0799 | 0.0054 | 0.0054 | 0 | 0.946 | 1 |
| Variances | SITE05 | 0.121 | 0.1207 | 0.0082 | 0.0081 | 0.0001 | 0.942 | 1 |
| Variances | SITE06 | 0.112 | 0.1117 | 0.0075 | 0.0075 | 0.0001 | 0.947 | 1 |
| Variances | SITE07 | 0.119 | 0.1188 | 0.0081 | 0.008 | 0.0001 | 0.94 | 1 |
| Variances | SITE08 | 0.095 | 0.0947 | 0.0063 | 0.0064 | 0 | 0.944 | 1 |
| Variances | SITE09 | 0.125 | 0.1246 | 0.0085 | 0.0084 | 0.0001 | 0.945 | 1 |
| Variances | LV_HI | 0.657 | 0.6526 | 0.0671 | 0.0668 | 0.0045 | 0.942 | 1 |
| Residual.Variances | POSS_CH | 0.142 | 0.1422 | 0 | 0 | 0 | 1 | 0 |
| Residual.Variances | HI_CH | 0.339 | 0.339 | 0 | 0 | 0 | 1 | 0 |
| Residual.Variances | LV_POSS | 0.746 | 0.7136 | 0.0602 | 0.0589 | 0.0047 | 0.888 | 1 |
| LV_HI.BY | HI_CH | 1 | 1 | 0 | 0 | 0 | 1 | 0 |
| LV_SYCOD.BY | SYCOD_CH | 1 | 1 | 0 | 0 | 0 | 1 | 0 |
| LV_SYCOD.ON | LV_HI | 0.172 | 0.1722 | 0.1739 | 0.1702 | 0.0302 | 0.939 | 0.19 |
| LV_SYCOD.ON | HIXCHR | -0.082 | -0.0799 | 0.1917 | 0.1873 | 0.0367 | 0.94 | 0.084 |
| LV_SYCOD.ON | CHR | -0.129 | -0.1287 | 0.1233 | 0.1217 | 0.0152 | 0.944 | 0.187 |
| LV_SYCOD.ON | AGE | -0.006 | -0.0057 | 0.0132 | 0.0128 | 0.0002 | 0.943 | 0.084 |
| LV_SYCOD.ON | FEMALE | -0.14 | -0.139 | 0.0972 | 0.0949 | 0.0095 | 0.943 | 0.312 |
| LV_SYCOD.ON | SITE02 | 0.005 | 0.0055 | 0.2256 | 0.2194 | 0.0509 | 0.946 | 0.054 |
| LV_SYCOD.ON | SITE03 | 0.23 | 0.2312 | 0.2386 | 0.2331 | 0.0569 | 0.943 | 0.173 |
| LV_SYCOD.ON | SITE04 | -0.463 | -0.4606 | 0.2173 | 0.2109 | 0.0472 | 0.942 | 0.584 |
| LV_SYCOD.ON | SITE05 | 0.287 | 0.2864 | 0.1941 | 0.1895 | 0.0377 | 0.945 | 0.329 |
| LV_SYCOD.ON | SITE06 | -0.189 | -0.1875 | 0.1958 | 0.1911 | 0.0384 | 0.942 | 0.177 |
| LV_SYCOD.ON | SITE07 | -0.18 | -0.1765 | 0.1905 | 0.1875 | 0.0363 | 0.946 | 0.162 |
| LV_SYCOD.ON | SITE08 | -0.49 | -0.4916 | 0.2026 | 0.1993 | 0.0411 | 0.947 | 0.691 |
| LV_SYCOD.ON | SITE09 | -0.004 | -0.0039 | 0.1896 | 0.1854 | 0.0359 | 0.943 | 0.057 |
| LV_HI.WITH | CHR | -0.002 | -0.0019 | 0.0185 | 0.0184 | 0.0003 | 0.947 | 0.055 |
| LV_HI.WITH | AGE | 0.084 | 0.0828 | 0.1883 | 0.1905 | 0.0355 | 0.953 | 0.069 |
| LV_HI.WITH | FEMALE | -0.034 | -0.0339 | 0.0241 | 0.0239 | 0.0006 | 0.948 | 0.294 |
| LV_HI.WITH | SITE02 | 0.006 | 0.0061 | 0.0135 | 0.0134 | 0.0002 | 0.95 | 0.078 |
| LV_HI.WITH | SITE03 | -0.001 | -0.0009 | 0.0113 | 0.0114 | 0.0001 | 0.953 | 0.046 |
| LV_HI.WITH | SITE04 | 0.014 | 0.0141 | 0.0135 | 0.0135 | 0.0002 | 0.944 | 0.179 |
| LV_HI.WITH | SITE05 | -0.017 | -0.0167 | 0.0165 | 0.0166 | 0.0003 | 0.948 | 0.17 |
| LV_HI.WITH | SITE06 | 0.021 | 0.0208 | 0.0161 | 0.0161 | 0.0003 | 0.95 | 0.248 |
| LV_HI.WITH | SITE07 | -0.002 | -0.0022 | 0.0168 | 0.0166 | 0.0003 | 0.948 | 0.053 |
| LV_HI.WITH | SITE08 | -0.003 | -0.0029 | 0.015 | 0.0148 | 0.0002 | 0.948 | 0.056 |
| LV_HI.WITH | SITE09 | -0.011 | -0.011 | 0.0169 | 0.0171 | 0.0003 | 0.951 | 0.095 |
| CHR.WITH | AGE | -0.075 | -0.0751 | 0.0729 | 0.0733 | 0.0053 | 0.952 | 0.174 |
| CHR.WITH | FEMALE | -0.006 | -0.0059 | 0.0092 | 0.0092 | 0.0001 | 0.948 | 0.096 |
| CHR.WITH | SITE02 | -0.003 | -0.003 | 0.0052 | 0.0052 | 0 | 0.946 | 0.091 |
| CHR.WITH | SITE03 | -0.003 | -0.0029 | 0.0044 | 0.0044 | 0 | 0.952 | 0.104 |
| CHR.WITH | SITE04 | 0.001 | 0.001 | 0.0052 | 0.0052 | 0 | 0.95 | 0.057 |
| CHR.WITH | SITE05 | -0.001 | -0.001 | 0.0064 | 0.0064 | 0 | 0.95 | 0.054 |
| CHR.WITH | SITE06 | -0.002 | -0.002 | 0.0062 | 0.0062 | 0 | 0.952 | 0.063 |
| CHR.WITH | SITE07 | -0.001 | -0.001 | 0.0064 | 0.0064 | 0 | 0.945 | 0.058 |
| CHR.WITH | SITE08 | 0.003 | 0.003 | 0.0058 | 0.0057 | 0 | 0.946 | 0.085 |
| CHR.WITH | SITE09 | 0.001 | 0.001 | 0.0066 | 0.0066 | 0 | 0.947 | 0.056 |
| AGE.WITH | FEMALE | -0.219 | -0.217 | 0.0964 | 0.0957 | 0.0093 | 0.948 | 0.621 |
| AGE.WITH | SITE02 | 0.346 | 0.3447 | 0.0566 | 0.0561 | 0.0032 | 0.945 | 1 |
| AGE.WITH | SITE03 | 0 | -0.0005 | 0.046 | 0.0456 | 0.0021 | 0.949 | 0.051 |
| AGE.WITH | SITE04 | -0.011 | -0.0103 | 0.0552 | 0.0537 | 0.003 | 0.942 | 0.059 |
| AGE.WITH | SITE05 | 0.209 | 0.2079 | 0.0676 | 0.0669 | 0.0046 | 0.947 | 0.879 |
| AGE.WITH | SITE06 | -0.012 | -0.0128 | 0.0649 | 0.0642 | 0.0042 | 0.947 | 0.057 |
| AGE.WITH | SITE07 | -0.198 | -0.1975 | 0.0677 | 0.0668 | 0.0046 | 0.947 | 0.845 |
| AGE.WITH | SITE08 | -0.211 | -0.2098 | 0.0603 | 0.06 | 0.0036 | 0.95 | 0.945 |
| AGE.WITH | SITE09 | -0.042 | -0.0405 | 0.0691 | 0.068 | 0.0048 | 0.946 | 0.093 |
| FEMALE.WITH | SITE02 | 0.005 | 0.005 | 0.0067 | 0.0067 | 0 | 0.951 | 0.117 |
| FEMALE.WITH | SITE03 | -0.002 | -0.002 | 0.0058 | 0.0057 | 0 | 0.947 | 0.068 |
| FEMALE.WITH | SITE04 | 0 | 0 | 0.0067 | 0.0067 | 0 | 0.949 | 0.051 |
| FEMALE.WITH | SITE05 | 0.001 | 0.001 | 0.0083 | 0.0083 | 0.0001 | 0.947 | 0.054 |
| FEMALE.WITH | SITE06 | -0.011 | -0.011 | 0.0081 | 0.0081 | 0.0001 | 0.95 | 0.275 |
| FEMALE.WITH | SITE07 | 0.01 | 0.01 | 0.0084 | 0.0083 | 0.0001 | 0.945 | 0.229 |
| FEMALE.WITH | SITE08 | 0 | -0.0001 | 0.0074 | 0.0074 | 0.0001 | 0.948 | 0.052 |
| FEMALE.WITH | SITE09 | 0.002 | 0.002 | 0.0086 | 0.0085 | 0.0001 | 0.947 | 0.057 |
| SITE02.WITH | SITE03 | -0.005 | -0.0051 | 0.0033 | 0.0032 | 0 | 0.947 | 0.352 |
| SITE02.WITH | SITE04 | -0.007 | -0.007 | 0.0038 | 0.0038 | 0 | 0.949 | 0.45 |
| SITE02.WITH | SITE05 | -0.012 | -0.012 | 0.0047 | 0.0047 | 0 | 0.95 | 0.727 |
| SITE02.WITH | SITE06 | -0.011 | -0.011 | 0.0046 | 0.0046 | 0 | 0.95 | 0.677 |
| SITE02.WITH | SITE07 | -0.012 | -0.012 | 0.0048 | 0.0047 | 0 | 0.949 | 0.72 |
| SITE02.WITH | SITE08 | -0.009 | -0.0089 | 0.0042 | 0.0042 | 0 | 0.948 | 0.567 |
| SITE02.WITH | SITE09 | -0.013 | -0.0129 | 0.0049 | 0.0048 | 0 | 0.947 | 0.765 |
| SITE03.WITH | SITE04 | -0.005 | -0.005 | 0.0032 | 0.0032 | 0 | 0.95 | 0.34 |
| SITE03.WITH | SITE05 | -0.008 | -0.008 | 0.004 | 0.004 | 0 | 0.949 | 0.524 |
| SITE03.WITH | SITE06 | -0.008 | -0.008 | 0.0039 | 0.0039 | 0 | 0.95 | 0.541 |
| SITE03.WITH | SITE07 | -0.008 | -0.0079 | 0.004 | 0.004 | 0 | 0.947 | 0.511 |
| SITE03.WITH | SITE08 | -0.006 | -0.006 | 0.0036 | 0.0036 | 0 | 0.949 | 0.389 |
| SITE03.WITH | SITE09 | -0.009 | -0.009 | 0.0042 | 0.0041 | 0 | 0.946 | 0.6 |
| SITE04.WITH | SITE05 | -0.012 | -0.012 | 0.0047 | 0.0047 | 0 | 0.947 | 0.728 |
| SITE04.WITH | SITE06 | -0.011 | -0.011 | 0.0046 | 0.0046 | 0 | 0.949 | 0.679 |
| SITE04.WITH | SITE07 | -0.012 | -0.0121 | 0.0048 | 0.0047 | 0 | 0.948 | 0.726 |
| SITE04.WITH | SITE08 | -0.009 | -0.009 | 0.0042 | 0.0042 | 0 | 0.948 | 0.568 |
| SITE04.WITH | SITE09 | -0.013 | -0.0129 | 0.0048 | 0.0048 | 0 | 0.947 | 0.764 |
| SITE05.WITH | SITE06 | -0.018 | -0.0179 | 0.0057 | 0.0056 | 0 | 0.946 | 0.89 |
| SITE05.WITH | SITE07 | -0.019 | -0.019 | 0.0059 | 0.0058 | 0 | 0.948 | 0.91 |
| SITE05.WITH | SITE08 | -0.015 | -0.015 | 0.0052 | 0.0052 | 0 | 0.949 | 0.829 |
| SITE05.WITH | SITE09 | -0.021 | -0.0209 | 0.006 | 0.006 | 0 | 0.949 | 0.942 |
| SITE06.WITH | SITE07 | -0.018 | -0.0179 | 0.0057 | 0.0056 | 0 | 0.945 | 0.895 |
| SITE06.WITH | SITE08 | -0.014 | -0.0139 | 0.0051 | 0.005 | 0 | 0.945 | 0.79 |
| SITE06.WITH | SITE09 | -0.019 | -0.0191 | 0.0058 | 0.0058 | 0 | 0.951 | 0.917 |
| SITE07.WITH | SITE08 | -0.015 | -0.015 | 0.0053 | 0.0052 | 0 | 0.945 | 0.826 |
| SITE07.WITH | SITE09 | -0.021 | -0.0208 | 0.0061 | 0.006 | 0 | 0.946 | 0.939 |
| SITE08.WITH | SITE09 | -0.016 | -0.016 | 0.0054 | 0.0053 | 0 | 0.948 | 0.852 |
| Means | CHR | 0.821 | 0.8212 | 0.0186 | 0.0185 | 0.0003 | 0.948 | 1 |
| Means | AGE | 19.053 | 19.0539 | 0.1925 | 0.1916 | 0.0371 | 0.947 | 1 |
| Means | FEMALE | 0.458 | 0.4579 | 0.0238 | 0.024 | 0.0006 | 0.951 | 1 |
| Means | SITE02 | 0.086 | 0.0862 | 0.0135 | 0.0135 | 0.0002 | 0.951 | 1 |
| Means | SITE03 | 0.06 | 0.0601 | 0.0115 | 0.0115 | 0.0001 | 0.951 | 0.999 |
| Means | SITE04 | 0.086 | 0.0858 | 0.0135 | 0.0135 | 0.0002 | 0.95 | 1 |
| Means | SITE05 | 0.14 | 0.1398 | 0.0166 | 0.0167 | 0.0003 | 0.95 | 1 |
| Means | SITE06 | 0.13 | 0.1299 | 0.0165 | 0.0162 | 0.0003 | 0.943 | 1 |
| Means | SITE07 | 0.14 | 0.1399 | 0.0167 | 0.0167 | 0.0003 | 0.949 | 1 |
| Means | SITE08 | 0.107 | 0.1071 | 0.0151 | 0.0149 | 0.0002 | 0.947 | 1 |
| Means | SITE09 | 0.149 | 0.1492 | 0.0173 | 0.0171 | 0.0003 | 0.947 | 1 |
| Intercepts | SYCOD_CH | 0.374 | 0.366 | 0.3106 | 0.3039 | 0.0966 | 0.944 | 0.228 |
| Intercepts | HI_CH | 0 | -0.0003 | 0.048 | 0.0481 | 0.0023 | 0.95 | 0.05 |
| Variances | CHR | 0.147 | 0.1468 | 0.01 | 0.01 | 0.0001 | 0.946 | 1 |
| Variances | AGE | 15.846 | 15.8095 | 1.0717 | 1.0721 | 1.1498 | 0.947 | 1 |
| Variances | FEMALE | 0.248 | 0.2477 | 0.017 | 0.0168 | 0.0003 | 0.942 | 1 |
| Variances | SITE02 | 0.079 | 0.0788 | 0.0054 | 0.0054 | 0 | 0.944 | 1 |
| Variances | SITE03 | 0.057 | 0.0569 | 0.0039 | 0.0039 | 0 | 0.941 | 1 |
| Variances | SITE04 | 0.079 | 0.0789 | 0.0053 | 0.0054 | 0 | 0.942 | 1 |
| Variances | SITE05 | 0.12 | 0.1197 | 0.0082 | 0.0081 | 0.0001 | 0.946 | 1 |
| Variances | SITE06 | 0.113 | 0.1127 | 0.0076 | 0.0077 | 0.0001 | 0.942 | 1 |
| Variances | SITE07 | 0.12 | 0.1198 | 0.0081 | 0.0081 | 0.0001 | 0.944 | 1 |
| Variances | SITE08 | 0.096 | 0.0957 | 0.0065 | 0.0065 | 0 | 0.943 | 1 |
| Variances | SITE09 | 0.127 | 0.1266 | 0.0086 | 0.0086 | 0.0001 | 0.942 | 1 |
| Variances | LV_HI | 0.659 | 0.6547 | 0.0675 | 0.0672 | 0.0046 | 0.943 | 1 |
| Residual.Variances | SYCOD_CH | 0.203 | 0.2033 | 0 | 0 | 0 | 1 | 0 |
| Residual.Variances | HI_CH | 0.338 | 0.3379 | 0 | 0 | 0 | 1 | 0 |
| Residual.Variances | LV_SYCOD | 0.724 | 0.6901 | 0.0617 | 0.0611 | 0.005 | 0.89 | 1 |
| LV_HI.BY | HI_CH | 1 | 1 | 0 | 0 | 0 | 1 | 0 |
| LV_VEMEM.BY | VEMEM_CH | 1 | 1 | 0 | 0 | 0 | 1 | 0 |
| LV_VEMEM.ON | LV_HI | 0.089 | 0.0851 | 0.1737 | 0.1699 | 0.0302 | 0.941 | 0.091 |
| LV_VEMEM.ON | HIXCHR | -0.24 | -0.2381 | 0.1922 | 0.1878 | 0.0369 | 0.938 | 0.26 |
| LV_VEMEM.ON | CHR | -0.096 | -0.0955 | 0.1244 | 0.1226 | 0.0155 | 0.945 | 0.123 |
| LV_VEMEM.ON | AGE | 0.007 | 0.0073 | 0.0132 | 0.0129 | 0.0002 | 0.944 | 0.095 |
| LV_VEMEM.ON | FEMALE | -0.152 | -0.1514 | 0.098 | 0.0954 | 0.0096 | 0.941 | 0.358 |
| LV_VEMEM.ON | SITE02 | 0.102 | 0.103 | 0.2258 | 0.2193 | 0.051 | 0.944 | 0.08 |
| LV_VEMEM.ON | SITE03 | 0.011 | 0.0128 | 0.2409 | 0.2348 | 0.058 | 0.945 | 0.055 |
| LV_VEMEM.ON | SITE04 | -0.45 | -0.4465 | 0.2208 | 0.2136 | 0.0488 | 0.942 | 0.55 |
| LV_VEMEM.ON | SITE05 | 0.077 | 0.0768 | 0.1947 | 0.1901 | 0.0379 | 0.945 | 0.078 |
| LV_VEMEM.ON | SITE06 | 0.268 | 0.2703 | 0.1979 | 0.1933 | 0.0392 | 0.944 | 0.296 |
| LV_VEMEM.ON | SITE07 | 0.16 | 0.1627 | 0.1907 | 0.1881 | 0.0364 | 0.946 | 0.146 |
| LV_VEMEM.ON | SITE08 | 0.139 | 0.1372 | 0.2044 | 0.2007 | 0.0418 | 0.945 | 0.114 |
| LV_VEMEM.ON | SITE09 | 0.288 | 0.2883 | 0.1912 | 0.1873 | 0.0366 | 0.945 | 0.344 |
| LV_HI.WITH | CHR | -0.001 | -0.0009 | 0.0186 | 0.0184 | 0.0003 | 0.946 | 0.055 |
| LV_HI.WITH | AGE | 0.083 | 0.0817 | 0.1883 | 0.1904 | 0.0354 | 0.952 | 0.068 |
| LV_HI.WITH | FEMALE | -0.031 | -0.0309 | 0.0241 | 0.0239 | 0.0006 | 0.948 | 0.253 |
| LV_HI.WITH | SITE02 | 0.005 | 0.0051 | 0.0137 | 0.0136 | 0.0002 | 0.949 | 0.069 |
| LV_HI.WITH | SITE03 | -0.001 | -0.0009 | 0.0113 | 0.0114 | 0.0001 | 0.953 | 0.045 |
| LV_HI.WITH | SITE04 | 0.015 | 0.0151 | 0.0135 | 0.0135 | 0.0002 | 0.945 | 0.197 |
| LV_HI.WITH | SITE05 | -0.017 | -0.0167 | 0.0165 | 0.0166 | 0.0003 | 0.947 | 0.167 |
| LV_HI.WITH | SITE06 | 0.021 | 0.0207 | 0.0161 | 0.0161 | 0.0003 | 0.949 | 0.247 |
| LV_HI.WITH | SITE07 | -0.002 | -0.0023 | 0.0168 | 0.0166 | 0.0003 | 0.948 | 0.054 |
| LV_HI.WITH | SITE08 | -0.003 | -0.0029 | 0.0149 | 0.0148 | 0.0002 | 0.949 | 0.057 |
| LV_HI.WITH | SITE09 | -0.012 | -0.012 | 0.0168 | 0.0169 | 0.0003 | 0.951 | 0.105 |
| CHR.WITH | AGE | -0.075 | -0.0751 | 0.0731 | 0.0735 | 0.0053 | 0.952 | 0.172 |
| CHR.WITH | FEMALE | -0.005 | -0.0049 | 0.0092 | 0.0092 | 0.0001 | 0.949 | 0.085 |
| CHR.WITH | SITE02 | -0.005 | -0.005 | 0.0053 | 0.0053 | 0 | 0.946 | 0.156 |
| CHR.WITH | SITE03 | -0.003 | -0.0029 | 0.0044 | 0.0044 | 0 | 0.951 | 0.105 |
| CHR.WITH | SITE04 | 0.002 | 0.002 | 0.0052 | 0.0052 | 0 | 0.951 | 0.073 |
| CHR.WITH | SITE05 | 0 | 0 | 0.0064 | 0.0064 | 0 | 0.948 | 0.052 |
| CHR.WITH | SITE06 | -0.002 | -0.002 | 0.0062 | 0.0062 | 0 | 0.95 | 0.063 |
| CHR.WITH | SITE07 | 0 | 0 | 0.0064 | 0.0064 | 0 | 0.946 | 0.054 |
| CHR.WITH | SITE08 | 0.003 | 0.003 | 0.0058 | 0.0057 | 0 | 0.946 | 0.083 |
| CHR.WITH | SITE09 | 0.001 | 0.001 | 0.0066 | 0.0065 | 0 | 0.947 | 0.056 |
| AGE.WITH | FEMALE | -0.221 | -0.219 | 0.0963 | 0.0957 | 0.0093 | 0.949 | 0.628 |
| AGE.WITH | SITE02 | 0.346 | 0.3447 | 0.0572 | 0.0568 | 0.0033 | 0.944 | 1 |
| AGE.WITH | SITE03 | 0 | -0.0006 | 0.046 | 0.0456 | 0.0021 | 0.949 | 0.051 |
| AGE.WITH | SITE04 | -0.012 | -0.0113 | 0.0552 | 0.0537 | 0.003 | 0.941 | 0.061 |
| AGE.WITH | SITE05 | 0.208 | 0.2067 | 0.0675 | 0.0668 | 0.0046 | 0.949 | 0.878 |
| AGE.WITH | SITE06 | -0.012 | -0.0127 | 0.0649 | 0.0641 | 0.0042 | 0.947 | 0.056 |
| AGE.WITH | SITE07 | -0.199 | -0.1985 | 0.0677 | 0.0668 | 0.0046 | 0.947 | 0.849 |
| AGE.WITH | SITE08 | -0.211 | -0.2096 | 0.0603 | 0.06 | 0.0036 | 0.951 | 0.944 |
| AGE.WITH | SITE09 | -0.038 | -0.0365 | 0.0685 | 0.0674 | 0.0047 | 0.946 | 0.085 |
| FEMALE.WITH | SITE02 | 0.004 | 0.004 | 0.0068 | 0.0068 | 0 | 0.951 | 0.091 |
| FEMALE.WITH | SITE03 | -0.002 | -0.002 | 0.0058 | 0.0057 | 0 | 0.946 | 0.068 |
| FEMALE.WITH | SITE04 | 0 | 0 | 0.0067 | 0.0067 | 0 | 0.949 | 0.051 |
| FEMALE.WITH | SITE05 | 0.001 | 0.001 | 0.0083 | 0.0083 | 0.0001 | 0.947 | 0.055 |
| FEMALE.WITH | SITE06 | -0.011 | -0.011 | 0.0081 | 0.0081 | 0.0001 | 0.95 | 0.276 |
| FEMALE.WITH | SITE07 | 0.01 | 0.01 | 0.0084 | 0.0083 | 0.0001 | 0.946 | 0.229 |
| FEMALE.WITH | SITE08 | 0 | -0.0001 | 0.0074 | 0.0074 | 0.0001 | 0.949 | 0.051 |
| FEMALE.WITH | SITE09 | 0.003 | 0.003 | 0.0085 | 0.0084 | 0.0001 | 0.947 | 0.065 |
| SITE02.WITH | SITE03 | -0.005 | -0.0051 | 0.0033 | 0.0033 | 0 | 0.945 | 0.345 |
| SITE02.WITH | SITE04 | -0.008 | -0.008 | 0.0039 | 0.0039 | 0 | 0.949 | 0.547 |
| SITE02.WITH | SITE05 | -0.012 | -0.012 | 0.0048 | 0.0048 | 0 | 0.949 | 0.72 |
| SITE02.WITH | SITE06 | -0.012 | -0.012 | 0.0046 | 0.0046 | 0 | 0.949 | 0.739 |
| SITE02.WITH | SITE07 | -0.012 | -0.012 | 0.0048 | 0.0048 | 0 | 0.948 | 0.711 |
| SITE02.WITH | SITE08 | -0.009 | -0.0089 | 0.0043 | 0.0042 | 0 | 0.949 | 0.558 |
| SITE02.WITH | SITE09 | -0.013 | -0.0129 | 0.0049 | 0.0049 | 0 | 0.946 | 0.761 |
| SITE03.WITH | SITE04 | -0.005 | -0.005 | 0.0032 | 0.0032 | 0 | 0.951 | 0.34 |
| SITE03.WITH | SITE05 | -0.008 | -0.008 | 0.004 | 0.004 | 0 | 0.951 | 0.522 |
| SITE03.WITH | SITE06 | -0.008 | -0.008 | 0.0039 | 0.0039 | 0 | 0.949 | 0.538 |
| SITE03.WITH | SITE07 | -0.008 | -0.0079 | 0.004 | 0.004 | 0 | 0.946 | 0.511 |
| SITE03.WITH | SITE08 | -0.006 | -0.006 | 0.0036 | 0.0036 | 0 | 0.951 | 0.389 |
| SITE03.WITH | SITE09 | -0.009 | -0.009 | 0.0042 | 0.0041 | 0 | 0.946 | 0.605 |
| SITE04.WITH | SITE05 | -0.012 | -0.012 | 0.0047 | 0.0047 | 0 | 0.947 | 0.727 |
| SITE04.WITH | SITE06 | -0.011 | -0.011 | 0.0046 | 0.0046 | 0 | 0.95 | 0.68 |
| SITE04.WITH | SITE07 | -0.012 | -0.0121 | 0.0047 | 0.0047 | 0 | 0.948 | 0.726 |
| SITE04.WITH | SITE08 | -0.009 | -0.009 | 0.0043 | 0.0042 | 0 | 0.946 | 0.572 |
| SITE04.WITH | SITE09 | -0.013 | -0.0129 | 0.0048 | 0.0048 | 0 | 0.948 | 0.772 |
| SITE05.WITH | SITE06 | -0.018 | -0.0179 | 0.0057 | 0.0056 | 0 | 0.945 | 0.89 |
| SITE05.WITH | SITE07 | -0.019 | -0.0191 | 0.0059 | 0.0058 | 0 | 0.947 | 0.91 |
| SITE05.WITH | SITE08 | -0.015 | -0.015 | 0.0052 | 0.0052 | 0 | 0.947 | 0.827 |
| SITE05.WITH | SITE09 | -0.02 | -0.0199 | 0.006 | 0.0059 | 0 | 0.947 | 0.923 |
| SITE06.WITH | SITE07 | -0.018 | -0.0179 | 0.0057 | 0.0056 | 0 | 0.944 | 0.892 |
| SITE06.WITH | SITE08 | -0.014 | -0.0139 | 0.0051 | 0.005 | 0 | 0.946 | 0.789 |
| SITE06.WITH | SITE09 | -0.019 | -0.0191 | 0.0058 | 0.0058 | 0 | 0.951 | 0.92 |
| SITE07.WITH | SITE08 | -0.015 | -0.015 | 0.0053 | 0.0052 | 0 | 0.945 | 0.825 |
| SITE07.WITH | SITE09 | -0.02 | -0.0199 | 0.006 | 0.0059 | 0 | 0.946 | 0.92 |
| SITE08.WITH | SITE09 | -0.016 | -0.016 | 0.0054 | 0.0053 | 0 | 0.947 | 0.859 |
| Means | CHR | 0.819 | 0.8192 | 0.0186 | 0.0185 | 0.0003 | 0.948 | 1 |
| Means | AGE | 19.058 | 19.0589 | 0.1925 | 0.1916 | 0.037 | 0.947 | 1 |
| Means | FEMALE | 0.458 | 0.4579 | 0.0238 | 0.024 | 0.0006 | 0.95 | 1 |
| Means | SITE02 | 0.088 | 0.0882 | 0.0137 | 0.0137 | 0.0002 | 0.95 | 1 |
| Means | SITE03 | 0.06 | 0.0602 | 0.0115 | 0.0115 | 0.0001 | 0.951 | 0.999 |
| Means | SITE04 | 0.086 | 0.0858 | 0.0135 | 0.0135 | 0.0002 | 0.95 | 1 |
| Means | SITE05 | 0.14 | 0.1398 | 0.0166 | 0.0167 | 0.0003 | 0.95 | 1 |
| Means | SITE06 | 0.13 | 0.1299 | 0.0165 | 0.0162 | 0.0003 | 0.945 | 1 |
| Means | SITE07 | 0.14 | 0.1399 | 0.0167 | 0.0167 | 0.0003 | 0.95 | 1 |
| Means | SITE08 | 0.107 | 0.107 | 0.0151 | 0.0149 | 0.0002 | 0.947 | 1 |
| Means | SITE09 | 0.147 | 0.1472 | 0.0171 | 0.017 | 0.0003 | 0.948 | 1 |
| Intercepts | VEMEM_CH | -0.075 | -0.0823 | 0.3114 | 0.3043 | 0.097 | 0.942 | 0.064 |
| Intercepts | HI_CH | 0 | -0.0002 | 0.048 | 0.0481 | 0.0023 | 0.951 | 0.049 |
| Variances | CHR | 0.148 | 0.1478 | 0.0101 | 0.01 | 0.0001 | 0.946 | 1 |
| Variances | AGE | 15.837 | 15.8003 | 1.0712 | 1.0715 | 1.1486 | 0.947 | 1 |
| Variances | FEMALE | 0.248 | 0.2477 | 0.017 | 0.0168 | 0.0003 | 0.942 | 1 |
| Variances | SITE02 | 0.081 | 0.0808 | 0.0055 | 0.0055 | 0 | 0.944 | 1 |
| Variances | SITE03 | 0.057 | 0.0569 | 0.0039 | 0.0039 | 0 | 0.941 | 1 |
| Variances | SITE04 | 0.079 | 0.0789 | 0.0053 | 0.0054 | 0 | 0.946 | 1 |
| Variances | SITE05 | 0.12 | 0.1197 | 0.0082 | 0.0081 | 0.0001 | 0.945 | 1 |
| Variances | SITE06 | 0.113 | 0.1127 | 0.0076 | 0.0077 | 0.0001 | 0.942 | 1 |
| Variances | SITE07 | 0.12 | 0.1198 | 0.0081 | 0.0081 | 0.0001 | 0.943 | 1 |
| Variances | SITE08 | 0.096 | 0.0957 | 0.0065 | 0.0065 | 0 | 0.943 | 1 |
| Variances | SITE09 | 0.125 | 0.1246 | 0.0085 | 0.0085 | 0.0001 | 0.943 | 1 |
| Variances | LV_HI | 0.66 | 0.6561 | 0.0675 | 0.0672 | 0.0046 | 0.944 | 1 |
| Residual.Variances | VEMEM_CH | 0.452 | 0.452 | 0 | 0 | 0 | 1 | 0 |
| Residual.Variances | HI_CH | 0.337 | 0.3366 | 0 | 0 | 0 | 1 | 0 |
| Residual.Variances | LV_VEMEM | 0.487 | 0.4529 | 0.0632 | 0.0621 | 0.0052 | 0.889 | 1 |

Table_S5a_SensitivityAnalysisStandardized_Medication

| **paramHeader** | **param** | **est** | **se** | **est_se** | **pval** | **zval** | **pval_exact** |
| --- | --- | --- | --- | --- | --- | --- | --- |
| LV_HI.BY | HI_CH | 0.821 | 0.025 | 32.421 | 0 | 32.84 | 0 |
| LV_DEPS.BY | DEPS_CH | 0.844 | 0.043 | 19.481 | 0 | 19.62791 | 0 |
| LV_CPZ.BY | CPZ_CH | 0.968 | 0.007 | 137.596 | 0 | 138.2857 | 0 |
| LV_DEPS.ON | LV_HI | -0.341 | 0.105 | -3.243 | 0.001 | -3.24762 | 0.001164 |
| LV_DEPS.ON | LV_CPZ | 0.031 | 0.055 | 0.56 | 0.576 | 0.563636 | 0.573002 |
| LV_DEPS.ON | AGE | -0.087 | 0.057 | -1.527 | 0.127 | -1.52632 | 0.126931 |
| LV_DEPS.ON | FEMALE | -0.035 | 0.059 | -0.595 | 0.552 | -0.59322 | 0.553034 |
| LV_DEPS.ON | SITE02 | -0.069 | 0.107 | -0.646 | 0.518 | -0.64486 | 0.519018 |
| LV_DEPS.ON | SITE03 | 0.129 | 0.071 | 1.83 | 0.067 | 1.816901 | 0.069232 |
| LV_DEPS.ON | SITE04 | 0.036 | 0.073 | 0.493 | 0.622 | 0.493151 | 0.621906 |
| LV_DEPS.ON | SITE05 | 0.116 | 0.08 | 1.453 | 0.146 | 1.45 | 0.147059 |
| LV_DEPS.ON | SITE06 | 0.046 | 0.082 | 0.559 | 0.576 | 0.560976 | 0.574814 |
| LV_DEPS.ON | SITE07 | 0.075 | 0.077 | 0.97 | 0.332 | 0.974026 | 0.330044 |
| LV_DEPS.ON | SITE08 | 0.025 | 0.084 | 0.295 | 0.768 | 0.297619 | 0.765994 |
| LV_DEPS.ON | SITE09 | 0.181 | 0.089 | 2.021 | 0.043 | 2.033708 | 0.041981 |
| LV_HI.WITH | AGE | 0.035 | 0.061 | 0.566 | 0.571 | 0.57377 | 0.566123 |
| LV_HI.WITH | FEMALE | -0.107 | 0.066 | -1.632 | 0.103 | -1.62121 | 0.104972 |
| LV_HI.WITH | SITE02 | -0.009 | 0.067 | -0.137 | 0.891 | -0.13433 | 0.893143 |
| LV_HI.WITH | SITE03 | 0.034 | 0.099 | 0.343 | 0.731 | 0.343434 | 0.731272 |
| LV_HI.WITH | SITE04 | 0.053 | 0.05 | 1.066 | 0.286 | 1.06 | 0.289145 |
| LV_HI.WITH | SITE05 | -0.088 | 0.069 | -1.287 | 0.198 | -1.27536 | 0.202181 |
| LV_HI.WITH | SITE06 | 0.092 | 0.048 | 1.912 | 0.056 | 1.916667 | 0.05528 |
| LV_HI.WITH | SITE07 | 0.018 | 0.071 | 0.253 | 0.8 | 0.253521 | 0.799866 |
| LV_HI.WITH | SITE08 | -0.042 | 0.054 | -0.773 | 0.439 | -0.77778 | 0.4367 |
| LV_HI.WITH | SITE09 | -0.038 | 0.065 | -0.591 | 0.555 | -0.58462 | 0.558806 |
| LV_HI.WITH | LV_CPZ | 0.037 | 0.1 | 0.372 | 0.71 | 0.37 | 0.711382 |
| AGE.WITH | LV_CPZ | -0.069 | 0.059 | -1.171 | 0.241 | -1.16949 | 0.242206 |
| FEMALE.WITH | LV_CPZ | -0.006 | 0.053 | -0.119 | 0.905 | -0.11321 | 0.909866 |
| SITE02.WITH | LV_CPZ | -0.062 | 0.044 | -1.413 | 0.158 | -1.40909 | 0.158808 |
| SITE03.WITH | LV_CPZ | 0.016 | 0.05 | 0.32 | 0.749 | 0.32 | 0.748968 |
| SITE04.WITH | LV_CPZ | 0.05 | 0.048 | 1.041 | 0.298 | 1.041667 | 0.297566 |
| SITE05.WITH | LV_CPZ | 0.023 | 0.04 | 0.581 | 0.561 | 0.575 | 0.565291 |
| SITE06.WITH | LV_CPZ | -0.008 | 0.078 | -0.102 | 0.919 | -0.10256 | 0.918309 |
| SITE07.WITH | LV_CPZ | -0.014 | 0.052 | -0.272 | 0.786 | -0.26923 | 0.787752 |
| SITE08.WITH | LV_CPZ | 0.131 | 0.062 | 2.105 | 0.035 | 2.112903 | 0.034609 |
| SITE09.WITH | LV_CPZ | -0.023 | 0.042 | -0.556 | 0.578 | -0.54762 | 0.583954 |
| AGE.WITH | FEMALE | -0.147 | 0.052 | -2.854 | 0.004 | -2.82692 | 0.0047 |
| AGE.WITH | SITE02 | 0.323 | 0.059 | 5.43 | 0 | 5.474576 | 4.39E-08 |
| AGE.WITH | SITE03 | -0.026 | 0.046 | -0.579 | 0.562 | -0.56522 | 0.571926 |
| AGE.WITH | SITE04 | 0.016 | 0.044 | 0.369 | 0.712 | 0.363636 | 0.71613 |
| AGE.WITH | SITE05 | 0.167 | 0.048 | 3.468 | 0.001 | 3.479167 | 0.000503 |
| AGE.WITH | SITE06 | -0.001 | 0.047 | -0.024 | 0.981 | -0.02128 | 0.983025 |
| AGE.WITH | SITE07 | -0.154 | 0.045 | -3.404 | 0.001 | -3.42222 | 0.000621 |
| AGE.WITH | SITE08 | -0.193 | 0.05 | -3.891 | 0 | -3.86 | 0.000113 |
| AGE.WITH | SITE09 | -0.028 | 0.048 | -0.573 | 0.567 | -0.58333 | 0.559669 |
| FEMALE.WITH | SITE02 | 0.043 | 0.053 | 0.807 | 0.42 | 0.811321 | 0.417181 |
| FEMALE.WITH | SITE03 | -0.022 | 0.052 | -0.426 | 0.67 | -0.42308 | 0.672239 |
| FEMALE.WITH | SITE04 | -0.044 | 0.052 | -0.858 | 0.391 | -0.84615 | 0.397467 |
| FEMALE.WITH | SITE05 | 0.012 | 0.053 | 0.224 | 0.823 | 0.226415 | 0.820879 |
| FEMALE.WITH | SITE06 | -0.069 | 0.051 | -1.335 | 0.182 | -1.35294 | 0.176074 |
| FEMALE.WITH | SITE07 | 0.085 | 0.053 | 1.596 | 0.111 | 1.603774 | 0.108764 |
| FEMALE.WITH | SITE08 | 0.011 | 0.053 | 0.215 | 0.83 | 0.207547 | 0.835583 |
| FEMALE.WITH | SITE09 | 0.038 | 0.053 | 0.713 | 0.476 | 0.716981 | 0.473386 |
| SITE02.WITH | SITE03 | -0.072 | 0.01 | -6.899 | 0 | -7.2 | 6.02E-13 |
| SITE02.WITH | SITE04 | -0.093 | 0.012 | -7.835 | 0 | -7.75 | 9.1E-15 |
| SITE02.WITH | SITE05 | -0.12 | 0.014 | -8.631 | 0 | -8.57143 | 0 |
| SITE02.WITH | SITE06 | -0.113 | 0.013 | -8.453 | 0 | -8.69231 | 0 |
| SITE02.WITH | SITE07 | -0.119 | 0.014 | -8.597 | 0 | -8.5 | 0 |
| SITE02.WITH | SITE08 | -0.104 | 0.013 | -8.201 | 0 | -8 | 1.33E-15 |
| SITE02.WITH | SITE09 | -0.124 | 0.014 | -8.726 | 0 | -8.85714 | 0 |
| SITE03.WITH | SITE04 | -0.076 | 0.011 | -7.037 | 0 | -6.90909 | 4.88E-12 |
| SITE03.WITH | SITE05 | -0.098 | 0.013 | -7.596 | 0 | -7.53846 | 4.75E-14 |
| SITE03.WITH | SITE06 | -0.093 | 0.012 | -7.474 | 0 | -7.75 | 9.1E-15 |
| SITE03.WITH | SITE07 | -0.097 | 0.013 | -7.573 | 0 | -7.46154 | 8.55E-14 |
| SITE03.WITH | SITE08 | -0.085 | 0.012 | -7.299 | 0 | -7.08333 | 1.41E-12 |
| SITE03.WITH | SITE09 | -0.102 | 0.013 | -7.661 | 0 | -7.84615 | 4.22E-15 |
| SITE04.WITH | SITE05 | -0.127 | 0.014 | -8.906 | 0 | -9.07143 | 0 |
| SITE04.WITH | SITE06 | -0.119 | 0.014 | -8.711 | 0 | -8.5 | 0 |
| SITE04.WITH | SITE07 | -0.125 | 0.014 | -8.869 | 0 | -8.92857 | 0 |
| SITE04.WITH | SITE08 | -0.11 | 0.013 | -8.436 | 0 | -8.46154 | 0 |
| SITE04.WITH | SITE09 | -0.131 | 0.015 | -9.011 | 0 | -8.73333 | 0 |
| SITE05.WITH | SITE06 | -0.153 | 0.016 | -9.85 | 0 | -9.5625 | 0 |
| SITE05.WITH | SITE07 | -0.161 | 0.016 | -10.081 | 0 | -10.0625 | 0 |
| SITE05.WITH | SITE08 | -0.141 | 0.015 | -9.457 | 0 | -9.4 | 0 |
| SITE05.WITH | SITE09 | -0.169 | 0.016 | -10.292 | 0 | -10.5625 | 0 |
| SITE06.WITH | SITE07 | -0.151 | 0.015 | -9.8 | 0 | -10.0667 | 0 |
| SITE06.WITH | SITE08 | -0.133 | 0.014 | -9.224 | 0 | -9.5 | 0 |
| SITE06.WITH | SITE09 | -0.159 | 0.016 | -9.993 | 0 | -9.9375 | 0 |
| SITE07.WITH | SITE08 | -0.14 | 0.015 | -9.412 | 0 | -9.33333 | 0 |
| SITE07.WITH | SITE09 | -0.167 | 0.016 | -10.235 | 0 | -10.4375 | 0 |
| SITE08.WITH | SITE09 | -0.146 | 0.015 | -9.583 | 0 | -9.73333 | 0 |
| Means | AGE | 4.854 | 0.172 | 28.231 | 0 | 28.22093 | 0 |
| Means | FEMALE | 0.896 | 0.048 | 18.781 | 0 | 18.66667 | 0 |
| Means | SITE02 | 0.297 | 0.029 | 10.324 | 0 | 10.24138 | 0 |
| Means | SITE03 | 0.244 | 0.028 | 8.69 | 0 | 8.714286 | 0 |
| Means | SITE04 | 0.314 | 0.029 | 10.795 | 0 | 10.82759 | 0 |
| Means | SITE05 | 0.404 | 0.031 | 13.114 | 0 | 13.03226 | 0 |
| Means | SITE06 | 0.38 | 0.03 | 12.542 | 0 | 12.66667 | 0 |
| Means | SITE07 | 0.399 | 0.031 | 13.004 | 0 | 12.87097 | 0 |
| Means | SITE08 | 0.35 | 0.03 | 11.788 | 0 | 11.66667 | 0 |
| Means | SITE09 | 0.418 | 0.031 | 13.436 | 0 | 13.48387 | 0 |
| Intercepts | DEPS_CH | -0.083 | 0.301 | -0.274 | 0.784 | -0.27575 | 0.782742 |
| Intercepts | HI_CH | -0.046 | 0.052 | -0.886 | 0.375 | -0.88462 | 0.376364 |
| Intercepts | CPZ_CH | 0.086 | 0.052 | 1.668 | 0.095 | 1.653846 | 0.098159 |
| Variances | AGE | 1 | 0 |  |  |  |  |
| Variances | FEMALE | 1 | 0 |  |  |  |  |
| Variances | SITE02 | 1 | 0 |  |  |  |  |
| Variances | SITE03 | 1 | 0 |  |  |  |  |
| Variances | SITE04 | 1 | 0 |  |  |  |  |
| Variances | SITE05 | 1 | 0 |  |  |  |  |
| Variances | SITE06 | 1 | 0 |  |  |  |  |
| Variances | SITE07 | 1 | 0 |  |  |  |  |
| Variances | SITE08 | 1 | 0 |  |  |  |  |
| Variances | SITE09 | 1 | 0 |  |  |  |  |
| Variances | LV_HI | 1 | 0 |  |  |  |  |
| Variances | LV_CPZ | 1 | 0 |  |  |  |  |
| Residual.Variances | DEPS_CH | 0.288 | 0.073 | 3.945 | 0 | 3.945205 | 7.97E-05 |
| Residual.Variances | HI_CH | 0.326 | 0.042 | 7.846 | 0 | 7.761905 | 8.44E-15 |
| Residual.Variances | CPZ_CH | 0.063 | 0.014 | 4.658 | 0 | 4.5 | 6.8E-06 |
| Residual.Variances | LV_DEPS | 0.819 | 0.083 | 9.879 | 0 | 9.86747 | 0 |
| LV_HI.BY | HI_CH | 0.821 | 0.025 | 32.45 | 0 | 32.84 | 0 |
| LV_GAF.BY | GAF_CH | 0.906 | 0.015 | 60.713 | 0 | 60.4 | 0 |
| LV_CPZ.BY | CPZ_CH | 0.968 | 0.007 | 137.347 | 0 | 138.2857 | 0 |
| LV_GAF.ON | LV_HI | 0.352 | 0.103 | 3.428 | 0.001 | 3.417476 | 0.000632 |
| LV_GAF.ON | LV_CPZ | 0.005 | 0.097 | 0.049 | 0.961 | 0.051546 | 0.95889 |
| LV_GAF.ON | AGE | -0.102 | 0.06 | -1.696 | 0.09 | -1.7 | 0.089131 |
| LV_GAF.ON | FEMALE | -0.02 | 0.057 | -0.356 | 0.722 | -0.35088 | 0.72568 |
| LV_GAF.ON | SITE02 | -0.049 | 0.074 | -0.667 | 0.505 | -0.66216 | 0.507867 |
| LV_GAF.ON | SITE03 | -0.126 | 0.062 | -2.03 | 0.042 | -2.03226 | 0.042128 |
| LV_GAF.ON | SITE04 | -0.056 | 0.066 | -0.844 | 0.399 | -0.84848 | 0.396168 |
| LV_GAF.ON | SITE05 | -0.076 | 0.08 | -0.952 | 0.341 | -0.95 | 0.342112 |
| LV_GAF.ON | SITE06 | -0.172 | 0.076 | -2.264 | 0.024 | -2.26316 | 0.023626 |
| LV_GAF.ON | SITE07 | -0.134 | 0.096 | -1.39 | 0.165 | -1.39583 | 0.162765 |
| LV_GAF.ON | SITE08 | -0.013 | 0.096 | -0.136 | 0.892 | -0.13542 | 0.892282 |
| LV_GAF.ON | SITE09 | -0.177 | 0.086 | -2.053 | 0.04 | -2.05814 | 0.039577 |
| LV_HI.WITH | AGE | 0.034 | 0.061 | 0.547 | 0.584 | 0.557377 | 0.57727 |
| LV_HI.WITH | FEMALE | -0.109 | 0.066 | -1.657 | 0.097 | -1.65152 | 0.098633 |
| LV_HI.WITH | SITE02 | -0.01 | 0.068 | -0.145 | 0.885 | -0.14706 | 0.883086 |
| LV_HI.WITH | SITE03 | 0.034 | 0.099 | 0.339 | 0.735 | 0.343434 | 0.731272 |
| LV_HI.WITH | SITE04 | 0.053 | 0.05 | 1.055 | 0.292 | 1.06 | 0.289145 |
| LV_HI.WITH | SITE05 | -0.089 | 0.069 | -1.298 | 0.194 | -1.28986 | 0.197101 |
| LV_HI.WITH | SITE06 | 0.091 | 0.048 | 1.897 | 0.058 | 1.895833 | 0.057982 |
| LV_HI.WITH | SITE07 | 0.017 | 0.071 | 0.242 | 0.809 | 0.239437 | 0.810767 |
| LV_HI.WITH | SITE08 | -0.042 | 0.054 | -0.785 | 0.432 | -0.77778 | 0.4367 |
| LV_HI.WITH | SITE09 | -0.039 | 0.065 | -0.603 | 0.547 | -0.6 | 0.548506 |
| LV_HI.WITH | LV_CPZ | 0.037 | 0.1 | 0.372 | 0.71 | 0.37 | 0.711382 |
| AGE.WITH | LV_CPZ | -0.069 | 0.059 | -1.172 | 0.241 | -1.16949 | 0.242206 |
| FEMALE.WITH | LV_CPZ | -0.007 | 0.053 | -0.125 | 0.901 | -0.13208 | 0.894925 |
| SITE02.WITH | LV_CPZ | -0.062 | 0.044 | -1.413 | 0.158 | -1.40909 | 0.158808 |
| SITE03.WITH | LV_CPZ | 0.016 | 0.05 | 0.319 | 0.749 | 0.32 | 0.748968 |
| SITE04.WITH | LV_CPZ | 0.05 | 0.048 | 1.04 | 0.298 | 1.041667 | 0.297566 |
| SITE05.WITH | LV_CPZ | 0.023 | 0.04 | 0.579 | 0.562 | 0.575 | 0.565291 |
| SITE06.WITH | LV_CPZ | -0.008 | 0.078 | -0.103 | 0.918 | -0.10256 | 0.918309 |
| SITE07.WITH | LV_CPZ | -0.014 | 0.052 | -0.273 | 0.785 | -0.26923 | 0.787752 |
| SITE08.WITH | LV_CPZ | 0.131 | 0.062 | 2.105 | 0.035 | 2.112903 | 0.034609 |
| SITE09.WITH | LV_CPZ | -0.023 | 0.042 | -0.557 | 0.577 | -0.54762 | 0.583954 |
| AGE.WITH | FEMALE | -0.149 | 0.052 | -2.887 | 0.004 | -2.86538 | 0.004165 |
| AGE.WITH | SITE02 | 0.323 | 0.059 | 5.422 | 0 | 5.474576 | 4.39E-08 |
| AGE.WITH | SITE03 | -0.027 | 0.046 | -0.589 | 0.556 | -0.58696 | 0.557233 |
| AGE.WITH | SITE04 | 0.016 | 0.044 | 0.355 | 0.722 | 0.363636 | 0.71613 |
| AGE.WITH | SITE05 | 0.166 | 0.048 | 3.454 | 0.001 | 3.458333 | 0.000544 |
| AGE.WITH | SITE06 | -0.002 | 0.047 | -0.039 | 0.969 | -0.04255 | 0.966058 |
| AGE.WITH | SITE07 | -0.154 | 0.045 | -3.42 | 0.001 | -3.42222 | 0.000621 |
| AGE.WITH | SITE08 | -0.194 | 0.05 | -3.904 | 0 | -3.88 | 0.000104 |
| AGE.WITH | SITE09 | -0.028 | 0.048 | -0.589 | 0.556 | -0.58333 | 0.559669 |
| FEMALE.WITH | SITE02 | 0.042 | 0.053 | 0.793 | 0.428 | 0.792453 | 0.428097 |
| FEMALE.WITH | SITE03 | -0.023 | 0.052 | -0.437 | 0.662 | -0.44231 | 0.658267 |
| FEMALE.WITH | SITE04 | -0.045 | 0.052 | -0.873 | 0.383 | -0.86538 | 0.386828 |
| FEMALE.WITH | SITE05 | 0.011 | 0.053 | 0.205 | 0.838 | 0.207547 | 0.835583 |
| FEMALE.WITH | SITE06 | -0.07 | 0.051 | -1.354 | 0.176 | -1.37255 | 0.169893 |
| FEMALE.WITH | SITE07 | 0.084 | 0.053 | 1.577 | 0.115 | 1.584906 | 0.112988 |
| FEMALE.WITH | SITE08 | 0.011 | 0.053 | 0.198 | 0.843 | 0.207547 | 0.835583 |
| FEMALE.WITH | SITE09 | 0.037 | 0.053 | 0.694 | 0.488 | 0.698113 | 0.485106 |
| SITE02.WITH | SITE03 | -0.073 | 0.011 | -6.899 | 0 | -6.63636 | 3.22E-11 |
| SITE02.WITH | SITE04 | -0.094 | 0.012 | -7.835 | 0 | -7.83333 | 4.66E-15 |
| SITE02.WITH | SITE05 | -0.12 | 0.014 | -8.631 | 0 | -8.57143 | 0 |
| SITE02.WITH | SITE06 | -0.113 | 0.013 | -8.453 | 0 | -8.69231 | 0 |
| SITE02.WITH | SITE07 | -0.119 | 0.014 | -8.598 | 0 | -8.5 | 0 |
| SITE02.WITH | SITE08 | -0.104 | 0.013 | -8.201 | 0 | -8 | 1.33E-15 |
| SITE02.WITH | SITE09 | -0.125 | 0.014 | -8.727 | 0 | -8.92857 | 0 |
| SITE03.WITH | SITE04 | -0.077 | 0.011 | -7.037 | 0 | -7 | 2.56E-12 |
| SITE03.WITH | SITE05 | -0.099 | 0.013 | -7.596 | 0 | -7.61538 | 2.62E-14 |
| SITE03.WITH | SITE06 | -0.093 | 0.012 | -7.474 | 0 | -7.75 | 9.1E-15 |
| SITE03.WITH | SITE07 | -0.097 | 0.013 | -7.573 | 0 | -7.46154 | 8.55E-14 |
| SITE03.WITH | SITE08 | -0.086 | 0.012 | -7.299 | 0 | -7.16667 | 7.68E-13 |
| SITE03.WITH | SITE09 | -0.102 | 0.013 | -7.661 | 0 | -7.84615 | 4.22E-15 |
| SITE04.WITH | SITE05 | -0.127 | 0.014 | -8.907 | 0 | -9.07143 | 0 |
| SITE04.WITH | SITE06 | -0.12 | 0.014 | -8.711 | 0 | -8.57143 | 0 |
| SITE04.WITH | SITE07 | -0.126 | 0.014 | -8.87 | 0 | -9 | 0 |
| SITE04.WITH | SITE08 | -0.11 | 0.013 | -8.437 | 0 | -8.46154 | 0 |
| SITE04.WITH | SITE09 | -0.131 | 0.015 | -9.012 | 0 | -8.73333 | 0 |
| SITE05.WITH | SITE06 | -0.154 | 0.016 | -9.85 | 0 | -9.625 | 0 |
| SITE05.WITH | SITE07 | -0.161 | 0.016 | -10.082 | 0 | -10.0625 | 0 |
| SITE05.WITH | SITE08 | -0.142 | 0.015 | -9.457 | 0 | -9.46667 | 0 |
| SITE05.WITH | SITE09 | -0.169 | 0.016 | -10.293 | 0 | -10.5625 | 0 |
| SITE06.WITH | SITE07 | -0.152 | 0.016 | -9.8 | 0 | -9.5 | 0 |
| SITE06.WITH | SITE08 | -0.133 | 0.014 | -9.224 | 0 | -9.5 | 0 |
| SITE06.WITH | SITE09 | -0.159 | 0.016 | -9.994 | 0 | -9.9375 | 0 |
| SITE07.WITH | SITE08 | -0.14 | 0.015 | -9.413 | 0 | -9.33333 | 0 |
| SITE07.WITH | SITE09 | -0.167 | 0.016 | -10.236 | 0 | -10.4375 | 0 |
| SITE08.WITH | SITE09 | -0.147 | 0.015 | -9.584 | 0 | -9.8 | 0 |
| Means | AGE | 4.852 | 0.172 | 28.218 | 0 | 28.2093 | 0 |
| Means | FEMALE | 0.898 | 0.048 | 18.76 | 0 | 18.70833 | 0 |
| Means | SITE02 | 0.298 | 0.029 | 10.322 | 0 | 10.27586 | 0 |
| Means | SITE03 | 0.244 | 0.028 | 8.689 | 0 | 8.714286 | 0 |
| Means | SITE04 | 0.314 | 0.029 | 10.793 | 0 | 10.82759 | 0 |
| Means | SITE05 | 0.404 | 0.031 | 13.111 | 0 | 13.03226 | 0 |
| Means | SITE06 | 0.38 | 0.03 | 12.54 | 0 | 12.66667 | 0 |
| Means | SITE07 | 0.4 | 0.031 | 13.001 | 0 | 12.90323 | 0 |
| Means | SITE08 | 0.351 | 0.03 | 11.786 | 0 | 11.7 | 0 |
| Means | SITE09 | 0.418 | 0.031 | 13.433 | 0 | 13.48387 | 0 |
| Intercepts | GAF_CH | 1.031 | 0.312 | 3.305 | 0.001 | 3.304487 | 0.000952 |
| Intercepts | HI_CH | -0.045 | 0.052 | -0.855 | 0.393 | -0.86538 | 0.386828 |
| Intercepts | CPZ_CH | 0.086 | 0.052 | 1.664 | 0.096 | 1.653846 | 0.098159 |
| Variances | AGE | 1 | 0 |  |  |  |  |
| Variances | FEMALE | 1 | 0 |  |  |  |  |
| Variances | SITE02 | 1 | 0 |  |  |  |  |
| Variances | SITE03 | 1 | 0 |  |  |  |  |
| Variances | SITE04 | 1 | 0 |  |  |  |  |
| Variances | SITE05 | 1 | 0 |  |  |  |  |
| Variances | SITE06 | 1 | 0 |  |  |  |  |
| Variances | SITE07 | 1 | 0 |  |  |  |  |
| Variances | SITE08 | 1 | 0 |  |  |  |  |
| Variances | SITE09 | 1 | 0 |  |  |  |  |
| Variances | LV_HI | 1 | 0 |  |  |  |  |
| Variances | LV_CPZ | 1 | 0 |  |  |  |  |
| Residual.Variances | GAF_CH | 0.179 | 0.027 | 6.606 | 0 | 6.62963 | 3.37E-11 |
| Residual.Variances | HI_CH | 0.326 | 0.042 | 7.838 | 0 | 7.761905 | 8.44E-15 |
| Residual.Variances | CPZ_CH | 0.064 | 0.014 | 4.657 | 0 | 4.571429 | 4.84E-06 |
| Residual.Variances | LV_GAF | 0.838 | 0.068 | 12.355 | 0 | 12.32353 | 0 |
| LV_HI.BY | HI_CH | 0.819 | 0.026 | 31.757 | 0 | 31.5 | 0 |
| LV_NEGS.BY | NEGS_CH | 0.899 | 0.022 | 39.997 | 0 | 40.86364 | 0 |
| LV_CPZ.BY | CPZ_CH | 0.968 | 0.007 | 136.404 | 0 | 138.2857 | 0 |
| LV_NEGS.ON | LV_HI | -0.363 | 0.082 | -4.404 | 0 | -4.42683 | 9.56E-06 |
| LV_NEGS.ON | LV_CPZ | 0.07 | 0.075 | 0.934 | 0.35 | 0.933333 | 0.350648 |
| LV_NEGS.ON | AGE | -0.012 | 0.065 | -0.184 | 0.854 | -0.18462 | 0.853531 |
| LV_NEGS.ON | FEMALE | 0.005 | 0.053 | 0.089 | 0.929 | 0.09434 | 0.924839 |
| LV_NEGS.ON | SITE02 | -0.192 | 0.086 | -2.231 | 0.026 | -2.23256 | 0.025578 |
| LV_NEGS.ON | SITE03 | 0.082 | 0.073 | 1.119 | 0.263 | 1.123288 | 0.261315 |
| LV_NEGS.ON | SITE04 | -0.124 | 0.072 | -1.725 | 0.085 | -1.72222 | 0.085029 |
| LV_NEGS.ON | SITE05 | -0.027 | 0.086 | -0.311 | 0.756 | -0.31395 | 0.753556 |
| LV_NEGS.ON | SITE06 | -0.101 | 0.087 | -1.171 | 0.242 | -1.16092 | 0.245675 |
| LV_NEGS.ON | SITE07 | -0.097 | 0.082 | -1.178 | 0.239 | -1.18293 | 0.236838 |
| LV_NEGS.ON | SITE08 | -0.164 | 0.083 | -1.973 | 0.048 | -1.9759 | 0.048166 |
| LV_NEGS.ON | SITE09 | 0.045 | 0.101 | 0.442 | 0.658 | 0.445545 | 0.655926 |
| LV_HI.WITH | AGE | 0.037 | 0.062 | 0.588 | 0.556 | 0.596774 | 0.550658 |
| LV_HI.WITH | FEMALE | -0.099 | 0.066 | -1.494 | 0.135 | -1.5 | 0.133614 |
| LV_HI.WITH | SITE02 | -0.006 | 0.069 | -0.088 | 0.93 | -0.08696 | 0.930706 |
| LV_HI.WITH | SITE03 | -0.012 | 0.092 | -0.131 | 0.896 | -0.13043 | 0.896222 |
| LV_HI.WITH | SITE04 | 0.058 | 0.051 | 1.14 | 0.254 | 1.137255 | 0.255432 |
| LV_HI.WITH | SITE05 | -0.085 | 0.07 | -1.222 | 0.222 | -1.21429 | 0.224639 |
| LV_HI.WITH | SITE06 | 0.098 | 0.049 | 2.015 | 0.044 | 2 | 0.0455 |
| LV_HI.WITH | SITE07 | 0.023 | 0.072 | 0.316 | 0.752 | 0.319444 | 0.74939 |
| LV_HI.WITH | SITE08 | -0.038 | 0.055 | -0.703 | 0.482 | -0.69091 | 0.489623 |
| LV_HI.WITH | SITE09 | -0.034 | 0.066 | -0.52 | 0.603 | -0.51515 | 0.606447 |
| LV_HI.WITH | LV_CPZ | 0.026 | 0.101 | 0.256 | 0.798 | 0.257426 | 0.79685 |
| AGE.WITH | LV_CPZ | -0.069 | 0.058 | -1.18 | 0.238 | -1.18966 | 0.234182 |
| FEMALE.WITH | LV_CPZ | -0.003 | 0.053 | -0.057 | 0.954 | -0.0566 | 0.954861 |
| SITE02.WITH | LV_CPZ | -0.062 | 0.044 | -1.4 | 0.162 | -1.40909 | 0.158808 |
| SITE03.WITH | LV_CPZ | 0.005 | 0.05 | 0.1 | 0.92 | 0.1 | 0.920344 |
| SITE04.WITH | LV_CPZ | 0.051 | 0.049 | 1.052 | 0.293 | 1.040816 | 0.297961 |
| SITE05.WITH | LV_CPZ | 0.024 | 0.04 | 0.6 | 0.549 | 0.6 | 0.548506 |
| SITE06.WITH | LV_CPZ | -0.007 | 0.078 | -0.093 | 0.926 | -0.08974 | 0.928491 |
| SITE07.WITH | LV_CPZ | -0.013 | 0.052 | -0.257 | 0.797 | -0.25 | 0.802587 |
| SITE08.WITH | LV_CPZ | 0.132 | 0.063 | 2.114 | 0.035 | 2.095238 | 0.03615 |
| SITE09.WITH | LV_CPZ | -0.022 | 0.042 | -0.535 | 0.593 | -0.52381 | 0.600411 |
| AGE.WITH | FEMALE | -0.148 | 0.052 | -2.859 | 0.004 | -2.84615 | 0.004425 |
| AGE.WITH | SITE02 | 0.323 | 0.059 | 5.428 | 0 | 5.474576 | 4.39E-08 |
| AGE.WITH | SITE03 | -0.026 | 0.047 | -0.55 | 0.582 | -0.55319 | 0.580132 |
| AGE.WITH | SITE04 | 0.016 | 0.044 | 0.366 | 0.714 | 0.363636 | 0.71613 |
| AGE.WITH | SITE05 | 0.167 | 0.048 | 3.465 | 0.001 | 3.479167 | 0.000503 |
| AGE.WITH | SITE06 | -0.001 | 0.047 | -0.027 | 0.979 | -0.02128 | 0.983025 |
| AGE.WITH | SITE07 | -0.154 | 0.045 | -3.406 | 0.001 | -3.42222 | 0.000621 |
| AGE.WITH | SITE08 | -0.193 | 0.05 | -3.893 | 0 | -3.86 | 0.000113 |
| AGE.WITH | SITE09 | -0.028 | 0.048 | -0.576 | 0.565 | -0.58333 | 0.559669 |
| FEMALE.WITH | SITE02 | 0.042 | 0.053 | 0.793 | 0.428 | 0.792453 | 0.428097 |
| FEMALE.WITH | SITE03 | -0.012 | 0.053 | -0.232 | 0.817 | -0.22642 | 0.820879 |
| FEMALE.WITH | SITE04 | -0.045 | 0.052 | -0.873 | 0.383 | -0.86538 | 0.386828 |
| FEMALE.WITH | SITE05 | 0.011 | 0.053 | 0.205 | 0.838 | 0.207547 | 0.835583 |
| FEMALE.WITH | SITE06 | -0.07 | 0.051 | -1.354 | 0.176 | -1.37255 | 0.169893 |
| FEMALE.WITH | SITE07 | 0.084 | 0.053 | 1.577 | 0.115 | 1.584906 | 0.112988 |
| FEMALE.WITH | SITE08 | 0.011 | 0.053 | 0.198 | 0.843 | 0.207547 | 0.835583 |
| FEMALE.WITH | SITE09 | 0.037 | 0.053 | 0.694 | 0.488 | 0.698113 | 0.485106 |
| SITE02.WITH | SITE03 | -0.071 | 0.01 | -6.793 | 0 | -7.1 | 1.25E-12 |
| SITE02.WITH | SITE04 | -0.094 | 0.012 | -7.835 | 0 | -7.83333 | 4.66E-15 |
| SITE02.WITH | SITE05 | -0.12 | 0.014 | -8.631 | 0 | -8.57143 | 0 |
| SITE02.WITH | SITE06 | -0.113 | 0.013 | -8.453 | 0 | -8.69231 | 0 |
| SITE02.WITH | SITE07 | -0.119 | 0.014 | -8.598 | 0 | -8.5 | 0 |
| SITE02.WITH | SITE08 | -0.104 | 0.013 | -8.201 | 0 | -8 | 1.33E-15 |
| SITE02.WITH | SITE09 | -0.125 | 0.014 | -8.727 | 0 | -8.92857 | 0 |
| SITE03.WITH | SITE04 | -0.075 | 0.011 | -6.925 | 0 | -6.81818 | 9.22E-12 |
| SITE03.WITH | SITE05 | -0.096 | 0.013 | -7.456 | 0 | -7.38462 | 1.53E-13 |
| SITE03.WITH | SITE06 | -0.09 | 0.012 | -7.34 | 0 | -7.5 | 6.37E-14 |
| SITE03.WITH | SITE07 | -0.095 | 0.013 | -7.434 | 0 | -7.30769 | 2.72E-13 |
| SITE03.WITH | SITE08 | -0.083 | 0.012 | -7.174 | 0 | -6.91667 | 4.62E-12 |
| SITE03.WITH | SITE09 | -0.099 | 0.013 | -7.517 | 0 | -7.61538 | 2.62E-14 |
| SITE04.WITH | SITE05 | -0.127 | 0.014 | -8.907 | 0 | -9.07143 | 0 |
| SITE04.WITH | SITE06 | -0.12 | 0.014 | -8.711 | 0 | -8.57143 | 0 |
| SITE04.WITH | SITE07 | -0.126 | 0.014 | -8.87 | 0 | -9 | 0 |
| SITE04.WITH | SITE08 | -0.11 | 0.013 | -8.437 | 0 | -8.46154 | 0 |
| SITE04.WITH | SITE09 | -0.131 | 0.015 | -9.012 | 0 | -8.73333 | 0 |
| SITE05.WITH | SITE06 | -0.154 | 0.016 | -9.85 | 0 | -9.625 | 0 |
| SITE05.WITH | SITE07 | -0.161 | 0.016 | -10.082 | 0 | -10.0625 | 0 |
| SITE05.WITH | SITE08 | -0.142 | 0.015 | -9.457 | 0 | -9.46667 | 0 |
| SITE05.WITH | SITE09 | -0.169 | 0.016 | -10.293 | 0 | -10.5625 | 0 |
| SITE06.WITH | SITE07 | -0.152 | 0.016 | -9.8 | 0 | -9.5 | 0 |
| SITE06.WITH | SITE08 | -0.133 | 0.014 | -9.224 | 0 | -9.5 | 0 |
| SITE06.WITH | SITE09 | -0.159 | 0.016 | -9.994 | 0 | -9.9375 | 0 |
| SITE07.WITH | SITE08 | -0.14 | 0.015 | -9.413 | 0 | -9.33333 | 0 |
| SITE07.WITH | SITE09 | -0.167 | 0.016 | -10.236 | 0 | -10.4375 | 0 |
| SITE08.WITH | SITE09 | -0.147 | 0.015 | -9.584 | 0 | -9.8 | 0 |
| Means | AGE | 4.848 | 0.172 | 28.258 | 0 | 28.18605 | 0 |
| Means | FEMALE | 0.898 | 0.048 | 18.76 | 0 | 18.70833 | 0 |
| Means | SITE02 | 0.298 | 0.029 | 10.322 | 0 | 10.27586 | 0 |
| Means | SITE03 | 0.237 | 0.028 | 8.482 | 0 | 8.464286 | 0 |
| Means | SITE04 | 0.314 | 0.029 | 10.793 | 0 | 10.82759 | 0 |
| Means | SITE05 | 0.404 | 0.031 | 13.111 | 0 | 13.03226 | 0 |
| Means | SITE06 | 0.38 | 0.03 | 12.54 | 0 | 12.66667 | 0 |
| Means | SITE07 | 0.4 | 0.031 | 13.001 | 0 | 12.90323 | 0 |
| Means | SITE08 | 0.351 | 0.03 | 11.786 | 0 | 11.7 | 0 |
| Means | SITE09 | 0.418 | 0.031 | 13.433 | 0 | 13.48387 | 0 |
| Intercepts | NEGS_CH | -0.035 | 0.331 | -0.105 | 0.916 | -0.10574 | 0.915788 |
| Intercepts | HI_CH | -0.056 | 0.052 | -1.078 | 0.281 | -1.07692 | 0.281515 |
| Intercepts | CPZ_CH | 0.084 | 0.052 | 1.619 | 0.105 | 1.615385 | 0.106227 |
| Variances | AGE | 1 | 0 |  |  |  |  |
| Variances | FEMALE | 1 | 0 |  |  |  |  |
| Variances | SITE02 | 1 | 0 |  |  |  |  |
| Variances | SITE03 | 1 | 0 |  |  |  |  |
| Variances | SITE04 | 1 | 0 |  |  |  |  |
| Variances | SITE05 | 1 | 0 |  |  |  |  |
| Variances | SITE06 | 1 | 0 |  |  |  |  |
| Variances | SITE07 | 1 | 0 |  |  |  |  |
| Variances | SITE08 | 1 | 0 |  |  |  |  |
| Variances | SITE09 | 1 | 0 |  |  |  |  |
| Variances | LV_HI | 1 | 0 |  |  |  |  |
| Variances | LV_CPZ | 1 | 0 |  |  |  |  |
| Residual.Variances | NEGS_CH | 0.192 | 0.04 | 4.76 | 0 | 4.8 | 1.59E-06 |
| Residual.Variances | HI_CH | 0.329 | 0.042 | 7.802 | 0 | 7.833333 | 4.66E-15 |
| Residual.Variances | CPZ_CH | 0.064 | 0.014 | 4.646 | 0 | 4.571429 | 4.84E-06 |
| Residual.Variances | LV_NEGS | 0.778 | 0.07 | 11.136 | 0 | 11.11429 | 0 |
| LV_HI.BY | HI_CH | 0.821 | 0.025 | 32.421 | 0 | 32.84 | 0 |
| LV_POSS.BY | POSS_CH | 0.914 | 0.02 | 44.91 | 0 | 45.7 | 0 |
| LV_CPZ.BY | CPZ_CH | 0.968 | 0.007 | 137.581 | 0 | 138.2857 | 0 |
| LV_POSS.ON | LV_HI | -0.174 | 0.115 | -1.521 | 0.128 | -1.51304 | 0.130269 |
| LV_POSS.ON | LV_CPZ | 0.137 | 0.059 | 2.337 | 0.019 | 2.322034 | 0.020231 |
| LV_POSS.ON | AGE | -0.009 | 0.061 | -0.151 | 0.88 | -0.14754 | 0.882705 |
| LV_POSS.ON | FEMALE | 0.056 | 0.054 | 1.041 | 0.298 | 1.037037 | 0.299719 |
| LV_POSS.ON | SITE02 | 0.015 | 0.108 | 0.134 | 0.893 | 0.138889 | 0.889538 |
| LV_POSS.ON | SITE03 | 0.051 | 0.067 | 0.769 | 0.442 | 0.761194 | 0.446541 |
| LV_POSS.ON | SITE04 | 0.025 | 0.07 | 0.356 | 0.722 | 0.357143 | 0.720985 |
| LV_POSS.ON | SITE05 | 0.13 | 0.083 | 1.564 | 0.118 | 1.566265 | 0.117287 |
| LV_POSS.ON | SITE06 | -0.02 | 0.078 | -0.253 | 0.8 | -0.25641 | 0.797634 |
| LV_POSS.ON | SITE07 | 0.039 | 0.075 | 0.521 | 0.602 | 0.52 | 0.603064 |
| LV_POSS.ON | SITE08 | 0.045 | 0.08 | 0.559 | 0.576 | 0.5625 | 0.573775 |
| LV_POSS.ON | SITE09 | 0.051 | 0.084 | 0.608 | 0.543 | 0.607143 | 0.543756 |
| LV_HI.WITH | AGE | 0.035 | 0.061 | 0.566 | 0.571 | 0.57377 | 0.566123 |
| LV_HI.WITH | FEMALE | -0.107 | 0.066 | -1.632 | 0.103 | -1.62121 | 0.104972 |
| LV_HI.WITH | SITE02 | -0.009 | 0.067 | -0.137 | 0.891 | -0.13433 | 0.893143 |
| LV_HI.WITH | SITE03 | 0.034 | 0.099 | 0.343 | 0.731 | 0.343434 | 0.731272 |
| LV_HI.WITH | SITE04 | 0.053 | 0.05 | 1.066 | 0.286 | 1.06 | 0.289145 |
| LV_HI.WITH | SITE05 | -0.088 | 0.069 | -1.287 | 0.198 | -1.27536 | 0.202181 |
| LV_HI.WITH | SITE06 | 0.092 | 0.048 | 1.912 | 0.056 | 1.916667 | 0.05528 |
| LV_HI.WITH | SITE07 | 0.018 | 0.071 | 0.253 | 0.8 | 0.253521 | 0.799866 |
| LV_HI.WITH | SITE08 | -0.042 | 0.054 | -0.773 | 0.439 | -0.77778 | 0.4367 |
| LV_HI.WITH | SITE09 | -0.038 | 0.065 | -0.591 | 0.555 | -0.58462 | 0.558806 |
| LV_HI.WITH | LV_CPZ | 0.037 | 0.1 | 0.369 | 0.712 | 0.37 | 0.711382 |
| AGE.WITH | LV_CPZ | -0.069 | 0.059 | -1.172 | 0.241 | -1.16949 | 0.242206 |
| FEMALE.WITH | LV_CPZ | -0.006 | 0.053 | -0.116 | 0.908 | -0.11321 | 0.909866 |
| SITE02.WITH | LV_CPZ | -0.062 | 0.044 | -1.413 | 0.158 | -1.40909 | 0.158808 |
| SITE03.WITH | LV_CPZ | 0.016 | 0.05 | 0.32 | 0.749 | 0.32 | 0.748968 |
| SITE04.WITH | LV_CPZ | 0.05 | 0.048 | 1.04 | 0.298 | 1.041667 | 0.297566 |
| SITE05.WITH | LV_CPZ | 0.023 | 0.04 | 0.58 | 0.562 | 0.575 | 0.565291 |
| SITE06.WITH | LV_CPZ | -0.008 | 0.078 | -0.103 | 0.918 | -0.10256 | 0.918309 |
| SITE07.WITH | LV_CPZ | -0.014 | 0.052 | -0.273 | 0.785 | -0.26923 | 0.787752 |
| SITE08.WITH | LV_CPZ | 0.131 | 0.062 | 2.104 | 0.035 | 2.112903 | 0.034609 |
| SITE09.WITH | LV_CPZ | -0.023 | 0.042 | -0.557 | 0.577 | -0.54762 | 0.583954 |
| AGE.WITH | FEMALE | -0.147 | 0.052 | -2.854 | 0.004 | -2.82692 | 0.0047 |
| AGE.WITH | SITE02 | 0.323 | 0.059 | 5.43 | 0 | 5.474576 | 4.39E-08 |
| AGE.WITH | SITE03 | -0.026 | 0.046 | -0.579 | 0.562 | -0.56522 | 0.571926 |
| AGE.WITH | SITE04 | 0.016 | 0.044 | 0.369 | 0.712 | 0.363636 | 0.71613 |
| AGE.WITH | SITE05 | 0.167 | 0.048 | 3.468 | 0.001 | 3.479167 | 0.000503 |
| AGE.WITH | SITE06 | -0.001 | 0.047 | -0.024 | 0.981 | -0.02128 | 0.983025 |
| AGE.WITH | SITE07 | -0.154 | 0.045 | -3.404 | 0.001 | -3.42222 | 0.000621 |
| AGE.WITH | SITE08 | -0.193 | 0.05 | -3.891 | 0 | -3.86 | 0.000113 |
| AGE.WITH | SITE09 | -0.028 | 0.048 | -0.573 | 0.567 | -0.58333 | 0.559669 |
| FEMALE.WITH | SITE02 | 0.043 | 0.053 | 0.807 | 0.42 | 0.811321 | 0.417181 |
| FEMALE.WITH | SITE03 | -0.022 | 0.052 | -0.426 | 0.67 | -0.42308 | 0.672239 |
| FEMALE.WITH | SITE04 | -0.044 | 0.052 | -0.858 | 0.391 | -0.84615 | 0.397467 |
| FEMALE.WITH | SITE05 | 0.012 | 0.053 | 0.224 | 0.823 | 0.226415 | 0.820879 |
| FEMALE.WITH | SITE06 | -0.069 | 0.051 | -1.335 | 0.182 | -1.35294 | 0.176074 |
| FEMALE.WITH | SITE07 | 0.085 | 0.053 | 1.596 | 0.111 | 1.603774 | 0.108764 |
| FEMALE.WITH | SITE08 | 0.011 | 0.053 | 0.215 | 0.83 | 0.207547 | 0.835583 |
| FEMALE.WITH | SITE09 | 0.038 | 0.053 | 0.713 | 0.476 | 0.716981 | 0.473386 |
| SITE02.WITH | SITE03 | -0.072 | 0.01 | -6.899 | 0 | -7.2 | 6.02E-13 |
| SITE02.WITH | SITE04 | -0.093 | 0.012 | -7.835 | 0 | -7.75 | 9.1E-15 |
| SITE02.WITH | SITE05 | -0.12 | 0.014 | -8.631 | 0 | -8.57143 | 0 |
| SITE02.WITH | SITE06 | -0.113 | 0.013 | -8.453 | 0 | -8.69231 | 0 |
| SITE02.WITH | SITE07 | -0.119 | 0.014 | -8.597 | 0 | -8.5 | 0 |
| SITE02.WITH | SITE08 | -0.104 | 0.013 | -8.201 | 0 | -8 | 1.33E-15 |
| SITE02.WITH | SITE09 | -0.124 | 0.014 | -8.726 | 0 | -8.85714 | 0 |
| SITE03.WITH | SITE04 | -0.076 | 0.011 | -7.037 | 0 | -6.90909 | 4.88E-12 |
| SITE03.WITH | SITE05 | -0.098 | 0.013 | -7.596 | 0 | -7.53846 | 4.75E-14 |
| SITE03.WITH | SITE06 | -0.093 | 0.012 | -7.474 | 0 | -7.75 | 9.1E-15 |
| SITE03.WITH | SITE07 | -0.097 | 0.013 | -7.573 | 0 | -7.46154 | 8.55E-14 |
| SITE03.WITH | SITE08 | -0.085 | 0.012 | -7.299 | 0 | -7.08333 | 1.41E-12 |
| SITE03.WITH | SITE09 | -0.102 | 0.013 | -7.661 | 0 | -7.84615 | 4.22E-15 |
| SITE04.WITH | SITE05 | -0.127 | 0.014 | -8.906 | 0 | -9.07143 | 0 |
| SITE04.WITH | SITE06 | -0.119 | 0.014 | -8.711 | 0 | -8.5 | 0 |
| SITE04.WITH | SITE07 | -0.125 | 0.014 | -8.869 | 0 | -8.92857 | 0 |
| SITE04.WITH | SITE08 | -0.11 | 0.013 | -8.436 | 0 | -8.46154 | 0 |
| SITE04.WITH | SITE09 | -0.131 | 0.015 | -9.011 | 0 | -8.73333 | 0 |
| SITE05.WITH | SITE06 | -0.153 | 0.016 | -9.85 | 0 | -9.5625 | 0 |
| SITE05.WITH | SITE07 | -0.161 | 0.016 | -10.081 | 0 | -10.0625 | 0 |
| SITE05.WITH | SITE08 | -0.141 | 0.015 | -9.457 | 0 | -9.4 | 0 |
| SITE05.WITH | SITE09 | -0.169 | 0.016 | -10.292 | 0 | -10.5625 | 0 |
| SITE06.WITH | SITE07 | -0.151 | 0.015 | -9.8 | 0 | -10.0667 | 0 |
| SITE06.WITH | SITE08 | -0.133 | 0.014 | -9.224 | 0 | -9.5 | 0 |
| SITE06.WITH | SITE09 | -0.159 | 0.016 | -9.993 | 0 | -9.9375 | 0 |
| SITE07.WITH | SITE08 | -0.14 | 0.015 | -9.412 | 0 | -9.33333 | 0 |
| SITE07.WITH | SITE09 | -0.167 | 0.016 | -10.235 | 0 | -10.4375 | 0 |
| SITE08.WITH | SITE09 | -0.146 | 0.015 | -9.583 | 0 | -9.73333 | 0 |
| Means | AGE | 4.854 | 0.172 | 28.231 | 0 | 28.22093 | 0 |
| Means | FEMALE | 0.896 | 0.048 | 18.781 | 0 | 18.66667 | 0 |
| Means | SITE02 | 0.297 | 0.029 | 10.324 | 0 | 10.24138 | 0 |
| Means | SITE03 | 0.244 | 0.028 | 8.69 | 0 | 8.714286 | 0 |
| Means | SITE04 | 0.314 | 0.029 | 10.795 | 0 | 10.82759 | 0 |
| Means | SITE05 | 0.404 | 0.031 | 13.114 | 0 | 13.03226 | 0 |
| Means | SITE06 | 0.38 | 0.03 | 12.542 | 0 | 12.66667 | 0 |
| Means | SITE07 | 0.399 | 0.031 | 13.004 | 0 | 12.87097 | 0 |
| Means | SITE08 | 0.35 | 0.03 | 11.788 | 0 | 11.66667 | 0 |
| Means | SITE09 | 0.418 | 0.031 | 13.436 | 0 | 13.48387 | 0 |
| Intercepts | POSS_CH | -0.838 | 0.352 | -2.38 | 0.017 | -2.38068 | 0.017281 |
| Intercepts | HI_CH | -0.046 | 0.052 | -0.886 | 0.375 | -0.88462 | 0.376364 |
| Intercepts | CPZ_CH | 0.086 | 0.052 | 1.67 | 0.095 | 1.653846 | 0.098159 |
| Variances | AGE | 1 | 0 |  |  |  |  |
| Variances | FEMALE | 1 | 0 |  |  |  |  |
| Variances | SITE02 | 1 | 0 |  |  |  |  |
| Variances | SITE03 | 1 | 0 |  |  |  |  |
| Variances | SITE04 | 1 | 0 |  |  |  |  |
| Variances | SITE05 | 1 | 0 |  |  |  |  |
| Variances | SITE06 | 1 | 0 |  |  |  |  |
| Variances | SITE07 | 1 | 0 |  |  |  |  |
| Variances | SITE08 | 1 | 0 |  |  |  |  |
| Variances | SITE09 | 1 | 0 |  |  |  |  |
| Variances | LV_HI | 1 | 0 |  |  |  |  |
| Variances | LV_CPZ | 1 | 0 |  |  |  |  |
| Residual.Variances | POSS_CH | 0.165 | 0.037 | 4.423 | 0 | 4.459459 | 8.22E-06 |
| Residual.Variances | HI_CH | 0.326 | 0.042 | 7.846 | 0 | 7.761905 | 8.44E-15 |
| Residual.Variances | CPZ_CH | 0.063 | 0.014 | 4.658 | 0 | 4.5 | 6.8E-06 |
| Residual.Variances | LV_POSS | 0.923 | 0.043 | 21.34 | 0 | 21.46512 | 0 |
| LV_HI.BY | HI_CH | 0.822 | 0.025 | 32.416 | 0 | 32.88 | 0 |
| LV_SYCOD.BY | SYCOD_CH | 0.892 | 0.018 | 50.345 | 0 | 49.55556 | 0 |
| LV_CPZ.BY | CPZ_CH | 0.968 | 0.007 | 136.777 | 0 | 138.2857 | 0 |
| LV_SYCOD.ON | LV_HI | 0.093 | 0.102 | 0.915 | 0.36 | 0.911765 | 0.361893 |
| LV_SYCOD.ON | LV_CPZ | -0.016 | 0.067 | -0.239 | 0.811 | -0.23881 | 0.811256 |
| LV_SYCOD.ON | AGE | 0.002 | 0.061 | 0.036 | 0.971 | 0.032787 | 0.973845 |
| LV_SYCOD.ON | FEMALE | -0.03 | 0.064 | -0.472 | 0.637 | -0.46875 | 0.639248 |
| LV_SYCOD.ON | SITE02 | -0.028 | 0.089 | -0.315 | 0.753 | -0.31461 | 0.75306 |
| LV_SYCOD.ON | SITE03 | 0.085 | 0.097 | 0.882 | 0.378 | 0.876289 | 0.380873 |
| LV_SYCOD.ON | SITE04 | -0.157 | 0.066 | -2.388 | 0.017 | -2.37879 | 0.01737 |
| LV_SYCOD.ON | SITE05 | 0.126 | 0.086 | 1.469 | 0.142 | 1.465116 | 0.142889 |
| LV_SYCOD.ON | SITE06 | -0.092 | 0.075 | -1.23 | 0.219 | -1.22667 | 0.219948 |
| LV_SYCOD.ON | SITE07 | -0.061 | 0.075 | -0.811 | 0.417 | -0.81333 | 0.416027 |
| LV_SYCOD.ON | SITE08 | -0.159 | 0.079 | -2.011 | 0.044 | -2.01266 | 0.044151 |
| LV_SYCOD.ON | SITE09 | -0.011 | 0.094 | -0.115 | 0.908 | -0.11702 | 0.906843 |
| LV_HI.WITH | AGE | 0.043 | 0.061 | 0.706 | 0.48 | 0.704918 | 0.480861 |
| LV_HI.WITH | FEMALE | -0.101 | 0.066 | -1.534 | 0.125 | -1.5303 | 0.125942 |
| LV_HI.WITH | SITE02 | -0.007 | 0.068 | -0.103 | 0.918 | -0.10294 | 0.91801 |
| LV_HI.WITH | SITE03 | 0.036 | 0.099 | 0.362 | 0.717 | 0.363636 | 0.71613 |
| LV_HI.WITH | SITE04 | 0.052 | 0.051 | 1.022 | 0.307 | 1.019608 | 0.307914 |
| LV_HI.WITH | SITE05 | -0.086 | 0.07 | -1.235 | 0.217 | -1.22857 | 0.219233 |
| LV_HI.WITH | SITE06 | 0.095 | 0.048 | 1.972 | 0.049 | 1.979167 | 0.047797 |
| LV_HI.WITH | SITE07 | 0.021 | 0.071 | 0.296 | 0.767 | 0.295775 | 0.767402 |
| LV_HI.WITH | SITE08 | -0.039 | 0.054 | -0.724 | 0.469 | -0.72222 | 0.470158 |
| LV_HI.WITH | SITE09 | -0.035 | 0.065 | -0.541 | 0.588 | -0.53846 | 0.590258 |
| LV_HI.WITH | LV_CPZ | 0.038 | 0.1 | 0.381 | 0.703 | 0.38 | 0.703945 |
| AGE.WITH | LV_CPZ | -0.069 | 0.059 | -1.167 | 0.243 | -1.16949 | 0.242206 |
| FEMALE.WITH | LV_CPZ | -0.007 | 0.053 | -0.139 | 0.889 | -0.13208 | 0.894925 |
| SITE02.WITH | LV_CPZ | -0.062 | 0.044 | -1.416 | 0.157 | -1.40909 | 0.158808 |
| SITE03.WITH | LV_CPZ | 0.016 | 0.05 | 0.316 | 0.752 | 0.32 | 0.748968 |
| SITE04.WITH | LV_CPZ | 0.052 | 0.049 | 1.056 | 0.291 | 1.061224 | 0.288588 |
| SITE05.WITH | LV_CPZ | 0.024 | 0.04 | 0.592 | 0.554 | 0.6 | 0.548506 |
| SITE06.WITH | LV_CPZ | -0.008 | 0.078 | -0.106 | 0.915 | -0.10256 | 0.918309 |
| SITE07.WITH | LV_CPZ | -0.015 | 0.052 | -0.278 | 0.781 | -0.28846 | 0.772993 |
| SITE08.WITH | LV_CPZ | 0.131 | 0.062 | 2.101 | 0.036 | 2.112903 | 0.034609 |
| SITE09.WITH | LV_CPZ | -0.024 | 0.042 | -0.563 | 0.573 | -0.57143 | 0.567709 |
| AGE.WITH | FEMALE | -0.148 | 0.052 | -2.856 | 0.004 | -2.84615 | 0.004425 |
| AGE.WITH | SITE02 | 0.326 | 0.06 | 5.449 | 0 | 5.433333 | 5.53E-08 |
| AGE.WITH | SITE03 | -0.026 | 0.046 | -0.574 | 0.566 | -0.56522 | 0.571926 |
| AGE.WITH | SITE04 | 0.021 | 0.045 | 0.462 | 0.644 | 0.466667 | 0.640738 |
| AGE.WITH | SITE05 | 0.162 | 0.049 | 3.33 | 0.001 | 3.306122 | 0.000946 |
| AGE.WITH | SITE06 | -0.001 | 0.047 | -0.015 | 0.988 | -0.02128 | 0.983025 |
| AGE.WITH | SITE07 | -0.155 | 0.045 | -3.401 | 0.001 | -3.44444 | 0.000572 |
| AGE.WITH | SITE08 | -0.194 | 0.05 | -3.89 | 0 | -3.88 | 0.000104 |
| AGE.WITH | SITE09 | -0.027 | 0.048 | -0.565 | 0.572 | -0.5625 | 0.573775 |
| FEMALE.WITH | SITE02 | 0.04 | 0.054 | 0.751 | 0.453 | 0.740741 | 0.458851 |
| FEMALE.WITH | SITE03 | -0.025 | 0.053 | -0.473 | 0.636 | -0.4717 | 0.637142 |
| FEMALE.WITH | SITE04 | -0.039 | 0.052 | -0.755 | 0.45 | -0.75 | 0.453255 |
| FEMALE.WITH | SITE05 | 0.015 | 0.053 | 0.287 | 0.774 | 0.283019 | 0.777162 |
| FEMALE.WITH | SITE06 | -0.073 | 0.052 | -1.411 | 0.158 | -1.40385 | 0.160365 |
| FEMALE.WITH | SITE07 | 0.081 | 0.053 | 1.521 | 0.128 | 1.528302 | 0.126438 |
| FEMALE.WITH | SITE08 | 0.008 | 0.053 | 0.148 | 0.883 | 0.150943 | 0.88002 |
| FEMALE.WITH | SITE09 | 0.034 | 0.053 | 0.634 | 0.526 | 0.641509 | 0.521192 |
| SITE02.WITH | SITE03 | -0.073 | 0.011 | -6.899 | 0 | -6.63636 | 3.22E-11 |
| SITE02.WITH | SITE04 | -0.093 | 0.012 | -7.775 | 0 | -7.75 | 9.1E-15 |
| SITE02.WITH | SITE05 | -0.12 | 0.014 | -8.599 | 0 | -8.57143 | 0 |
| SITE02.WITH | SITE06 | -0.114 | 0.014 | -8.454 | 0 | -8.14286 | 4.44E-16 |
| SITE02.WITH | SITE07 | -0.12 | 0.014 | -8.599 | 0 | -8.57143 | 0 |
| SITE02.WITH | SITE08 | -0.105 | 0.013 | -8.202 | 0 | -8.07692 | 6.66E-16 |
| SITE02.WITH | SITE09 | -0.126 | 0.014 | -8.728 | 0 | -9 | 0 |
| SITE03.WITH | SITE04 | -0.076 | 0.011 | -6.994 | 0 | -6.90909 | 4.88E-12 |
| SITE03.WITH | SITE05 | -0.098 | 0.013 | -7.574 | 0 | -7.53846 | 4.75E-14 |
| SITE03.WITH | SITE06 | -0.094 | 0.013 | -7.475 | 0 | -7.23077 | 4.8E-13 |
| SITE03.WITH | SITE07 | -0.098 | 0.013 | -7.574 | 0 | -7.53846 | 4.75E-14 |
| SITE03.WITH | SITE08 | -0.086 | 0.012 | -7.299 | 0 | -7.16667 | 7.68E-13 |
| SITE03.WITH | SITE09 | -0.103 | 0.013 | -7.662 | 0 | -7.92308 | 2.22E-15 |
| SITE04.WITH | SITE05 | -0.125 | 0.014 | -8.783 | 0 | -8.92857 | 0 |
| SITE04.WITH | SITE06 | -0.119 | 0.014 | -8.63 | 0 | -8.5 | 0 |
| SITE04.WITH | SITE07 | -0.125 | 0.014 | -8.783 | 0 | -8.92857 | 0 |
| SITE04.WITH | SITE08 | -0.109 | 0.013 | -8.362 | 0 | -8.38462 | 0 |
| SITE04.WITH | SITE09 | -0.13 | 0.015 | -8.921 | 0 | -8.66667 | 0 |
| SITE05.WITH | SITE06 | -0.153 | 0.016 | -9.803 | 0 | -9.5625 | 0 |
| SITE05.WITH | SITE07 | -0.161 | 0.016 | -10.031 | 0 | -10.0625 | 0 |
| SITE05.WITH | SITE08 | -0.141 | 0.015 | -9.415 | 0 | -9.4 | 0 |
| SITE05.WITH | SITE09 | -0.169 | 0.016 | -10.239 | 0 | -10.5625 | 0 |
| SITE06.WITH | SITE07 | -0.153 | 0.016 | -9.803 | 0 | -9.5625 | 0 |
| SITE06.WITH | SITE08 | -0.135 | 0.015 | -9.226 | 0 | -9 | 0 |
| SITE06.WITH | SITE09 | -0.161 | 0.016 | -9.996 | 0 | -10.0625 | 0 |
| SITE07.WITH | SITE08 | -0.141 | 0.015 | -9.415 | 0 | -9.4 | 0 |
| SITE07.WITH | SITE09 | -0.169 | 0.016 | -10.239 | 0 | -10.5625 | 0 |
| SITE08.WITH | SITE09 | -0.148 | 0.015 | -9.586 | 0 | -9.86667 | 0 |
| Means | AGE | 4.866 | 0.174 | 27.895 | 0 | 27.96552 | 0 |
| Means | FEMALE | 0.905 | 0.048 | 18.696 | 0 | 18.85417 | 0 |
| Means | SITE02 | 0.299 | 0.029 | 10.318 | 0 | 10.31034 | 0 |
| Means | SITE03 | 0.245 | 0.028 | 8.687 | 0 | 8.75 | 0 |
| Means | SITE04 | 0.31 | 0.029 | 10.635 | 0 | 10.68966 | 0 |
| Means | SITE05 | 0.401 | 0.031 | 12.992 | 0 | 12.93548 | 0 |
| Means | SITE06 | 0.382 | 0.031 | 12.532 | 0 | 12.32258 | 0 |
| Means | SITE07 | 0.401 | 0.031 | 12.992 | 0 | 12.93548 | 0 |
| Means | SITE08 | 0.352 | 0.03 | 11.78 | 0 | 11.73333 | 0 |
| Means | SITE09 | 0.42 | 0.031 | 13.423 | 0 | 13.54839 | 0 |
| Intercepts | SYCOD_CH | 0.811 | 0.314 | 2.583 | 0.01 | 2.582803 | 0.0098 |
| Intercepts | HI_CH | -0.053 | 0.053 | -1 | 0.317 | -1 | 0.317311 |
| Intercepts | CPZ_CH | 0.087 | 0.052 | 1.664 | 0.096 | 1.673077 | 0.094312 |
| Variances | AGE | 1 | 0 |  |  |  |  |
| Variances | FEMALE | 1 | 0 |  |  |  |  |
| Variances | SITE02 | 1 | 0 |  |  |  |  |
| Variances | SITE03 | 1 | 0 |  |  |  |  |
| Variances | SITE04 | 1 | 0 |  |  |  |  |
| Variances | SITE05 | 1 | 0 |  |  |  |  |
| Variances | SITE06 | 1 | 0 |  |  |  |  |
| Variances | SITE07 | 1 | 0 |  |  |  |  |
| Variances | SITE08 | 1 | 0 |  |  |  |  |
| Variances | SITE09 | 1 | 0 |  |  |  |  |
| Variances | LV_HI | 1 | 0 |  |  |  |  |
| Variances | LV_CPZ | 1 | 0 |  |  |  |  |
| Residual.Variances | SYCOD_CH | 0.204 | 0.032 | 6.445 | 0 | 6.375 | 1.83E-10 |
| Residual.Variances | HI_CH | 0.325 | 0.042 | 7.803 | 0 | 7.738095 | 9.99E-15 |
| Residual.Variances | CPZ_CH | 0.064 | 0.014 | 4.659 | 0 | 4.571429 | 4.84E-06 |
| Residual.Variances | LV_SYCOD | 0.907 | 0.03 | 30.113 | 0 | 30.23333 | 0 |
| LV_HI.BY | HI_CH | 0.823 | 0.025 | 32.653 | 0 | 32.92 | 0 |
| LV_VEMEM.BY | VEMEM_CH | 0.746 | 0.044 | 16.91 | 0 | 16.95455 | 0 |
| LV_CPZ.BY | CPZ_CH | 0.968 | 0.007 | 136.747 | 0 | 138.2857 | 0 |
| LV_VEMEM.ON | LV_HI | -0.155 | 0.093 | -1.667 | 0.096 | -1.66667 | 0.095581 |
| LV_VEMEM.ON | LV_CPZ | 0.065 | 0.07 | 0.937 | 0.349 | 0.928571 | 0.353111 |
| LV_VEMEM.ON | AGE | 0.071 | 0.072 | 0.984 | 0.325 | 0.986111 | 0.324079 |
| LV_VEMEM.ON | FEMALE | -0.134 | 0.07 | -1.933 | 0.053 | -1.91429 | 0.055584 |
| LV_VEMEM.ON | SITE02 | 0.005 | 0.096 | 0.048 | 0.961 | 0.052083 | 0.958462 |
| LV_VEMEM.ON | SITE03 | -0.007 | 0.076 | -0.095 | 0.924 | -0.09211 | 0.926614 |
| LV_VEMEM.ON | SITE04 | -0.2 | 0.104 | -1.933 | 0.053 | -1.92308 | 0.05447 |
| LV_VEMEM.ON | SITE05 | 0.045 | 0.084 | 0.535 | 0.592 | 0.535714 | 0.592156 |
| LV_VEMEM.ON | SITE06 | 0.115 | 0.094 | 1.222 | 0.222 | 1.223404 | 0.221177 |
| LV_VEMEM.ON | SITE07 | 0.082 | 0.08 | 1.031 | 0.303 | 1.025 | 0.305363 |
| LV_VEMEM.ON | SITE08 | 0.075 | 0.094 | 0.793 | 0.428 | 0.797872 | 0.424945 |
| LV_VEMEM.ON | SITE09 | 0.149 | 0.099 | 1.494 | 0.135 | 1.505051 | 0.132311 |
| LV_HI.WITH | AGE | 0.044 | 0.061 | 0.72 | 0.472 | 0.721311 | 0.470718 |
| LV_HI.WITH | FEMALE | -0.099 | 0.066 | -1.51 | 0.131 | -1.5 | 0.133614 |
| LV_HI.WITH | SITE02 | -0.006 | 0.068 | -0.095 | 0.924 | -0.08824 | 0.92969 |
| LV_HI.WITH | SITE03 | 0.036 | 0.099 | 0.367 | 0.714 | 0.363636 | 0.71613 |
| LV_HI.WITH | SITE04 | 0.052 | 0.051 | 1.032 | 0.302 | 1.019608 | 0.307914 |
| LV_HI.WITH | SITE05 | -0.085 | 0.069 | -1.224 | 0.221 | -1.23188 | 0.217992 |
| LV_HI.WITH | SITE06 | 0.096 | 0.048 | 1.986 | 0.047 | 2 | 0.0455 |
| LV_HI.WITH | SITE07 | 0.022 | 0.071 | 0.307 | 0.759 | 0.309859 | 0.756668 |
| LV_HI.WITH | SITE08 | -0.038 | 0.054 | -0.712 | 0.477 | -0.7037 | 0.481617 |
| LV_HI.WITH | SITE09 | -0.04 | 0.065 | -0.61 | 0.542 | -0.61538 | 0.538301 |
| LV_HI.WITH | LV_CPZ | 0.039 | 0.1 | 0.387 | 0.699 | 0.39 | 0.696537 |
| AGE.WITH | LV_CPZ | -0.069 | 0.059 | -1.167 | 0.243 | -1.16949 | 0.242206 |
| FEMALE.WITH | LV_CPZ | -0.008 | 0.053 | -0.15 | 0.881 | -0.15094 | 0.88002 |
| SITE02.WITH | LV_CPZ | -0.062 | 0.044 | -1.415 | 0.157 | -1.40909 | 0.158808 |
| SITE03.WITH | LV_CPZ | 0.016 | 0.05 | 0.316 | 0.752 | 0.32 | 0.748968 |
| SITE04.WITH | LV_CPZ | 0.052 | 0.049 | 1.057 | 0.291 | 1.061224 | 0.288588 |
| SITE05.WITH | LV_CPZ | 0.024 | 0.04 | 0.592 | 0.554 | 0.6 | 0.548506 |
| SITE06.WITH | LV_CPZ | -0.008 | 0.078 | -0.106 | 0.916 | -0.10256 | 0.918309 |
| SITE07.WITH | LV_CPZ | -0.015 | 0.052 | -0.278 | 0.781 | -0.28846 | 0.772993 |
| SITE08.WITH | LV_CPZ | 0.131 | 0.062 | 2.102 | 0.036 | 2.112903 | 0.034609 |
| SITE09.WITH | LV_CPZ | -0.023 | 0.042 | -0.547 | 0.584 | -0.54762 | 0.583954 |
| AGE.WITH | FEMALE | -0.15 | 0.052 | -2.879 | 0.004 | -2.88462 | 0.003919 |
| AGE.WITH | SITE02 | 0.326 | 0.06 | 5.443 | 0 | 5.433333 | 5.53E-08 |
| AGE.WITH | SITE03 | -0.027 | 0.046 | -0.581 | 0.561 | -0.58696 | 0.557233 |
| AGE.WITH | SITE04 | 0.02 | 0.045 | 0.453 | 0.651 | 0.444444 | 0.656721 |
| AGE.WITH | SITE05 | 0.161 | 0.049 | 3.319 | 0.001 | 3.285714 | 0.001017 |
| AGE.WITH | SITE06 | -0.001 | 0.047 | -0.026 | 0.979 | -0.02128 | 0.983025 |
| AGE.WITH | SITE07 | -0.155 | 0.045 | -3.413 | 0.001 | -3.44444 | 0.000572 |
| AGE.WITH | SITE08 | -0.195 | 0.05 | -3.899 | 0 | -3.9 | 9.62E-05 |
| AGE.WITH | SITE09 | -0.024 | 0.049 | -0.502 | 0.616 | -0.4898 | 0.624278 |
| FEMALE.WITH | SITE02 | 0.039 | 0.054 | 0.736 | 0.462 | 0.722222 | 0.470158 |
| FEMALE.WITH | SITE03 | -0.025 | 0.053 | -0.485 | 0.628 | -0.4717 | 0.637142 |
| FEMALE.WITH | SITE04 | -0.04 | 0.052 | -0.77 | 0.441 | -0.76923 | 0.441756 |
| FEMALE.WITH | SITE05 | 0.014 | 0.053 | 0.267 | 0.789 | 0.264151 | 0.791664 |
| FEMALE.WITH | SITE06 | -0.074 | 0.052 | -1.43 | 0.153 | -1.42308 | 0.154714 |
| FEMALE.WITH | SITE07 | 0.08 | 0.053 | 1.502 | 0.133 | 1.509434 | 0.131188 |
| FEMALE.WITH | SITE08 | 0.007 | 0.053 | 0.131 | 0.896 | 0.132075 | 0.894925 |
| FEMALE.WITH | SITE09 | 0.04 | 0.054 | 0.755 | 0.45 | 0.740741 | 0.458851 |
| SITE02.WITH | SITE03 | -0.074 | 0.011 | -6.9 | 0 | -6.72727 | 1.73E-11 |
| SITE02.WITH | SITE04 | -0.093 | 0.012 | -7.775 | 0 | -7.75 | 9.1E-15 |
| SITE02.WITH | SITE05 | -0.12 | 0.014 | -8.599 | 0 | -8.57143 | 0 |
| SITE02.WITH | SITE06 | -0.115 | 0.014 | -8.455 | 0 | -8.21429 | 2.22E-16 |
| SITE02.WITH | SITE07 | -0.12 | 0.014 | -8.599 | 0 | -8.57143 | 0 |
| SITE02.WITH | SITE08 | -0.106 | 0.013 | -8.203 | 0 | -8.15385 | 4.44E-16 |
| SITE02.WITH | SITE09 | -0.125 | 0.014 | -8.697 | 0 | -8.92857 | 0 |
| SITE03.WITH | SITE04 | -0.076 | 0.011 | -6.994 | 0 | -6.90909 | 4.88E-12 |
| SITE03.WITH | SITE05 | -0.099 | 0.013 | -7.574 | 0 | -7.61538 | 2.62E-14 |
| SITE03.WITH | SITE06 | -0.094 | 0.013 | -7.475 | 0 | -7.23077 | 4.8E-13 |
| SITE03.WITH | SITE07 | -0.099 | 0.013 | -7.574 | 0 | -7.61538 | 2.62E-14 |
| SITE03.WITH | SITE08 | -0.087 | 0.012 | -7.299 | 0 | -7.25 | 4.17E-13 |
| SITE03.WITH | SITE09 | -0.102 | 0.013 | -7.641 | 0 | -7.84615 | 4.22E-15 |
| SITE04.WITH | SITE05 | -0.125 | 0.014 | -8.784 | 0 | -8.92857 | 0 |
| SITE04.WITH | SITE06 | -0.119 | 0.014 | -8.63 | 0 | -8.5 | 0 |
| SITE04.WITH | SITE07 | -0.125 | 0.014 | -8.784 | 0 | -8.92857 | 0 |
| SITE04.WITH | SITE08 | -0.11 | 0.013 | -8.362 | 0 | -8.46154 | 0 |
| SITE04.WITH | SITE09 | -0.129 | 0.015 | -8.889 | 0 | -8.6 | 0 |
| SITE05.WITH | SITE06 | -0.154 | 0.016 | -9.803 | 0 | -9.625 | 0 |
| SITE05.WITH | SITE07 | -0.162 | 0.016 | -10.032 | 0 | -10.125 | 0 |
| SITE05.WITH | SITE08 | -0.142 | 0.015 | -9.415 | 0 | -9.46667 | 0 |
| SITE05.WITH | SITE09 | -0.167 | 0.016 | -10.189 | 0 | -10.4375 | 0 |
| SITE06.WITH | SITE07 | -0.154 | 0.016 | -9.803 | 0 | -9.625 | 0 |
| SITE06.WITH | SITE08 | -0.135 | 0.015 | -9.226 | 0 | -9 | 0 |
| SITE06.WITH | SITE09 | -0.159 | 0.016 | -9.95 | 0 | -9.9375 | 0 |
| SITE07.WITH | SITE08 | -0.142 | 0.015 | -9.415 | 0 | -9.46667 | 0 |
| SITE07.WITH | SITE09 | -0.167 | 0.016 | -10.189 | 0 | -10.4375 | 0 |
| SITE08.WITH | SITE09 | -0.147 | 0.015 | -9.545 | 0 | -9.8 | 0 |
| Means | AGE | 4.862 | 0.174 | 27.902 | 0 | 27.94253 | 0 |
| Means | FEMALE | 0.908 | 0.049 | 18.674 | 0 | 18.53061 | 0 |
| Means | SITE02 | 0.3 | 0.029 | 10.317 | 0 | 10.34483 | 0 |
| Means | SITE03 | 0.245 | 0.028 | 8.686 | 0 | 8.75 | 0 |
| Means | SITE04 | 0.311 | 0.029 | 10.634 | 0 | 10.72414 | 0 |
| Means | SITE05 | 0.402 | 0.031 | 12.989 | 0 | 12.96774 | 0 |
| Means | SITE06 | 0.383 | 0.031 | 12.53 | 0 | 12.35484 | 0 |
| Means | SITE07 | 0.402 | 0.031 | 12.989 | 0 | 12.96774 | 0 |
| Means | SITE08 | 0.353 | 0.03 | 11.778 | 0 | 11.76667 | 0 |
| Means | SITE09 | 0.416 | 0.031 | 13.314 | 0 | 13.41935 | 0 |
| Intercepts | VEMEM_CH | 0.04 | 0.321 | 0.123 | 0.902 | 0.124611 | 0.900832 |
| Intercepts | HI_CH | -0.054 | 0.053 | -1.026 | 0.305 | -1.01887 | 0.308266 |
| Intercepts | CPZ_CH | 0.086 | 0.052 | 1.658 | 0.097 | 1.653846 | 0.098159 |
| Variances | AGE | 1 | 0 |  |  |  |  |
| Variances | FEMALE | 1 | 0 |  |  |  |  |
| Variances | SITE02 | 1 | 0 |  |  |  |  |
| Variances | SITE03 | 1 | 0 |  |  |  |  |
| Variances | SITE04 | 1 | 0 |  |  |  |  |
| Variances | SITE05 | 1 | 0 |  |  |  |  |
| Variances | SITE06 | 1 | 0 |  |  |  |  |
| Variances | SITE07 | 1 | 0 |  |  |  |  |
| Variances | SITE08 | 1 | 0 |  |  |  |  |
| Variances | SITE09 | 1 | 0 |  |  |  |  |
| Variances | LV_HI | 1 | 0 |  |  |  |  |
| Variances | LV_CPZ | 1 | 0 |  |  |  |  |
| Residual.Variances | VEMEM_CH | 0.443 | 0.066 | 6.725 | 0 | 6.712121 | 1.92E-11 |
| Residual.Variances | HI_CH | 0.323 | 0.041 | 7.8 | 0 | 7.878049 | 3.33E-15 |
| Residual.Variances | CPZ_CH | 0.064 | 0.014 | 4.658 | 0 | 4.571429 | 4.84E-06 |
| Residual.Variances | LV_VEMEM | 0.87 | 0.053 | 16.562 | 0 | 16.41509 | 0 |

Table_S5b_SensitivityAnalysisStandardized_Med_Corr

| **paramHeader** | **param** | **est** | **se** | **est_se** | **pval** | **zval** | **p_exact** | **p_bonf** |
| --- | --- | --- | --- | --- | --- | --- | --- | --- |
| LV_DEPS.ON | LV_HI | -0.341 | 0.105 | -3.243 | 0.001 | -3.24762 | 0.001164 | 0.006982 |
| LV_GAF.ON | LV_HI | 0.352 | 0.103 | 3.428 | 0.001 | 3.417476 | 0.000632 | 0.003792 |
| LV_NEGS.ON | LV_HI | -0.363 | 0.082 | -4.404 | 0 | -4.42683 | 9.56E-06 | 5.74E-05 |
| LV_POSS.ON | LV_HI | -0.174 | 0.115 | -1.521 | 0.128 | -1.51304 | 0.130269 | 0.781612 |
| LV_SYCOD.ON | LV_HI | 0.093 | 0.102 | 0.915 | 0.36 | 0.911765 | 0.361893 | 1 |
| LV_VEMEM.ON | LV_HI | -0.155 | 0.093 | -1.667 | 0.096 | -1.66667 | 0.095581 | 0.573484 |

Table_S5c_SensitivityAnalysis_Medication_ModelSum

| **behav** | **neuro** | **estimator** | **n_obs** | **n_parameters** | **aic** | **bic** |
| --- | --- | --- | --- | --- | --- | --- |
| PosS | fchi | MLR | 357 | 104 | 5671.17 | 6074.454 |
| NegS | fchi | MLR | 356 | 104 | 5888.308 | 6291.301 |
| DepS | fchi | MLR | 357 | 104 | 5811.37 | 6214.655 |
| gaf | fchi | MLR | 356 | 104 | 6460.786 | 6863.779 |
| SyCod | fchi | MLR | 353 | 104 | 6355.813 | 6757.926 |
| VeMem | fchi | MLR | 352 | 104 | 5674.989 | 6076.807 |

Table_S6a_PredAnalysisStand_FCtoPredictClin

| **paramHeader** | **param** | **est** | **se** | **est_se** | **pval** | **zval** | **pval_exact** |
| --- | --- | --- | --- | --- | --- | --- | --- |
| LV_DEPS.BY | DEPS_CH | 0.849 | 0.035 | 24.128 | 0 | 24.25714 | 0 |
| LV_DEPS.ON | HI_CH | 0.062 | 0.066 | 0.942 | 0.346 | 0.939394 | 0.347528524 |
| LV_DEPS.ON | AGE | -0.134 | 0.09 | -1.496 | 0.135 | -1.48889 | 0.13651663 |
| LV_DEPS.ON | FEMALE | 0.025 | 0.074 | 0.333 | 0.739 | 0.337838 | 0.735485394 |
| LV_DEPS.ON | SITE02 | 0.303 | 0.119 | 2.535 | 0.011 | 2.546218 | 0.010889701 |
| LV_DEPS.ON | SITE03 | 0.275 | 0.117 | 2.351 | 0.019 | 2.350427 | 0.018751868 |
| LV_DEPS.ON | SITE04 | 0.355 | 0.135 | 2.627 | 0.009 | 2.62963 | 0.008547794 |
| LV_DEPS.ON | SITE05 | 0.451 | 0.161 | 2.812 | 0.005 | 2.801242 | 0.005090629 |
| LV_DEPS.ON | SITE06 | 0.339 | 0.176 | 1.922 | 0.055 | 1.926136 | 0.054087349 |
| LV_DEPS.ON | SITE07 | 0.331 | 0.178 | 1.859 | 0.063 | 1.859551 | 0.062949139 |
| LV_DEPS.ON | SITE08 | 0.254 | 0.178 | 1.423 | 0.155 | 1.426966 | 0.153589611 |
| LV_DEPS.ON | SITE09 | 0.442 | 0.169 | 2.612 | 0.009 | 2.615385 | 0.008912703 |
| HI_CH.WITH | AGE | 0.02 | 0.067 | 0.294 | 0.769 | 0.298507 | 0.765315881 |
| HI_CH.WITH | FEMALE | -0.005 | 0.063 | -0.075 | 0.94 | -0.07937 | 0.936742244 |
| HI_CH.WITH | SITE02 | -0.021 | 0.056 | -0.375 | 0.708 | -0.375 | 0.707660467 |
| HI_CH.WITH | SITE03 | -0.036 | 0.106 | -0.336 | 0.737 | -0.33962 | 0.734140725 |
| HI_CH.WITH | SITE04 | 0.024 | 0.046 | 0.512 | 0.609 | 0.521739 | 0.601851976 |
| HI_CH.WITH | SITE05 | -0.004 | 0.064 | -0.059 | 0.953 | -0.0625 | 0.950164662 |
| HI_CH.WITH | SITE06 | 0.063 | 0.068 | 0.934 | 0.35 | 0.926471 | 0.354201475 |
| HI_CH.WITH | SITE07 | 0.015 | 0.057 | 0.259 | 0.796 | 0.263158 | 0.792428882 |
| HI_CH.WITH | SITE08 | -0.051 | 0.057 | -0.903 | 0.367 | -0.89474 | 0.370927779 |
| HI_CH.WITH | SITE09 | 0.009 | 0.059 | 0.16 | 0.873 | 0.152542 | 0.878759173 |
| AGE.WITH | FEMALE | -0.158 | 0.06 | -2.617 | 0.009 | -2.63333 | 0.008455132 |
| AGE.WITH | SITE02 | 0.262 | 0.083 | 3.175 | 0.002 | 3.156627 | 0.001596056 |
| AGE.WITH | SITE03 | -0.009 | 0.052 | -0.171 | 0.865 | -0.17308 | 0.862590964 |
| AGE.WITH | SITE04 | -0.027 | 0.045 | -0.596 | 0.551 | -0.6 | 0.548506236 |
| AGE.WITH | SITE05 | 0.218 | 0.056 | 3.907 | 0 | 3.892857 | 9.90705E-05 |
| AGE.WITH | SITE06 | 0.03 | 0.058 | 0.513 | 0.608 | 0.517241 | 0.60498767 |
| AGE.WITH | SITE07 | -0.156 | 0.055 | -2.814 | 0.005 | -2.83636 | 0.004563046 |
| AGE.WITH | SITE08 | -0.184 | 0.063 | -2.909 | 0.004 | -2.92063 | 0.003493189 |
| AGE.WITH | SITE09 | -0.008 | 0.06 | -0.13 | 0.896 | -0.13333 | 0.893929767 |
| FEMALE.WITH | SITE02 | 0.07 | 0.062 | 1.133 | 0.257 | 1.129032 | 0.258884226 |
| FEMALE.WITH | SITE03 | -0.002 | 0.062 | -0.033 | 0.973 | -0.03226 | 0.974266251 |
| FEMALE.WITH | SITE04 | -0.031 | 0.061 | -0.501 | 0.616 | -0.5082 | 0.61131539 |
| FEMALE.WITH | SITE05 | 0.002 | 0.062 | 0.028 | 0.977 | 0.032258 | 0.974266251 |
| FEMALE.WITH | SITE06 | -0.124 | 0.059 | -2.095 | 0.036 | -2.10169 | 0.035580009 |
| FEMALE.WITH | SITE07 | 0.056 | 0.062 | 0.902 | 0.367 | 0.903226 | 0.366406064 |
| FEMALE.WITH | SITE08 | 0.031 | 0.062 | 0.49 | 0.624 | 0.5 | 0.617075077 |
| FEMALE.WITH | SITE09 | 0.036 | 0.062 | 0.574 | 0.566 | 0.580645 | 0.561479628 |
| SITE02.WITH | SITE03 | -0.052 | 0.01 | -5.045 | 0 | -5.2 | 1.99289E-07 |
| SITE02.WITH | SITE04 | -0.076 | 0.013 | -5.99 | 0 | -5.84615 | 5.03069E-09 |
| SITE02.WITH | SITE05 | -0.104 | 0.016 | -6.619 | 0 | -6.5 | 8.032E-11 |
| SITE02.WITH | SITE06 | -0.108 | 0.016 | -6.667 | 0 | -6.75 | 1.47846E-11 |
| SITE02.WITH | SITE07 | -0.106 | 0.016 | -6.643 | 0 | -6.625 | 3.47249E-11 |
| SITE02.WITH | SITE08 | -0.093 | 0.015 | -6.419 | 0 | -6.2 | 5.64632E-10 |
| SITE02.WITH | SITE09 | -0.109 | 0.016 | -6.689 | 0 | -6.8125 | 9.59166E-12 |
| SITE03.WITH | SITE04 | -0.064 | 0.012 | -5.427 | 0 | -5.33333 | 9.64261E-08 |
| SITE03.WITH | SITE05 | -0.089 | 0.015 | -5.881 | 0 | -5.93333 | 2.96845E-09 |
| SITE03.WITH | SITE06 | -0.091 | 0.015 | -5.915 | 0 | -6.06667 | 1.30592E-09 |
| SITE03.WITH | SITE07 | -0.09 | 0.015 | -5.898 | 0 | -6 | 1.97318E-09 |
| SITE03.WITH | SITE08 | -0.079 | 0.014 | -5.74 | 0 | -5.64286 | 1.67251E-08 |
| SITE03.WITH | SITE09 | -0.093 | 0.016 | -5.93 | 0 | -5.8125 | 6.15467E-09 |
| SITE04.WITH | SITE05 | -0.128 | 0.017 | -7.563 | 0 | -7.52941 | 5.10703E-14 |
| SITE04.WITH | SITE06 | -0.132 | 0.017 | -7.635 | 0 | -7.76471 | 8.21565E-15 |
| SITE04.WITH | SITE07 | -0.13 | 0.017 | -7.6 | 0 | -7.64706 | 2.06501E-14 |
| SITE04.WITH | SITE08 | -0.114 | 0.016 | -7.269 | 0 | -7.125 | 1.04072E-12 |
| SITE04.WITH | SITE09 | -0.134 | 0.017 | -7.669 | 0 | -7.88235 | 3.10862E-15 |
| SITE05.WITH | SITE06 | -0.183 | 0.02 | -9.094 | 0 | -9.15 | 0 |
| SITE05.WITH | SITE07 | -0.18 | 0.02 | -9.035 | 0 | -9 | 0 |
| SITE05.WITH | SITE08 | -0.158 | 0.019 | -8.492 | 0 | -8.31579 | 0 |
| SITE05.WITH | SITE09 | -0.185 | 0.02 | -9.152 | 0 | -9.25 | 0 |
| SITE06.WITH | SITE07 | -0.185 | 0.02 | -9.158 | 0 | -9.25 | 0 |
| SITE06.WITH | SITE08 | -0.163 | 0.019 | -8.594 | 0 | -8.57895 | 0 |
| SITE06.WITH | SITE09 | -0.191 | 0.021 | -9.28 | 0 | -9.09524 | 0 |
| SITE07.WITH | SITE08 | -0.16 | 0.019 | -8.543 | 0 | -8.42105 | 0 |
| SITE07.WITH | SITE09 | -0.188 | 0.02 | -9.217 | 0 | -9.4 | 0 |
| SITE08.WITH | SITE09 | -0.165 | 0.019 | -8.642 | 0 | -8.68421 | 0 |
| Means | HI_CH | 0.045 | 0.062 | 0.713 | 0.476 | 0.725806 | 0.467957431 |
| Means | AGE | 4.948 | 0.217 | 22.796 | 0 | 22.80184 | 0 |
| Means | FEMALE | 0.922 | 0.057 | 16.04 | 0 | 16.17544 | 0 |
| Means | SITE02 | 0.248 | 0.033 | 7.518 | 0 | 7.515152 | 5.68434E-14 |
| Means | SITE03 | 0.211 | 0.032 | 6.491 | 0 | 6.59375 | 4.28855E-11 |
| Means | SITE04 | 0.305 | 0.034 | 8.974 | 0 | 8.970588 | 0 |
| Means | SITE05 | 0.421 | 0.037 | 11.511 | 0 | 11.37838 | 0 |
| Means | SITE06 | 0.434 | 0.037 | 11.749 | 0 | 11.72973 | 0 |
| Means | SITE07 | 0.427 | 0.037 | 11.631 | 0 | 11.54054 | 0 |
| Means | SITE08 | 0.375 | 0.035 | 10.592 | 0 | 10.71429 | 0 |
| Means | SITE09 | 0.44 | 0.037 | 11.864 | 0 | 11.89189 | 0 |
| Intercepts | DEPS_CH | -0.484 | 0.587 | -0.824 | 0.41 | -0.82453 | 0.409637616 |
| Variances | HI_CH | 1 | 0 |  |  |  |  |
| Variances | AGE | 1 | 0 |  |  |  |  |
| Variances | FEMALE | 1 | 0 |  |  |  |  |
| Variances | SITE02 | 1 | 0 |  |  |  |  |
| Variances | SITE03 | 1 | 0 |  |  |  |  |
| Variances | SITE04 | 1 | 0 |  |  |  |  |
| Variances | SITE05 | 1 | 0 |  |  |  |  |
| Variances | SITE06 | 1 | 0 |  |  |  |  |
| Variances | SITE07 | 1 | 0 |  |  |  |  |
| Variances | SITE08 | 1 | 0 |  |  |  |  |
| Variances | SITE09 | 1 | 0 |  |  |  |  |
| Residual.Variances | DEPS_CH | 0.279 | 0.06 | 4.669 | 0 | 4.65 | 3.31935E-06 |
| Residual.Variances | LV_DEPS | 0.887 | 0.049 | 18.193 | 0 | 18.10204 | 0 |
| LV_GAF.BY | GAF_CH | 0.91 | 0.018 | 50.533 | 0 | 50.55556 | 0 |
| LV_GAF.ON | HI_CH | -0.071 | 0.06 | -1.171 | 0.242 | -1.18333 | 0.236677067 |
| LV_GAF.ON | AGE | -0.054 | 0.066 | -0.82 | 0.412 | -0.81818 | 0.413253375 |
| LV_GAF.ON | FEMALE | -0.103 | 0.064 | -1.605 | 0.108 | -1.60938 | 0.107534368 |
| LV_GAF.ON | SITE02 | 0.052 | 0.135 | 0.384 | 0.701 | 0.385185 | 0.700100219 |
| LV_GAF.ON | SITE03 | -0.025 | 0.117 | -0.216 | 0.829 | -0.21368 | 0.830800342 |
| LV_GAF.ON | SITE04 | 0.028 | 0.16 | 0.177 | 0.859 | 0.175 | 0.861079634 |
| LV_GAF.ON | SITE05 | 0.036 | 0.198 | 0.182 | 0.856 | 0.181818 | 0.855725415 |
| LV_GAF.ON | SITE06 | -0.012 | 0.204 | -0.058 | 0.954 | -0.05882 | 0.953092667 |
| LV_GAF.ON | SITE07 | 0.057 | 0.207 | 0.276 | 0.783 | 0.275362 | 0.783037891 |
| LV_GAF.ON | SITE08 | 0.093 | 0.186 | 0.498 | 0.618 | 0.5 | 0.617075077 |
| LV_GAF.ON | SITE09 | -0.047 | 0.208 | -0.226 | 0.822 | -0.22596 | 0.821231335 |
| HI_CH.WITH | AGE | 0.008 | 0.068 | 0.122 | 0.903 | 0.117647 | 0.906347316 |
| HI_CH.WITH | FEMALE | -0.003 | 0.063 | -0.044 | 0.965 | -0.04762 | 0.962019851 |
| HI_CH.WITH | SITE02 | -0.02 | 0.056 | -0.363 | 0.717 | -0.35714 | 0.720984862 |
| HI_CH.WITH | SITE03 | -0.035 | 0.107 | -0.331 | 0.741 | -0.3271 | 0.743590131 |
| HI_CH.WITH | SITE04 | 0.025 | 0.047 | 0.531 | 0.596 | 0.531915 | 0.594784941 |
| HI_CH.WITH | SITE05 | -0.003 | 0.064 | -0.041 | 0.967 | -0.04688 | 0.962612853 |
| HI_CH.WITH | SITE06 | 0.065 | 0.068 | 0.953 | 0.34 | 0.955882 | 0.339131676 |
| HI_CH.WITH | SITE07 | 0.016 | 0.057 | 0.28 | 0.78 | 0.280702 | 0.778939163 |
| HI_CH.WITH | SITE08 | -0.036 | 0.057 | -0.64 | 0.522 | -0.63158 | 0.527662047 |
| HI_CH.WITH | SITE09 | -0.01 | 0.058 | -0.177 | 0.86 | -0.17241 | 0.863112229 |
| AGE.WITH | FEMALE | -0.16 | 0.06 | -2.654 | 0.008 | -2.66667 | 0.007660761 |
| AGE.WITH | SITE02 | 0.263 | 0.083 | 3.165 | 0.002 | 3.168675 | 0.001531357 |
| AGE.WITH | SITE03 | -0.01 | 0.053 | -0.188 | 0.851 | -0.18868 | 0.850344219 |
| AGE.WITH | SITE04 | -0.028 | 0.045 | -0.625 | 0.532 | -0.62222 | 0.533795756 |
| AGE.WITH | SITE05 | 0.217 | 0.056 | 3.877 | 0 | 3.875 | 0.000106625 |
| AGE.WITH | SITE06 | 0.028 | 0.058 | 0.481 | 0.631 | 0.482759 | 0.629267135 |
| AGE.WITH | SITE07 | -0.159 | 0.056 | -2.851 | 0.004 | -2.83929 | 0.004521465 |
| AGE.WITH | SITE08 | -0.173 | 0.064 | -2.711 | 0.007 | -2.70313 | 0.006869091 |
| AGE.WITH | SITE09 | -0.013 | 0.061 | -0.212 | 0.832 | -0.21311 | 0.831237457 |
| FEMALE.WITH | SITE02 | 0.068 | 0.062 | 1.105 | 0.269 | 1.096774 | 0.272740114 |
| FEMALE.WITH | SITE03 | -0.004 | 0.062 | -0.058 | 0.954 | -0.06452 | 0.948559265 |
| FEMALE.WITH | SITE04 | -0.033 | 0.062 | -0.537 | 0.591 | -0.53226 | 0.594547272 |
| FEMALE.WITH | SITE05 | -0.001 | 0.062 | -0.02 | 0.984 | -0.01613 | 0.987131452 |
| FEMALE.WITH | SITE06 | -0.128 | 0.059 | -2.148 | 0.032 | -2.16949 | 0.030045387 |
| FEMALE.WITH | SITE07 | 0.053 | 0.063 | 0.853 | 0.393 | 0.84127 | 0.400196782 |
| FEMALE.WITH | SITE08 | 0.039 | 0.063 | 0.63 | 0.528 | 0.619048 | 0.535884989 |
| FEMALE.WITH | SITE09 | 0.043 | 0.063 | 0.687 | 0.492 | 0.68254 | 0.494897759 |
| SITE02.WITH | SITE03 | -0.053 | 0.01 | -5.045 | 0 | -5.3 | 1.15803E-07 |
| SITE02.WITH | SITE04 | -0.076 | 0.013 | -5.99 | 0 | -5.84615 | 5.03069E-09 |
| SITE02.WITH | SITE05 | -0.105 | 0.016 | -6.62 | 0 | -6.5625 | 5.2913E-11 |
| SITE02.WITH | SITE06 | -0.108 | 0.016 | -6.667 | 0 | -6.75 | 1.47846E-11 |
| SITE02.WITH | SITE07 | -0.107 | 0.016 | -6.644 | 0 | -6.6875 | 2.27016E-11 |
| SITE02.WITH | SITE08 | -0.092 | 0.014 | -6.386 | 0 | -6.57143 | 4.98348E-11 |
| SITE02.WITH | SITE09 | -0.108 | 0.016 | -6.667 | 0 | -6.75 | 1.47846E-11 |
| SITE03.WITH | SITE04 | -0.065 | 0.012 | -5.427 | 0 | -5.41667 | 6.07204E-08 |
| SITE03.WITH | SITE05 | -0.089 | 0.015 | -5.882 | 0 | -5.93333 | 2.96845E-09 |
| SITE03.WITH | SITE06 | -0.092 | 0.016 | -5.915 | 0 | -5.75 | 8.92434E-09 |
| SITE03.WITH | SITE07 | -0.091 | 0.015 | -5.899 | 0 | -6.06667 | 1.30592E-09 |
| SITE03.WITH | SITE08 | -0.078 | 0.014 | -5.716 | 0 | -5.57143 | 2.52659E-08 |
| SITE03.WITH | SITE09 | -0.092 | 0.016 | -5.915 | 0 | -5.75 | 8.92434E-09 |
| SITE04.WITH | SITE05 | -0.129 | 0.017 | -7.564 | 0 | -7.58824 | 3.24185E-14 |
| SITE04.WITH | SITE06 | -0.133 | 0.017 | -7.636 | 0 | -7.82353 | 5.10703E-15 |
| SITE04.WITH | SITE07 | -0.131 | 0.017 | -7.601 | 0 | -7.70588 | 1.31006E-14 |
| SITE04.WITH | SITE08 | -0.113 | 0.016 | -7.221 | 0 | -7.0625 | 1.63536E-12 |
| SITE04.WITH | SITE09 | -0.133 | 0.017 | -7.636 | 0 | -7.82353 | 5.10703E-15 |
| SITE05.WITH | SITE06 | -0.184 | 0.02 | -9.097 | 0 | -9.2 | 0 |
| SITE05.WITH | SITE07 | -0.182 | 0.02 | -9.038 | 0 | -9.1 | 0 |
| SITE05.WITH | SITE08 | -0.157 | 0.019 | -8.416 | 0 | -8.26316 | 2.22045E-16 |
| SITE05.WITH | SITE09 | -0.184 | 0.02 | -9.097 | 0 | -9.2 | 0 |
| SITE06.WITH | SITE07 | -0.187 | 0.02 | -9.161 | 0 | -9.35 | 0 |
| SITE06.WITH | SITE08 | -0.161 | 0.019 | -8.515 | 0 | -8.47368 | 0 |
| SITE06.WITH | SITE09 | -0.19 | 0.021 | -9.223 | 0 | -9.04762 | 0 |
| SITE07.WITH | SITE08 | -0.159 | 0.019 | -8.466 | 0 | -8.36842 | 0 |
| SITE07.WITH | SITE09 | -0.187 | 0.02 | -9.161 | 0 | -9.35 | 0 |
| SITE08.WITH | SITE09 | -0.161 | 0.019 | -8.515 | 0 | -8.47368 | 0 |
| Means | HI_CH | 0.042 | 0.063 | 0.671 | 0.502 | 0.666667 | 0.504985075 |
| Means | AGE | 4.957 | 0.219 | 22.664 | 0 | 22.6347 | 0 |
| Means | FEMALE | 0.929 | 0.058 | 15.987 | 0 | 16.01724 | 0 |
| Means | SITE02 | 0.249 | 0.033 | 7.517 | 0 | 7.545455 | 4.50751E-14 |
| Means | SITE03 | 0.211 | 0.033 | 6.49 | 0 | 6.393939 | 1.61666E-10 |
| Means | SITE04 | 0.306 | 0.034 | 8.97 | 0 | 9 | 0 |
| Means | SITE05 | 0.423 | 0.037 | 11.503 | 0 | 11.43243 | 0 |
| Means | SITE06 | 0.436 | 0.037 | 11.74 | 0 | 11.78378 | 0 |
| Means | SITE07 | 0.429 | 0.037 | 11.623 | 0 | 11.59459 | 0 |
| Means | SITE08 | 0.37 | 0.035 | 10.442 | 0 | 10.57143 | 0 |
| Means | SITE09 | 0.436 | 0.037 | 11.74 | 0 | 11.78378 | 0 |
| Intercepts | GAF_CH | 0.473 | 0.565 | 0.838 | 0.402 | 0.837168 | 0.402498062 |
| Variances | HI_CH | 1 | 0 |  |  |  |  |
| Variances | AGE | 1 | 0 |  |  |  |  |
| Variances | FEMALE | 1 | 0 |  |  |  |  |
| Variances | SITE02 | 1 | 0 |  |  |  |  |
| Variances | SITE03 | 1 | 0 |  |  |  |  |
| Variances | SITE04 | 1 | 0 |  |  |  |  |
| Variances | SITE05 | 1 | 0 |  |  |  |  |
| Variances | SITE06 | 1 | 0 |  |  |  |  |
| Variances | SITE07 | 1 | 0 |  |  |  |  |
| Variances | SITE08 | 1 | 0 |  |  |  |  |
| Variances | SITE09 | 1 | 0 |  |  |  |  |
| Residual.Variances | GAF_CH | 0.171 | 0.033 | 5.226 | 0 | 5.181818 | 2.19733E-07 |
| Residual.Variances | LV_GAF | 0.966 | 0.025 | 38.001 | 0 | 38.64 | 0 |
| LV_NEGS.BY | NEGS_CH | 0.852 | 0.029 | 29.717 | 0 | 29.37931 | 0 |
| LV_NEGS.ON | HI_CH | -0.16 | 0.05 | -3.188 | 0.001 | -3.2 | 0.001374276 |
| LV_NEGS.ON | AGE | -0.018 | 0.073 | -0.251 | 0.802 | -0.24658 | 0.805236889 |
| LV_NEGS.ON | FEMALE | 0.051 | 0.072 | 0.712 | 0.476 | 0.708333 | 0.478738282 |
| LV_NEGS.ON | SITE02 | -0.091 | 0.124 | -0.728 | 0.467 | -0.73387 | 0.463027383 |
| LV_NEGS.ON | SITE03 | 0.026 | 0.114 | 0.229 | 0.819 | 0.22807 | 0.819591686 |
| LV_NEGS.ON | SITE04 | -0.111 | 0.16 | -0.692 | 0.489 | -0.69375 | 0.487839004 |
| LV_NEGS.ON | SITE05 | -0.01 | 0.193 | -0.054 | 0.957 | -0.05181 | 0.958677321 |
| LV_NEGS.ON | SITE06 | -0.032 | 0.205 | -0.155 | 0.877 | -0.1561 | 0.875956121 |
| LV_NEGS.ON | SITE07 | -0.127 | 0.201 | -0.629 | 0.529 | -0.63184 | 0.527490913 |
| LV_NEGS.ON | SITE08 | -0.12 | 0.183 | -0.656 | 0.512 | -0.65574 | 0.511992904 |
| LV_NEGS.ON | SITE09 | 0.066 | 0.207 | 0.321 | 0.748 | 0.318841 | 0.749847405 |
| HI_CH.WITH | AGE | 0.012 | 0.068 | 0.174 | 0.862 | 0.176471 | 0.859924253 |
| HI_CH.WITH | FEMALE | -0.012 | 0.063 | -0.192 | 0.848 | -0.19048 | 0.848935999 |
| HI_CH.WITH | SITE02 | -0.034 | 0.056 | -0.596 | 0.551 | -0.60714 | 0.543756111 |
| HI_CH.WITH | SITE03 | -0.036 | 0.106 | -0.341 | 0.733 | -0.33962 | 0.734140725 |
| HI_CH.WITH | SITE04 | 0.023 | 0.046 | 0.497 | 0.619 | 0.5 | 0.617075077 |
| HI_CH.WITH | SITE05 | -0.005 | 0.064 | -0.074 | 0.941 | -0.07813 | 0.937728621 |
| HI_CH.WITH | SITE06 | 0.062 | 0.068 | 0.92 | 0.358 | 0.911765 | 0.361892593 |
| HI_CH.WITH | SITE07 | 0.014 | 0.057 | 0.241 | 0.809 | 0.245614 | 0.805981024 |
| HI_CH.WITH | SITE08 | -0.038 | 0.056 | -0.672 | 0.501 | -0.67857 | 0.497409451 |
| HI_CH.WITH | SITE09 | 0.008 | 0.059 | 0.143 | 0.886 | 0.135593 | 0.892142865 |
| AGE.WITH | FEMALE | -0.166 | 0.06 | -2.754 | 0.006 | -2.76667 | 0.005663263 |
| AGE.WITH | SITE02 | 0.262 | 0.086 | 3.056 | 0.002 | 3.046512 | 0.002315135 |
| AGE.WITH | SITE03 | -0.01 | 0.053 | -0.184 | 0.854 | -0.18868 | 0.850344219 |
| AGE.WITH | SITE04 | -0.028 | 0.045 | -0.618 | 0.537 | -0.62222 | 0.533795756 |
| AGE.WITH | SITE05 | 0.217 | 0.056 | 3.883 | 0 | 3.875 | 0.000106625 |
| AGE.WITH | SITE06 | 0.028 | 0.058 | 0.489 | 0.625 | 0.482759 | 0.629267135 |
| AGE.WITH | SITE07 | -0.158 | 0.056 | -2.843 | 0.004 | -2.82143 | 0.004781028 |
| AGE.WITH | SITE08 | -0.172 | 0.064 | -2.705 | 0.007 | -2.6875 | 0.00719891 |
| AGE.WITH | SITE09 | -0.009 | 0.061 | -0.154 | 0.878 | -0.14754 | 0.882705033 |
| FEMALE.WITH | SITE02 | 0.054 | 0.062 | 0.868 | 0.385 | 0.870968 | 0.383771767 |
| FEMALE.WITH | SITE03 | -0.002 | 0.062 | -0.031 | 0.975 | -0.03226 | 0.974266251 |
| FEMALE.WITH | SITE04 | -0.031 | 0.062 | -0.498 | 0.618 | -0.5 | 0.617075077 |
| FEMALE.WITH | SITE05 | 0.002 | 0.062 | 0.033 | 0.974 | 0.032258 | 0.974266251 |
| FEMALE.WITH | SITE06 | -0.124 | 0.059 | -2.092 | 0.036 | -2.10169 | 0.035580009 |
| FEMALE.WITH | SITE07 | 0.057 | 0.063 | 0.907 | 0.364 | 0.904762 | 0.365591538 |
| FEMALE.WITH | SITE08 | 0.042 | 0.063 | 0.677 | 0.499 | 0.666667 | 0.504985075 |
| FEMALE.WITH | SITE09 | 0.036 | 0.063 | 0.579 | 0.563 | 0.571429 | 0.567709166 |
| SITE02.WITH | SITE03 | -0.051 | 0.01 | -4.97 | 0 | -5.1 | 3.39653E-07 |
| SITE02.WITH | SITE04 | -0.073 | 0.013 | -5.866 | 0 | -5.61538 | 1.96125E-08 |
| SITE02.WITH | SITE05 | -0.102 | 0.016 | -6.453 | 0 | -6.375 | 1.82963E-10 |
| SITE02.WITH | SITE06 | -0.105 | 0.016 | -6.497 | 0 | -6.5625 | 5.2913E-11 |
| SITE02.WITH | SITE07 | -0.103 | 0.016 | -6.475 | 0 | -6.4375 | 1.21457E-10 |
| SITE02.WITH | SITE08 | -0.089 | 0.014 | -6.236 | 0 | -6.35714 | 2.05541E-10 |
| SITE02.WITH | SITE09 | -0.106 | 0.016 | -6.518 | 0 | -6.625 | 3.47249E-11 |
| SITE03.WITH | SITE04 | -0.065 | 0.012 | -5.427 | 0 | -5.41667 | 6.07204E-08 |
| SITE03.WITH | SITE05 | -0.089 | 0.015 | -5.882 | 0 | -5.93333 | 2.96845E-09 |
| SITE03.WITH | SITE06 | -0.092 | 0.016 | -5.915 | 0 | -5.75 | 8.92434E-09 |
| SITE03.WITH | SITE07 | -0.091 | 0.015 | -5.899 | 0 | -6.06667 | 1.30592E-09 |
| SITE03.WITH | SITE08 | -0.078 | 0.014 | -5.716 | 0 | -5.57143 | 2.52659E-08 |
| SITE03.WITH | SITE09 | -0.093 | 0.016 | -5.931 | 0 | -5.8125 | 6.15467E-09 |
| SITE04.WITH | SITE05 | -0.129 | 0.017 | -7.564 | 0 | -7.58824 | 3.24185E-14 |
| SITE04.WITH | SITE06 | -0.133 | 0.017 | -7.636 | 0 | -7.82353 | 5.10703E-15 |
| SITE04.WITH | SITE07 | -0.131 | 0.017 | -7.601 | 0 | -7.70588 | 1.31006E-14 |
| SITE04.WITH | SITE08 | -0.113 | 0.016 | -7.221 | 0 | -7.0625 | 1.63536E-12 |
| SITE04.WITH | SITE09 | -0.135 | 0.018 | -7.67 | 0 | -7.5 | 6.37268E-14 |
| SITE05.WITH | SITE06 | -0.184 | 0.02 | -9.097 | 0 | -9.2 | 0 |
| SITE05.WITH | SITE07 | -0.182 | 0.02 | -9.038 | 0 | -9.1 | 0 |
| SITE05.WITH | SITE08 | -0.157 | 0.019 | -8.416 | 0 | -8.26316 | 2.22045E-16 |
| SITE05.WITH | SITE09 | -0.187 | 0.02 | -9.155 | 0 | -9.35 | 0 |
| SITE06.WITH | SITE07 | -0.187 | 0.02 | -9.161 | 0 | -9.35 | 0 |
| SITE06.WITH | SITE08 | -0.161 | 0.019 | -8.515 | 0 | -8.47368 | 0 |
| SITE06.WITH | SITE09 | -0.193 | 0.021 | -9.284 | 0 | -9.19048 | 0 |
| SITE07.WITH | SITE08 | -0.159 | 0.019 | -8.466 | 0 | -8.36842 | 0 |
| SITE07.WITH | SITE09 | -0.19 | 0.021 | -9.22 | 0 | -9.04762 | 0 |
| SITE08.WITH | SITE09 | -0.164 | 0.019 | -8.562 | 0 | -8.63158 | 0 |
| Means | HI_CH | 0.047 | 0.063 | 0.747 | 0.455 | 0.746032 | 0.455648238 |
| Means | AGE | 4.953 | 0.218 | 22.69 | 0 | 22.72018 | 0 |
| Means | FEMALE | 0.921 | 0.058 | 15.978 | 0 | 15.87931 | 0 |
| Means | SITE02 | 0.24 | 0.033 | 7.277 | 0 | 7.272727 | 3.52385E-13 |
| Means | SITE03 | 0.211 | 0.033 | 6.49 | 0 | 6.393939 | 1.61666E-10 |
| Means | SITE04 | 0.306 | 0.034 | 8.97 | 0 | 9 | 0 |
| Means | SITE05 | 0.423 | 0.037 | 11.503 | 0 | 11.43243 | 0 |
| Means | SITE06 | 0.436 | 0.037 | 11.74 | 0 | 11.78378 | 0 |
| Means | SITE07 | 0.429 | 0.037 | 11.623 | 0 | 11.59459 | 0 |
| Means | SITE08 | 0.37 | 0.035 | 10.442 | 0 | 10.57143 | 0 |
| Means | SITE09 | 0.442 | 0.037 | 11.855 | 0 | 11.94595 | 0 |
| Intercepts | NEGS_CH | -0.012 | 0.591 | -0.02 | 0.984 | -0.0203 | 0.983800411 |
| Variances | HI_CH | 1 | 0 |  |  |  |  |
| Variances | AGE | 1 | 0 |  |  |  |  |
| Variances | FEMALE | 1 | 0 |  |  |  |  |
| Variances | SITE02 | 1 | 0 |  |  |  |  |
| Variances | SITE03 | 1 | 0 |  |  |  |  |
| Variances | SITE04 | 1 | 0 |  |  |  |  |
| Variances | SITE05 | 1 | 0 |  |  |  |  |
| Variances | SITE06 | 1 | 0 |  |  |  |  |
| Variances | SITE07 | 1 | 0 |  |  |  |  |
| Variances | SITE08 | 1 | 0 |  |  |  |  |
| Variances | SITE09 | 1 | 0 |  |  |  |  |
| Residual.Variances | NEGS_CH | 0.273 | 0.049 | 5.593 | 0 | 5.571429 | 2.52659E-08 |
| Residual.Variances | LV_NEGS | 0.927 | 0.031 | 30.14 | 0 | 29.90323 | 0 |

Table_S6b_PredAnalysisStand_FCtoPredictClin_MSum

| **behav** | **neuro** | **estimator** | **n_obs** | **n_parameters** | **aic** | **bic** |
| --- | --- | --- | --- | --- | --- | --- |
| NegS | fchi | MLR | 257 | 90 | 2582.105 | 2901.522 |
| DepS | fchi | MLR | 259 | 90 | 2490.654 | 2810.769 |
| gaf | fchi | MLR | 257 | 90 | 3068.781 | 3388.198 |

Table_S6c_PredAnalysisStand_ClinToPredictFC

| **paramHeader** | **param** | **est** | **se** | **est_se** | **pval** | **zval** | **pval_exact** |
| --- | --- | --- | --- | --- | --- | --- | --- |
| LV_HI.BY | HI_CH | 0.819 | 0.025 | 32.206 | 0 | 32.76 | 0 |
| LV_HI.ON | DEPS_CH | 0.047 | 0.078 | 0.609 | 0.543 | 0.602564 | 0.546798707 |
| LV_HI.ON | AGE | 0.066 | 0.112 | 0.59 | 0.555 | 0.589286 | 0.555669626 |
| LV_HI.ON | FEMALE | -0.016 | 0.087 | -0.188 | 0.851 | -0.18391 | 0.854085593 |
| LV_HI.ON | SITE02 | -0.024 | 0.134 | -0.182 | 0.856 | -0.1791 | 0.857855666 |
| LV_HI.ON | SITE03 | 0.051 | 0.114 | 0.445 | 0.656 | 0.447368 | 0.654609073 |
| LV_HI.ON | SITE04 | 0.094 | 0.126 | 0.751 | 0.452 | 0.746032 | 0.455648238 |
| LV_HI.ON | SITE05 | -0.092 | 0.146 | -0.626 | 0.531 | -0.63014 | 0.528604963 |
| LV_HI.ON | SITE06 | -0.051 | 0.136 | -0.372 | 0.71 | -0.375 | 0.707660467 |
| LV_HI.ON | SITE07 | -0.111 | 0.144 | -0.773 | 0.44 | -0.77083 | 0.440805727 |
| LV_HI.ON | SITE08 | -0.024 | 0.138 | -0.171 | 0.865 | -0.17391 | 0.861933802 |
| LV_HI.ON | SITE09 | -0.143 | 0.149 | -0.959 | 0.338 | -0.95973 | 0.337190343 |
| DEPS_CH.WITH | AGE | 0.035 | 0.067 | 0.529 | 0.597 | 0.522388 | 0.601400168 |
| DEPS_CH.WITH | FEMALE | -0.045 | 0.064 | -0.707 | 0.48 | -0.70313 | 0.481977854 |
| DEPS_CH.WITH | SITE02 | -0.04 | 0.075 | -0.526 | 0.599 | -0.53333 | 0.593802857 |
| DEPS_CH.WITH | SITE03 | 0.102 | 0.059 | 1.734 | 0.083 | 1.728814 | 0.083842469 |
| DEPS_CH.WITH | SITE04 | -0.13 | 0.067 | -1.94 | 0.052 | -1.9403 | 0.052343423 |
| DEPS_CH.WITH | SITE05 | 0.076 | 0.048 | 1.581 | 0.114 | 1.583333 | 0.113345509 |
| DEPS_CH.WITH | SITE06 | 0.018 | 0.074 | 0.241 | 0.81 | 0.243243 | 0.807816969 |
| DEPS_CH.WITH | SITE07 | -0.025 | 0.056 | -0.44 | 0.66 | -0.44643 | 0.655287698 |
| DEPS_CH.WITH | SITE08 | 0.081 | 0.068 | 1.192 | 0.233 | 1.191176 | 0.233584315 |
| DEPS_CH.WITH | SITE09 | -0.025 | 0.064 | -0.389 | 0.697 | -0.39063 | 0.696074444 |
| AGE.WITH | FEMALE | -0.159 | 0.063 | -2.521 | 0.012 | -2.52381 | 0.011609081 |
| AGE.WITH | SITE02 | 0.336 | 0.076 | 4.441 | 0 | 4.421053 | 9.82212E-06 |
| AGE.WITH | SITE03 | 0.007 | 0.055 | 0.134 | 0.893 | 0.127273 | 0.898724545 |
| AGE.WITH | SITE04 | 0.011 | 0.059 | 0.185 | 0.853 | 0.186441 | 0.852099195 |
| AGE.WITH | SITE05 | 0.175 | 0.054 | 3.258 | 0.001 | 3.240741 | 0.001192196 |
| AGE.WITH | SITE06 | -0.025 | 0.057 | -0.434 | 0.664 | -0.4386 | 0.66095394 |
| AGE.WITH | SITE07 | -0.159 | 0.056 | -2.82 | 0.005 | -2.83929 | 0.004521465 |
| AGE.WITH | SITE08 | -0.214 | 0.066 | -3.236 | 0.001 | -3.24242 | 0.001185175 |
| AGE.WITH | SITE09 | -0.021 | 0.06 | -0.346 | 0.73 | -0.35 | 0.726338698 |
| FEMALE.WITH | SITE02 | 0.04 | 0.065 | 0.613 | 0.54 | 0.615385 | 0.538300749 |
| FEMALE.WITH | SITE03 | -0.061 | 0.06 | -1.015 | 0.31 | -1.01667 | 0.309312028 |
| FEMALE.WITH | SITE04 | -0.036 | 0.064 | -0.57 | 0.569 | -0.5625 | 0.573775404 |
| FEMALE.WITH | SITE05 | 0 | 0.065 | -0.006 | 0.995 | 0 | 1 |
| FEMALE.WITH | SITE06 | -0.094 | 0.063 | -1.489 | 0.137 | -1.49206 | 0.135682501 |
| FEMALE.WITH | SITE07 | 0.057 | 0.065 | 0.876 | 0.381 | 0.876923 | 0.380528421 |
| FEMALE.WITH | SITE08 | 0.001 | 0.065 | 0.018 | 0.986 | 0.015385 | 0.987725337 |
| FEMALE.WITH | SITE09 | 0.07 | 0.065 | 1.081 | 0.28 | 1.076923 | 0.281514633 |
| SITE02.WITH | SITE03 | -0.054 | 0.012 | -4.589 | 0 | -4.5 | 6.79535E-06 |
| SITE02.WITH | SITE04 | -0.095 | 0.015 | -6.43 | 0 | -6.33333 | 2.39921E-10 |
| SITE02.WITH | SITE05 | -0.134 | 0.018 | -7.387 | 0 | -7.44444 | 9.72555E-14 |
| SITE02.WITH | SITE06 | -0.134 | 0.018 | -7.387 | 0 | -7.44444 | 9.72555E-14 |
| SITE02.WITH | SITE07 | -0.128 | 0.018 | -7.265 | 0 | -7.11111 | 1.15108E-12 |
| SITE02.WITH | SITE08 | -0.109 | 0.016 | -6.858 | 0 | -6.8125 | 9.59166E-12 |
| SITE02.WITH | SITE09 | -0.143 | 0.019 | -7.53 | 0 | -7.52632 | 5.21805E-14 |
| SITE03.WITH | SITE04 | -0.053 | 0.012 | -4.561 | 0 | -4.41667 | 1.00235E-05 |
| SITE03.WITH | SITE05 | -0.075 | 0.015 | -4.866 | 0 | -5 | 5.73303E-07 |
| SITE03.WITH | SITE06 | -0.075 | 0.015 | -4.866 | 0 | -5 | 5.73303E-07 |
| SITE03.WITH | SITE07 | -0.071 | 0.015 | -4.831 | 0 | -4.73333 | 2.20862E-06 |
| SITE03.WITH | SITE08 | -0.061 | 0.013 | -4.706 | 0 | -4.69231 | 2.7014E-06 |
| SITE03.WITH | SITE09 | -0.08 | 0.016 | -4.906 | 0 | -5 | 5.73303E-07 |
| SITE04.WITH | SITE05 | -0.131 | 0.018 | -7.268 | 0 | -7.27778 | 3.39284E-13 |
| SITE04.WITH | SITE06 | -0.131 | 0.018 | -7.268 | 0 | -7.27778 | 3.39284E-13 |
| SITE04.WITH | SITE07 | -0.124 | 0.017 | -7.153 | 0 | -7.29412 | 3.00648E-13 |
| SITE04.WITH | SITE08 | -0.107 | 0.016 | -6.763 | 0 | -6.6875 | 2.27016E-11 |
| SITE04.WITH | SITE09 | -0.139 | 0.019 | -7.405 | 0 | -7.31579 | 2.55795E-13 |
| SITE05.WITH | SITE06 | -0.185 | 0.021 | -8.753 | 0 | -8.80952 | 0 |
| SITE05.WITH | SITE07 | -0.176 | 0.021 | -8.552 | 0 | -8.38095 | 0 |
| SITE05.WITH | SITE08 | -0.151 | 0.019 | -7.906 | 0 | -7.94737 | 1.9984E-15 |
| SITE05.WITH | SITE09 | -0.197 | 0.022 | -8.996 | 0 | -8.95455 | 0 |
| SITE06.WITH | SITE07 | -0.176 | 0.021 | -8.552 | 0 | -8.38095 | 0 |
| SITE06.WITH | SITE08 | -0.151 | 0.019 | -7.906 | 0 | -7.94737 | 1.9984E-15 |
| SITE06.WITH | SITE09 | -0.197 | 0.022 | -8.996 | 0 | -8.95455 | 0 |
| SITE07.WITH | SITE08 | -0.144 | 0.019 | -7.757 | 0 | -7.57895 | 3.4861E-14 |
| SITE07.WITH | SITE09 | -0.187 | 0.021 | -8.778 | 0 | -8.90476 | 0 |
| SITE08.WITH | SITE09 | -0.161 | 0.02 | -8.082 | 0 | -8.05 | 8.88178E-16 |
| Means | DEPS_CH | -0.24 | 0.065 | -3.661 | 0 | -3.69231 | 0.000222228 |
| Means | AGE | 4.784 | 0.208 | 22.999 | 0 | 23 | 0 |
| Means | FEMALE | 0.923 | 0.06 | 15.345 | 0 | 15.38333 | 0 |
| Means | SITE02 | 0.312 | 0.036 | 8.75 | 0 | 8.666667 | 0 |
| Means | SITE03 | 0.174 | 0.033 | 5.213 | 0 | 5.272727 | 1.34411E-07 |
| Means | SITE04 | 0.304 | 0.035 | 8.559 | 0 | 8.685714 | 0 |
| Means | SITE05 | 0.43 | 0.038 | 11.176 | 0 | 11.31579 | 0 |
| Means | SITE06 | 0.43 | 0.038 | 11.176 | 0 | 11.31579 | 0 |
| Means | SITE07 | 0.409 | 0.038 | 10.793 | 0 | 10.76316 | 0 |
| Means | SITE08 | 0.351 | 0.036 | 9.622 | 0 | 9.75 | 0 |
| Means | SITE09 | 0.457 | 0.039 | 11.646 | 0 | 11.71795 | 0 |
| Intercepts | HI_CH | -0.101 | 0.489 | -0.208 | 0.836 | -0.20654 | 0.836366024 |
| Variances | DEPS_CH | 1 | 0 |  |  |  |  |
| Variances | AGE | 1 | 0 |  |  |  |  |
| Variances | FEMALE | 1 | 0 |  |  |  |  |
| Variances | SITE02 | 1 | 0 |  |  |  |  |
| Variances | SITE03 | 1 | 0 |  |  |  |  |
| Variances | SITE04 | 1 | 0 |  |  |  |  |
| Variances | SITE05 | 1 | 0 |  |  |  |  |
| Variances | SITE06 | 1 | 0 |  |  |  |  |
| Variances | SITE07 | 1 | 0 |  |  |  |  |
| Variances | SITE08 | 1 | 0 |  |  |  |  |
| Variances | SITE09 | 1 | 0 |  |  |  |  |
| Residual.Variances | HI_CH | 0.33 | 0.042 | 7.918 | 0 | 7.857143 | 3.9968E-15 |
| Residual.Variances | LV_HI | 0.949 | 0.034 | 27.961 | 0 | 27.91176 | 0 |
| LV_HI.BY | HI_CH | 0.821 | 0.025 | 32.966 | 0 | 32.84 | 0 |
| LV_HI.ON | GAF_CH | 0.023 | 0.092 | 0.25 | 0.803 | 0.25 | 0.802587349 |
| LV_HI.ON | AGE | 0.061 | 0.112 | 0.545 | 0.585 | 0.544643 | 0.585999173 |
| LV_HI.ON | FEMALE | -0.013 | 0.086 | -0.155 | 0.877 | -0.15116 | 0.879847302 |
| LV_HI.ON | SITE02 | -0.008 | 0.132 | -0.058 | 0.954 | -0.06061 | 0.951672947 |
| LV_HI.ON | SITE03 | 0.066 | 0.113 | 0.581 | 0.561 | 0.584071 | 0.559172682 |
| LV_HI.ON | SITE04 | 0.104 | 0.121 | 0.857 | 0.392 | 0.859504 | 0.390062441 |
| LV_HI.ON | SITE05 | -0.065 | 0.142 | -0.456 | 0.648 | -0.45775 | 0.647134592 |
| LV_HI.ON | SITE06 | -0.027 | 0.132 | -0.204 | 0.838 | -0.20455 | 0.837927275 |
| LV_HI.ON | SITE07 | -0.088 | 0.141 | -0.626 | 0.532 | -0.62411 | 0.532553065 |
| LV_HI.ON | SITE08 | -0.008 | 0.132 | -0.06 | 0.953 | -0.06061 | 0.951672947 |
| LV_HI.ON | SITE09 | -0.105 | 0.146 | -0.717 | 0.474 | -0.71918 | 0.472031202 |
| GAF_CH.WITH | AGE | -0.019 | 0.06 | -0.318 | 0.75 | -0.31667 | 0.751496544 |
| GAF_CH.WITH | FEMALE | 0.001 | 0.065 | 0.015 | 0.988 | 0.015385 | 0.987725337 |
| GAF_CH.WITH | SITE02 | 0.031 | 0.046 | 0.663 | 0.507 | 0.673913 | 0.500366599 |
| GAF_CH.WITH | SITE03 | -0.013 | 0.023 | -0.555 | 0.579 | -0.56522 | 0.571925909 |
| GAF_CH.WITH | SITE04 | 0.068 | 0.037 | 1.82 | 0.069 | 1.837838 | 0.066086304 |
| GAF_CH.WITH | SITE05 | -0.019 | 0.068 | -0.283 | 0.777 | -0.27941 | 0.779928844 |
| GAF_CH.WITH | SITE06 | -0.016 | 0.046 | -0.342 | 0.732 | -0.34783 | 0.727970797 |
| GAF_CH.WITH | SITE07 | -0.148 | 0.078 | -1.901 | 0.057 | -1.89744 | 0.057770432 |
| GAF_CH.WITH | SITE08 | 0.063 | 0.088 | 0.715 | 0.475 | 0.715909 | 0.474047485 |
| GAF_CH.WITH | SITE09 | 0.026 | 0.05 | 0.519 | 0.604 | 0.52 | 0.603063575 |
| AGE.WITH | FEMALE | -0.167 | 0.063 | -2.671 | 0.008 | -2.65079 | 0.008030289 |
| AGE.WITH | SITE02 | 0.336 | 0.075 | 4.457 | 0 | 4.48 | 7.4643E-06 |
| AGE.WITH | SITE03 | 0.008 | 0.054 | 0.149 | 0.881 | 0.148148 | 0.882225852 |
| AGE.WITH | SITE04 | 0.012 | 0.059 | 0.21 | 0.834 | 0.20339 | 0.838830348 |
| AGE.WITH | SITE05 | 0.176 | 0.054 | 3.293 | 0.001 | 3.259259 | 0.001117035 |
| AGE.WITH | SITE06 | -0.023 | 0.057 | -0.398 | 0.691 | -0.40351 | 0.686573983 |
| AGE.WITH | SITE07 | -0.156 | 0.056 | -2.783 | 0.005 | -2.78571 | 0.005340993 |
| AGE.WITH | SITE08 | -0.196 | 0.066 | -2.961 | 0.003 | -2.9697 | 0.002980936 |
| AGE.WITH | SITE09 | -0.037 | 0.061 | -0.603 | 0.547 | -0.60656 | 0.544144694 |
| FEMALE.WITH | SITE02 | 0.038 | 0.065 | 0.591 | 0.554 | 0.584615 | 0.558806361 |
| FEMALE.WITH | SITE03 | -0.062 | 0.06 | -1.028 | 0.304 | -1.03333 | 0.301447933 |
| FEMALE.WITH | SITE04 | -0.038 | 0.064 | -0.591 | 0.554 | -0.59375 | 0.552679336 |
| FEMALE.WITH | SITE05 | -0.002 | 0.065 | -0.036 | 0.971 | -0.03077 | 0.975453579 |
| FEMALE.WITH | SITE06 | -0.095 | 0.063 | -1.519 | 0.129 | -1.50794 | 0.131570778 |
| FEMALE.WITH | SITE07 | 0.055 | 0.065 | 0.847 | 0.397 | 0.846154 | 0.397466925 |
| FEMALE.WITH | SITE08 | 0.012 | 0.065 | 0.188 | 0.85 | 0.184615 | 0.853530718 |
| FEMALE.WITH | SITE09 | 0.079 | 0.065 | 1.223 | 0.221 | 1.215385 | 0.224219433 |
| SITE02.WITH | SITE03 | -0.054 | 0.012 | -4.589 | 0 | -4.5 | 6.79535E-06 |
| SITE02.WITH | SITE04 | -0.094 | 0.015 | -6.43 | 0 | -6.26667 | 3.68858E-10 |
| SITE02.WITH | SITE05 | -0.133 | 0.018 | -7.386 | 0 | -7.38889 | 1.48104E-13 |
| SITE02.WITH | SITE06 | -0.133 | 0.018 | -7.386 | 0 | -7.38889 | 1.48104E-13 |
| SITE02.WITH | SITE07 | -0.127 | 0.017 | -7.265 | 0 | -7.47059 | 7.99361E-14 |
| SITE02.WITH | SITE08 | -0.107 | 0.016 | -6.795 | 0 | -6.6875 | 2.27016E-11 |
| SITE02.WITH | SITE09 | -0.144 | 0.019 | -7.562 | 0 | -7.57895 | 3.4861E-14 |
| SITE03.WITH | SITE04 | -0.053 | 0.012 | -4.561 | 0 | -4.41667 | 1.00235E-05 |
| SITE03.WITH | SITE05 | -0.075 | 0.015 | -4.866 | 0 | -5 | 5.73303E-07 |
| SITE03.WITH | SITE06 | -0.075 | 0.015 | -4.866 | 0 | -5 | 5.73303E-07 |
| SITE03.WITH | SITE07 | -0.071 | 0.015 | -4.831 | 0 | -4.73333 | 2.20862E-06 |
| SITE03.WITH | SITE08 | -0.06 | 0.013 | -4.685 | 0 | -4.61538 | 3.92368E-06 |
| SITE03.WITH | SITE09 | -0.081 | 0.016 | -4.915 | 0 | -5.0625 | 4.13794E-07 |
| SITE04.WITH | SITE05 | -0.13 | 0.018 | -7.268 | 0 | -7.22222 | 5.11369E-13 |
| SITE04.WITH | SITE06 | -0.13 | 0.018 | -7.268 | 0 | -7.22222 | 5.11369E-13 |
| SITE04.WITH | SITE07 | -0.124 | 0.017 | -7.152 | 0 | -7.29412 | 3.00648E-13 |
| SITE04.WITH | SITE08 | -0.104 | 0.015 | -6.703 | 0 | -6.93333 | 4.11049E-12 |
| SITE04.WITH | SITE09 | -0.14 | 0.019 | -7.435 | 0 | -7.36842 | 1.72751E-13 |
| SITE05.WITH | SITE06 | -0.184 | 0.021 | -8.752 | 0 | -8.7619 | 0 |
| SITE05.WITH | SITE07 | -0.175 | 0.02 | -8.551 | 0 | -8.75 | 0 |
| SITE05.WITH | SITE08 | -0.147 | 0.019 | -7.81 | 0 | -7.73684 | 1.02141E-14 |
| SITE05.WITH | SITE09 | -0.199 | 0.022 | -9.051 | 0 | -9.04545 | 0 |
| SITE06.WITH | SITE07 | -0.175 | 0.02 | -8.551 | 0 | -8.75 | 0 |
| SITE06.WITH | SITE08 | -0.147 | 0.019 | -7.81 | 0 | -7.73684 | 1.02141E-14 |
| SITE06.WITH | SITE09 | -0.199 | 0.022 | -9.051 | 0 | -9.04545 | 0 |
| SITE07.WITH | SITE08 | -0.14 | 0.018 | -7.667 | 0 | -7.77778 | 7.32747E-15 |
| SITE07.WITH | SITE09 | -0.189 | 0.021 | -8.828 | 0 | -9 | 0 |
| SITE08.WITH | SITE09 | -0.159 | 0.02 | -8.019 | 0 | -7.95 | 1.77636E-15 |
| Means | GAF_CH | 0.173 | 0.066 | 2.605 | 0.009 | 2.621212 | 0.008761773 |
| Means | AGE | 4.773 | 0.207 | 23.017 | 0 | 23.05797 | 0 |
| Means | FEMALE | 0.927 | 0.06 | 15.383 | 0 | 15.45 | 0 |
| Means | SITE02 | 0.311 | 0.036 | 8.751 | 0 | 8.638889 | 0 |
| Means | SITE03 | 0.174 | 0.033 | 5.213 | 0 | 5.272727 | 1.34411E-07 |
| Means | SITE04 | 0.303 | 0.035 | 8.56 | 0 | 8.657143 | 0 |
| Means | SITE05 | 0.429 | 0.038 | 11.18 | 0 | 11.28947 | 0 |
| Means | SITE06 | 0.429 | 0.038 | 11.18 | 0 | 11.28947 | 0 |
| Means | SITE07 | 0.408 | 0.038 | 10.797 | 0 | 10.73684 | 0 |
| Means | SITE08 | 0.343 | 0.036 | 9.46 | 0 | 9.527778 | 0 |
| Means | SITE09 | 0.463 | 0.039 | 11.762 | 0 | 11.87179 | 0 |
| Intercepts | HI_CH | -0.149 | 0.478 | -0.311 | 0.756 | -0.31172 | 0.755256762 |
| Variances | GAF_CH | 1 | 0 |  |  |  |  |
| Variances | AGE | 1 | 0 |  |  |  |  |
| Variances | FEMALE | 1 | 0 |  |  |  |  |
| Variances | SITE02 | 1 | 0 |  |  |  |  |
| Variances | SITE03 | 1 | 0 |  |  |  |  |
| Variances | SITE04 | 1 | 0 |  |  |  |  |
| Variances | SITE05 | 1 | 0 |  |  |  |  |
| Variances | SITE06 | 1 | 0 |  |  |  |  |
| Variances | SITE07 | 1 | 0 |  |  |  |  |
| Variances | SITE08 | 1 | 0 |  |  |  |  |
| Variances | SITE09 | 1 | 0 |  |  |  |  |
| Residual.Variances | HI_CH | 0.326 | 0.041 | 7.978 | 0 | 7.95122 | 1.77636E-15 |
| Residual.Variances | LV_HI | 0.956 | 0.033 | 28.87 | 0 | 28.9697 | 0 |
| LV_HI.BY | HI_CH | 0.808 | 0.027 | 29.78 | 0 | 29.92593 | 0 |
| LV_HI.ON | NEGS_CH | -0.059 | 0.097 | -0.606 | 0.544 | -0.60825 | 0.543023386 |
| LV_HI.ON | AGE | 0.075 | 0.114 | 0.655 | 0.512 | 0.657895 | 0.510605773 |
| LV_HI.ON | FEMALE | -0.01 | 0.088 | -0.111 | 0.912 | -0.11364 | 0.90952606 |
| LV_HI.ON | SITE02 | -0.019 | 0.133 | -0.139 | 0.889 | -0.14286 | 0.886403006 |
| LV_HI.ON | SITE03 | 0.069 | 0.116 | 0.592 | 0.554 | 0.594828 | 0.551958726 |
| LV_HI.ON | SITE04 | 0.1 | 0.123 | 0.812 | 0.417 | 0.813008 | 0.416213397 |
| LV_HI.ON | SITE05 | -0.06 | 0.143 | -0.42 | 0.675 | -0.41958 | 0.674791995 |
| LV_HI.ON | SITE06 | -0.027 | 0.132 | -0.202 | 0.84 | -0.20455 | 0.837927275 |
| LV_HI.ON | SITE07 | -0.091 | 0.14 | -0.651 | 0.515 | -0.65 | 0.515692222 |
| LV_HI.ON | SITE08 | -0.004 | 0.133 | -0.028 | 0.977 | -0.03008 | 0.976007089 |
| LV_HI.ON | SITE09 | -0.12 | 0.144 | -0.836 | 0.403 | -0.83333 | 0.404656762 |
| NEGS_CH.WITH | AGE | 0.011 | 0.066 | 0.163 | 0.871 | 0.166667 | 0.867632335 |
| NEGS_CH.WITH | FEMALE | 0.104 | 0.062 | 1.679 | 0.093 | 1.677419 | 0.093460507 |
| NEGS_CH.WITH | SITE02 | 0.018 | 0.062 | 0.291 | 0.771 | 0.290323 | 0.771569466 |
| NEGS_CH.WITH | SITE03 | 0.02 | 0.022 | 0.91 | 0.363 | 0.909091 | 0.363302141 |
| NEGS_CH.WITH | SITE04 | -0.155 | 0.057 | -2.73 | 0.006 | -2.7193 | 0.006542059 |
| NEGS_CH.WITH | SITE05 | 0.142 | 0.044 | 3.197 | 0.001 | 3.227273 | 0.001249763 |
| NEGS_CH.WITH | SITE06 | -0.012 | 0.075 | -0.155 | 0.877 | -0.16 | 0.872881074 |
| NEGS_CH.WITH | SITE07 | -0.024 | 0.053 | -0.459 | 0.646 | -0.45283 | 0.650671024 |
| NEGS_CH.WITH | SITE08 | -0.002 | 0.058 | -0.033 | 0.974 | -0.03448 | 0.972492191 |
| NEGS_CH.WITH | SITE09 | 0.026 | 0.069 | 0.379 | 0.705 | 0.376812 | 0.706313624 |
| AGE.WITH | FEMALE | -0.161 | 0.063 | -2.563 | 0.01 | -2.55556 | 0.010601844 |
| AGE.WITH | SITE02 | 0.336 | 0.076 | 4.434 | 0 | 4.421053 | 9.82212E-06 |
| AGE.WITH | SITE03 | 0.007 | 0.055 | 0.127 | 0.899 | 0.127273 | 0.898724545 |
| AGE.WITH | SITE04 | 0.01 | 0.059 | 0.173 | 0.862 | 0.169492 | 0.865410038 |
| AGE.WITH | SITE05 | 0.174 | 0.054 | 3.243 | 0.001 | 3.222222 | 0.001272004 |
| AGE.WITH | SITE06 | -0.026 | 0.057 | -0.452 | 0.651 | -0.45614 | 0.648289056 |
| AGE.WITH | SITE07 | -0.16 | 0.056 | -2.839 | 0.005 | -2.85714 | 0.004274734 |
| AGE.WITH | SITE08 | -0.2 | 0.067 | -3 | 0.003 | -2.98507 | 0.002835091 |
| AGE.WITH | SITE09 | -0.022 | 0.06 | -0.363 | 0.716 | -0.36667 | 0.713867673 |
| FEMALE.WITH | SITE02 | 0.04 | 0.065 | 0.613 | 0.54 | 0.615385 | 0.538300749 |
| FEMALE.WITH | SITE03 | -0.061 | 0.06 | -1.015 | 0.31 | -1.01667 | 0.309312028 |
| FEMALE.WITH | SITE04 | -0.036 | 0.064 | -0.57 | 0.569 | -0.5625 | 0.573775404 |
| FEMALE.WITH | SITE05 | 0 | 0.065 | -0.006 | 0.995 | 0 | 1 |
| FEMALE.WITH | SITE06 | -0.094 | 0.063 | -1.489 | 0.137 | -1.49206 | 0.135682501 |
| FEMALE.WITH | SITE07 | 0.057 | 0.065 | 0.876 | 0.381 | 0.876923 | 0.380528421 |
| FEMALE.WITH | SITE08 | 0.014 | 0.065 | 0.213 | 0.832 | 0.215385 | 0.829467464 |
| FEMALE.WITH | SITE09 | 0.07 | 0.065 | 1.081 | 0.28 | 1.076923 | 0.281514633 |
| SITE02.WITH | SITE03 | -0.054 | 0.012 | -4.589 | 0 | -4.5 | 6.79535E-06 |
| SITE02.WITH | SITE04 | -0.095 | 0.015 | -6.43 | 0 | -6.33333 | 2.39921E-10 |
| SITE02.WITH | SITE05 | -0.134 | 0.018 | -7.387 | 0 | -7.44444 | 9.72555E-14 |
| SITE02.WITH | SITE06 | -0.134 | 0.018 | -7.387 | 0 | -7.44444 | 9.72555E-14 |
| SITE02.WITH | SITE07 | -0.128 | 0.018 | -7.265 | 0 | -7.11111 | 1.15108E-12 |
| SITE02.WITH | SITE08 | -0.107 | 0.016 | -6.796 | 0 | -6.6875 | 2.27016E-11 |
| SITE02.WITH | SITE09 | -0.143 | 0.019 | -7.53 | 0 | -7.52632 | 5.21805E-14 |
| SITE03.WITH | SITE04 | -0.053 | 0.012 | -4.561 | 0 | -4.41667 | 1.00235E-05 |
| SITE03.WITH | SITE05 | -0.075 | 0.015 | -4.866 | 0 | -5 | 5.73303E-07 |
| SITE03.WITH | SITE06 | -0.075 | 0.015 | -4.866 | 0 | -5 | 5.73303E-07 |
| SITE03.WITH | SITE07 | -0.071 | 0.015 | -4.831 | 0 | -4.73333 | 2.20862E-06 |
| SITE03.WITH | SITE08 | -0.06 | 0.013 | -4.685 | 0 | -4.61538 | 3.92368E-06 |
| SITE03.WITH | SITE09 | -0.08 | 0.016 | -4.906 | 0 | -5 | 5.73303E-07 |
| SITE04.WITH | SITE05 | -0.131 | 0.018 | -7.268 | 0 | -7.27778 | 3.39284E-13 |
| SITE04.WITH | SITE06 | -0.131 | 0.018 | -7.268 | 0 | -7.27778 | 3.39284E-13 |
| SITE04.WITH | SITE07 | -0.124 | 0.017 | -7.153 | 0 | -7.29412 | 3.00648E-13 |
| SITE04.WITH | SITE08 | -0.104 | 0.016 | -6.703 | 0 | -6.5 | 8.032E-11 |
| SITE04.WITH | SITE09 | -0.139 | 0.019 | -7.405 | 0 | -7.31579 | 2.55795E-13 |
| SITE05.WITH | SITE06 | -0.185 | 0.021 | -8.753 | 0 | -8.80952 | 0 |
| SITE05.WITH | SITE07 | -0.176 | 0.021 | -8.552 | 0 | -8.38095 | 0 |
| SITE05.WITH | SITE08 | -0.148 | 0.019 | -7.811 | 0 | -7.78947 | 6.66134E-15 |
| SITE05.WITH | SITE09 | -0.197 | 0.022 | -8.996 | 0 | -8.95455 | 0 |
| SITE06.WITH | SITE07 | -0.176 | 0.021 | -8.552 | 0 | -8.38095 | 0 |
| SITE06.WITH | SITE08 | -0.148 | 0.019 | -7.811 | 0 | -7.78947 | 6.66134E-15 |
| SITE06.WITH | SITE09 | -0.197 | 0.022 | -8.996 | 0 | -8.95455 | 0 |
| SITE07.WITH | SITE08 | -0.141 | 0.018 | -7.667 | 0 | -7.83333 | 4.66294E-15 |
| SITE07.WITH | SITE09 | -0.187 | 0.021 | -8.778 | 0 | -8.90476 | 0 |
| SITE08.WITH | SITE09 | -0.157 | 0.02 | -7.981 | 0 | -7.85 | 4.21885E-15 |
| Means | NEGS_CH | -0.254 | 0.064 | -3.959 | 0 | -3.96875 | 7.22506E-05 |
| Means | AGE | 4.798 | 0.209 | 22.954 | 0 | 22.95694 | 0 |
| Means | FEMALE | 0.923 | 0.06 | 15.345 | 0 | 15.38333 | 0 |
| Means | SITE02 | 0.312 | 0.036 | 8.75 | 0 | 8.666667 | 0 |
| Means | SITE03 | 0.174 | 0.033 | 5.213 | 0 | 5.272727 | 1.34411E-07 |
| Means | SITE04 | 0.304 | 0.035 | 8.559 | 0 | 8.685714 | 0 |
| Means | SITE05 | 0.43 | 0.038 | 11.176 | 0 | 11.31579 | 0 |
| Means | SITE06 | 0.43 | 0.038 | 11.176 | 0 | 11.31579 | 0 |
| Means | SITE07 | 0.409 | 0.038 | 10.793 | 0 | 10.76316 | 0 |
| Means | SITE08 | 0.343 | 0.036 | 9.458 | 0 | 9.527778 | 0 |
| Means | SITE09 | 0.457 | 0.039 | 11.646 | 0 | 11.71795 | 0 |
| Intercepts | HI_CH | -0.212 | 0.483 | -0.438 | 0.661 | -0.43892 | 0.660717044 |
| Variances | NEGS_CH | 1 | 0 |  |  |  |  |
| Variances | AGE | 1 | 0 |  |  |  |  |
| Variances | FEMALE | 1 | 0 |  |  |  |  |
| Variances | SITE02 | 1 | 0 |  |  |  |  |
| Variances | SITE03 | 1 | 0 |  |  |  |  |
| Variances | SITE04 | 1 | 0 |  |  |  |  |
| Variances | SITE05 | 1 | 0 |  |  |  |  |
| Variances | SITE06 | 1 | 0 |  |  |  |  |
| Variances | SITE07 | 1 | 0 |  |  |  |  |
| Variances | SITE08 | 1 | 0 |  |  |  |  |
| Variances | SITE09 | 1 | 0 |  |  |  |  |
| Residual.Variances | HI_CH | 0.347 | 0.044 | 7.929 | 0 | 7.886364 | 3.10862E-15 |
| Residual.Variances | LV_HI | 0.947 | 0.037 | 25.493 | 0 | 25.59459 | 0 |

Table_S6d_PredAnalysisStand_ClinToPredictFC_MSum

| **behav** | **neuro** | **estimator** | **n_obs** | **n_parameters** | **aic** | **bic** |
| --- | --- | --- | --- | --- | --- | --- |
| NegS | fchi | MLR | 237 | 90 | 2368.701 | 2680.826 |
| DepS | fchi | MLR | 237 | 90 | 2416.982 | 2729.107 |
| gaf | fchi | MLR | 238 | 90 | 2746.497 | 3059.001 |

Table_S7a_SensAnalysisStand_WithinHippo

| **paramHeader** | **param** | **est** | **se** | **est_se** | **pval** | **zval** | **pval_exact** |
| --- | --- | --- | --- | --- | --- | --- | --- |
| LV_HIH.BY | HIH_CH | 0.818 | 0.024 | 33.938 | 0 | 34.08333 | 0 |
| LV_DEPS.BY | DEPS_CH | 0.863 | 0.036 | 23.861 | 0 | 23.97222 | 0 |
| LV_DEPS.ON | LV_HIH | 0.016 | 0.038 | 0.411 | 0.681 | 0.421053 | 0.673717 |
| LV_DEPS.ON | HIHXCHR | -0.151 | 0.063 | -2.406 | 0.016 | -2.39683 | 0.016538 |
| LV_DEPS.ON | CHR | -0.149 | 0.032 | -4.586 | 0 | -4.65625 | 3.22E-06 |
| LV_DEPS.ON | AGE | -0.092 | 0.047 | -1.958 | 0.05 | -1.95745 | 0.050295 |
| LV_DEPS.ON | FEMALE | -0.027 | 0.05 | -0.54 | 0.589 | -0.54 | 0.589197 |
| LV_DEPS.ON | SITE02 | -0.026 | 0.093 | -0.275 | 0.784 | -0.27957 | 0.779808 |
| LV_DEPS.ON | SITE03 | 0.111 | 0.064 | 1.728 | 0.084 | 1.734375 | 0.082852 |
| LV_DEPS.ON | SITE04 | 0.041 | 0.066 | 0.625 | 0.532 | 0.621212 | 0.53446 |
| LV_DEPS.ON | SITE05 | 0.111 | 0.075 | 1.482 | 0.138 | 1.48 | 0.138873 |
| LV_DEPS.ON | SITE06 | 0.046 | 0.077 | 0.603 | 0.546 | 0.597403 | 0.550239 |
| LV_DEPS.ON | SITE07 | 0.064 | 0.072 | 0.89 | 0.373 | 0.888889 | 0.374063 |
| LV_DEPS.ON | SITE08 | 0.026 | 0.077 | 0.343 | 0.732 | 0.337662 | 0.735618 |
| LV_DEPS.ON | SITE09 | 0.161 | 0.083 | 1.937 | 0.053 | 1.939759 | 0.052409 |
| LV_HIH.WITH | CHR | -0.003 | 0.045 | -0.067 | 0.946 | -0.06667 | 0.946847 |
| LV_HIH.WITH | AGE | 0.03 | 0.06 | 0.51 | 0.61 | 0.5 | 0.617075 |
| LV_HIH.WITH | FEMALE | -0.079 | 0.058 | -1.362 | 0.173 | -1.36207 | 0.173176 |
| LV_HIH.WITH | SITE02 | 0.047 | 0.068 | 0.693 | 0.488 | 0.691176 | 0.489455 |
| LV_HIH.WITH | SITE03 | -0.009 | 0.089 | -0.103 | 0.918 | -0.10112 | 0.919452 |
| LV_HIH.WITH | SITE04 | 0.072 | 0.044 | 1.63 | 0.103 | 1.636364 | 0.101764 |
| LV_HIH.WITH | SITE05 | -0.063 | 0.062 | -1.02 | 0.308 | -1.01613 | 0.309568 |
| LV_HIH.WITH | SITE06 | 0.077 | 0.043 | 1.791 | 0.073 | 1.790698 | 0.073342 |
| LV_HIH.WITH | SITE07 | -0.01 | 0.058 | -0.181 | 0.857 | -0.17241 | 0.863112 |
| LV_HIH.WITH | SITE08 | -0.033 | 0.057 | -0.582 | 0.561 | -0.57895 | 0.562625 |
| LV_HIH.WITH | SITE09 | -0.045 | 0.058 | -0.781 | 0.435 | -0.77586 | 0.43783 |
| CHR.WITH | AGE | -0.049 | 0.051 | -0.956 | 0.339 | -0.96078 | 0.336661 |
| CHR.WITH | FEMALE | -0.032 | 0.048 | -0.67 | 0.503 | -0.66667 | 0.504985 |
| CHR.WITH | SITE02 | -0.046 | 0.053 | -0.881 | 0.378 | -0.86792 | 0.385436 |
| CHR.WITH | SITE03 | -0.034 | 0.052 | -0.645 | 0.519 | -0.65385 | 0.513211 |
| CHR.WITH | SITE04 | 0.017 | 0.046 | 0.377 | 0.706 | 0.369565 | 0.711706 |
| CHR.WITH | SITE05 | -0.001 | 0.048 | -0.022 | 0.982 | -0.02083 | 0.983379 |
| CHR.WITH | SITE06 | -0.017 | 0.049 | -0.347 | 0.728 | -0.34694 | 0.728637 |
| CHR.WITH | SITE07 | -0.004 | 0.048 | -0.087 | 0.931 | -0.08333 | 0.933586 |
| CHR.WITH | SITE08 | 0.024 | 0.045 | 0.538 | 0.591 | 0.533333 | 0.593803 |
| CHR.WITH | SITE09 | 0.008 | 0.047 | 0.17 | 0.865 | 0.170213 | 0.864843 |
| AGE.WITH | FEMALE | -0.11 | 0.048 | -2.305 | 0.021 | -2.29167 | 0.021925 |
| AGE.WITH | SITE02 | 0.304 | 0.052 | 5.894 | 0 | 5.846154 | 5.03E-09 |
| AGE.WITH | SITE03 | 0 | 0.044 | 0.003 | 0.998 | 0 | 1 |
| AGE.WITH | SITE04 | -0.013 | 0.038 | -0.343 | 0.732 | -0.34211 | 0.732272 |
| AGE.WITH | SITE05 | 0.155 | 0.043 | 3.59 | 0 | 3.604651 | 0.000313 |
| AGE.WITH | SITE06 | -0.009 | 0.04 | -0.222 | 0.824 | -0.225 | 0.821979 |
| AGE.WITH | SITE07 | -0.143 | 0.046 | -3.123 | 0.002 | -3.1087 | 0.001879 |
| AGE.WITH | SITE08 | -0.17 | 0.049 | -3.474 | 0.001 | -3.46939 | 0.000522 |
| AGE.WITH | SITE09 | -0.03 | 0.044 | -0.676 | 0.499 | -0.68182 | 0.495354 |
| FEMALE.WITH | SITE02 | 0.029 | 0.048 | 0.608 | 0.543 | 0.604167 | 0.545733 |
| FEMALE.WITH | SITE03 | -0.015 | 0.048 | -0.317 | 0.751 | -0.3125 | 0.754661 |
| FEMALE.WITH | SITE04 | -0.003 | 0.048 | -0.071 | 0.943 | -0.0625 | 0.950165 |
| FEMALE.WITH | SITE05 | 0.005 | 0.048 | 0.104 | 0.917 | 0.104167 | 0.917037 |
| FEMALE.WITH | SITE06 | -0.06 | 0.047 | -1.282 | 0.2 | -1.2766 | 0.201745 |
| FEMALE.WITH | SITE07 | 0.065 | 0.048 | 1.343 | 0.179 | 1.354167 | 0.175683 |
| FEMALE.WITH | SITE08 | 0.003 | 0.048 | 0.053 | 0.958 | 0.0625 | 0.950165 |
| FEMALE.WITH | SITE09 | 0.013 | 0.048 | 0.276 | 0.783 | 0.270833 | 0.786519 |
| SITE02.WITH | SITE03 | -0.078 | 0.01 | -7.882 | 0 | -7.8 | 6.22E-15 |
| SITE02.WITH | SITE04 | -0.096 | 0.011 | -8.758 | 0 | -8.72727 | 0 |
| SITE02.WITH | SITE05 | -0.125 | 0.013 | -9.754 | 0 | -9.61538 | 0 |
| SITE02.WITH | SITE06 | -0.119 | 0.012 | -9.584 | 0 | -9.91667 | 0 |
| SITE02.WITH | SITE07 | -0.124 | 0.013 | -9.722 | 0 | -9.53846 | 0 |
| SITE02.WITH | SITE08 | -0.106 | 0.012 | -9.176 | 0 | -8.83333 | 0 |
| SITE02.WITH | SITE09 | -0.128 | 0.013 | -9.848 | 0 | -9.84615 | 0 |
| SITE03.WITH | SITE04 | -0.078 | 0.01 | -7.882 | 0 | -7.8 | 6.22E-15 |
| SITE03.WITH | SITE05 | -0.102 | 0.012 | -8.584 | 0 | -8.5 | 0 |
| SITE03.WITH | SITE06 | -0.097 | 0.011 | -8.467 | 0 | -8.81818 | 0 |
| SITE03.WITH | SITE07 | -0.101 | 0.012 | -8.562 | 0 | -8.41667 | 0 |
| SITE03.WITH | SITE08 | -0.087 | 0.011 | -8.182 | 0 | -7.90909 | 2.66E-15 |
| SITE03.WITH | SITE09 | -0.105 | 0.012 | -8.647 | 0 | -8.75 | 0 |
| SITE04.WITH | SITE05 | -0.125 | 0.013 | -9.754 | 0 | -9.61538 | 0 |
| SITE04.WITH | SITE06 | -0.119 | 0.012 | -9.584 | 0 | -9.91667 | 0 |
| SITE04.WITH | SITE07 | -0.124 | 0.013 | -9.722 | 0 | -9.53846 | 0 |
| SITE04.WITH | SITE08 | -0.106 | 0.012 | -9.176 | 0 | -8.83333 | 0 |
| SITE04.WITH | SITE09 | -0.128 | 0.013 | -9.848 | 0 | -9.84615 | 0 |
| SITE05.WITH | SITE06 | -0.155 | 0.014 | -10.939 | 0 | -11.0714 | 0 |
| SITE05.WITH | SITE07 | -0.162 | 0.014 | -11.146 | 0 | -11.5714 | 0 |
| SITE05.WITH | SITE08 | -0.139 | 0.013 | -10.342 | 0 | -10.6923 | 0 |
| SITE05.WITH | SITE09 | -0.168 | 0.015 | -11.338 | 0 | -11.2 | 0 |
| SITE06.WITH | SITE07 | -0.154 | 0.014 | -10.893 | 0 | -11 | 0 |
| SITE06.WITH | SITE08 | -0.132 | 0.013 | -10.14 | 0 | -10.1538 | 0 |
| SITE06.WITH | SITE09 | -0.16 | 0.014 | -11.072 | 0 | -11.4286 | 0 |
| SITE07.WITH | SITE08 | -0.138 | 0.013 | -10.303 | 0 | -10.6154 | 0 |
| SITE07.WITH | SITE09 | -0.166 | 0.015 | -11.287 | 0 | -11.0667 | 0 |
| SITE08.WITH | SITE09 | -0.143 | 0.014 | -10.454 | 0 | -10.2143 | 0 |
| Means | CHR | 2.139 | 0.134 | 16.002 | 0 | 15.96269 | 0 |
| Means | AGE | 4.784 | 0.15 | 31.887 | 0 | 31.89333 | 0 |
| Means | FEMALE | 0.91 | 0.044 | 20.762 | 0 | 20.68182 | 0 |
| Means | SITE02 | 0.309 | 0.026 | 11.778 | 0 | 11.88462 | 0 |
| Means | SITE03 | 0.252 | 0.025 | 9.888 | 0 | 10.08 | 0 |
| Means | SITE04 | 0.309 | 0.026 | 11.779 | 0 | 11.88462 | 0 |
| Means | SITE05 | 0.404 | 0.028 | 14.483 | 0 | 14.42857 | 0 |
| Means | SITE06 | 0.384 | 0.028 | 13.971 | 0 | 13.71429 | 0 |
| Means | SITE07 | 0.4 | 0.028 | 14.384 | 0 | 14.28571 | 0 |
| Means | SITE08 | 0.344 | 0.027 | 12.827 | 0 | 12.74074 | 0 |
| Means | SITE09 | 0.415 | 0.028 | 14.776 | 0 | 14.82143 | 0 |
| Intercepts | DEPS_CH | 0.238 | 0.266 | 0.895 | 0.371 | 0.894737 | 0.370928 |
| Intercepts | HIH_CH | -0.051 | 0.048 | -1.072 | 0.284 | -1.0625 | 0.288009 |
| Variances | CHR | 1 | 0 |  |  |  |  |
| Variances | AGE | 1 | 0 |  |  |  |  |
| Variances | FEMALE | 1 | 0 |  |  |  |  |
| Variances | SITE02 | 1 | 0 |  |  |  |  |
| Variances | SITE03 | 1 | 0 |  |  |  |  |
| Variances | SITE04 | 1 | 0 |  |  |  |  |
| Variances | SITE05 | 1 | 0 |  |  |  |  |
| Variances | SITE06 | 1 | 0 |  |  |  |  |
| Variances | SITE07 | 1 | 0 |  |  |  |  |
| Variances | SITE08 | 1 | 0 |  |  |  |  |
| Variances | SITE09 | 1 | 0 |  |  |  |  |
| Variances | LV_HIH | 1 | 0 |  |  |  |  |
| Residual.Variances | DEPS_CH | 0.255 | 0.062 | 4.079 | 0 | 4.112903 | 3.91E-05 |
| Residual.Variances | HIH_CH | 0.331 | 0.039 | 8.413 | 0 | 8.487179 | 0 |
| Residual.Variances | LV_DEPS | 0.815 | 0.097 | 8.439 | 0 | 8.402062 | 0 |
| LV_HIL.BY | HIL_CH | 0.82 | 0.023 | 35.23 | 0 | 35.65217 | 0 |
| LV_DEPS.BY | DEPS_CH | 0.864 | 0.037 | 23.543 | 0 | 23.35135 | 0 |
| LV_DEPS.ON | LV_HIL | -0.044 | 0.035 | -1.242 | 0.214 | -1.25714 | 0.208702 |
| LV_DEPS.ON | HILXCHR | 0.008 | 0.084 | 0.095 | 0.924 | 0.095238 | 0.924126 |
| LV_DEPS.ON | CHR | -0.149 | 0.032 | -4.593 | 0 | -4.65625 | 3.22E-06 |
| LV_DEPS.ON | AGE | -0.102 | 0.051 | -2.009 | 0.045 | -2 | 0.0455 |
| LV_DEPS.ON | FEMALE | -0.001 | 0.064 | -0.023 | 0.982 | -0.01563 | 0.987534 |
| LV_DEPS.ON | SITE02 | -0.034 | 0.092 | -0.368 | 0.713 | -0.36957 | 0.711706 |
| LV_DEPS.ON | SITE03 | 0.108 | 0.065 | 1.648 | 0.099 | 1.661538 | 0.096605 |
| LV_DEPS.ON | SITE04 | 0.022 | 0.065 | 0.333 | 0.739 | 0.338462 | 0.735015 |
| LV_DEPS.ON | SITE05 | 0.129 | 0.067 | 1.922 | 0.055 | 1.925373 | 0.054183 |
| LV_DEPS.ON | SITE06 | 0.022 | 0.071 | 0.312 | 0.755 | 0.309859 | 0.756668 |
| LV_DEPS.ON | SITE07 | 0.055 | 0.068 | 0.815 | 0.415 | 0.808824 | 0.418617 |
| LV_DEPS.ON | SITE08 | 0.037 | 0.072 | 0.519 | 0.603 | 0.513889 | 0.60733 |
| LV_DEPS.ON | SITE09 | 0.17 | 0.08 | 2.12 | 0.034 | 2.125 | 0.033587 |
| LV_HIL.WITH | CHR | -0.004 | 0.049 | -0.075 | 0.941 | -0.08163 | 0.934939 |
| LV_HIL.WITH | AGE | 0.013 | 0.059 | 0.224 | 0.823 | 0.220339 | 0.825607 |
| LV_HIL.WITH | FEMALE | -0.117 | 0.061 | -1.936 | 0.053 | -1.91803 | 0.055107 |
| LV_HIL.WITH | SITE02 | -0.041 | 0.064 | -0.633 | 0.527 | -0.64063 | 0.521766 |
| LV_HIL.WITH | SITE03 | 0.095 | 0.073 | 1.299 | 0.194 | 1.30137 | 0.193132 |
| LV_HIL.WITH | SITE04 | 0.1 | 0.049 | 2.051 | 0.04 | 2.040816 | 0.041269 |
| LV_HIL.WITH | SITE05 | -0.08 | 0.066 | -1.213 | 0.225 | -1.21212 | 0.225466 |
| LV_HIL.WITH | SITE06 | 0.049 | 0.047 | 1.043 | 0.297 | 1.042553 | 0.297155 |
| LV_HIL.WITH | SITE07 | -0.086 | 0.06 | -1.448 | 0.147 | -1.43333 | 0.151763 |
| LV_HIL.WITH | SITE08 | 0.02 | 0.052 | 0.389 | 0.697 | 0.384615 | 0.700522 |
| LV_HIL.WITH | SITE09 | 0.003 | 0.055 | 0.059 | 0.953 | 0.054545 | 0.956501 |
| CHR.WITH | AGE | -0.049 | 0.051 | -0.956 | 0.339 | -0.96078 | 0.336661 |
| CHR.WITH | FEMALE | -0.032 | 0.048 | -0.67 | 0.503 | -0.66667 | 0.504985 |
| CHR.WITH | SITE02 | -0.046 | 0.053 | -0.881 | 0.378 | -0.86792 | 0.385436 |
| CHR.WITH | SITE03 | -0.034 | 0.052 | -0.645 | 0.519 | -0.65385 | 0.513211 |
| CHR.WITH | SITE04 | 0.017 | 0.046 | 0.377 | 0.706 | 0.369565 | 0.711706 |
| CHR.WITH | SITE05 | -0.001 | 0.048 | -0.022 | 0.982 | -0.02083 | 0.983379 |
| CHR.WITH | SITE06 | -0.017 | 0.049 | -0.347 | 0.728 | -0.34694 | 0.728637 |
| CHR.WITH | SITE07 | -0.004 | 0.048 | -0.087 | 0.931 | -0.08333 | 0.933586 |
| CHR.WITH | SITE08 | 0.024 | 0.045 | 0.538 | 0.591 | 0.533333 | 0.593803 |
| CHR.WITH | SITE09 | 0.008 | 0.047 | 0.17 | 0.865 | 0.170213 | 0.864843 |
| AGE.WITH | FEMALE | -0.11 | 0.048 | -2.305 | 0.021 | -2.29167 | 0.021925 |
| AGE.WITH | SITE02 | 0.304 | 0.052 | 5.894 | 0 | 5.846154 | 5.03E-09 |
| AGE.WITH | SITE03 | 0 | 0.044 | 0.003 | 0.998 | 0 | 1 |
| AGE.WITH | SITE04 | -0.013 | 0.038 | -0.343 | 0.732 | -0.34211 | 0.732272 |
| AGE.WITH | SITE05 | 0.155 | 0.043 | 3.59 | 0 | 3.604651 | 0.000313 |
| AGE.WITH | SITE06 | -0.009 | 0.04 | -0.222 | 0.824 | -0.225 | 0.821979 |
| AGE.WITH | SITE07 | -0.143 | 0.046 | -3.123 | 0.002 | -3.1087 | 0.001879 |
| AGE.WITH | SITE08 | -0.17 | 0.049 | -3.474 | 0.001 | -3.46939 | 0.000522 |
| AGE.WITH | SITE09 | -0.03 | 0.044 | -0.676 | 0.499 | -0.68182 | 0.495354 |
| FEMALE.WITH | SITE02 | 0.029 | 0.048 | 0.608 | 0.543 | 0.604167 | 0.545733 |
| FEMALE.WITH | SITE03 | -0.015 | 0.048 | -0.317 | 0.751 | -0.3125 | 0.754661 |
| FEMALE.WITH | SITE04 | -0.003 | 0.048 | -0.071 | 0.943 | -0.0625 | 0.950165 |
| FEMALE.WITH | SITE05 | 0.005 | 0.048 | 0.104 | 0.917 | 0.104167 | 0.917037 |
| FEMALE.WITH | SITE06 | -0.06 | 0.047 | -1.282 | 0.2 | -1.2766 | 0.201745 |
| FEMALE.WITH | SITE07 | 0.065 | 0.048 | 1.343 | 0.179 | 1.354167 | 0.175683 |
| FEMALE.WITH | SITE08 | 0.003 | 0.048 | 0.053 | 0.958 | 0.0625 | 0.950165 |
| FEMALE.WITH | SITE09 | 0.013 | 0.048 | 0.276 | 0.783 | 0.270833 | 0.786519 |
| SITE02.WITH | SITE03 | -0.078 | 0.01 | -7.882 | 0 | -7.8 | 6.22E-15 |
| SITE02.WITH | SITE04 | -0.096 | 0.011 | -8.758 | 0 | -8.72727 | 0 |
| SITE02.WITH | SITE05 | -0.125 | 0.013 | -9.754 | 0 | -9.61538 | 0 |
| SITE02.WITH | SITE06 | -0.119 | 0.012 | -9.584 | 0 | -9.91667 | 0 |
| SITE02.WITH | SITE07 | -0.124 | 0.013 | -9.722 | 0 | -9.53846 | 0 |
| SITE02.WITH | SITE08 | -0.106 | 0.012 | -9.176 | 0 | -8.83333 | 0 |
| SITE02.WITH | SITE09 | -0.128 | 0.013 | -9.848 | 0 | -9.84615 | 0 |
| SITE03.WITH | SITE04 | -0.078 | 0.01 | -7.882 | 0 | -7.8 | 6.22E-15 |
| SITE03.WITH | SITE05 | -0.102 | 0.012 | -8.584 | 0 | -8.5 | 0 |
| SITE03.WITH | SITE06 | -0.097 | 0.011 | -8.467 | 0 | -8.81818 | 0 |
| SITE03.WITH | SITE07 | -0.101 | 0.012 | -8.562 | 0 | -8.41667 | 0 |
| SITE03.WITH | SITE08 | -0.087 | 0.011 | -8.182 | 0 | -7.90909 | 2.66E-15 |
| SITE03.WITH | SITE09 | -0.105 | 0.012 | -8.647 | 0 | -8.75 | 0 |
| SITE04.WITH | SITE05 | -0.125 | 0.013 | -9.754 | 0 | -9.61538 | 0 |
| SITE04.WITH | SITE06 | -0.119 | 0.012 | -9.584 | 0 | -9.91667 | 0 |
| SITE04.WITH | SITE07 | -0.124 | 0.013 | -9.722 | 0 | -9.53846 | 0 |
| SITE04.WITH | SITE08 | -0.106 | 0.012 | -9.176 | 0 | -8.83333 | 0 |
| SITE04.WITH | SITE09 | -0.128 | 0.013 | -9.848 | 0 | -9.84615 | 0 |
| SITE05.WITH | SITE06 | -0.155 | 0.014 | -10.939 | 0 | -11.0714 | 0 |
| SITE05.WITH | SITE07 | -0.162 | 0.014 | -11.146 | 0 | -11.5714 | 0 |
| SITE05.WITH | SITE08 | -0.139 | 0.013 | -10.342 | 0 | -10.6923 | 0 |
| SITE05.WITH | SITE09 | -0.168 | 0.015 | -11.338 | 0 | -11.2 | 0 |
| SITE06.WITH | SITE07 | -0.154 | 0.014 | -10.893 | 0 | -11 | 0 |
| SITE06.WITH | SITE08 | -0.132 | 0.013 | -10.14 | 0 | -10.1538 | 0 |
| SITE06.WITH | SITE09 | -0.16 | 0.014 | -11.072 | 0 | -11.4286 | 0 |
| SITE07.WITH | SITE08 | -0.138 | 0.013 | -10.303 | 0 | -10.6154 | 0 |
| SITE07.WITH | SITE09 | -0.166 | 0.015 | -11.287 | 0 | -11.0667 | 0 |
| SITE08.WITH | SITE09 | -0.143 | 0.014 | -10.454 | 0 | -10.2143 | 0 |
| Means | CHR | 2.139 | 0.134 | 16.002 | 0 | 15.96269 | 0 |
| Means | AGE | 4.784 | 0.15 | 31.887 | 0 | 31.89333 | 0 |
| Means | FEMALE | 0.91 | 0.044 | 20.76 | 0 | 20.68182 | 0 |
| Means | SITE02 | 0.309 | 0.026 | 11.778 | 0 | 11.88462 | 0 |
| Means | SITE03 | 0.252 | 0.026 | 9.889 | 0 | 9.692308 | 0 |
| Means | SITE04 | 0.31 | 0.026 | 11.779 | 0 | 11.92308 | 0 |
| Means | SITE05 | 0.404 | 0.028 | 14.483 | 0 | 14.42857 | 0 |
| Means | SITE06 | 0.384 | 0.028 | 13.971 | 0 | 13.71429 | 0 |
| Means | SITE07 | 0.4 | 0.028 | 14.383 | 0 | 14.28571 | 0 |
| Means | SITE08 | 0.344 | 0.027 | 12.828 | 0 | 12.74074 | 0 |
| Means | SITE09 | 0.415 | 0.028 | 14.776 | 0 | 14.82143 | 0 |
| Intercepts | DEPS_CH | 0.268 | 0.278 | 0.964 | 0.335 | 0.964029 | 0.335031 |
| Intercepts | HIL_CH | -0.062 | 0.047 | -1.313 | 0.189 | -1.31915 | 0.187119 |
| Variances | CHR | 1 | 0 |  |  |  |  |
| Variances | AGE | 1 | 0 |  |  |  |  |
| Variances | FEMALE | 1 | 0 |  |  |  |  |
| Variances | SITE02 | 1 | 0 |  |  |  |  |
| Variances | SITE03 | 1 | 0 |  |  |  |  |
| Variances | SITE04 | 1 | 0 |  |  |  |  |
| Variances | SITE05 | 1 | 0 |  |  |  |  |
| Variances | SITE06 | 1 | 0 |  |  |  |  |
| Variances | SITE07 | 1 | 0 |  |  |  |  |
| Variances | SITE08 | 1 | 0 |  |  |  |  |
| Variances | SITE09 | 1 | 0 |  |  |  |  |
| Variances | LV_HIL | 1 | 0 |  |  |  |  |
| Residual.Variances | DEPS_CH | 0.253 | 0.063 | 3.989 | 0 | 4.015873 | 5.92E-05 |
| Residual.Variances | HIL_CH | 0.328 | 0.038 | 8.58 | 0 | 8.631579 | 0 |
| Residual.Variances | LV_DEPS | 0.929 | 0.021 | 43.938 | 0 | 44.2381 | 0 |
| LV_HIR.BY | HIR_CH | 0.83 | 0.029 | 28.965 | 0 | 28.62069 | 0 |
| LV_DEPS.BY | DEPS_CH | 0.863 | 0.036 | 23.734 | 0 | 23.97222 | 0 |
| LV_DEPS.ON | LV_HIR | -0.011 | 0.042 | -0.259 | 0.795 | -0.2619 | 0.793395 |
| LV_DEPS.ON | HIRXCHR | -0.149 | 0.048 | -3.102 | 0.002 | -3.10417 | 0.001908 |
| LV_DEPS.ON | CHR | -0.149 | 0.032 | -4.621 | 0 | -4.65625 | 3.22E-06 |
| LV_DEPS.ON | AGE | -0.102 | 0.048 | -2.152 | 0.031 | -2.125 | 0.033587 |
| LV_DEPS.ON | FEMALE | -0.005 | 0.051 | -0.104 | 0.918 | -0.09804 | 0.921901 |
| LV_DEPS.ON | SITE02 | -0.048 | 0.093 | -0.516 | 0.606 | -0.51613 | 0.605764 |
| LV_DEPS.ON | SITE03 | 0.073 | 0.059 | 1.252 | 0.211 | 1.237288 | 0.21598 |
| LV_DEPS.ON | SITE04 | -0.006 | 0.063 | -0.089 | 0.929 | -0.09524 | 0.924126 |
| LV_DEPS.ON | SITE05 | 0.119 | 0.069 | 1.742 | 0.082 | 1.724638 | 0.084593 |
| LV_DEPS.ON | SITE06 | 0.023 | 0.071 | 0.323 | 0.747 | 0.323944 | 0.745981 |
| LV_DEPS.ON | SITE07 | 0.073 | 0.067 | 1.097 | 0.273 | 1.089552 | 0.27591 |
| LV_DEPS.ON | SITE08 | 0.03 | 0.073 | 0.411 | 0.681 | 0.410959 | 0.681103 |
| LV_DEPS.ON | SITE09 | 0.15 | 0.079 | 1.897 | 0.058 | 1.898734 | 0.057599 |
| LV_HIR.WITH | CHR | 0.014 | 0.045 | 0.308 | 0.758 | 0.311111 | 0.755716 |
| LV_HIR.WITH | AGE | -0.017 | 0.055 | -0.31 | 0.757 | -0.30909 | 0.757252 |
| LV_HIR.WITH | FEMALE | -0.009 | 0.058 | -0.159 | 0.873 | -0.15517 | 0.876685 |
| LV_HIR.WITH | SITE02 | -0.012 | 0.065 | -0.178 | 0.859 | -0.18462 | 0.853531 |
| LV_HIR.WITH | SITE03 | -0.1 | 0.073 | -1.371 | 0.17 | -1.36986 | 0.17073 |
| LV_HIR.WITH | SITE04 | -0.039 | 0.058 | -0.671 | 0.502 | -0.67241 | 0.50132 |
| LV_HIR.WITH | SITE05 | 0.004 | 0.05 | 0.077 | 0.939 | 0.08 | 0.936237 |
| LV_HIR.WITH | SITE06 | 0.031 | 0.046 | 0.659 | 0.51 | 0.673913 | 0.500367 |
| LV_HIR.WITH | SITE07 | 0.067 | 0.07 | 0.966 | 0.334 | 0.957143 | 0.338495 |
| LV_HIR.WITH | SITE08 | 0.034 | 0.053 | 0.634 | 0.526 | 0.641509 | 0.521192 |
| LV_HIR.WITH | SITE09 | -0.041 | 0.055 | -0.751 | 0.452 | -0.74545 | 0.455997 |
| CHR.WITH | AGE | -0.049 | 0.051 | -0.956 | 0.339 | -0.96078 | 0.336661 |
| CHR.WITH | FEMALE | -0.032 | 0.048 | -0.67 | 0.503 | -0.66667 | 0.504985 |
| CHR.WITH | SITE02 | -0.046 | 0.053 | -0.881 | 0.378 | -0.86792 | 0.385436 |
| CHR.WITH | SITE03 | -0.034 | 0.052 | -0.645 | 0.519 | -0.65385 | 0.513211 |
| CHR.WITH | SITE04 | 0.017 | 0.046 | 0.377 | 0.706 | 0.369565 | 0.711706 |
| CHR.WITH | SITE05 | -0.001 | 0.048 | -0.022 | 0.982 | -0.02083 | 0.983379 |
| CHR.WITH | SITE06 | -0.017 | 0.049 | -0.347 | 0.728 | -0.34694 | 0.728637 |
| CHR.WITH | SITE07 | -0.004 | 0.048 | -0.087 | 0.931 | -0.08333 | 0.933586 |
| CHR.WITH | SITE08 | 0.024 | 0.045 | 0.538 | 0.591 | 0.533333 | 0.593803 |
| CHR.WITH | SITE09 | 0.008 | 0.047 | 0.17 | 0.865 | 0.170213 | 0.864843 |
| AGE.WITH | FEMALE | -0.11 | 0.048 | -2.305 | 0.021 | -2.29167 | 0.021925 |
| AGE.WITH | SITE02 | 0.304 | 0.052 | 5.894 | 0 | 5.846154 | 5.03E-09 |
| AGE.WITH | SITE03 | 0 | 0.044 | 0.003 | 0.998 | 0 | 1 |
| AGE.WITH | SITE04 | -0.013 | 0.038 | -0.343 | 0.732 | -0.34211 | 0.732272 |
| AGE.WITH | SITE05 | 0.155 | 0.043 | 3.59 | 0 | 3.604651 | 0.000313 |
| AGE.WITH | SITE06 | -0.009 | 0.04 | -0.222 | 0.824 | -0.225 | 0.821979 |
| AGE.WITH | SITE07 | -0.143 | 0.046 | -3.123 | 0.002 | -3.1087 | 0.001879 |
| AGE.WITH | SITE08 | -0.17 | 0.049 | -3.474 | 0.001 | -3.46939 | 0.000522 |
| AGE.WITH | SITE09 | -0.03 | 0.044 | -0.676 | 0.499 | -0.68182 | 0.495354 |
| FEMALE.WITH | SITE02 | 0.029 | 0.048 | 0.608 | 0.543 | 0.604167 | 0.545733 |
| FEMALE.WITH | SITE03 | -0.015 | 0.048 | -0.317 | 0.751 | -0.3125 | 0.754661 |
| FEMALE.WITH | SITE04 | -0.003 | 0.048 | -0.071 | 0.943 | -0.0625 | 0.950165 |
| FEMALE.WITH | SITE05 | 0.005 | 0.048 | 0.104 | 0.917 | 0.104167 | 0.917037 |
| FEMALE.WITH | SITE06 | -0.06 | 0.047 | -1.282 | 0.2 | -1.2766 | 0.201745 |
| FEMALE.WITH | SITE07 | 0.065 | 0.048 | 1.343 | 0.179 | 1.354167 | 0.175683 |
| FEMALE.WITH | SITE08 | 0.003 | 0.048 | 0.053 | 0.958 | 0.0625 | 0.950165 |
| FEMALE.WITH | SITE09 | 0.013 | 0.048 | 0.276 | 0.783 | 0.270833 | 0.786519 |
| SITE02.WITH | SITE03 | -0.078 | 0.01 | -7.882 | 0 | -7.8 | 6.22E-15 |
| SITE02.WITH | SITE04 | -0.096 | 0.011 | -8.758 | 0 | -8.72727 | 0 |
| SITE02.WITH | SITE05 | -0.125 | 0.013 | -9.754 | 0 | -9.61538 | 0 |
| SITE02.WITH | SITE06 | -0.119 | 0.012 | -9.584 | 0 | -9.91667 | 0 |
| SITE02.WITH | SITE07 | -0.124 | 0.013 | -9.722 | 0 | -9.53846 | 0 |
| SITE02.WITH | SITE08 | -0.106 | 0.012 | -9.176 | 0 | -8.83333 | 0 |
| SITE02.WITH | SITE09 | -0.128 | 0.013 | -9.848 | 0 | -9.84615 | 0 |
| SITE03.WITH | SITE04 | -0.078 | 0.01 | -7.882 | 0 | -7.8 | 6.22E-15 |
| SITE03.WITH | SITE05 | -0.102 | 0.012 | -8.584 | 0 | -8.5 | 0 |
| SITE03.WITH | SITE06 | -0.097 | 0.011 | -8.467 | 0 | -8.81818 | 0 |
| SITE03.WITH | SITE07 | -0.101 | 0.012 | -8.562 | 0 | -8.41667 | 0 |
| SITE03.WITH | SITE08 | -0.087 | 0.011 | -8.182 | 0 | -7.90909 | 2.66E-15 |
| SITE03.WITH | SITE09 | -0.105 | 0.012 | -8.647 | 0 | -8.75 | 0 |
| SITE04.WITH | SITE05 | -0.125 | 0.013 | -9.754 | 0 | -9.61538 | 0 |
| SITE04.WITH | SITE06 | -0.119 | 0.012 | -9.584 | 0 | -9.91667 | 0 |
| SITE04.WITH | SITE07 | -0.124 | 0.013 | -9.722 | 0 | -9.53846 | 0 |
| SITE04.WITH | SITE08 | -0.106 | 0.012 | -9.176 | 0 | -8.83333 | 0 |
| SITE04.WITH | SITE09 | -0.128 | 0.013 | -9.848 | 0 | -9.84615 | 0 |
| SITE05.WITH | SITE06 | -0.155 | 0.014 | -10.939 | 0 | -11.0714 | 0 |
| SITE05.WITH | SITE07 | -0.162 | 0.014 | -11.146 | 0 | -11.5714 | 0 |
| SITE05.WITH | SITE08 | -0.139 | 0.013 | -10.342 | 0 | -10.6923 | 0 |
| SITE05.WITH | SITE09 | -0.168 | 0.015 | -11.338 | 0 | -11.2 | 0 |
| SITE06.WITH | SITE07 | -0.154 | 0.014 | -10.893 | 0 | -11 | 0 |
| SITE06.WITH | SITE08 | -0.132 | 0.013 | -10.14 | 0 | -10.1538 | 0 |
| SITE06.WITH | SITE09 | -0.16 | 0.014 | -11.072 | 0 | -11.4286 | 0 |
| SITE07.WITH | SITE08 | -0.138 | 0.013 | -10.303 | 0 | -10.6154 | 0 |
| SITE07.WITH | SITE09 | -0.166 | 0.015 | -11.287 | 0 | -11.0667 | 0 |
| SITE08.WITH | SITE09 | -0.143 | 0.014 | -10.454 | 0 | -10.2143 | 0 |
| Means | CHR | 2.139 | 0.134 | 16.002 | 0 | 15.96269 | 0 |
| Means | AGE | 4.784 | 0.15 | 31.887 | 0 | 31.89333 | 0 |
| Means | FEMALE | 0.91 | 0.044 | 20.763 | 0 | 20.68182 | 0 |
| Means | SITE02 | 0.309 | 0.026 | 11.778 | 0 | 11.88462 | 0 |
| Means | SITE03 | 0.252 | 0.025 | 9.888 | 0 | 10.08 | 0 |
| Means | SITE04 | 0.309 | 0.026 | 11.778 | 0 | 11.88462 | 0 |
| Means | SITE05 | 0.404 | 0.028 | 14.484 | 0 | 14.42857 | 0 |
| Means | SITE06 | 0.384 | 0.028 | 13.971 | 0 | 13.71429 | 0 |
| Means | SITE07 | 0.4 | 0.028 | 14.385 | 0 | 14.28571 | 0 |
| Means | SITE08 | 0.344 | 0.027 | 12.828 | 0 | 12.74074 | 0 |
| Means | SITE09 | 0.415 | 0.028 | 14.775 | 0 | 14.82143 | 0 |
| Intercepts | DEPS_CH | 0.299 | 0.265 | 1.128 | 0.259 | 1.128302 | 0.259192 |
| Intercepts | HIR_CH | 0.013 | 0.048 | 0.27 | 0.787 | 0.270833 | 0.786519 |
| Variances | CHR | 1 | 0 |  |  |  |  |
| Variances | AGE | 1 | 0 |  |  |  |  |
| Variances | FEMALE | 1 | 0 |  |  |  |  |
| Variances | SITE02 | 1 | 0 |  |  |  |  |
| Variances | SITE03 | 1 | 0 |  |  |  |  |
| Variances | SITE04 | 1 | 0 |  |  |  |  |
| Variances | SITE05 | 1 | 0 |  |  |  |  |
| Variances | SITE06 | 1 | 0 |  |  |  |  |
| Variances | SITE07 | 1 | 0 |  |  |  |  |
| Variances | SITE08 | 1 | 0 |  |  |  |  |
| Variances | SITE09 | 1 | 0 |  |  |  |  |
| Variances | LV_HIR | 1 | 0 |  |  |  |  |
| Residual.Variances | DEPS_CH | 0.254 | 0.063 | 4.051 | 0 | 4.031746 | 5.54E-05 |
| Residual.Variances | HIR_CH | 0.311 | 0.048 | 6.53 | 0 | 6.479167 | 9.22E-11 |
| Residual.Variances | LV_DEPS | 0.799 | 0.076 | 10.524 | 0 | 10.51316 | 0 |
| LV_HIH.BY | HIH_CH | 0.818 | 0.024 | 33.83 | 0 | 34.08333 | 0 |
| LV_GAF.BY | GAF_CH | 0.91 | 0.013 | 67.482 | 0 | 70 | 0 |
| LV_GAF.ON | LV_HIH | -0.052 | 0.056 | -0.935 | 0.35 | -0.92857 | 0.353111 |
| LV_GAF.ON | HIHXCHR | 0.139 | 0.045 | 3.084 | 0.002 | 3.088889 | 0.002009 |
| LV_GAF.ON | CHR | 0.125 | 0.035 | 3.549 | 0 | 3.571429 | 0.000355 |
| LV_GAF.ON | AGE | -0.104 | 0.052 | -1.988 | 0.047 | -2 | 0.0455 |
| LV_GAF.ON | FEMALE | -0.009 | 0.052 | -0.178 | 0.859 | -0.17308 | 0.862591 |
| LV_GAF.ON | SITE02 | -0.083 | 0.069 | -1.207 | 0.227 | -1.2029 | 0.229016 |
| LV_GAF.ON | SITE03 | -0.107 | 0.055 | -1.946 | 0.052 | -1.94545 | 0.05172 |
| LV_GAF.ON | SITE04 | -0.044 | 0.06 | -0.73 | 0.465 | -0.73333 | 0.463355 |
| LV_GAF.ON | SITE05 | -0.061 | 0.074 | -0.824 | 0.41 | -0.82432 | 0.409755 |
| LV_GAF.ON | SITE06 | -0.136 | 0.072 | -1.892 | 0.059 | -1.88889 | 0.058907 |
| LV_GAF.ON | SITE07 | -0.138 | 0.089 | -1.552 | 0.121 | -1.55056 | 0.121007 |
| LV_GAF.ON | SITE08 | -0.008 | 0.09 | -0.094 | 0.925 | -0.08889 | 0.92917 |
| LV_GAF.ON | SITE09 | -0.159 | 0.079 | -2.004 | 0.045 | -2.01266 | 0.044151 |
| LV_HIH.WITH | CHR | -0.002 | 0.045 | -0.054 | 0.957 | -0.04444 | 0.96455 |
| LV_HIH.WITH | AGE | 0.028 | 0.059 | 0.47 | 0.638 | 0.474576 | 0.635089 |
| LV_HIH.WITH | FEMALE | -0.084 | 0.058 | -1.437 | 0.151 | -1.44828 | 0.14754 |
| LV_HIH.WITH | SITE02 | 0.044 | 0.066 | 0.678 | 0.498 | 0.666667 | 0.504985 |
| LV_HIH.WITH | SITE03 | -0.008 | 0.089 | -0.095 | 0.924 | -0.08989 | 0.928377 |
| LV_HIH.WITH | SITE04 | 0.071 | 0.044 | 1.615 | 0.106 | 1.613636 | 0.106606 |
| LV_HIH.WITH | SITE05 | -0.064 | 0.062 | -1.036 | 0.3 | -1.03226 | 0.301951 |
| LV_HIH.WITH | SITE06 | 0.072 | 0.043 | 1.664 | 0.096 | 1.674419 | 0.094048 |
| LV_HIH.WITH | SITE07 | -0.008 | 0.058 | -0.134 | 0.894 | -0.13793 | 0.890295 |
| LV_HIH.WITH | SITE08 | -0.033 | 0.058 | -0.568 | 0.57 | -0.56897 | 0.56938 |
| LV_HIH.WITH | SITE09 | -0.045 | 0.058 | -0.775 | 0.438 | -0.77586 | 0.43783 |
| CHR.WITH | AGE | -0.048 | 0.051 | -0.942 | 0.346 | -0.94118 | 0.346614 |
| CHR.WITH | FEMALE | -0.031 | 0.048 | -0.649 | 0.516 | -0.64583 | 0.518387 |
| CHR.WITH | SITE02 | -0.046 | 0.053 | -0.875 | 0.382 | -0.86792 | 0.385436 |
| CHR.WITH | SITE03 | -0.034 | 0.052 | -0.64 | 0.522 | -0.65385 | 0.513211 |
| CHR.WITH | SITE04 | 0.018 | 0.046 | 0.384 | 0.701 | 0.391304 | 0.695572 |
| CHR.WITH | SITE05 | -0.001 | 0.048 | -0.013 | 0.989 | -0.02083 | 0.983379 |
| CHR.WITH | SITE06 | -0.017 | 0.049 | -0.339 | 0.735 | -0.34694 | 0.728637 |
| CHR.WITH | SITE07 | -0.004 | 0.048 | -0.078 | 0.938 | -0.08333 | 0.933586 |
| CHR.WITH | SITE08 | 0.025 | 0.045 | 0.546 | 0.585 | 0.555556 | 0.578515 |
| CHR.WITH | SITE09 | 0.009 | 0.047 | 0.18 | 0.857 | 0.191489 | 0.848142 |
| AGE.WITH | FEMALE | -0.111 | 0.048 | -2.336 | 0.02 | -2.3125 | 0.02075 |
| AGE.WITH | SITE02 | 0.304 | 0.052 | 5.886 | 0 | 5.846154 | 5.03E-09 |
| AGE.WITH | SITE03 | 0 | 0.044 | -0.006 | 0.995 | 0 | 1 |
| AGE.WITH | SITE04 | -0.014 | 0.038 | -0.355 | 0.722 | -0.36842 | 0.712559 |
| AGE.WITH | SITE05 | 0.155 | 0.043 | 3.577 | 0 | 3.604651 | 0.000313 |
| AGE.WITH | SITE06 | -0.01 | 0.04 | -0.237 | 0.813 | -0.25 | 0.802587 |
| AGE.WITH | SITE07 | -0.143 | 0.046 | -3.137 | 0.002 | -3.1087 | 0.001879 |
| AGE.WITH | SITE08 | -0.171 | 0.049 | -3.485 | 0 | -3.4898 | 0.000483 |
| AGE.WITH | SITE09 | -0.03 | 0.044 | -0.691 | 0.49 | -0.68182 | 0.495354 |
| FEMALE.WITH | SITE02 | 0.029 | 0.048 | 0.595 | 0.552 | 0.604167 | 0.545733 |
| FEMALE.WITH | SITE03 | -0.016 | 0.048 | -0.328 | 0.743 | -0.33333 | 0.738883 |
| FEMALE.WITH | SITE04 | -0.004 | 0.048 | -0.085 | 0.932 | -0.08333 | 0.933586 |
| FEMALE.WITH | SITE05 | 0.004 | 0.048 | 0.086 | 0.931 | 0.083333 | 0.933586 |
| FEMALE.WITH | SITE06 | -0.061 | 0.047 | -1.299 | 0.194 | -1.29787 | 0.194331 |
| FEMALE.WITH | SITE07 | 0.064 | 0.048 | 1.326 | 0.185 | 1.333333 | 0.182422 |
| FEMALE.WITH | SITE08 | 0.002 | 0.048 | 0.038 | 0.97 | 0.041667 | 0.966764 |
| FEMALE.WITH | SITE09 | 0.012 | 0.048 | 0.258 | 0.797 | 0.25 | 0.802587 |
| SITE02.WITH | SITE03 | -0.078 | 0.01 | -7.882 | 0 | -7.8 | 6.22E-15 |
| SITE02.WITH | SITE04 | -0.096 | 0.011 | -8.758 | 0 | -8.72727 | 0 |
| SITE02.WITH | SITE05 | -0.125 | 0.013 | -9.754 | 0 | -9.61538 | 0 |
| SITE02.WITH | SITE06 | -0.119 | 0.012 | -9.584 | 0 | -9.91667 | 0 |
| SITE02.WITH | SITE07 | -0.124 | 0.013 | -9.722 | 0 | -9.53846 | 0 |
| SITE02.WITH | SITE08 | -0.107 | 0.012 | -9.176 | 0 | -8.91667 | 0 |
| SITE02.WITH | SITE09 | -0.129 | 0.013 | -9.848 | 0 | -9.92308 | 0 |
| SITE03.WITH | SITE04 | -0.078 | 0.01 | -7.882 | 0 | -7.8 | 6.22E-15 |
| SITE03.WITH | SITE05 | -0.102 | 0.012 | -8.584 | 0 | -8.5 | 0 |
| SITE03.WITH | SITE06 | -0.097 | 0.011 | -8.468 | 0 | -8.81818 | 0 |
| SITE03.WITH | SITE07 | -0.101 | 0.012 | -8.562 | 0 | -8.41667 | 0 |
| SITE03.WITH | SITE08 | -0.087 | 0.011 | -8.182 | 0 | -7.90909 | 2.66E-15 |
| SITE03.WITH | SITE09 | -0.105 | 0.012 | -8.648 | 0 | -8.75 | 0 |
| SITE04.WITH | SITE05 | -0.125 | 0.013 | -9.754 | 0 | -9.61538 | 0 |
| SITE04.WITH | SITE06 | -0.119 | 0.012 | -9.584 | 0 | -9.91667 | 0 |
| SITE04.WITH | SITE07 | -0.124 | 0.013 | -9.722 | 0 | -9.53846 | 0 |
| SITE04.WITH | SITE08 | -0.107 | 0.012 | -9.176 | 0 | -8.91667 | 0 |
| SITE04.WITH | SITE09 | -0.129 | 0.013 | -9.848 | 0 | -9.92308 | 0 |
| SITE05.WITH | SITE06 | -0.156 | 0.014 | -10.94 | 0 | -11.1429 | 0 |
| SITE05.WITH | SITE07 | -0.162 | 0.015 | -11.147 | 0 | -10.8 | 0 |
| SITE05.WITH | SITE08 | -0.139 | 0.013 | -10.343 | 0 | -10.6923 | 0 |
| SITE05.WITH | SITE09 | -0.168 | 0.015 | -11.339 | 0 | -11.2 | 0 |
| SITE06.WITH | SITE07 | -0.154 | 0.014 | -10.894 | 0 | -11 | 0 |
| SITE06.WITH | SITE08 | -0.133 | 0.013 | -10.14 | 0 | -10.2308 | 0 |
| SITE06.WITH | SITE09 | -0.16 | 0.014 | -11.073 | 0 | -11.4286 | 0 |
| SITE07.WITH | SITE08 | -0.138 | 0.013 | -10.304 | 0 | -10.6154 | 0 |
| SITE07.WITH | SITE09 | -0.167 | 0.015 | -11.287 | 0 | -11.1333 | 0 |
| SITE08.WITH | SITE09 | -0.143 | 0.014 | -10.454 | 0 | -10.2143 | 0 |
| Means | CHR | 2.136 | 0.134 | 15.998 | 0 | 15.9403 | 0 |
| Means | AGE | 4.782 | 0.15 | 31.873 | 0 | 31.88 | 0 |
| Means | FEMALE | 0.912 | 0.044 | 20.746 | 0 | 20.72727 | 0 |
| Means | SITE02 | 0.31 | 0.026 | 11.776 | 0 | 11.92308 | 0 |
| Means | SITE03 | 0.252 | 0.026 | 9.888 | 0 | 9.692308 | 0 |
| Means | SITE04 | 0.31 | 0.026 | 11.776 | 0 | 11.92308 | 0 |
| Means | SITE05 | 0.404 | 0.028 | 14.482 | 0 | 14.42857 | 0 |
| Means | SITE06 | 0.385 | 0.028 | 13.967 | 0 | 13.75 | 0 |
| Means | SITE07 | 0.401 | 0.028 | 14.381 | 0 | 14.32143 | 0 |
| Means | SITE08 | 0.344 | 0.027 | 12.826 | 0 | 12.74074 | 0 |
| Means | SITE09 | 0.416 | 0.028 | 14.774 | 0 | 14.85714 | 0 |
| Intercepts | GAF_CH | 0.723 | 0.278 | 2.601 | 0.009 | 2.600719 | 0.009303 |
| Intercepts | HIH_CH | -0.052 | 0.048 | -1.09 | 0.276 | -1.08333 | 0.27866 |
| Variances | CHR | 1 | 0 |  |  |  |  |
| Variances | AGE | 1 | 0 |  |  |  |  |
| Variances | FEMALE | 1 | 0 |  |  |  |  |
| Variances | SITE02 | 1 | 0 |  |  |  |  |
| Variances | SITE03 | 1 | 0 |  |  |  |  |
| Variances | SITE04 | 1 | 0 |  |  |  |  |
| Variances | SITE05 | 1 | 0 |  |  |  |  |
| Variances | SITE06 | 1 | 0 |  |  |  |  |
| Variances | SITE07 | 1 | 0 |  |  |  |  |
| Variances | SITE08 | 1 | 0 |  |  |  |  |
| Variances | SITE09 | 1 | 0 |  |  |  |  |
| Variances | LV_HIH | 1 | 0 |  |  |  |  |
| Residual.Variances | GAF_CH | 0.172 | 0.025 | 7.032 | 0 | 6.88 | 5.99E-12 |
| Residual.Variances | HIH_CH | 0.331 | 0.04 | 8.382 | 0 | 8.275 | 2.22E-16 |
| Residual.Variances | LV_GAF | 0.864 | 0.052 | 16.668 | 0 | 16.61538 | 0 |
| LV_HIL.BY | HIL_CH | 0.819 | 0.023 | 34.968 | 0 | 35.6087 | 0 |
| LV_GAF.BY | GAF_CH | 0.91 | 0.014 | 67.308 | 0 | 65 | 0 |
| LV_GAF.ON | LV_HIL | 0.022 | 0.083 | 0.263 | 0.792 | 0.26506 | 0.790963 |
| LV_GAF.ON | HILXCHR | 0.108 | 0.053 | 2.02 | 0.043 | 2.037736 | 0.041576 |
| LV_GAF.ON | CHR | 0.125 | 0.035 | 3.555 | 0 | 3.571429 | 0.000355 |
| LV_GAF.ON | AGE | -0.106 | 0.055 | -1.917 | 0.055 | -1.92727 | 0.053946 |
| LV_GAF.ON | FEMALE | -0.005 | 0.053 | -0.101 | 0.92 | -0.09434 | 0.924839 |
| LV_GAF.ON | SITE02 | -0.068 | 0.072 | -0.949 | 0.343 | -0.94444 | 0.344943 |
| LV_GAF.ON | SITE03 | -0.137 | 0.057 | -2.39 | 0.017 | -2.40351 | 0.016239 |
| LV_GAF.ON | SITE04 | -0.055 | 0.062 | -0.888 | 0.375 | -0.8871 | 0.375027 |
| LV_GAF.ON | SITE05 | -0.056 | 0.075 | -0.754 | 0.451 | -0.74667 | 0.455265 |
| LV_GAF.ON | SITE06 | -0.133 | 0.072 | -1.847 | 0.065 | -1.84722 | 0.064715 |
| LV_GAF.ON | SITE07 | -0.119 | 0.091 | -1.304 | 0.192 | -1.30769 | 0.190978 |
| LV_GAF.ON | SITE08 | -0.027 | 0.088 | -0.307 | 0.759 | -0.30682 | 0.758982 |
| LV_GAF.ON | SITE09 | -0.167 | 0.078 | -2.135 | 0.033 | -2.14103 | 0.032272 |
| LV_HIL.WITH | CHR | -0.004 | 0.049 | -0.075 | 0.94 | -0.08163 | 0.934939 |
| LV_HIL.WITH | AGE | 0.014 | 0.058 | 0.234 | 0.815 | 0.241379 | 0.809261 |
| LV_HIL.WITH | FEMALE | -0.119 | 0.061 | -1.959 | 0.05 | -1.95082 | 0.051079 |
| LV_HIL.WITH | SITE02 | -0.037 | 0.062 | -0.589 | 0.556 | -0.59677 | 0.550658 |
| LV_HIL.WITH | SITE03 | 0.092 | 0.073 | 1.269 | 0.204 | 1.260274 | 0.207571 |
| LV_HIL.WITH | SITE04 | 0.1 | 0.049 | 2.06 | 0.039 | 2.040816 | 0.041269 |
| LV_HIL.WITH | SITE05 | -0.081 | 0.065 | -1.242 | 0.214 | -1.24615 | 0.212708 |
| LV_HIL.WITH | SITE06 | 0.047 | 0.047 | 0.984 | 0.325 | 1 | 0.317311 |
| LV_HIL.WITH | SITE07 | -0.084 | 0.06 | -1.41 | 0.159 | -1.4 | 0.161513 |
| LV_HIL.WITH | SITE08 | 0.021 | 0.052 | 0.408 | 0.683 | 0.403846 | 0.686326 |
| LV_HIL.WITH | SITE09 | 0.001 | 0.055 | 0.023 | 0.982 | 0.018182 | 0.985494 |
| CHR.WITH | AGE | -0.048 | 0.051 | -0.942 | 0.346 | -0.94118 | 0.346614 |
| CHR.WITH | FEMALE | -0.031 | 0.048 | -0.649 | 0.516 | -0.64583 | 0.518387 |
| CHR.WITH | SITE02 | -0.046 | 0.053 | -0.875 | 0.382 | -0.86792 | 0.385436 |
| CHR.WITH | SITE03 | -0.034 | 0.052 | -0.64 | 0.522 | -0.65385 | 0.513211 |
| CHR.WITH | SITE04 | 0.018 | 0.046 | 0.384 | 0.701 | 0.391304 | 0.695572 |
| CHR.WITH | SITE05 | -0.001 | 0.048 | -0.013 | 0.989 | -0.02083 | 0.983379 |
| CHR.WITH | SITE06 | -0.017 | 0.049 | -0.339 | 0.735 | -0.34694 | 0.728637 |
| CHR.WITH | SITE07 | -0.004 | 0.048 | -0.078 | 0.938 | -0.08333 | 0.933586 |
| CHR.WITH | SITE08 | 0.025 | 0.045 | 0.546 | 0.585 | 0.555556 | 0.578515 |
| CHR.WITH | SITE09 | 0.009 | 0.047 | 0.18 | 0.857 | 0.191489 | 0.848142 |
| AGE.WITH | FEMALE | -0.111 | 0.048 | -2.336 | 0.02 | -2.3125 | 0.02075 |
| AGE.WITH | SITE02 | 0.304 | 0.052 | 5.886 | 0 | 5.846154 | 5.03E-09 |
| AGE.WITH | SITE03 | 0 | 0.044 | -0.006 | 0.995 | 0 | 1 |
| AGE.WITH | SITE04 | -0.014 | 0.038 | -0.355 | 0.722 | -0.36842 | 0.712559 |
| AGE.WITH | SITE05 | 0.155 | 0.043 | 3.577 | 0 | 3.604651 | 0.000313 |
| AGE.WITH | SITE06 | -0.01 | 0.04 | -0.237 | 0.813 | -0.25 | 0.802587 |
| AGE.WITH | SITE07 | -0.143 | 0.046 | -3.137 | 0.002 | -3.1087 | 0.001879 |
| AGE.WITH | SITE08 | -0.171 | 0.049 | -3.485 | 0 | -3.4898 | 0.000483 |
| AGE.WITH | SITE09 | -0.03 | 0.044 | -0.691 | 0.49 | -0.68182 | 0.495354 |
| FEMALE.WITH | SITE02 | 0.029 | 0.048 | 0.595 | 0.552 | 0.604167 | 0.545733 |
| FEMALE.WITH | SITE03 | -0.016 | 0.048 | -0.328 | 0.743 | -0.33333 | 0.738883 |
| FEMALE.WITH | SITE04 | -0.004 | 0.048 | -0.085 | 0.932 | -0.08333 | 0.933586 |
| FEMALE.WITH | SITE05 | 0.004 | 0.048 | 0.086 | 0.931 | 0.083333 | 0.933586 |
| FEMALE.WITH | SITE06 | -0.061 | 0.047 | -1.299 | 0.194 | -1.29787 | 0.194331 |
| FEMALE.WITH | SITE07 | 0.064 | 0.048 | 1.326 | 0.185 | 1.333333 | 0.182422 |
| FEMALE.WITH | SITE08 | 0.002 | 0.048 | 0.038 | 0.97 | 0.041667 | 0.966764 |
| FEMALE.WITH | SITE09 | 0.012 | 0.048 | 0.258 | 0.797 | 0.25 | 0.802587 |
| SITE02.WITH | SITE03 | -0.078 | 0.01 | -7.882 | 0 | -7.8 | 6.22E-15 |
| SITE02.WITH | SITE04 | -0.096 | 0.011 | -8.758 | 0 | -8.72727 | 0 |
| SITE02.WITH | SITE05 | -0.125 | 0.013 | -9.754 | 0 | -9.61538 | 0 |
| SITE02.WITH | SITE06 | -0.119 | 0.012 | -9.584 | 0 | -9.91667 | 0 |
| SITE02.WITH | SITE07 | -0.124 | 0.013 | -9.722 | 0 | -9.53846 | 0 |
| SITE02.WITH | SITE08 | -0.107 | 0.012 | -9.176 | 0 | -8.91667 | 0 |
| SITE02.WITH | SITE09 | -0.129 | 0.013 | -9.848 | 0 | -9.92308 | 0 |
| SITE03.WITH | SITE04 | -0.078 | 0.01 | -7.882 | 0 | -7.8 | 6.22E-15 |
| SITE03.WITH | SITE05 | -0.102 | 0.012 | -8.584 | 0 | -8.5 | 0 |
| SITE03.WITH | SITE06 | -0.097 | 0.011 | -8.468 | 0 | -8.81818 | 0 |
| SITE03.WITH | SITE07 | -0.101 | 0.012 | -8.562 | 0 | -8.41667 | 0 |
| SITE03.WITH | SITE08 | -0.087 | 0.011 | -8.182 | 0 | -7.90909 | 2.66E-15 |
| SITE03.WITH | SITE09 | -0.105 | 0.012 | -8.648 | 0 | -8.75 | 0 |
| SITE04.WITH | SITE05 | -0.125 | 0.013 | -9.754 | 0 | -9.61538 | 0 |
| SITE04.WITH | SITE06 | -0.119 | 0.012 | -9.584 | 0 | -9.91667 | 0 |
| SITE04.WITH | SITE07 | -0.124 | 0.013 | -9.722 | 0 | -9.53846 | 0 |
| SITE04.WITH | SITE08 | -0.107 | 0.012 | -9.176 | 0 | -8.91667 | 0 |
| SITE04.WITH | SITE09 | -0.129 | 0.013 | -9.848 | 0 | -9.92308 | 0 |
| SITE05.WITH | SITE06 | -0.156 | 0.014 | -10.94 | 0 | -11.1429 | 0 |
| SITE05.WITH | SITE07 | -0.162 | 0.015 | -11.147 | 0 | -10.8 | 0 |
| SITE05.WITH | SITE08 | -0.139 | 0.013 | -10.343 | 0 | -10.6923 | 0 |
| SITE05.WITH | SITE09 | -0.168 | 0.015 | -11.339 | 0 | -11.2 | 0 |
| SITE06.WITH | SITE07 | -0.154 | 0.014 | -10.894 | 0 | -11 | 0 |
| SITE06.WITH | SITE08 | -0.133 | 0.013 | -10.14 | 0 | -10.2308 | 0 |
| SITE06.WITH | SITE09 | -0.16 | 0.014 | -11.073 | 0 | -11.4286 | 0 |
| SITE07.WITH | SITE08 | -0.138 | 0.013 | -10.304 | 0 | -10.6154 | 0 |
| SITE07.WITH | SITE09 | -0.167 | 0.015 | -11.287 | 0 | -11.1333 | 0 |
| SITE08.WITH | SITE09 | -0.143 | 0.014 | -10.454 | 0 | -10.2143 | 0 |
| Means | CHR | 2.136 | 0.134 | 15.998 | 0 | 15.9403 | 0 |
| Means | AGE | 4.782 | 0.15 | 31.872 | 0 | 31.88 | 0 |
| Means | FEMALE | 0.912 | 0.044 | 20.745 | 0 | 20.72727 | 0 |
| Means | SITE02 | 0.31 | 0.026 | 11.777 | 0 | 11.92308 | 0 |
| Means | SITE03 | 0.252 | 0.026 | 9.888 | 0 | 9.692308 | 0 |
| Means | SITE04 | 0.31 | 0.026 | 11.776 | 0 | 11.92308 | 0 |
| Means | SITE05 | 0.404 | 0.028 | 14.482 | 0 | 14.42857 | 0 |
| Means | SITE06 | 0.385 | 0.028 | 13.967 | 0 | 13.75 | 0 |
| Means | SITE07 | 0.401 | 0.028 | 14.382 | 0 | 14.32143 | 0 |
| Means | SITE08 | 0.344 | 0.027 | 12.826 | 0 | 12.74074 | 0 |
| Means | SITE09 | 0.416 | 0.028 | 14.773 | 0 | 14.85714 | 0 |
| Intercepts | GAF_CH | 0.736 | 0.282 | 2.607 | 0.009 | 2.609929 | 0.009056 |
| Intercepts | HIL_CH | -0.064 | 0.047 | -1.349 | 0.177 | -1.3617 | 0.173292 |
| Variances | CHR | 1 | 0 |  |  |  |  |
| Variances | AGE | 1 | 0 |  |  |  |  |
| Variances | FEMALE | 1 | 0 |  |  |  |  |
| Variances | SITE02 | 1 | 0 |  |  |  |  |
| Variances | SITE03 | 1 | 0 |  |  |  |  |
| Variances | SITE04 | 1 | 0 |  |  |  |  |
| Variances | SITE05 | 1 | 0 |  |  |  |  |
| Variances | SITE06 | 1 | 0 |  |  |  |  |
| Variances | SITE07 | 1 | 0 |  |  |  |  |
| Variances | SITE08 | 1 | 0 |  |  |  |  |
| Variances | SITE09 | 1 | 0 |  |  |  |  |
| Variances | LV_HIL | 1 | 0 |  |  |  |  |
| Residual.Variances | GAF_CH | 0.172 | 0.025 | 7.002 | 0 | 6.88 | 5.99E-12 |
| Residual.Variances | HIL_CH | 0.328 | 0.038 | 8.551 | 0 | 8.631579 | 0 |
| Residual.Variances | LV_GAF | 0.872 | 0.054 | 16.059 | 0 | 16.14815 | 0 |
| LV_HIR.BY | HIR_CH | 0.83 | 0.029 | 28.966 | 0 | 28.62069 | 0 |
| LV_GAF.BY | GAF_CH | 0.909 | 0.013 | 67.718 | 0 | 69.92308 | 0 |
| LV_GAF.ON | LV_HIR | 0.032 | 0.067 | 0.473 | 0.636 | 0.477612 | 0.632926 |
| LV_GAF.ON | HIRXCHR | 0.143 | 0.053 | 2.712 | 0.007 | 2.698113 | 0.006973 |
| LV_GAF.ON | CHR | 0.125 | 0.035 | 3.599 | 0 | 3.571429 | 0.000355 |
| LV_GAF.ON | AGE | -0.094 | 0.054 | -1.72 | 0.086 | -1.74074 | 0.081729 |
| LV_GAF.ON | FEMALE | -0.025 | 0.049 | -0.515 | 0.607 | -0.5102 | 0.609908 |
| LV_GAF.ON | SITE02 | -0.061 | 0.067 | -0.918 | 0.359 | -0.91045 | 0.362586 |
| LV_GAF.ON | SITE03 | -0.066 | 0.055 | -1.197 | 0.231 | -1.2 | 0.230139 |
| LV_GAF.ON | SITE04 | 0.001 | 0.062 | 0.019 | 0.985 | 0.016129 | 0.987131 |
| LV_GAF.ON | SITE05 | -0.064 | 0.074 | -0.865 | 0.387 | -0.86486 | 0.387113 |
| LV_GAF.ON | SITE06 | -0.114 | 0.071 | -1.601 | 0.109 | -1.60563 | 0.108354 |
| LV_GAF.ON | SITE07 | -0.145 | 0.082 | -1.771 | 0.076 | -1.76829 | 0.077012 |
| LV_GAF.ON | SITE08 | -0.009 | 0.087 | -0.103 | 0.918 | -0.10345 | 0.917607 |
| LV_GAF.ON | SITE09 | -0.142 | 0.077 | -1.841 | 0.066 | -1.84416 | 0.06516 |
| LV_HIR.WITH | CHR | 0.015 | 0.045 | 0.342 | 0.733 | 0.333333 | 0.738883 |
| LV_HIR.WITH | AGE | -0.021 | 0.054 | -0.388 | 0.698 | -0.38889 | 0.697358 |
| LV_HIR.WITH | FEMALE | -0.015 | 0.058 | -0.262 | 0.793 | -0.25862 | 0.795928 |
| LV_HIR.WITH | SITE02 | -0.015 | 0.064 | -0.227 | 0.82 | -0.23438 | 0.814694 |
| LV_HIR.WITH | SITE03 | -0.1 | 0.073 | -1.377 | 0.169 | -1.36986 | 0.17073 |
| LV_HIR.WITH | SITE04 | -0.04 | 0.058 | -0.695 | 0.487 | -0.68966 | 0.490411 |
| LV_HIR.WITH | SITE05 | 0.002 | 0.05 | 0.043 | 0.966 | 0.04 | 0.968093 |
| LV_HIR.WITH | SITE06 | 0.025 | 0.047 | 0.538 | 0.591 | 0.531915 | 0.594785 |
| LV_HIR.WITH | SITE07 | 0.069 | 0.07 | 0.984 | 0.325 | 0.985714 | 0.324273 |
| LV_HIR.WITH | SITE08 | 0.034 | 0.054 | 0.629 | 0.529 | 0.62963 | 0.528937 |
| LV_HIR.WITH | SITE09 | -0.042 | 0.054 | -0.767 | 0.443 | -0.77778 | 0.4367 |
| CHR.WITH | AGE | -0.048 | 0.051 | -0.942 | 0.346 | -0.94118 | 0.346614 |
| CHR.WITH | FEMALE | -0.031 | 0.048 | -0.649 | 0.516 | -0.64583 | 0.518387 |
| CHR.WITH | SITE02 | -0.046 | 0.053 | -0.875 | 0.382 | -0.86792 | 0.385436 |
| CHR.WITH | SITE03 | -0.034 | 0.052 | -0.64 | 0.522 | -0.65385 | 0.513211 |
| CHR.WITH | SITE04 | 0.018 | 0.046 | 0.384 | 0.701 | 0.391304 | 0.695572 |
| CHR.WITH | SITE05 | -0.001 | 0.048 | -0.013 | 0.989 | -0.02083 | 0.983379 |
| CHR.WITH | SITE06 | -0.017 | 0.049 | -0.339 | 0.735 | -0.34694 | 0.728637 |
| CHR.WITH | SITE07 | -0.004 | 0.048 | -0.078 | 0.938 | -0.08333 | 0.933586 |
| CHR.WITH | SITE08 | 0.025 | 0.045 | 0.546 | 0.585 | 0.555556 | 0.578515 |
| CHR.WITH | SITE09 | 0.009 | 0.047 | 0.18 | 0.857 | 0.191489 | 0.848142 |
| AGE.WITH | FEMALE | -0.111 | 0.048 | -2.336 | 0.02 | -2.3125 | 0.02075 |
| AGE.WITH | SITE02 | 0.304 | 0.052 | 5.886 | 0 | 5.846154 | 5.03E-09 |
| AGE.WITH | SITE03 | 0 | 0.044 | -0.006 | 0.995 | 0 | 1 |
| AGE.WITH | SITE04 | -0.014 | 0.038 | -0.355 | 0.722 | -0.36842 | 0.712559 |
| AGE.WITH | SITE05 | 0.155 | 0.043 | 3.577 | 0 | 3.604651 | 0.000313 |
| AGE.WITH | SITE06 | -0.01 | 0.04 | -0.237 | 0.813 | -0.25 | 0.802587 |
| AGE.WITH | SITE07 | -0.143 | 0.046 | -3.137 | 0.002 | -3.1087 | 0.001879 |
| AGE.WITH | SITE08 | -0.171 | 0.049 | -3.485 | 0 | -3.4898 | 0.000483 |
| AGE.WITH | SITE09 | -0.03 | 0.044 | -0.691 | 0.49 | -0.68182 | 0.495354 |
| FEMALE.WITH | SITE02 | 0.029 | 0.048 | 0.595 | 0.552 | 0.604167 | 0.545733 |
| FEMALE.WITH | SITE03 | -0.016 | 0.048 | -0.328 | 0.743 | -0.33333 | 0.738883 |
| FEMALE.WITH | SITE04 | -0.004 | 0.048 | -0.085 | 0.932 | -0.08333 | 0.933586 |
| FEMALE.WITH | SITE05 | 0.004 | 0.048 | 0.086 | 0.931 | 0.083333 | 0.933586 |
| FEMALE.WITH | SITE06 | -0.061 | 0.047 | -1.299 | 0.194 | -1.29787 | 0.194331 |
| FEMALE.WITH | SITE07 | 0.064 | 0.048 | 1.326 | 0.185 | 1.333333 | 0.182422 |
| FEMALE.WITH | SITE08 | 0.002 | 0.048 | 0.038 | 0.97 | 0.041667 | 0.966764 |
| FEMALE.WITH | SITE09 | 0.012 | 0.048 | 0.258 | 0.797 | 0.25 | 0.802587 |
| SITE02.WITH | SITE03 | -0.078 | 0.01 | -7.882 | 0 | -7.8 | 6.22E-15 |
| SITE02.WITH | SITE04 | -0.096 | 0.011 | -8.758 | 0 | -8.72727 | 0 |
| SITE02.WITH | SITE05 | -0.125 | 0.013 | -9.754 | 0 | -9.61538 | 0 |
| SITE02.WITH | SITE06 | -0.119 | 0.012 | -9.584 | 0 | -9.91667 | 0 |
| SITE02.WITH | SITE07 | -0.124 | 0.013 | -9.722 | 0 | -9.53846 | 0 |
| SITE02.WITH | SITE08 | -0.107 | 0.012 | -9.176 | 0 | -8.91667 | 0 |
| SITE02.WITH | SITE09 | -0.129 | 0.013 | -9.848 | 0 | -9.92308 | 0 |
| SITE03.WITH | SITE04 | -0.078 | 0.01 | -7.882 | 0 | -7.8 | 6.22E-15 |
| SITE03.WITH | SITE05 | -0.102 | 0.012 | -8.584 | 0 | -8.5 | 0 |
| SITE03.WITH | SITE06 | -0.097 | 0.011 | -8.468 | 0 | -8.81818 | 0 |
| SITE03.WITH | SITE07 | -0.101 | 0.012 | -8.562 | 0 | -8.41667 | 0 |
| SITE03.WITH | SITE08 | -0.087 | 0.011 | -8.182 | 0 | -7.90909 | 2.66E-15 |
| SITE03.WITH | SITE09 | -0.105 | 0.012 | -8.648 | 0 | -8.75 | 0 |
| SITE04.WITH | SITE05 | -0.125 | 0.013 | -9.754 | 0 | -9.61538 | 0 |
| SITE04.WITH | SITE06 | -0.119 | 0.012 | -9.584 | 0 | -9.91667 | 0 |
| SITE04.WITH | SITE07 | -0.124 | 0.013 | -9.722 | 0 | -9.53846 | 0 |
| SITE04.WITH | SITE08 | -0.107 | 0.012 | -9.176 | 0 | -8.91667 | 0 |
| SITE04.WITH | SITE09 | -0.129 | 0.013 | -9.848 | 0 | -9.92308 | 0 |
| SITE05.WITH | SITE06 | -0.156 | 0.014 | -10.94 | 0 | -11.1429 | 0 |
| SITE05.WITH | SITE07 | -0.162 | 0.015 | -11.147 | 0 | -10.8 | 0 |
| SITE05.WITH | SITE08 | -0.139 | 0.013 | -10.343 | 0 | -10.6923 | 0 |
| SITE05.WITH | SITE09 | -0.168 | 0.015 | -11.339 | 0 | -11.2 | 0 |
| SITE06.WITH | SITE07 | -0.154 | 0.014 | -10.894 | 0 | -11 | 0 |
| SITE06.WITH | SITE08 | -0.133 | 0.013 | -10.14 | 0 | -10.2308 | 0 |
| SITE06.WITH | SITE09 | -0.16 | 0.014 | -11.073 | 0 | -11.4286 | 0 |
| SITE07.WITH | SITE08 | -0.138 | 0.013 | -10.304 | 0 | -10.6154 | 0 |
| SITE07.WITH | SITE09 | -0.167 | 0.015 | -11.287 | 0 | -11.1333 | 0 |
| SITE08.WITH | SITE09 | -0.143 | 0.014 | -10.454 | 0 | -10.2143 | 0 |
| Means | CHR | 2.136 | 0.134 | 15.998 | 0 | 15.9403 | 0 |
| Means | AGE | 4.782 | 0.15 | 31.872 | 0 | 31.88 | 0 |
| Means | FEMALE | 0.912 | 0.044 | 20.744 | 0 | 20.72727 | 0 |
| Means | SITE02 | 0.31 | 0.026 | 11.777 | 0 | 11.92308 | 0 |
| Means | SITE03 | 0.252 | 0.026 | 9.888 | 0 | 9.692308 | 0 |
| Means | SITE04 | 0.31 | 0.026 | 11.777 | 0 | 11.92308 | 0 |
| Means | SITE05 | 0.404 | 0.028 | 14.481 | 0 | 14.42857 | 0 |
| Means | SITE06 | 0.385 | 0.028 | 13.968 | 0 | 13.75 | 0 |
| Means | SITE07 | 0.401 | 0.028 | 14.381 | 0 | 14.32143 | 0 |
| Means | SITE08 | 0.344 | 0.027 | 12.826 | 0 | 12.74074 | 0 |
| Means | SITE09 | 0.416 | 0.028 | 14.773 | 0 | 14.85714 | 0 |
| Intercepts | GAF_CH | 0.652 | 0.284 | 2.292 | 0.022 | 2.295775 | 0.021689 |
| Intercepts | HIR_CH | 0.014 | 0.048 | 0.294 | 0.769 | 0.291667 | 0.770541 |
| Variances | CHR | 1 | 0 |  |  |  |  |
| Variances | AGE | 1 | 0 |  |  |  |  |
| Variances | FEMALE | 1 | 0 |  |  |  |  |
| Variances | SITE02 | 1 | 0 |  |  |  |  |
| Variances | SITE03 | 1 | 0 |  |  |  |  |
| Variances | SITE04 | 1 | 0 |  |  |  |  |
| Variances | SITE05 | 1 | 0 |  |  |  |  |
| Variances | SITE06 | 1 | 0 |  |  |  |  |
| Variances | SITE07 | 1 | 0 |  |  |  |  |
| Variances | SITE08 | 1 | 0 |  |  |  |  |
| Variances | SITE09 | 1 | 0 |  |  |  |  |
| Variances | LV_HIR | 1 | 0 |  |  |  |  |
| Residual.Variances | GAF_CH | 0.173 | 0.024 | 7.076 | 0 | 7.208333 | 5.66E-13 |
| Residual.Variances | HIR_CH | 0.311 | 0.048 | 6.532 | 0 | 6.479167 | 9.22E-11 |
| Residual.Variances | LV_GAF | 0.809 | 0.079 | 10.293 | 0 | 10.24051 | 0 |
| LV_HIH.BY | HIH_CH | 0.815 | 0.024 | 33.487 | 0 | 33.95833 | 0 |
| LV_NEGS.BY | NEGS_CH | 0.909 | 0.019 | 47.338 | 0 | 47.84211 | 0 |
| LV_NEGS.ON | LV_HIH | 0.12 | 0.085 | 1.406 | 0.16 | 1.411765 | 0.158019 |
| LV_NEGS.ON | HIHXCHR | -0.212 | 0.057 | -3.699 | 0 | -3.7193 | 0.0002 |
| LV_NEGS.ON | CHR | -0.128 | 0.036 | -3.607 | 0 | -3.55556 | 0.000377 |
| LV_NEGS.ON | AGE | -0.012 | 0.054 | -0.221 | 0.825 | -0.22222 | 0.824141 |
| LV_NEGS.ON | FEMALE | -0.002 | 0.046 | -0.036 | 0.972 | -0.04348 | 0.96532 |
| LV_NEGS.ON | SITE02 | -0.147 | 0.076 | -1.923 | 0.054 | -1.93421 | 0.053087 |
| LV_NEGS.ON | SITE03 | 0.058 | 0.065 | 0.885 | 0.376 | 0.892308 | 0.372228 |
| LV_NEGS.ON | SITE04 | -0.113 | 0.065 | -1.734 | 0.083 | -1.73846 | 0.08213 |
| LV_NEGS.ON | SITE05 | -0.039 | 0.079 | -0.493 | 0.622 | -0.49367 | 0.621539 |
| LV_NEGS.ON | SITE06 | -0.1 | 0.08 | -1.25 | 0.211 | -1.25 | 0.2113 |
| LV_NEGS.ON | SITE07 | -0.078 | 0.075 | -1.046 | 0.296 | -1.04 | 0.29834 |
| LV_NEGS.ON | SITE08 | -0.152 | 0.075 | -2.021 | 0.043 | -2.02667 | 0.042697 |
| LV_NEGS.ON | SITE09 | 0.033 | 0.091 | 0.362 | 0.717 | 0.362637 | 0.716876 |
| LV_HIH.WITH | CHR | -0.009 | 0.045 | -0.193 | 0.847 | -0.2 | 0.841481 |
| LV_HIH.WITH | AGE | 0.034 | 0.061 | 0.554 | 0.579 | 0.557377 | 0.57727 |
| LV_HIH.WITH | FEMALE | -0.072 | 0.059 | -1.225 | 0.221 | -1.22034 | 0.222336 |
| LV_HIH.WITH | SITE02 | 0.056 | 0.07 | 0.801 | 0.423 | 0.8 | 0.423711 |
| LV_HIH.WITH | SITE03 | -0.061 | 0.078 | -0.784 | 0.433 | -0.78205 | 0.434184 |
| LV_HIH.WITH | SITE04 | 0.077 | 0.045 | 1.713 | 0.087 | 1.711111 | 0.087061 |
| LV_HIH.WITH | SITE05 | -0.064 | 0.062 | -1.018 | 0.309 | -1.03226 | 0.301951 |
| LV_HIH.WITH | SITE06 | 0.083 | 0.043 | 1.901 | 0.057 | 1.930233 | 0.053578 |
| LV_HIH.WITH | SITE07 | 0 | 0.058 | -0.005 | 0.996 | 0 | 1 |
| LV_HIH.WITH | SITE08 | -0.029 | 0.058 | -0.493 | 0.622 | -0.5 | 0.617075 |
| LV_HIH.WITH | SITE09 | -0.043 | 0.059 | -0.739 | 0.46 | -0.72881 | 0.466116 |
| CHR.WITH | AGE | -0.048 | 0.051 | -0.953 | 0.34 | -0.94118 | 0.346614 |
| CHR.WITH | FEMALE | -0.031 | 0.048 | -0.649 | 0.516 | -0.64583 | 0.518387 |
| CHR.WITH | SITE02 | -0.046 | 0.053 | -0.875 | 0.382 | -0.86792 | 0.385436 |
| CHR.WITH | SITE03 | -0.039 | 0.053 | -0.731 | 0.465 | -0.73585 | 0.461823 |
| CHR.WITH | SITE04 | 0.018 | 0.046 | 0.384 | 0.701 | 0.391304 | 0.695572 |
| CHR.WITH | SITE05 | -0.001 | 0.048 | -0.013 | 0.989 | -0.02083 | 0.983379 |
| CHR.WITH | SITE06 | -0.017 | 0.049 | -0.339 | 0.735 | -0.34694 | 0.728637 |
| CHR.WITH | SITE07 | -0.004 | 0.048 | -0.078 | 0.938 | -0.08333 | 0.933586 |
| CHR.WITH | SITE08 | 0.025 | 0.045 | 0.546 | 0.585 | 0.555556 | 0.578515 |
| CHR.WITH | SITE09 | 0.009 | 0.047 | 0.18 | 0.857 | 0.191489 | 0.848142 |
| AGE.WITH | FEMALE | -0.11 | 0.048 | -2.312 | 0.021 | -2.29167 | 0.021925 |
| AGE.WITH | SITE02 | 0.304 | 0.052 | 5.892 | 0 | 5.846154 | 5.03E-09 |
| AGE.WITH | SITE03 | 0.001 | 0.045 | 0.032 | 0.975 | 0.022222 | 0.982271 |
| AGE.WITH | SITE04 | -0.013 | 0.038 | -0.345 | 0.73 | -0.34211 | 0.732272 |
| AGE.WITH | SITE05 | 0.155 | 0.043 | 3.587 | 0 | 3.604651 | 0.000313 |
| AGE.WITH | SITE06 | -0.009 | 0.04 | -0.225 | 0.822 | -0.225 | 0.821979 |
| AGE.WITH | SITE07 | -0.143 | 0.046 | -3.126 | 0.002 | -3.1087 | 0.001879 |
| AGE.WITH | SITE08 | -0.17 | 0.049 | -3.477 | 0.001 | -3.46939 | 0.000522 |
| AGE.WITH | SITE09 | -0.03 | 0.044 | -0.679 | 0.497 | -0.68182 | 0.495354 |
| FEMALE.WITH | SITE02 | 0.029 | 0.048 | 0.595 | 0.552 | 0.604167 | 0.545733 |
| FEMALE.WITH | SITE03 | -0.007 | 0.048 | -0.144 | 0.885 | -0.14583 | 0.884053 |
| FEMALE.WITH | SITE04 | -0.004 | 0.048 | -0.085 | 0.932 | -0.08333 | 0.933586 |
| FEMALE.WITH | SITE05 | 0.004 | 0.048 | 0.086 | 0.931 | 0.083333 | 0.933586 |
| FEMALE.WITH | SITE06 | -0.061 | 0.047 | -1.299 | 0.194 | -1.29787 | 0.194331 |
| FEMALE.WITH | SITE07 | 0.064 | 0.048 | 1.326 | 0.185 | 1.333333 | 0.182422 |
| FEMALE.WITH | SITE08 | 0.002 | 0.048 | 0.038 | 0.97 | 0.041667 | 0.966764 |
| FEMALE.WITH | SITE09 | 0.012 | 0.048 | 0.258 | 0.797 | 0.25 | 0.802587 |
| SITE02.WITH | SITE03 | -0.077 | 0.01 | -7.789 | 0 | -7.7 | 1.35E-14 |
| SITE02.WITH | SITE04 | -0.096 | 0.011 | -8.758 | 0 | -8.72727 | 0 |
| SITE02.WITH | SITE05 | -0.125 | 0.013 | -9.754 | 0 | -9.61538 | 0 |
| SITE02.WITH | SITE06 | -0.119 | 0.012 | -9.584 | 0 | -9.91667 | 0 |
| SITE02.WITH | SITE07 | -0.124 | 0.013 | -9.722 | 0 | -9.53846 | 0 |
| SITE02.WITH | SITE08 | -0.107 | 0.012 | -9.176 | 0 | -8.91667 | 0 |
| SITE02.WITH | SITE09 | -0.129 | 0.013 | -9.848 | 0 | -9.92308 | 0 |
| SITE03.WITH | SITE04 | -0.077 | 0.01 | -7.789 | 0 | -7.7 | 1.35E-14 |
| SITE03.WITH | SITE05 | -0.1 | 0.012 | -8.464 | 0 | -8.33333 | 0 |
| SITE03.WITH | SITE06 | -0.095 | 0.011 | -8.353 | 0 | -8.63636 | 0 |
| SITE03.WITH | SITE07 | -0.099 | 0.012 | -8.443 | 0 | -8.25 | 2.22E-16 |
| SITE03.WITH | SITE08 | -0.085 | 0.011 | -8.078 | 0 | -7.72727 | 1.09E-14 |
| SITE03.WITH | SITE09 | -0.103 | 0.012 | -8.525 | 0 | -8.58333 | 0 |
| SITE04.WITH | SITE05 | -0.125 | 0.013 | -9.754 | 0 | -9.61538 | 0 |
| SITE04.WITH | SITE06 | -0.119 | 0.012 | -9.584 | 0 | -9.91667 | 0 |
| SITE04.WITH | SITE07 | -0.124 | 0.013 | -9.722 | 0 | -9.53846 | 0 |
| SITE04.WITH | SITE08 | -0.107 | 0.012 | -9.176 | 0 | -8.91667 | 0 |
| SITE04.WITH | SITE09 | -0.129 | 0.013 | -9.848 | 0 | -9.92308 | 0 |
| SITE05.WITH | SITE06 | -0.156 | 0.014 | -10.94 | 0 | -11.1429 | 0 |
| SITE05.WITH | SITE07 | -0.162 | 0.015 | -11.147 | 0 | -10.8 | 0 |
| SITE05.WITH | SITE08 | -0.139 | 0.013 | -10.343 | 0 | -10.6923 | 0 |
| SITE05.WITH | SITE09 | -0.168 | 0.015 | -11.339 | 0 | -11.2 | 0 |
| SITE06.WITH | SITE07 | -0.154 | 0.014 | -10.894 | 0 | -11 | 0 |
| SITE06.WITH | SITE08 | -0.133 | 0.013 | -10.14 | 0 | -10.2308 | 0 |
| SITE06.WITH | SITE09 | -0.16 | 0.014 | -11.073 | 0 | -11.4286 | 0 |
| SITE07.WITH | SITE08 | -0.138 | 0.013 | -10.304 | 0 | -10.6154 | 0 |
| SITE07.WITH | SITE09 | -0.167 | 0.015 | -11.287 | 0 | -11.1333 | 0 |
| SITE08.WITH | SITE09 | -0.143 | 0.014 | -10.454 | 0 | -10.2143 | 0 |
| Means | CHR | 2.136 | 0.134 | 15.998 | 0 | 15.9403 | 0 |
| Means | AGE | 4.779 | 0.15 | 31.911 | 0 | 31.86 | 0 |
| Means | FEMALE | 0.912 | 0.044 | 20.742 | 0 | 20.72727 | 0 |
| Means | SITE02 | 0.31 | 0.026 | 11.777 | 0 | 11.92308 | 0 |
| Means | SITE03 | 0.247 | 0.025 | 9.707 | 0 | 9.88 | 0 |
| Means | SITE04 | 0.31 | 0.026 | 11.777 | 0 | 11.92308 | 0 |
| Means | SITE05 | 0.404 | 0.028 | 14.48 | 0 | 14.42857 | 0 |
| Means | SITE06 | 0.385 | 0.028 | 13.969 | 0 | 13.75 | 0 |
| Means | SITE07 | 0.401 | 0.028 | 14.381 | 0 | 14.32143 | 0 |
| Means | SITE08 | 0.344 | 0.027 | 12.825 | 0 | 12.74074 | 0 |
| Means | SITE09 | 0.416 | 0.028 | 14.773 | 0 | 14.85714 | 0 |
| Intercepts | NEGS_CH | 0.239 | 0.3 | 0.799 | 0.425 | 0.796667 | 0.425645 |
| Intercepts | HIH_CH | -0.062 | 0.048 | -1.308 | 0.191 | -1.29167 | 0.196473 |
| Variances | CHR | 1 | 0 |  |  |  |  |
| Variances | AGE | 1 | 0 |  |  |  |  |
| Variances | FEMALE | 1 | 0 |  |  |  |  |
| Variances | SITE02 | 1 | 0 |  |  |  |  |
| Variances | SITE03 | 1 | 0 |  |  |  |  |
| Variances | SITE04 | 1 | 0 |  |  |  |  |
| Variances | SITE05 | 1 | 0 |  |  |  |  |
| Variances | SITE06 | 1 | 0 |  |  |  |  |
| Variances | SITE07 | 1 | 0 |  |  |  |  |
| Variances | SITE08 | 1 | 0 |  |  |  |  |
| Variances | SITE09 | 1 | 0 |  |  |  |  |
| Variances | LV_HIH | 1 | 0 |  |  |  |  |
| Residual.Variances | NEGS_CH | 0.175 | 0.035 | 5.004 | 0 | 5 | 5.73E-07 |
| Residual.Variances | HIH_CH | 0.336 | 0.04 | 8.484 | 0 | 8.4 | 0 |
| Residual.Variances | LV_NEGS | 0.761 | 0.087 | 8.71 | 0 | 8.747126 | 0 |
| LV_HIL.BY | HIL_CH | 0.82 | 0.023 | 35.263 | 0 | 35.65217 | 0 |
| LV_NEGS.BY | NEGS_CH | 0.909 | 0.019 | 47.121 | 0 | 47.84211 | 0 |
| LV_NEGS.ON | LV_HIL | -0.047 | 0.083 | -0.567 | 0.571 | -0.56627 | 0.571214 |
| LV_NEGS.ON | HILXCHR | -0.009 | 0.074 | -0.124 | 0.902 | -0.12162 | 0.903199 |
| LV_NEGS.ON | CHR | -0.128 | 0.036 | -3.564 | 0 | -3.55556 | 0.000377 |
| LV_NEGS.ON | AGE | -0.026 | 0.06 | -0.429 | 0.668 | -0.43333 | 0.664773 |
| LV_NEGS.ON | FEMALE | 0.021 | 0.057 | 0.368 | 0.713 | 0.368421 | 0.712559 |
| LV_NEGS.ON | SITE02 | -0.155 | 0.084 | -1.845 | 0.065 | -1.84524 | 0.065003 |
| LV_NEGS.ON | SITE03 | 0.078 | 0.072 | 1.078 | 0.281 | 1.083333 | 0.27866 |
| LV_NEGS.ON | SITE04 | -0.129 | 0.068 | -1.911 | 0.056 | -1.89706 | 0.05782 |
| LV_NEGS.ON | SITE05 | -0.017 | 0.083 | -0.211 | 0.833 | -0.20482 | 0.837713 |
| LV_NEGS.ON | SITE06 | -0.124 | 0.081 | -1.527 | 0.127 | -1.53086 | 0.125803 |
| LV_NEGS.ON | SITE07 | -0.092 | 0.078 | -1.181 | 0.238 | -1.17949 | 0.238204 |
| LV_NEGS.ON | SITE08 | -0.136 | 0.076 | -1.801 | 0.072 | -1.78947 | 0.073539 |
| LV_NEGS.ON | SITE09 | 0.044 | 0.093 | 0.473 | 0.636 | 0.473118 | 0.636129 |
| LV_HIL.WITH | CHR | -0.004 | 0.049 | -0.079 | 0.937 | -0.08163 | 0.934939 |
| LV_HIL.WITH | AGE | 0.014 | 0.059 | 0.229 | 0.819 | 0.237288 | 0.812433 |
| LV_HIL.WITH | FEMALE | -0.117 | 0.061 | -1.927 | 0.054 | -1.91803 | 0.055107 |
| LV_HIL.WITH | SITE02 | -0.04 | 0.064 | -0.62 | 0.535 | -0.625 | 0.531971 |
| LV_HIL.WITH | SITE03 | 0.094 | 0.075 | 1.26 | 0.208 | 1.253333 | 0.210084 |
| LV_HIL.WITH | SITE04 | 0.101 | 0.049 | 2.056 | 0.04 | 2.061224 | 0.039282 |
| LV_HIL.WITH | SITE05 | -0.08 | 0.066 | -1.21 | 0.226 | -1.21212 | 0.225466 |
| LV_HIL.WITH | SITE06 | 0.05 | 0.047 | 1.049 | 0.294 | 1.06383 | 0.287406 |
| LV_HIL.WITH | SITE07 | -0.086 | 0.06 | -1.443 | 0.149 | -1.43333 | 0.151763 |
| LV_HIL.WITH | SITE08 | 0.02 | 0.052 | 0.394 | 0.693 | 0.384615 | 0.700522 |
| LV_HIL.WITH | SITE09 | 0.003 | 0.055 | 0.056 | 0.955 | 0.054545 | 0.956501 |
| CHR.WITH | AGE | -0.048 | 0.051 | -0.953 | 0.34 | -0.94118 | 0.346614 |
| CHR.WITH | FEMALE | -0.031 | 0.048 | -0.649 | 0.516 | -0.64583 | 0.518387 |
| CHR.WITH | SITE02 | -0.046 | 0.053 | -0.875 | 0.382 | -0.86792 | 0.385436 |
| CHR.WITH | SITE03 | -0.039 | 0.053 | -0.731 | 0.465 | -0.73585 | 0.461823 |
| CHR.WITH | SITE04 | 0.018 | 0.046 | 0.384 | 0.701 | 0.391304 | 0.695572 |
| CHR.WITH | SITE05 | -0.001 | 0.048 | -0.013 | 0.989 | -0.02083 | 0.983379 |
| CHR.WITH | SITE06 | -0.017 | 0.049 | -0.339 | 0.735 | -0.34694 | 0.728637 |
| CHR.WITH | SITE07 | -0.004 | 0.048 | -0.078 | 0.938 | -0.08333 | 0.933586 |
| CHR.WITH | SITE08 | 0.025 | 0.045 | 0.546 | 0.585 | 0.555556 | 0.578515 |
| CHR.WITH | SITE09 | 0.009 | 0.047 | 0.18 | 0.857 | 0.191489 | 0.848142 |
| AGE.WITH | FEMALE | -0.11 | 0.048 | -2.312 | 0.021 | -2.29167 | 0.021925 |
| AGE.WITH | SITE02 | 0.304 | 0.052 | 5.892 | 0 | 5.846154 | 5.03E-09 |
| AGE.WITH | SITE03 | 0.001 | 0.045 | 0.032 | 0.975 | 0.022222 | 0.982271 |
| AGE.WITH | SITE04 | -0.013 | 0.038 | -0.345 | 0.73 | -0.34211 | 0.732272 |
| AGE.WITH | SITE05 | 0.155 | 0.043 | 3.587 | 0 | 3.604651 | 0.000313 |
| AGE.WITH | SITE06 | -0.009 | 0.04 | -0.225 | 0.822 | -0.225 | 0.821979 |
| AGE.WITH | SITE07 | -0.143 | 0.046 | -3.126 | 0.002 | -3.1087 | 0.001879 |
| AGE.WITH | SITE08 | -0.17 | 0.049 | -3.477 | 0.001 | -3.46939 | 0.000522 |
| AGE.WITH | SITE09 | -0.03 | 0.044 | -0.679 | 0.497 | -0.68182 | 0.495354 |
| FEMALE.WITH | SITE02 | 0.029 | 0.048 | 0.595 | 0.552 | 0.604167 | 0.545733 |
| FEMALE.WITH | SITE03 | -0.007 | 0.048 | -0.144 | 0.885 | -0.14583 | 0.884053 |
| FEMALE.WITH | SITE04 | -0.004 | 0.048 | -0.085 | 0.932 | -0.08333 | 0.933586 |
| FEMALE.WITH | SITE05 | 0.004 | 0.048 | 0.086 | 0.931 | 0.083333 | 0.933586 |
| FEMALE.WITH | SITE06 | -0.061 | 0.047 | -1.299 | 0.194 | -1.29787 | 0.194331 |
| FEMALE.WITH | SITE07 | 0.064 | 0.048 | 1.326 | 0.185 | 1.333333 | 0.182422 |
| FEMALE.WITH | SITE08 | 0.002 | 0.048 | 0.038 | 0.97 | 0.041667 | 0.966764 |
| FEMALE.WITH | SITE09 | 0.012 | 0.048 | 0.258 | 0.797 | 0.25 | 0.802587 |
| SITE02.WITH | SITE03 | -0.077 | 0.01 | -7.789 | 0 | -7.7 | 1.35E-14 |
| SITE02.WITH | SITE04 | -0.096 | 0.011 | -8.758 | 0 | -8.72727 | 0 |
| SITE02.WITH | SITE05 | -0.125 | 0.013 | -9.754 | 0 | -9.61538 | 0 |
| SITE02.WITH | SITE06 | -0.119 | 0.012 | -9.584 | 0 | -9.91667 | 0 |
| SITE02.WITH | SITE07 | -0.124 | 0.013 | -9.722 | 0 | -9.53846 | 0 |
| SITE02.WITH | SITE08 | -0.107 | 0.012 | -9.176 | 0 | -8.91667 | 0 |
| SITE02.WITH | SITE09 | -0.129 | 0.013 | -9.848 | 0 | -9.92308 | 0 |
| SITE03.WITH | SITE04 | -0.077 | 0.01 | -7.789 | 0 | -7.7 | 1.35E-14 |
| SITE03.WITH | SITE05 | -0.1 | 0.012 | -8.464 | 0 | -8.33333 | 0 |
| SITE03.WITH | SITE06 | -0.095 | 0.011 | -8.353 | 0 | -8.63636 | 0 |
| SITE03.WITH | SITE07 | -0.099 | 0.012 | -8.443 | 0 | -8.25 | 2.22E-16 |
| SITE03.WITH | SITE08 | -0.085 | 0.011 | -8.078 | 0 | -7.72727 | 1.09E-14 |
| SITE03.WITH | SITE09 | -0.103 | 0.012 | -8.525 | 0 | -8.58333 | 0 |
| SITE04.WITH | SITE05 | -0.125 | 0.013 | -9.754 | 0 | -9.61538 | 0 |
| SITE04.WITH | SITE06 | -0.119 | 0.012 | -9.584 | 0 | -9.91667 | 0 |
| SITE04.WITH | SITE07 | -0.124 | 0.013 | -9.722 | 0 | -9.53846 | 0 |
| SITE04.WITH | SITE08 | -0.107 | 0.012 | -9.176 | 0 | -8.91667 | 0 |
| SITE04.WITH | SITE09 | -0.129 | 0.013 | -9.848 | 0 | -9.92308 | 0 |
| SITE05.WITH | SITE06 | -0.156 | 0.014 | -10.94 | 0 | -11.1429 | 0 |
| SITE05.WITH | SITE07 | -0.162 | 0.015 | -11.147 | 0 | -10.8 | 0 |
| SITE05.WITH | SITE08 | -0.139 | 0.013 | -10.343 | 0 | -10.6923 | 0 |
| SITE05.WITH | SITE09 | -0.168 | 0.015 | -11.339 | 0 | -11.2 | 0 |
| SITE06.WITH | SITE07 | -0.154 | 0.014 | -10.894 | 0 | -11 | 0 |
| SITE06.WITH | SITE08 | -0.133 | 0.013 | -10.14 | 0 | -10.2308 | 0 |
| SITE06.WITH | SITE09 | -0.16 | 0.014 | -11.073 | 0 | -11.4286 | 0 |
| SITE07.WITH | SITE08 | -0.138 | 0.013 | -10.304 | 0 | -10.6154 | 0 |
| SITE07.WITH | SITE09 | -0.167 | 0.015 | -11.287 | 0 | -11.1333 | 0 |
| SITE08.WITH | SITE09 | -0.143 | 0.014 | -10.454 | 0 | -10.2143 | 0 |
| Means | CHR | 2.136 | 0.134 | 15.998 | 0 | 15.9403 | 0 |
| Means | AGE | 4.779 | 0.15 | 31.911 | 0 | 31.86 | 0 |
| Means | FEMALE | 0.912 | 0.044 | 20.74 | 0 | 20.72727 | 0 |
| Means | SITE02 | 0.31 | 0.026 | 11.776 | 0 | 11.92308 | 0 |
| Means | SITE03 | 0.247 | 0.025 | 9.708 | 0 | 9.88 | 0 |
| Means | SITE04 | 0.31 | 0.026 | 11.778 | 0 | 11.92308 | 0 |
| Means | SITE05 | 0.404 | 0.028 | 14.48 | 0 | 14.42857 | 0 |
| Means | SITE06 | 0.385 | 0.028 | 13.968 | 0 | 13.75 | 0 |
| Means | SITE07 | 0.4 | 0.028 | 14.38 | 0 | 14.28571 | 0 |
| Means | SITE08 | 0.344 | 0.027 | 12.826 | 0 | 12.74074 | 0 |
| Means | SITE09 | 0.416 | 0.028 | 14.773 | 0 | 14.85714 | 0 |
| Intercepts | NEGS_CH | 0.281 | 0.313 | 0.899 | 0.369 | 0.897764 | 0.369312 |
| Intercepts | HIL_CH | -0.062 | 0.047 | -1.318 | 0.187 | -1.31915 | 0.187119 |
| Variances | CHR | 1 | 0 |  |  |  |  |
| Variances | AGE | 1 | 0 |  |  |  |  |
| Variances | FEMALE | 1 | 0 |  |  |  |  |
| Variances | SITE02 | 1 | 0 |  |  |  |  |
| Variances | SITE03 | 1 | 0 |  |  |  |  |
| Variances | SITE04 | 1 | 0 |  |  |  |  |
| Variances | SITE05 | 1 | 0 |  |  |  |  |
| Variances | SITE06 | 1 | 0 |  |  |  |  |
| Variances | SITE07 | 1 | 0 |  |  |  |  |
| Variances | SITE08 | 1 | 0 |  |  |  |  |
| Variances | SITE09 | 1 | 0 |  |  |  |  |
| Variances | LV_HIL | 1 | 0 |  |  |  |  |
| Residual.Variances | NEGS_CH | 0.173 | 0.035 | 4.946 | 0 | 4.942857 | 7.7E-07 |
| Residual.Variances | HIL_CH | 0.327 | 0.038 | 8.578 | 0 | 8.605263 | 0 |
| Residual.Variances | LV_NEGS | 0.912 | 0.024 | 38.445 | 0 | 38 | 0 |
| LV_HIR.BY | HIR_CH | 0.831 | 0.029 | 29.069 | 0 | 28.65517 | 0 |
| LV_NEGS.BY | NEGS_CH | 0.909 | 0.019 | 47.189 | 0 | 47.84211 | 0 |
| LV_NEGS.ON | LV_HIR | -0.046 | 0.076 | -0.599 | 0.549 | -0.60526 | 0.545004 |
| LV_NEGS.ON | HIRXCHR | -0.099 | 0.053 | -1.86 | 0.063 | -1.86792 | 0.061773 |
| LV_NEGS.ON | CHR | -0.128 | 0.035 | -3.63 | 0 | -3.65714 | 0.000255 |
| LV_NEGS.ON | AGE | -0.027 | 0.057 | -0.482 | 0.63 | -0.47368 | 0.635725 |
| LV_NEGS.ON | FEMALE | 0.024 | 0.048 | 0.503 | 0.615 | 0.5 | 0.617075 |
| LV_NEGS.ON | SITE02 | -0.163 | 0.084 | -1.934 | 0.053 | -1.94048 | 0.052322 |
| LV_NEGS.ON | SITE03 | 0.04 | 0.062 | 0.643 | 0.52 | 0.645161 | 0.518823 |
| LV_NEGS.ON | SITE04 | -0.155 | 0.067 | -2.295 | 0.022 | -2.31343 | 0.020699 |
| LV_NEGS.ON | SITE05 | -0.021 | 0.084 | -0.254 | 0.8 | -0.25 | 0.802587 |
| LV_NEGS.ON | SITE06 | -0.126 | 0.082 | -1.549 | 0.121 | -1.53659 | 0.124395 |
| LV_NEGS.ON | SITE07 | -0.076 | 0.079 | -0.962 | 0.336 | -0.96203 | 0.336037 |
| LV_NEGS.ON | SITE08 | -0.142 | 0.076 | -1.864 | 0.062 | -1.86842 | 0.061703 |
| LV_NEGS.ON | SITE09 | 0.028 | 0.095 | 0.296 | 0.767 | 0.294737 | 0.768195 |
| LV_HIR.WITH | CHR | 0.012 | 0.045 | 0.275 | 0.784 | 0.266667 | 0.789726 |
| LV_HIR.WITH | AGE | -0.018 | 0.055 | -0.324 | 0.746 | -0.32727 | 0.743462 |
| LV_HIR.WITH | FEMALE | -0.008 | 0.058 | -0.13 | 0.897 | -0.13793 | 0.890295 |
| LV_HIR.WITH | SITE02 | -0.012 | 0.065 | -0.187 | 0.852 | -0.18462 | 0.853531 |
| LV_HIR.WITH | SITE03 | -0.116 | 0.073 | -1.58 | 0.114 | -1.58904 | 0.112051 |
| LV_HIR.WITH | SITE04 | -0.038 | 0.058 | -0.656 | 0.512 | -0.65517 | 0.512357 |
| LV_HIR.WITH | SITE05 | 0.004 | 0.05 | 0.089 | 0.929 | 0.08 | 0.936237 |
| LV_HIR.WITH | SITE06 | 0.031 | 0.047 | 0.674 | 0.501 | 0.659574 | 0.509527 |
| LV_HIR.WITH | SITE07 | 0.071 | 0.069 | 1.03 | 0.303 | 1.028986 | 0.303486 |
| LV_HIR.WITH | SITE08 | 0.035 | 0.053 | 0.662 | 0.508 | 0.660377 | 0.509012 |
| LV_HIR.WITH | SITE09 | -0.039 | 0.054 | -0.716 | 0.474 | -0.72222 | 0.470158 |
| CHR.WITH | AGE | -0.048 | 0.051 | -0.953 | 0.34 | -0.94118 | 0.346614 |
| CHR.WITH | FEMALE | -0.031 | 0.048 | -0.649 | 0.516 | -0.64583 | 0.518387 |
| CHR.WITH | SITE02 | -0.046 | 0.053 | -0.875 | 0.382 | -0.86792 | 0.385436 |
| CHR.WITH | SITE03 | -0.039 | 0.053 | -0.731 | 0.465 | -0.73585 | 0.461823 |
| CHR.WITH | SITE04 | 0.018 | 0.046 | 0.384 | 0.701 | 0.391304 | 0.695572 |
| CHR.WITH | SITE05 | -0.001 | 0.048 | -0.013 | 0.989 | -0.02083 | 0.983379 |
| CHR.WITH | SITE06 | -0.017 | 0.049 | -0.339 | 0.735 | -0.34694 | 0.728637 |
| CHR.WITH | SITE07 | -0.004 | 0.048 | -0.078 | 0.938 | -0.08333 | 0.933586 |
| CHR.WITH | SITE08 | 0.025 | 0.045 | 0.546 | 0.585 | 0.555556 | 0.578515 |
| CHR.WITH | SITE09 | 0.009 | 0.047 | 0.18 | 0.857 | 0.191489 | 0.848142 |
| AGE.WITH | FEMALE | -0.11 | 0.048 | -2.312 | 0.021 | -2.29167 | 0.021925 |
| AGE.WITH | SITE02 | 0.304 | 0.052 | 5.892 | 0 | 5.846154 | 5.03E-09 |
| AGE.WITH | SITE03 | 0.001 | 0.045 | 0.032 | 0.975 | 0.022222 | 0.982271 |
| AGE.WITH | SITE04 | -0.013 | 0.038 | -0.345 | 0.73 | -0.34211 | 0.732272 |
| AGE.WITH | SITE05 | 0.155 | 0.043 | 3.587 | 0 | 3.604651 | 0.000313 |
| AGE.WITH | SITE06 | -0.009 | 0.04 | -0.225 | 0.822 | -0.225 | 0.821979 |
| AGE.WITH | SITE07 | -0.143 | 0.046 | -3.126 | 0.002 | -3.1087 | 0.001879 |
| AGE.WITH | SITE08 | -0.17 | 0.049 | -3.477 | 0.001 | -3.46939 | 0.000522 |
| AGE.WITH | SITE09 | -0.03 | 0.044 | -0.679 | 0.497 | -0.68182 | 0.495354 |
| FEMALE.WITH | SITE02 | 0.029 | 0.048 | 0.595 | 0.552 | 0.604167 | 0.545733 |
| FEMALE.WITH | SITE03 | -0.007 | 0.048 | -0.144 | 0.885 | -0.14583 | 0.884053 |
| FEMALE.WITH | SITE04 | -0.004 | 0.048 | -0.085 | 0.932 | -0.08333 | 0.933586 |
| FEMALE.WITH | SITE05 | 0.004 | 0.048 | 0.086 | 0.931 | 0.083333 | 0.933586 |
| FEMALE.WITH | SITE06 | -0.061 | 0.047 | -1.299 | 0.194 | -1.29787 | 0.194331 |
| FEMALE.WITH | SITE07 | 0.064 | 0.048 | 1.326 | 0.185 | 1.333333 | 0.182422 |
| FEMALE.WITH | SITE08 | 0.002 | 0.048 | 0.038 | 0.97 | 0.041667 | 0.966764 |
| FEMALE.WITH | SITE09 | 0.012 | 0.048 | 0.258 | 0.797 | 0.25 | 0.802587 |
| SITE02.WITH | SITE03 | -0.077 | 0.01 | -7.789 | 0 | -7.7 | 1.35E-14 |
| SITE02.WITH | SITE04 | -0.096 | 0.011 | -8.758 | 0 | -8.72727 | 0 |
| SITE02.WITH | SITE05 | -0.125 | 0.013 | -9.754 | 0 | -9.61538 | 0 |
| SITE02.WITH | SITE06 | -0.119 | 0.012 | -9.584 | 0 | -9.91667 | 0 |
| SITE02.WITH | SITE07 | -0.124 | 0.013 | -9.722 | 0 | -9.53846 | 0 |
| SITE02.WITH | SITE08 | -0.107 | 0.012 | -9.176 | 0 | -8.91667 | 0 |
| SITE02.WITH | SITE09 | -0.129 | 0.013 | -9.848 | 0 | -9.92308 | 0 |
| SITE03.WITH | SITE04 | -0.077 | 0.01 | -7.789 | 0 | -7.7 | 1.35E-14 |
| SITE03.WITH | SITE05 | -0.1 | 0.012 | -8.464 | 0 | -8.33333 | 0 |
| SITE03.WITH | SITE06 | -0.095 | 0.011 | -8.353 | 0 | -8.63636 | 0 |
| SITE03.WITH | SITE07 | -0.099 | 0.012 | -8.443 | 0 | -8.25 | 2.22E-16 |
| SITE03.WITH | SITE08 | -0.085 | 0.011 | -8.078 | 0 | -7.72727 | 1.09E-14 |
| SITE03.WITH | SITE09 | -0.103 | 0.012 | -8.525 | 0 | -8.58333 | 0 |
| SITE04.WITH | SITE05 | -0.125 | 0.013 | -9.754 | 0 | -9.61538 | 0 |
| SITE04.WITH | SITE06 | -0.119 | 0.012 | -9.584 | 0 | -9.91667 | 0 |
| SITE04.WITH | SITE07 | -0.124 | 0.013 | -9.722 | 0 | -9.53846 | 0 |
| SITE04.WITH | SITE08 | -0.107 | 0.012 | -9.176 | 0 | -8.91667 | 0 |
| SITE04.WITH | SITE09 | -0.129 | 0.013 | -9.848 | 0 | -9.92308 | 0 |
| SITE05.WITH | SITE06 | -0.156 | 0.014 | -10.94 | 0 | -11.1429 | 0 |
| SITE05.WITH | SITE07 | -0.162 | 0.015 | -11.147 | 0 | -10.8 | 0 |
| SITE05.WITH | SITE08 | -0.139 | 0.013 | -10.343 | 0 | -10.6923 | 0 |
| SITE05.WITH | SITE09 | -0.168 | 0.015 | -11.339 | 0 | -11.2 | 0 |
| SITE06.WITH | SITE07 | -0.154 | 0.014 | -10.894 | 0 | -11 | 0 |
| SITE06.WITH | SITE08 | -0.133 | 0.013 | -10.14 | 0 | -10.2308 | 0 |
| SITE06.WITH | SITE09 | -0.16 | 0.014 | -11.073 | 0 | -11.4286 | 0 |
| SITE07.WITH | SITE08 | -0.138 | 0.013 | -10.304 | 0 | -10.6154 | 0 |
| SITE07.WITH | SITE09 | -0.167 | 0.015 | -11.287 | 0 | -11.1333 | 0 |
| SITE08.WITH | SITE09 | -0.143 | 0.014 | -10.454 | 0 | -10.2143 | 0 |
| Means | CHR | 2.136 | 0.134 | 15.998 | 0 | 15.9403 | 0 |
| Means | AGE | 4.779 | 0.15 | 31.911 | 0 | 31.86 | 0 |
| Means | FEMALE | 0.912 | 0.044 | 20.744 | 0 | 20.72727 | 0 |
| Means | SITE02 | 0.31 | 0.026 | 11.777 | 0 | 11.92308 | 0 |
| Means | SITE03 | 0.247 | 0.025 | 9.708 | 0 | 9.88 | 0 |
| Means | SITE04 | 0.31 | 0.026 | 11.777 | 0 | 11.92308 | 0 |
| Means | SITE05 | 0.404 | 0.028 | 14.481 | 0 | 14.42857 | 0 |
| Means | SITE06 | 0.385 | 0.028 | 13.968 | 0 | 13.75 | 0 |
| Means | SITE07 | 0.401 | 0.028 | 14.381 | 0 | 14.32143 | 0 |
| Means | SITE08 | 0.344 | 0.027 | 12.826 | 0 | 12.74074 | 0 |
| Means | SITE09 | 0.416 | 0.028 | 14.773 | 0 | 14.85714 | 0 |
| Intercepts | NEGS_CH | 0.309 | 0.302 | 1.024 | 0.306 | 1.023179 | 0.306223 |
| Intercepts | HIR_CH | 0.009 | 0.048 | 0.183 | 0.855 | 0.1875 | 0.851269 |
| Variances | CHR | 1 | 0 |  |  |  |  |
| Variances | AGE | 1 | 0 |  |  |  |  |
| Variances | FEMALE | 1 | 0 |  |  |  |  |
| Variances | SITE02 | 1 | 0 |  |  |  |  |
| Variances | SITE03 | 1 | 0 |  |  |  |  |
| Variances | SITE04 | 1 | 0 |  |  |  |  |
| Variances | SITE05 | 1 | 0 |  |  |  |  |
| Variances | SITE06 | 1 | 0 |  |  |  |  |
| Variances | SITE07 | 1 | 0 |  |  |  |  |
| Variances | SITE08 | 1 | 0 |  |  |  |  |
| Variances | SITE09 | 1 | 0 |  |  |  |  |
| Variances | LV_HIR | 1 | 0 |  |  |  |  |
| Residual.Variances | NEGS_CH | 0.174 | 0.035 | 4.966 | 0 | 4.971429 | 6.65E-07 |
| Residual.Variances | HIR_CH | 0.31 | 0.047 | 6.531 | 0 | 6.595745 | 4.23E-11 |
| Residual.Variances | LV_NEGS | 0.839 | 0.062 | 13.589 | 0 | 13.53226 | 0 |
| LV_HIH.BY | HIH_CH | 0.818 | 0.024 | 33.941 | 0 | 34.08333 | 0 |
| LV_POSS.BY | POSS_CH | 0.926 | 0.016 | 57.266 | 0 | 57.875 | 0 |
| LV_POSS.ON | LV_HIH | -0.116 | 0.076 | -1.527 | 0.127 | -1.52632 | 0.126931 |
| LV_POSS.ON | HIHXCHR | -0.035 | 0.074 | -0.468 | 0.64 | -0.47297 | 0.636232 |
| LV_POSS.ON | CHR | -0.261 | 0.034 | -7.789 | 0 | -7.67647 | 1.64E-14 |
| LV_POSS.ON | AGE | 0.003 | 0.054 | 0.05 | 0.96 | 0.055556 | 0.955696 |
| LV_POSS.ON | FEMALE | 0.064 | 0.046 | 1.382 | 0.167 | 1.391304 | 0.164133 |
| LV_POSS.ON | SITE02 | 0.001 | 0.092 | 0.006 | 0.996 | 0.01087 | 0.991328 |
| LV_POSS.ON | SITE03 | 0.055 | 0.06 | 0.927 | 0.354 | 0.916667 | 0.359317 |
| LV_POSS.ON | SITE04 | 0.041 | 0.064 | 0.634 | 0.526 | 0.640625 | 0.521766 |
| LV_POSS.ON | SITE05 | 0.135 | 0.078 | 1.731 | 0.084 | 1.730769 | 0.083493 |
| LV_POSS.ON | SITE06 | 0.003 | 0.073 | 0.035 | 0.972 | 0.041096 | 0.967219 |
| LV_POSS.ON | SITE07 | 0.044 | 0.069 | 0.635 | 0.525 | 0.637681 | 0.523681 |
| LV_POSS.ON | SITE08 | 0.055 | 0.072 | 0.757 | 0.449 | 0.763889 | 0.444933 |
| LV_POSS.ON | SITE09 | 0.066 | 0.077 | 0.854 | 0.393 | 0.857143 | 0.391366 |
| LV_HIH.WITH | CHR | -0.003 | 0.044 | -0.067 | 0.947 | -0.06818 | 0.945641 |
| LV_HIH.WITH | AGE | 0.029 | 0.06 | 0.479 | 0.632 | 0.483333 | 0.628859 |
| LV_HIH.WITH | FEMALE | -0.081 | 0.058 | -1.381 | 0.167 | -1.39655 | 0.162548 |
| LV_HIH.WITH | SITE02 | 0.04 | 0.066 | 0.615 | 0.538 | 0.606061 | 0.544475 |
| LV_HIH.WITH | SITE03 | -0.007 | 0.09 | -0.074 | 0.941 | -0.07778 | 0.938005 |
| LV_HIH.WITH | SITE04 | 0.072 | 0.044 | 1.625 | 0.104 | 1.636364 | 0.101764 |
| LV_HIH.WITH | SITE05 | -0.062 | 0.061 | -1.006 | 0.315 | -1.01639 | 0.309442 |
| LV_HIH.WITH | SITE06 | 0.077 | 0.043 | 1.793 | 0.073 | 1.790698 | 0.073342 |
| LV_HIH.WITH | SITE07 | -0.01 | 0.058 | -0.166 | 0.869 | -0.17241 | 0.863112 |
| LV_HIH.WITH | SITE08 | -0.035 | 0.058 | -0.602 | 0.547 | -0.60345 | 0.546211 |
| LV_HIH.WITH | SITE09 | -0.042 | 0.058 | -0.72 | 0.471 | -0.72414 | 0.468981 |
| CHR.WITH | AGE | -0.049 | 0.051 | -0.956 | 0.339 | -0.96078 | 0.336661 |
| CHR.WITH | FEMALE | -0.032 | 0.048 | -0.67 | 0.503 | -0.66667 | 0.504985 |
| CHR.WITH | SITE02 | -0.046 | 0.053 | -0.881 | 0.378 | -0.86792 | 0.385436 |
| CHR.WITH | SITE03 | -0.034 | 0.052 | -0.645 | 0.519 | -0.65385 | 0.513211 |
| CHR.WITH | SITE04 | 0.017 | 0.046 | 0.377 | 0.706 | 0.369565 | 0.711706 |
| CHR.WITH | SITE05 | -0.001 | 0.048 | -0.022 | 0.982 | -0.02083 | 0.983379 |
| CHR.WITH | SITE06 | -0.017 | 0.049 | -0.347 | 0.728 | -0.34694 | 0.728637 |
| CHR.WITH | SITE07 | -0.004 | 0.048 | -0.087 | 0.931 | -0.08333 | 0.933586 |
| CHR.WITH | SITE08 | 0.024 | 0.045 | 0.538 | 0.591 | 0.533333 | 0.593803 |
| CHR.WITH | SITE09 | 0.008 | 0.047 | 0.17 | 0.865 | 0.170213 | 0.864843 |
| AGE.WITH | FEMALE | -0.11 | 0.048 | -2.305 | 0.021 | -2.29167 | 0.021925 |
| AGE.WITH | SITE02 | 0.304 | 0.052 | 5.894 | 0 | 5.846154 | 5.03E-09 |
| AGE.WITH | SITE03 | 0 | 0.044 | 0.003 | 0.998 | 0 | 1 |
| AGE.WITH | SITE04 | -0.013 | 0.038 | -0.343 | 0.732 | -0.34211 | 0.732272 |
| AGE.WITH | SITE05 | 0.155 | 0.043 | 3.59 | 0 | 3.604651 | 0.000313 |
| AGE.WITH | SITE06 | -0.009 | 0.04 | -0.222 | 0.824 | -0.225 | 0.821979 |
| AGE.WITH | SITE07 | -0.143 | 0.046 | -3.123 | 0.002 | -3.1087 | 0.001879 |
| AGE.WITH | SITE08 | -0.17 | 0.049 | -3.474 | 0.001 | -3.46939 | 0.000522 |
| AGE.WITH | SITE09 | -0.03 | 0.044 | -0.676 | 0.499 | -0.68182 | 0.495354 |
| FEMALE.WITH | SITE02 | 0.029 | 0.048 | 0.608 | 0.543 | 0.604167 | 0.545733 |
| FEMALE.WITH | SITE03 | -0.015 | 0.048 | -0.317 | 0.751 | -0.3125 | 0.754661 |
| FEMALE.WITH | SITE04 | -0.003 | 0.048 | -0.071 | 0.943 | -0.0625 | 0.950165 |
| FEMALE.WITH | SITE05 | 0.005 | 0.048 | 0.104 | 0.917 | 0.104167 | 0.917037 |
| FEMALE.WITH | SITE06 | -0.06 | 0.047 | -1.282 | 0.2 | -1.2766 | 0.201745 |
| FEMALE.WITH | SITE07 | 0.065 | 0.048 | 1.343 | 0.179 | 1.354167 | 0.175683 |
| FEMALE.WITH | SITE08 | 0.003 | 0.048 | 0.053 | 0.958 | 0.0625 | 0.950165 |
| FEMALE.WITH | SITE09 | 0.013 | 0.048 | 0.276 | 0.783 | 0.270833 | 0.786519 |
| SITE02.WITH | SITE03 | -0.078 | 0.01 | -7.882 | 0 | -7.8 | 6.22E-15 |
| SITE02.WITH | SITE04 | -0.096 | 0.011 | -8.758 | 0 | -8.72727 | 0 |
| SITE02.WITH | SITE05 | -0.125 | 0.013 | -9.754 | 0 | -9.61538 | 0 |
| SITE02.WITH | SITE06 | -0.119 | 0.012 | -9.584 | 0 | -9.91667 | 0 |
| SITE02.WITH | SITE07 | -0.124 | 0.013 | -9.722 | 0 | -9.53846 | 0 |
| SITE02.WITH | SITE08 | -0.106 | 0.012 | -9.176 | 0 | -8.83333 | 0 |
| SITE02.WITH | SITE09 | -0.128 | 0.013 | -9.848 | 0 | -9.84615 | 0 |
| SITE03.WITH | SITE04 | -0.078 | 0.01 | -7.882 | 0 | -7.8 | 6.22E-15 |
| SITE03.WITH | SITE05 | -0.102 | 0.012 | -8.584 | 0 | -8.5 | 0 |
| SITE03.WITH | SITE06 | -0.097 | 0.011 | -8.467 | 0 | -8.81818 | 0 |
| SITE03.WITH | SITE07 | -0.101 | 0.012 | -8.562 | 0 | -8.41667 | 0 |
| SITE03.WITH | SITE08 | -0.087 | 0.011 | -8.182 | 0 | -7.90909 | 2.66E-15 |
| SITE03.WITH | SITE09 | -0.105 | 0.012 | -8.647 | 0 | -8.75 | 0 |
| SITE04.WITH | SITE05 | -0.125 | 0.013 | -9.754 | 0 | -9.61538 | 0 |
| SITE04.WITH | SITE06 | -0.119 | 0.012 | -9.584 | 0 | -9.91667 | 0 |
| SITE04.WITH | SITE07 | -0.124 | 0.013 | -9.722 | 0 | -9.53846 | 0 |
| SITE04.WITH | SITE08 | -0.106 | 0.012 | -9.176 | 0 | -8.83333 | 0 |
| SITE04.WITH | SITE09 | -0.128 | 0.013 | -9.848 | 0 | -9.84615 | 0 |
| SITE05.WITH | SITE06 | -0.155 | 0.014 | -10.939 | 0 | -11.0714 | 0 |
| SITE05.WITH | SITE07 | -0.162 | 0.014 | -11.146 | 0 | -11.5714 | 0 |
| SITE05.WITH | SITE08 | -0.139 | 0.013 | -10.342 | 0 | -10.6923 | 0 |
| SITE05.WITH | SITE09 | -0.168 | 0.015 | -11.338 | 0 | -11.2 | 0 |
| SITE06.WITH | SITE07 | -0.154 | 0.014 | -10.893 | 0 | -11 | 0 |
| SITE06.WITH | SITE08 | -0.132 | 0.013 | -10.14 | 0 | -10.1538 | 0 |
| SITE06.WITH | SITE09 | -0.16 | 0.014 | -11.072 | 0 | -11.4286 | 0 |
| SITE07.WITH | SITE08 | -0.138 | 0.013 | -10.303 | 0 | -10.6154 | 0 |
| SITE07.WITH | SITE09 | -0.166 | 0.015 | -11.287 | 0 | -11.0667 | 0 |
| SITE08.WITH | SITE09 | -0.143 | 0.014 | -10.454 | 0 | -10.2143 | 0 |
| Means | CHR | 2.139 | 0.134 | 16.002 | 0 | 15.96269 | 0 |
| Means | AGE | 4.784 | 0.15 | 31.888 | 0 | 31.89333 | 0 |
| Means | FEMALE | 0.91 | 0.044 | 20.764 | 0 | 20.68182 | 0 |
| Means | SITE02 | 0.309 | 0.026 | 11.778 | 0 | 11.88462 | 0 |
| Means | SITE03 | 0.252 | 0.025 | 9.889 | 0 | 10.08 | 0 |
| Means | SITE04 | 0.309 | 0.026 | 11.778 | 0 | 11.88462 | 0 |
| Means | SITE05 | 0.404 | 0.028 | 14.484 | 0 | 14.42857 | 0 |
| Means | SITE06 | 0.384 | 0.028 | 13.97 | 0 | 13.71429 | 0 |
| Means | SITE07 | 0.4 | 0.028 | 14.384 | 0 | 14.28571 | 0 |
| Means | SITE08 | 0.344 | 0.027 | 12.827 | 0 | 12.74074 | 0 |
| Means | SITE09 | 0.415 | 0.028 | 14.776 | 0 | 14.82143 | 0 |
| Intercepts | POSS_CH | -0.334 | 0.298 | -1.12 | 0.263 | -1.12081 | 0.262371 |
| Intercepts | HIH_CH | -0.052 | 0.048 | -1.097 | 0.273 | -1.08333 | 0.27866 |
| Variances | CHR | 1 | 0 |  |  |  |  |
| Variances | AGE | 1 | 0 |  |  |  |  |
| Variances | FEMALE | 1 | 0 |  |  |  |  |
| Variances | SITE02 | 1 | 0 |  |  |  |  |
| Variances | SITE03 | 1 | 0 |  |  |  |  |
| Variances | SITE04 | 1 | 0 |  |  |  |  |
| Variances | SITE05 | 1 | 0 |  |  |  |  |
| Variances | SITE06 | 1 | 0 |  |  |  |  |
| Variances | SITE07 | 1 | 0 |  |  |  |  |
| Variances | SITE08 | 1 | 0 |  |  |  |  |
| Variances | SITE09 | 1 | 0 |  |  |  |  |
| Variances | LV_HIH | 1 | 0 |  |  |  |  |
| Residual.Variances | POSS_CH | 0.143 | 0.03 | 4.769 | 0 | 4.766667 | 1.87E-06 |
| Residual.Variances | HIH_CH | 0.331 | 0.039 | 8.413 | 0 | 8.487179 | 0 |
| Residual.Variances | LV_POSS | 0.868 | 0.052 | 16.672 | 0 | 16.69231 | 0 |
| LV_HIL.BY | HIL_CH | 0.82 | 0.023 | 35.26 | 0 | 35.65217 | 0 |
| LV_POSS.BY | POSS_CH | 0.926 | 0.016 | 56.476 | 0 | 57.875 | 0 |
| LV_POSS.ON | LV_HIL | -0.022 | 0.038 | -0.573 | 0.567 | -0.57895 | 0.562625 |
| LV_POSS.ON | HILXCHR | 0.059 | 0.063 | 0.935 | 0.35 | 0.936508 | 0.349012 |
| LV_POSS.ON | CHR | -0.26 | 0.034 | -7.682 | 0 | -7.64706 | 2.07E-14 |
| LV_POSS.ON | AGE | -0.007 | 0.059 | -0.113 | 0.91 | -0.11864 | 0.905557 |
| LV_POSS.ON | FEMALE | 0.091 | 0.05 | 1.814 | 0.07 | 1.82 | 0.068759 |
| LV_POSS.ON | SITE02 | -0.002 | 0.092 | -0.023 | 0.982 | -0.02174 | 0.982656 |
| LV_POSS.ON | SITE03 | 0.037 | 0.061 | 0.599 | 0.549 | 0.606557 | 0.544145 |
| LV_POSS.ON | SITE04 | 0.014 | 0.067 | 0.213 | 0.831 | 0.208955 | 0.834483 |
| LV_POSS.ON | SITE05 | 0.153 | 0.072 | 2.115 | 0.034 | 2.125 | 0.033587 |
| LV_POSS.ON | SITE06 | -0.021 | 0.071 | -0.288 | 0.774 | -0.29577 | 0.767402 |
| LV_POSS.ON | SITE07 | 0.045 | 0.07 | 0.642 | 0.521 | 0.642857 | 0.520317 |
| LV_POSS.ON | SITE08 | 0.055 | 0.072 | 0.765 | 0.445 | 0.763889 | 0.444933 |
| LV_POSS.ON | SITE09 | 0.069 | 0.076 | 0.906 | 0.365 | 0.907895 | 0.363934 |
| LV_HIL.WITH | CHR | -0.004 | 0.049 | -0.075 | 0.941 | -0.08163 | 0.934939 |
| LV_HIL.WITH | AGE | 0.013 | 0.059 | 0.213 | 0.831 | 0.220339 | 0.825607 |
| LV_HIL.WITH | FEMALE | -0.117 | 0.061 | -1.933 | 0.053 | -1.91803 | 0.055107 |
| LV_HIL.WITH | SITE02 | -0.039 | 0.062 | -0.632 | 0.527 | -0.62903 | 0.529328 |
| LV_HIL.WITH | SITE03 | 0.094 | 0.073 | 1.296 | 0.195 | 1.287671 | 0.19786 |
| LV_HIL.WITH | SITE04 | 0.1 | 0.049 | 2.059 | 0.04 | 2.040816 | 0.041269 |
| LV_HIL.WITH | SITE05 | -0.079 | 0.066 | -1.205 | 0.228 | -1.19697 | 0.231318 |
| LV_HIL.WITH | SITE06 | 0.048 | 0.047 | 1.016 | 0.31 | 1.021277 | 0.307123 |
| LV_HIL.WITH | SITE07 | -0.086 | 0.059 | -1.452 | 0.147 | -1.45763 | 0.144943 |
| LV_HIL.WITH | SITE08 | 0.021 | 0.052 | 0.407 | 0.684 | 0.403846 | 0.686326 |
| LV_HIL.WITH | SITE09 | 0.002 | 0.055 | 0.044 | 0.965 | 0.036364 | 0.970992 |
| CHR.WITH | AGE | -0.049 | 0.051 | -0.956 | 0.339 | -0.96078 | 0.336661 |
| CHR.WITH | FEMALE | -0.032 | 0.048 | -0.67 | 0.503 | -0.66667 | 0.504985 |
| CHR.WITH | SITE02 | -0.046 | 0.053 | -0.881 | 0.378 | -0.86792 | 0.385436 |
| CHR.WITH | SITE03 | -0.034 | 0.052 | -0.645 | 0.519 | -0.65385 | 0.513211 |
| CHR.WITH | SITE04 | 0.017 | 0.046 | 0.377 | 0.706 | 0.369565 | 0.711706 |
| CHR.WITH | SITE05 | -0.001 | 0.048 | -0.022 | 0.982 | -0.02083 | 0.983379 |
| CHR.WITH | SITE06 | -0.017 | 0.049 | -0.347 | 0.728 | -0.34694 | 0.728637 |
| CHR.WITH | SITE07 | -0.004 | 0.048 | -0.087 | 0.931 | -0.08333 | 0.933586 |
| CHR.WITH | SITE08 | 0.024 | 0.045 | 0.538 | 0.591 | 0.533333 | 0.593803 |
| CHR.WITH | SITE09 | 0.008 | 0.047 | 0.17 | 0.865 | 0.170213 | 0.864843 |
| AGE.WITH | FEMALE | -0.11 | 0.048 | -2.305 | 0.021 | -2.29167 | 0.021925 |
| AGE.WITH | SITE02 | 0.304 | 0.052 | 5.894 | 0 | 5.846154 | 5.03E-09 |
| AGE.WITH | SITE03 | 0 | 0.044 | 0.003 | 0.998 | 0 | 1 |
| AGE.WITH | SITE04 | -0.013 | 0.038 | -0.343 | 0.732 | -0.34211 | 0.732272 |
| AGE.WITH | SITE05 | 0.155 | 0.043 | 3.59 | 0 | 3.604651 | 0.000313 |
| AGE.WITH | SITE06 | -0.009 | 0.04 | -0.222 | 0.824 | -0.225 | 0.821979 |
| AGE.WITH | SITE07 | -0.143 | 0.046 | -3.123 | 0.002 | -3.1087 | 0.001879 |
| AGE.WITH | SITE08 | -0.17 | 0.049 | -3.474 | 0.001 | -3.46939 | 0.000522 |
| AGE.WITH | SITE09 | -0.03 | 0.044 | -0.676 | 0.499 | -0.68182 | 0.495354 |
| FEMALE.WITH | SITE02 | 0.029 | 0.048 | 0.608 | 0.543 | 0.604167 | 0.545733 |
| FEMALE.WITH | SITE03 | -0.015 | 0.048 | -0.317 | 0.751 | -0.3125 | 0.754661 |
| FEMALE.WITH | SITE04 | -0.003 | 0.048 | -0.071 | 0.943 | -0.0625 | 0.950165 |
| FEMALE.WITH | SITE05 | 0.005 | 0.048 | 0.104 | 0.917 | 0.104167 | 0.917037 |
| FEMALE.WITH | SITE06 | -0.06 | 0.047 | -1.282 | 0.2 | -1.2766 | 0.201745 |
| FEMALE.WITH | SITE07 | 0.065 | 0.048 | 1.343 | 0.179 | 1.354167 | 0.175683 |
| FEMALE.WITH | SITE08 | 0.003 | 0.048 | 0.053 | 0.958 | 0.0625 | 0.950165 |
| FEMALE.WITH | SITE09 | 0.013 | 0.048 | 0.276 | 0.783 | 0.270833 | 0.786519 |
| SITE02.WITH | SITE03 | -0.078 | 0.01 | -7.882 | 0 | -7.8 | 6.22E-15 |
| SITE02.WITH | SITE04 | -0.096 | 0.011 | -8.758 | 0 | -8.72727 | 0 |
| SITE02.WITH | SITE05 | -0.125 | 0.013 | -9.754 | 0 | -9.61538 | 0 |
| SITE02.WITH | SITE06 | -0.119 | 0.012 | -9.584 | 0 | -9.91667 | 0 |
| SITE02.WITH | SITE07 | -0.124 | 0.013 | -9.722 | 0 | -9.53846 | 0 |
| SITE02.WITH | SITE08 | -0.106 | 0.012 | -9.176 | 0 | -8.83333 | 0 |
| SITE02.WITH | SITE09 | -0.128 | 0.013 | -9.848 | 0 | -9.84615 | 0 |
| SITE03.WITH | SITE04 | -0.078 | 0.01 | -7.882 | 0 | -7.8 | 6.22E-15 |
| SITE03.WITH | SITE05 | -0.102 | 0.012 | -8.584 | 0 | -8.5 | 0 |
| SITE03.WITH | SITE06 | -0.097 | 0.011 | -8.467 | 0 | -8.81818 | 0 |
| SITE03.WITH | SITE07 | -0.101 | 0.012 | -8.562 | 0 | -8.41667 | 0 |
| SITE03.WITH | SITE08 | -0.087 | 0.011 | -8.182 | 0 | -7.90909 | 2.66E-15 |
| SITE03.WITH | SITE09 | -0.105 | 0.012 | -8.647 | 0 | -8.75 | 0 |
| SITE04.WITH | SITE05 | -0.125 | 0.013 | -9.754 | 0 | -9.61538 | 0 |
| SITE04.WITH | SITE06 | -0.119 | 0.012 | -9.584 | 0 | -9.91667 | 0 |
| SITE04.WITH | SITE07 | -0.124 | 0.013 | -9.722 | 0 | -9.53846 | 0 |
| SITE04.WITH | SITE08 | -0.106 | 0.012 | -9.176 | 0 | -8.83333 | 0 |
| SITE04.WITH | SITE09 | -0.128 | 0.013 | -9.848 | 0 | -9.84615 | 0 |
| SITE05.WITH | SITE06 | -0.155 | 0.014 | -10.939 | 0 | -11.0714 | 0 |
| SITE05.WITH | SITE07 | -0.162 | 0.014 | -11.146 | 0 | -11.5714 | 0 |
| SITE05.WITH | SITE08 | -0.139 | 0.013 | -10.342 | 0 | -10.6923 | 0 |
| SITE05.WITH | SITE09 | -0.168 | 0.015 | -11.338 | 0 | -11.2 | 0 |
| SITE06.WITH | SITE07 | -0.154 | 0.014 | -10.893 | 0 | -11 | 0 |
| SITE06.WITH | SITE08 | -0.132 | 0.013 | -10.14 | 0 | -10.1538 | 0 |
| SITE06.WITH | SITE09 | -0.16 | 0.014 | -11.072 | 0 | -11.4286 | 0 |
| SITE07.WITH | SITE08 | -0.138 | 0.013 | -10.303 | 0 | -10.6154 | 0 |
| SITE07.WITH | SITE09 | -0.166 | 0.015 | -11.287 | 0 | -11.0667 | 0 |
| SITE08.WITH | SITE09 | -0.143 | 0.014 | -10.454 | 0 | -10.2143 | 0 |
| Means | CHR | 2.139 | 0.134 | 16.002 | 0 | 15.96269 | 0 |
| Means | AGE | 4.784 | 0.15 | 31.887 | 0 | 31.89333 | 0 |
| Means | FEMALE | 0.91 | 0.044 | 20.76 | 0 | 20.68182 | 0 |
| Means | SITE02 | 0.309 | 0.026 | 11.778 | 0 | 11.88462 | 0 |
| Means | SITE03 | 0.252 | 0.026 | 9.889 | 0 | 9.692308 | 0 |
| Means | SITE04 | 0.31 | 0.026 | 11.779 | 0 | 11.92308 | 0 |
| Means | SITE05 | 0.404 | 0.028 | 14.483 | 0 | 14.42857 | 0 |
| Means | SITE06 | 0.384 | 0.028 | 13.971 | 0 | 13.71429 | 0 |
| Means | SITE07 | 0.4 | 0.028 | 14.382 | 0 | 14.28571 | 0 |
| Means | SITE08 | 0.344 | 0.027 | 12.828 | 0 | 12.74074 | 0 |
| Means | SITE09 | 0.415 | 0.028 | 14.776 | 0 | 14.82143 | 0 |
| Intercepts | POSS_CH | -0.303 | 0.327 | -0.928 | 0.353 | -0.92661 | 0.354131 |
| Intercepts | HIL_CH | -0.062 | 0.047 | -1.313 | 0.189 | -1.31915 | 0.187119 |
| Variances | CHR | 1 | 0 |  |  |  |  |
| Variances | AGE | 1 | 0 |  |  |  |  |
| Variances | FEMALE | 1 | 0 |  |  |  |  |
| Variances | SITE02 | 1 | 0 |  |  |  |  |
| Variances | SITE03 | 1 | 0 |  |  |  |  |
| Variances | SITE04 | 1 | 0 |  |  |  |  |
| Variances | SITE05 | 1 | 0 |  |  |  |  |
| Variances | SITE06 | 1 | 0 |  |  |  |  |
| Variances | SITE07 | 1 | 0 |  |  |  |  |
| Variances | SITE08 | 1 | 0 |  |  |  |  |
| Variances | SITE09 | 1 | 0 |  |  |  |  |
| Variances | LV_HIL | 1 | 0 |  |  |  |  |
| Residual.Variances | POSS_CH | 0.142 | 0.03 | 4.688 | 0 | 4.733333 | 2.21E-06 |
| Residual.Variances | HIL_CH | 0.328 | 0.038 | 8.591 | 0 | 8.631579 | 0 |
| Residual.Variances | LV_POSS | 0.889 | 0.036 | 24.644 | 0 | 24.69444 | 0 |
| LV_HIR.BY | HIR_CH | 0.83 | 0.029 | 29.101 | 0 | 28.62069 | 0 |
| LV_POSS.BY | POSS_CH | 0.926 | 0.016 | 56.637 | 0 | 57.875 | 0 |
| LV_POSS.ON | LV_HIR | 0 | 0.062 | -0.001 | 1 | 0 | 1 |
| LV_POSS.ON | HIRXCHR | -0.094 | 0.043 | -2.166 | 0.03 | -2.18605 | 0.028812 |
| LV_POSS.ON | CHR | -0.261 | 0.033 | -7.804 | 0 | -7.90909 | 2.66E-15 |
| LV_POSS.ON | AGE | -0.001 | 0.055 | -0.017 | 0.986 | -0.01818 | 0.985494 |
| LV_POSS.ON | FEMALE | 0.076 | 0.045 | 1.687 | 0.092 | 1.688889 | 0.091241 |
| LV_POSS.ON | SITE02 | -0.018 | 0.094 | -0.187 | 0.852 | -0.19149 | 0.848142 |
| LV_POSS.ON | SITE03 | 0.033 | 0.059 | 0.572 | 0.567 | 0.559322 | 0.575942 |
| LV_POSS.ON | SITE04 | 0.01 | 0.065 | 0.16 | 0.873 | 0.153846 | 0.877731 |
| LV_POSS.ON | SITE05 | 0.137 | 0.074 | 1.847 | 0.065 | 1.851351 | 0.064119 |
| LV_POSS.ON | SITE06 | -0.012 | 0.07 | -0.17 | 0.865 | -0.17143 | 0.863887 |
| LV_POSS.ON | SITE07 | 0.051 | 0.068 | 0.747 | 0.455 | 0.75 | 0.453255 |
| LV_POSS.ON | SITE08 | 0.054 | 0.071 | 0.759 | 0.448 | 0.760563 | 0.446918 |
| LV_POSS.ON | SITE09 | 0.058 | 0.076 | 0.766 | 0.444 | 0.763158 | 0.445369 |
| LV_HIR.WITH | CHR | 0.014 | 0.045 | 0.309 | 0.757 | 0.311111 | 0.755716 |
| LV_HIR.WITH | AGE | -0.018 | 0.055 | -0.335 | 0.738 | -0.32727 | 0.743462 |
| LV_HIR.WITH | FEMALE | -0.01 | 0.058 | -0.169 | 0.865 | -0.17241 | 0.863112 |
| LV_HIR.WITH | SITE02 | -0.019 | 0.064 | -0.299 | 0.765 | -0.29688 | 0.766562 |
| LV_HIR.WITH | SITE03 | -0.098 | 0.073 | -1.342 | 0.18 | -1.34247 | 0.179445 |
| LV_HIR.WITH | SITE04 | -0.04 | 0.058 | -0.684 | 0.494 | -0.68966 | 0.490411 |
| LV_HIR.WITH | SITE05 | 0.005 | 0.05 | 0.093 | 0.926 | 0.1 | 0.920344 |
| LV_HIR.WITH | SITE06 | 0.032 | 0.047 | 0.679 | 0.497 | 0.680851 | 0.495966 |
| LV_HIR.WITH | SITE07 | 0.069 | 0.069 | 0.988 | 0.323 | 1 | 0.317311 |
| LV_HIR.WITH | SITE08 | 0.032 | 0.053 | 0.594 | 0.552 | 0.603774 | 0.545994 |
| LV_HIR.WITH | SITE09 | -0.037 | 0.054 | -0.676 | 0.499 | -0.68519 | 0.493227 |
| CHR.WITH | AGE | -0.049 | 0.051 | -0.956 | 0.339 | -0.96078 | 0.336661 |
| CHR.WITH | FEMALE | -0.032 | 0.048 | -0.67 | 0.503 | -0.66667 | 0.504985 |
| CHR.WITH | SITE02 | -0.046 | 0.053 | -0.881 | 0.378 | -0.86792 | 0.385436 |
| CHR.WITH | SITE03 | -0.034 | 0.052 | -0.645 | 0.519 | -0.65385 | 0.513211 |
| CHR.WITH | SITE04 | 0.017 | 0.046 | 0.377 | 0.706 | 0.369565 | 0.711706 |
| CHR.WITH | SITE05 | -0.001 | 0.048 | -0.022 | 0.982 | -0.02083 | 0.983379 |
| CHR.WITH | SITE06 | -0.017 | 0.049 | -0.347 | 0.728 | -0.34694 | 0.728637 |
| CHR.WITH | SITE07 | -0.004 | 0.048 | -0.087 | 0.931 | -0.08333 | 0.933586 |
| CHR.WITH | SITE08 | 0.024 | 0.045 | 0.538 | 0.591 | 0.533333 | 0.593803 |
| CHR.WITH | SITE09 | 0.008 | 0.047 | 0.17 | 0.865 | 0.170213 | 0.864843 |
| AGE.WITH | FEMALE | -0.11 | 0.048 | -2.305 | 0.021 | -2.29167 | 0.021925 |
| AGE.WITH | SITE02 | 0.304 | 0.052 | 5.894 | 0 | 5.846154 | 5.03E-09 |
| AGE.WITH | SITE03 | 0 | 0.044 | 0.003 | 0.998 | 0 | 1 |
| AGE.WITH | SITE04 | -0.013 | 0.038 | -0.343 | 0.732 | -0.34211 | 0.732272 |
| AGE.WITH | SITE05 | 0.155 | 0.043 | 3.59 | 0 | 3.604651 | 0.000313 |
| AGE.WITH | SITE06 | -0.009 | 0.04 | -0.222 | 0.824 | -0.225 | 0.821979 |
| AGE.WITH | SITE07 | -0.143 | 0.046 | -3.123 | 0.002 | -3.1087 | 0.001879 |
| AGE.WITH | SITE08 | -0.17 | 0.049 | -3.474 | 0.001 | -3.46939 | 0.000522 |
| AGE.WITH | SITE09 | -0.03 | 0.044 | -0.676 | 0.499 | -0.68182 | 0.495354 |
| FEMALE.WITH | SITE02 | 0.029 | 0.048 | 0.608 | 0.543 | 0.604167 | 0.545733 |
| FEMALE.WITH | SITE03 | -0.015 | 0.048 | -0.317 | 0.751 | -0.3125 | 0.754661 |
| FEMALE.WITH | SITE04 | -0.003 | 0.048 | -0.071 | 0.943 | -0.0625 | 0.950165 |
| FEMALE.WITH | SITE05 | 0.005 | 0.048 | 0.104 | 0.917 | 0.104167 | 0.917037 |
| FEMALE.WITH | SITE06 | -0.06 | 0.047 | -1.282 | 0.2 | -1.2766 | 0.201745 |
| FEMALE.WITH | SITE07 | 0.065 | 0.048 | 1.343 | 0.179 | 1.354167 | 0.175683 |
| FEMALE.WITH | SITE08 | 0.003 | 0.048 | 0.053 | 0.958 | 0.0625 | 0.950165 |
| FEMALE.WITH | SITE09 | 0.013 | 0.048 | 0.276 | 0.783 | 0.270833 | 0.786519 |
| SITE02.WITH | SITE03 | -0.078 | 0.01 | -7.882 | 0 | -7.8 | 6.22E-15 |
| SITE02.WITH | SITE04 | -0.096 | 0.011 | -8.758 | 0 | -8.72727 | 0 |
| SITE02.WITH | SITE05 | -0.125 | 0.013 | -9.754 | 0 | -9.61538 | 0 |
| SITE02.WITH | SITE06 | -0.119 | 0.012 | -9.584 | 0 | -9.91667 | 0 |
| SITE02.WITH | SITE07 | -0.124 | 0.013 | -9.722 | 0 | -9.53846 | 0 |
| SITE02.WITH | SITE08 | -0.106 | 0.012 | -9.176 | 0 | -8.83333 | 0 |
| SITE02.WITH | SITE09 | -0.128 | 0.013 | -9.848 | 0 | -9.84615 | 0 |
| SITE03.WITH | SITE04 | -0.078 | 0.01 | -7.882 | 0 | -7.8 | 6.22E-15 |
| SITE03.WITH | SITE05 | -0.102 | 0.012 | -8.584 | 0 | -8.5 | 0 |
| SITE03.WITH | SITE06 | -0.097 | 0.011 | -8.467 | 0 | -8.81818 | 0 |
| SITE03.WITH | SITE07 | -0.101 | 0.012 | -8.562 | 0 | -8.41667 | 0 |
| SITE03.WITH | SITE08 | -0.087 | 0.011 | -8.182 | 0 | -7.90909 | 2.66E-15 |
| SITE03.WITH | SITE09 | -0.105 | 0.012 | -8.647 | 0 | -8.75 | 0 |
| SITE04.WITH | SITE05 | -0.125 | 0.013 | -9.754 | 0 | -9.61538 | 0 |
| SITE04.WITH | SITE06 | -0.119 | 0.012 | -9.584 | 0 | -9.91667 | 0 |
| SITE04.WITH | SITE07 | -0.124 | 0.013 | -9.722 | 0 | -9.53846 | 0 |
| SITE04.WITH | SITE08 | -0.106 | 0.012 | -9.176 | 0 | -8.83333 | 0 |
| SITE04.WITH | SITE09 | -0.128 | 0.013 | -9.848 | 0 | -9.84615 | 0 |
| SITE05.WITH | SITE06 | -0.155 | 0.014 | -10.939 | 0 | -11.0714 | 0 |
| SITE05.WITH | SITE07 | -0.162 | 0.014 | -11.146 | 0 | -11.5714 | 0 |
| SITE05.WITH | SITE08 | -0.139 | 0.013 | -10.342 | 0 | -10.6923 | 0 |
| SITE05.WITH | SITE09 | -0.168 | 0.015 | -11.338 | 0 | -11.2 | 0 |
| SITE06.WITH | SITE07 | -0.154 | 0.014 | -10.893 | 0 | -11 | 0 |
| SITE06.WITH | SITE08 | -0.132 | 0.013 | -10.14 | 0 | -10.1538 | 0 |
| SITE06.WITH | SITE09 | -0.16 | 0.014 | -11.072 | 0 | -11.4286 | 0 |
| SITE07.WITH | SITE08 | -0.138 | 0.013 | -10.303 | 0 | -10.6154 | 0 |
| SITE07.WITH | SITE09 | -0.166 | 0.015 | -11.287 | 0 | -11.0667 | 0 |
| SITE08.WITH | SITE09 | -0.143 | 0.014 | -10.454 | 0 | -10.2143 | 0 |
| Means | CHR | 2.139 | 0.134 | 16.002 | 0 | 15.96269 | 0 |
| Means | AGE | 4.784 | 0.15 | 31.888 | 0 | 31.89333 | 0 |
| Means | FEMALE | 0.91 | 0.044 | 20.764 | 0 | 20.68182 | 0 |
| Means | SITE02 | 0.309 | 0.026 | 11.778 | 0 | 11.88462 | 0 |
| Means | SITE03 | 0.252 | 0.025 | 9.889 | 0 | 10.08 | 0 |
| Means | SITE04 | 0.309 | 0.026 | 11.778 | 0 | 11.88462 | 0 |
| Means | SITE05 | 0.404 | 0.028 | 14.484 | 0 | 14.42857 | 0 |
| Means | SITE06 | 0.384 | 0.028 | 13.97 | 0 | 13.71429 | 0 |
| Means | SITE07 | 0.4 | 0.028 | 14.384 | 0 | 14.28571 | 0 |
| Means | SITE08 | 0.344 | 0.027 | 12.827 | 0 | 12.74074 | 0 |
| Means | SITE09 | 0.415 | 0.028 | 14.776 | 0 | 14.82143 | 0 |
| Intercepts | POSS_CH | -0.302 | 0.314 | -0.965 | 0.335 | -0.96178 | 0.336158 |
| Intercepts | HIR_CH | 0.012 | 0.048 | 0.239 | 0.811 | 0.25 | 0.802587 |
| Variances | CHR | 1 | 0 |  |  |  |  |
| Variances | AGE | 1 | 0 |  |  |  |  |
| Variances | FEMALE | 1 | 0 |  |  |  |  |
| Variances | SITE02 | 1 | 0 |  |  |  |  |
| Variances | SITE03 | 1 | 0 |  |  |  |  |
| Variances | SITE04 | 1 | 0 |  |  |  |  |
| Variances | SITE05 | 1 | 0 |  |  |  |  |
| Variances | SITE06 | 1 | 0 |  |  |  |  |
| Variances | SITE07 | 1 | 0 |  |  |  |  |
| Variances | SITE08 | 1 | 0 |  |  |  |  |
| Variances | SITE09 | 1 | 0 |  |  |  |  |
| Variances | LV_HIR | 1 | 0 |  |  |  |  |
| Residual.Variances | POSS_CH | 0.143 | 0.03 | 4.72 | 0 | 4.766667 | 1.87E-06 |
| Residual.Variances | HIR_CH | 0.311 | 0.047 | 6.559 | 0 | 6.617021 | 3.67E-11 |
| Residual.Variances | LV_POSS | 0.854 | 0.039 | 21.771 | 0 | 21.89744 | 0 |
| LV_HIH.BY | HIH_CH | 0.819 | 0.024 | 33.886 | 0 | 34.125 | 0 |
| LV_SYCOD.BY | SYCOD_CH | 0.892 | 0.016 | 55.496 | 0 | 55.75 | 0 |
| LV_SYCOD.ON | LV_HIH | 0.028 | 0.215 | 0.131 | 0.895 | 0.130233 | 0.896382 |
| LV_SYCOD.ON | HIHXCHR | 0.007 | 0.091 | 0.082 | 0.934 | 0.076923 | 0.938685 |
| LV_SYCOD.ON | CHR | -0.056 | 0.049 | -1.143 | 0.253 | -1.14286 | 0.253098 |
| LV_SYCOD.ON | AGE | -0.029 | 0.053 | -0.538 | 0.59 | -0.54717 | 0.584262 |
| LV_SYCOD.ON | FEMALE | -0.082 | 0.057 | -1.432 | 0.152 | -1.4386 | 0.150265 |
| LV_SYCOD.ON | SITE02 | 0.005 | 0.089 | 0.062 | 0.951 | 0.05618 | 0.955199 |
| LV_SYCOD.ON | SITE03 | 0.06 | 0.086 | 0.704 | 0.482 | 0.697674 | 0.485381 |
| LV_SYCOD.ON | SITE04 | -0.141 | 0.059 | -2.399 | 0.016 | -2.38983 | 0.016856 |
| LV_SYCOD.ON | SITE05 | 0.112 | 0.079 | 1.429 | 0.153 | 1.417722 | 0.156272 |
| LV_SYCOD.ON | SITE06 | -0.067 | 0.069 | -0.974 | 0.33 | -0.97101 | 0.331541 |
| LV_SYCOD.ON | SITE07 | -0.07 | 0.067 | -1.03 | 0.303 | -1.04478 | 0.296126 |
| LV_SYCOD.ON | SITE08 | -0.167 | 0.069 | -2.415 | 0.016 | -2.42029 | 0.015508 |
| LV_SYCOD.ON | SITE09 | -0.001 | 0.084 | -0.013 | 0.989 | -0.0119 | 0.990502 |
| LV_HIH.WITH | CHR | -0.008 | 0.045 | -0.175 | 0.861 | -0.17778 | 0.858898 |
| LV_HIH.WITH | AGE | 0.039 | 0.059 | 0.66 | 0.509 | 0.661017 | 0.508601 |
| LV_HIH.WITH | FEMALE | -0.077 | 0.059 | -1.302 | 0.193 | -1.30508 | 0.191864 |
| LV_HIH.WITH | SITE02 | 0.047 | 0.066 | 0.702 | 0.483 | 0.712121 | 0.47639 |
| LV_HIH.WITH | SITE03 | -0.005 | 0.09 | -0.058 | 0.954 | -0.05556 | 0.955696 |
| LV_HIH.WITH | SITE04 | 0.069 | 0.045 | 1.541 | 0.123 | 1.533333 | 0.125194 |
| LV_HIH.WITH | SITE05 | -0.056 | 0.062 | -0.907 | 0.364 | -0.90323 | 0.366406 |
| LV_HIH.WITH | SITE06 | 0.078 | 0.044 | 1.791 | 0.073 | 1.772727 | 0.076274 |
| LV_HIH.WITH | SITE07 | -0.008 | 0.058 | -0.133 | 0.894 | -0.13793 | 0.890295 |
| LV_HIH.WITH | SITE08 | -0.033 | 0.058 | -0.568 | 0.57 | -0.56897 | 0.56938 |
| LV_HIH.WITH | SITE09 | -0.04 | 0.058 | -0.695 | 0.487 | -0.68966 | 0.490411 |
| CHR.WITH | AGE | -0.049 | 0.051 | -0.961 | 0.337 | -0.96078 | 0.336661 |
| CHR.WITH | FEMALE | -0.033 | 0.048 | -0.685 | 0.493 | -0.6875 | 0.491768 |
| CHR.WITH | SITE02 | -0.03 | 0.051 | -0.578 | 0.563 | -0.58824 | 0.556374 |
| CHR.WITH | SITE03 | -0.034 | 0.053 | -0.649 | 0.517 | -0.64151 | 0.521192 |
| CHR.WITH | SITE04 | 0.014 | 0.047 | 0.291 | 0.771 | 0.297872 | 0.765801 |
| CHR.WITH | SITE05 | -0.004 | 0.049 | -0.092 | 0.927 | -0.08163 | 0.934939 |
| CHR.WITH | SITE06 | -0.018 | 0.05 | -0.352 | 0.724 | -0.36 | 0.718847 |
| CHR.WITH | SITE07 | -0.004 | 0.049 | -0.092 | 0.927 | -0.08163 | 0.934939 |
| CHR.WITH | SITE08 | 0.024 | 0.046 | 0.533 | 0.594 | 0.521739 | 0.601852 |
| CHR.WITH | SITE09 | 0.008 | 0.048 | 0.165 | 0.869 | 0.166667 | 0.867632 |
| AGE.WITH | FEMALE | -0.11 | 0.048 | -2.305 | 0.021 | -2.29167 | 0.021925 |
| AGE.WITH | SITE02 | 0.31 | 0.052 | 5.938 | 0 | 5.961538 | 2.5E-09 |
| AGE.WITH | SITE03 | 0 | 0.045 | 0.007 | 0.995 | 0 | 1 |
| AGE.WITH | SITE04 | -0.01 | 0.039 | -0.256 | 0.798 | -0.25641 | 0.797634 |
| AGE.WITH | SITE05 | 0.151 | 0.044 | 3.466 | 0.001 | 3.431818 | 0.0006 |
| AGE.WITH | SITE06 | -0.009 | 0.041 | -0.216 | 0.829 | -0.21951 | 0.826251 |
| AGE.WITH | SITE07 | -0.143 | 0.046 | -3.122 | 0.002 | -3.1087 | 0.001879 |
| AGE.WITH | SITE08 | -0.171 | 0.049 | -3.474 | 0.001 | -3.4898 | 0.000483 |
| AGE.WITH | SITE09 | -0.03 | 0.044 | -0.67 | 0.503 | -0.68182 | 0.495354 |
| FEMALE.WITH | SITE02 | 0.034 | 0.048 | 0.704 | 0.481 | 0.708333 | 0.478738 |
| FEMALE.WITH | SITE03 | -0.018 | 0.048 | -0.373 | 0.709 | -0.375 | 0.70766 |
| FEMALE.WITH | SITE04 | 0.001 | 0.048 | 0.017 | 0.987 | 0.020833 | 0.983379 |
| FEMALE.WITH | SITE05 | 0.007 | 0.048 | 0.143 | 0.886 | 0.145833 | 0.884053 |
| FEMALE.WITH | SITE06 | -0.065 | 0.047 | -1.369 | 0.171 | -1.38298 | 0.166671 |
| FEMALE.WITH | SITE07 | 0.061 | 0.048 | 1.257 | 0.209 | 1.270833 | 0.203788 |
| FEMALE.WITH | SITE08 | -0.001 | 0.048 | -0.023 | 0.981 | -0.02083 | 0.983379 |
| FEMALE.WITH | SITE09 | 0.009 | 0.048 | 0.184 | 0.854 | 0.1875 | 0.851269 |
| SITE02.WITH | SITE03 | -0.078 | 0.01 | -7.839 | 0 | -7.8 | 6.22E-15 |
| SITE02.WITH | SITE04 | -0.094 | 0.011 | -8.641 | 0 | -8.54545 | 0 |
| SITE02.WITH | SITE05 | -0.124 | 0.013 | -9.642 | 0 | -9.53846 | 0 |
| SITE02.WITH | SITE06 | -0.119 | 0.012 | -9.507 | 0 | -9.91667 | 0 |
| SITE02.WITH | SITE07 | -0.124 | 0.013 | -9.642 | 0 | -9.53846 | 0 |
| SITE02.WITH | SITE08 | -0.106 | 0.012 | -9.108 | 0 | -8.83333 | 0 |
| SITE02.WITH | SITE09 | -0.128 | 0.013 | -9.765 | 0 | -9.84615 | 0 |
| SITE03.WITH | SITE04 | -0.078 | 0.01 | -7.839 | 0 | -7.8 | 6.22E-15 |
| SITE03.WITH | SITE05 | -0.102 | 0.012 | -8.563 | 0 | -8.5 | 0 |
| SITE03.WITH | SITE06 | -0.098 | 0.012 | -8.468 | 0 | -8.16667 | 2.22E-16 |
| SITE03.WITH | SITE07 | -0.102 | 0.012 | -8.563 | 0 | -8.5 | 0 |
| SITE03.WITH | SITE08 | -0.088 | 0.011 | -8.183 | 0 | -8 | 1.33E-15 |
| SITE03.WITH | SITE09 | -0.106 | 0.012 | -8.649 | 0 | -8.83333 | 0 |
| SITE04.WITH | SITE05 | -0.124 | 0.013 | -9.642 | 0 | -9.53846 | 0 |
| SITE04.WITH | SITE06 | -0.119 | 0.012 | -9.507 | 0 | -9.91667 | 0 |
| SITE04.WITH | SITE07 | -0.124 | 0.013 | -9.642 | 0 | -9.53846 | 0 |
| SITE04.WITH | SITE08 | -0.106 | 0.012 | -9.108 | 0 | -8.83333 | 0 |
| SITE04.WITH | SITE09 | -0.128 | 0.013 | -9.765 | 0 | -9.84615 | 0 |
| SITE05.WITH | SITE06 | -0.156 | 0.014 | -10.897 | 0 | -11.1429 | 0 |
| SITE05.WITH | SITE07 | -0.162 | 0.015 | -11.101 | 0 | -10.8 | 0 |
| SITE05.WITH | SITE08 | -0.139 | 0.014 | -10.306 | 0 | -9.92857 | 0 |
| SITE05.WITH | SITE09 | -0.168 | 0.015 | -11.291 | 0 | -11.2 | 0 |
| SITE06.WITH | SITE07 | -0.156 | 0.014 | -10.897 | 0 | -11.1429 | 0 |
| SITE06.WITH | SITE08 | -0.134 | 0.013 | -10.142 | 0 | -10.3077 | 0 |
| SITE06.WITH | SITE09 | -0.162 | 0.015 | -11.076 | 0 | -10.8 | 0 |
| SITE07.WITH | SITE08 | -0.139 | 0.014 | -10.306 | 0 | -9.92857 | 0 |
| SITE07.WITH | SITE09 | -0.168 | 0.015 | -11.291 | 0 | -11.2 | 0 |
| SITE08.WITH | SITE09 | -0.145 | 0.014 | -10.457 | 0 | -10.3571 | 0 |
| Means | CHR | 2.141 | 0.135 | 15.901 | 0 | 15.85926 | 0 |
| Means | AGE | 4.786 | 0.151 | 31.628 | 0 | 31.69536 | 0 |
| Means | FEMALE | 0.92 | 0.044 | 20.664 | 0 | 20.90909 | 0 |
| Means | SITE02 | 0.307 | 0.026 | 11.63 | 0 | 11.80769 | 0 |
| Means | SITE03 | 0.254 | 0.026 | 9.885 | 0 | 9.769231 | 0 |
| Means | SITE04 | 0.307 | 0.026 | 11.63 | 0 | 11.80769 | 0 |
| Means | SITE05 | 0.403 | 0.028 | 14.371 | 0 | 14.39286 | 0 |
| Means | SITE06 | 0.387 | 0.028 | 13.958 | 0 | 13.82143 | 0 |
| Means | SITE07 | 0.403 | 0.028 | 14.371 | 0 | 14.39286 | 0 |
| Means | SITE08 | 0.346 | 0.027 | 12.819 | 0 | 12.81481 | 0 |
| Means | SITE09 | 0.418 | 0.028 | 14.761 | 0 | 14.92857 | 0 |
| Intercepts | SYCOD_CH | 1.118 | 0.32 | 3.497 | 0 | 3.49375 | 0.000476 |
| Intercepts | HIH_CH | -0.056 | 0.048 | -1.161 | 0.246 | -1.16667 | 0.243345 |
| Variances | CHR | 1 | 0 |  |  |  |  |
| Variances | AGE | 1 | 0 |  |  |  |  |
| Variances | FEMALE | 1 | 0 |  |  |  |  |
| Variances | SITE02 | 1 | 0 |  |  |  |  |
| Variances | SITE03 | 1 | 0 |  |  |  |  |
| Variances | SITE04 | 1 | 0 |  |  |  |  |
| Variances | SITE05 | 1 | 0 |  |  |  |  |
| Variances | SITE06 | 1 | 0 |  |  |  |  |
| Variances | SITE07 | 1 | 0 |  |  |  |  |
| Variances | SITE08 | 1 | 0 |  |  |  |  |
| Variances | SITE09 | 1 | 0 |  |  |  |  |
| Variances | LV_HIH | 1 | 0 |  |  |  |  |
| Residual.Variances | SYCOD_CH | 0.204 | 0.029 | 7.104 | 0 | 7.034483 | 2E-12 |
| Residual.Variances | HIH_CH | 0.33 | 0.04 | 8.343 | 0 | 8.25 | 2.22E-16 |
| Residual.Variances | LV_SYCOD | 0.918 | 0.025 | 37.08 | 0 | 36.72 | 0 |
| LV_HIL.BY | HIL_CH | 0.82 | 0.023 | 34.963 | 0 | 35.65217 | 0 |
| LV_SYCOD.BY | SYCOD_CH | 0.893 | 0.016 | 55.224 | 0 | 55.8125 | 0 |
| LV_SYCOD.ON | LV_HIL | 0.39 | 0.197 | 1.981 | 0.048 | 1.979695 | 0.047738 |
| LV_SYCOD.ON | HILXCHR | -0.129 | 0.086 | -1.502 | 0.133 | -1.5 | 0.133614 |
| LV_SYCOD.ON | CHR | -0.052 | 0.049 | -1.069 | 0.285 | -1.06122 | 0.288588 |
| LV_SYCOD.ON | AGE | -0.02 | 0.054 | -0.367 | 0.714 | -0.37037 | 0.711107 |
| LV_SYCOD.ON | FEMALE | -0.073 | 0.057 | -1.289 | 0.197 | -1.2807 | 0.200298 |
| LV_SYCOD.ON | SITE02 | 0.001 | 0.087 | 0.007 | 0.994 | 0.011494 | 0.990829 |
| LV_SYCOD.ON | SITE03 | 0.062 | 0.087 | 0.71 | 0.478 | 0.712644 | 0.476066 |
| LV_SYCOD.ON | SITE04 | -0.15 | 0.06 | -2.522 | 0.012 | -2.5 | 0.012419 |
| LV_SYCOD.ON | SITE05 | 0.105 | 0.078 | 1.342 | 0.18 | 1.346154 | 0.178253 |
| LV_SYCOD.ON | SITE06 | -0.069 | 0.069 | -0.998 | 0.318 | -1 | 0.317311 |
| LV_SYCOD.ON | SITE07 | -0.065 | 0.068 | -0.962 | 0.336 | -0.95588 | 0.339132 |
| LV_SYCOD.ON | SITE08 | -0.173 | 0.069 | -2.532 | 0.011 | -2.50725 | 0.012168 |
| LV_SYCOD.ON | SITE09 | -0.011 | 0.084 | -0.133 | 0.894 | -0.13095 | 0.895813 |
| LV_HIL.WITH | CHR | -0.01 | 0.049 | -0.195 | 0.845 | -0.20408 | 0.83829 |
| LV_HIL.WITH | AGE | 0.012 | 0.059 | 0.208 | 0.836 | 0.20339 | 0.83883 |
| LV_HIL.WITH | FEMALE | -0.115 | 0.061 | -1.87 | 0.061 | -1.88525 | 0.059397 |
| LV_HIL.WITH | SITE02 | -0.033 | 0.064 | -0.52 | 0.603 | -0.51563 | 0.606116 |
| LV_HIL.WITH | SITE03 | 0.095 | 0.073 | 1.304 | 0.192 | 1.30137 | 0.193132 |
| LV_HIL.WITH | SITE04 | 0.104 | 0.05 | 2.101 | 0.036 | 2.08 | 0.037526 |
| LV_HIL.WITH | SITE05 | -0.091 | 0.065 | -1.388 | 0.165 | -1.4 | 0.161513 |
| LV_HIL.WITH | SITE06 | 0.056 | 0.047 | 1.19 | 0.234 | 1.191489 | 0.233462 |
| LV_HIL.WITH | SITE07 | -0.085 | 0.06 | -1.416 | 0.157 | -1.41667 | 0.15658 |
| LV_HIL.WITH | SITE08 | 0.023 | 0.052 | 0.436 | 0.663 | 0.442308 | 0.658267 |
| LV_HIL.WITH | SITE09 | 0.007 | 0.055 | 0.126 | 0.9 | 0.127273 | 0.898725 |
| CHR.WITH | AGE | -0.049 | 0.051 | -0.961 | 0.337 | -0.96078 | 0.336661 |
| CHR.WITH | FEMALE | -0.033 | 0.048 | -0.685 | 0.493 | -0.6875 | 0.491768 |
| CHR.WITH | SITE02 | -0.03 | 0.051 | -0.578 | 0.563 | -0.58824 | 0.556374 |
| CHR.WITH | SITE03 | -0.034 | 0.053 | -0.649 | 0.517 | -0.64151 | 0.521192 |
| CHR.WITH | SITE04 | 0.014 | 0.047 | 0.291 | 0.771 | 0.297872 | 0.765801 |
| CHR.WITH | SITE05 | -0.004 | 0.049 | -0.092 | 0.927 | -0.08163 | 0.934939 |
| CHR.WITH | SITE06 | -0.018 | 0.05 | -0.352 | 0.724 | -0.36 | 0.718847 |
| CHR.WITH | SITE07 | -0.004 | 0.049 | -0.092 | 0.927 | -0.08163 | 0.934939 |
| CHR.WITH | SITE08 | 0.024 | 0.046 | 0.533 | 0.594 | 0.521739 | 0.601852 |
| CHR.WITH | SITE09 | 0.008 | 0.048 | 0.165 | 0.869 | 0.166667 | 0.867632 |
| AGE.WITH | FEMALE | -0.11 | 0.048 | -2.305 | 0.021 | -2.29167 | 0.021925 |
| AGE.WITH | SITE02 | 0.31 | 0.052 | 5.938 | 0 | 5.961538 | 2.5E-09 |
| AGE.WITH | SITE03 | 0 | 0.045 | 0.007 | 0.995 | 0 | 1 |
| AGE.WITH | SITE04 | -0.01 | 0.039 | -0.256 | 0.798 | -0.25641 | 0.797634 |
| AGE.WITH | SITE05 | 0.151 | 0.044 | 3.466 | 0.001 | 3.431818 | 0.0006 |
| AGE.WITH | SITE06 | -0.009 | 0.041 | -0.216 | 0.829 | -0.21951 | 0.826251 |
| AGE.WITH | SITE07 | -0.143 | 0.046 | -3.122 | 0.002 | -3.1087 | 0.001879 |
| AGE.WITH | SITE08 | -0.171 | 0.049 | -3.474 | 0.001 | -3.4898 | 0.000483 |
| AGE.WITH | SITE09 | -0.03 | 0.044 | -0.67 | 0.503 | -0.68182 | 0.495354 |
| FEMALE.WITH | SITE02 | 0.034 | 0.048 | 0.704 | 0.481 | 0.708333 | 0.478738 |
| FEMALE.WITH | SITE03 | -0.018 | 0.048 | -0.373 | 0.709 | -0.375 | 0.70766 |
| FEMALE.WITH | SITE04 | 0.001 | 0.048 | 0.017 | 0.987 | 0.020833 | 0.983379 |
| FEMALE.WITH | SITE05 | 0.007 | 0.048 | 0.143 | 0.886 | 0.145833 | 0.884053 |
| FEMALE.WITH | SITE06 | -0.065 | 0.047 | -1.369 | 0.171 | -1.38298 | 0.166671 |
| FEMALE.WITH | SITE07 | 0.061 | 0.048 | 1.257 | 0.209 | 1.270833 | 0.203788 |
| FEMALE.WITH | SITE08 | -0.001 | 0.048 | -0.023 | 0.981 | -0.02083 | 0.983379 |
| FEMALE.WITH | SITE09 | 0.009 | 0.048 | 0.184 | 0.854 | 0.1875 | 0.851269 |
| SITE02.WITH | SITE03 | -0.078 | 0.01 | -7.839 | 0 | -7.8 | 6.22E-15 |
| SITE02.WITH | SITE04 | -0.094 | 0.011 | -8.641 | 0 | -8.54545 | 0 |
| SITE02.WITH | SITE05 | -0.124 | 0.013 | -9.642 | 0 | -9.53846 | 0 |
| SITE02.WITH | SITE06 | -0.119 | 0.012 | -9.508 | 0 | -9.91667 | 0 |
| SITE02.WITH | SITE07 | -0.124 | 0.013 | -9.642 | 0 | -9.53846 | 0 |
| SITE02.WITH | SITE08 | -0.106 | 0.012 | -9.108 | 0 | -8.83333 | 0 |
| SITE02.WITH | SITE09 | -0.128 | 0.013 | -9.765 | 0 | -9.84615 | 0 |
| SITE03.WITH | SITE04 | -0.078 | 0.01 | -7.839 | 0 | -7.8 | 6.22E-15 |
| SITE03.WITH | SITE05 | -0.102 | 0.012 | -8.563 | 0 | -8.5 | 0 |
| SITE03.WITH | SITE06 | -0.098 | 0.012 | -8.468 | 0 | -8.16667 | 2.22E-16 |
| SITE03.WITH | SITE07 | -0.102 | 0.012 | -8.563 | 0 | -8.5 | 0 |
| SITE03.WITH | SITE08 | -0.088 | 0.011 | -8.183 | 0 | -8 | 1.33E-15 |
| SITE03.WITH | SITE09 | -0.106 | 0.012 | -8.649 | 0 | -8.83333 | 0 |
| SITE04.WITH | SITE05 | -0.124 | 0.013 | -9.642 | 0 | -9.53846 | 0 |
| SITE04.WITH | SITE06 | -0.119 | 0.012 | -9.507 | 0 | -9.91667 | 0 |
| SITE04.WITH | SITE07 | -0.124 | 0.013 | -9.642 | 0 | -9.53846 | 0 |
| SITE04.WITH | SITE08 | -0.106 | 0.012 | -9.108 | 0 | -8.83333 | 0 |
| SITE04.WITH | SITE09 | -0.128 | 0.013 | -9.765 | 0 | -9.84615 | 0 |
| SITE05.WITH | SITE06 | -0.156 | 0.014 | -10.897 | 0 | -11.1429 | 0 |
| SITE05.WITH | SITE07 | -0.162 | 0.015 | -11.101 | 0 | -10.8 | 0 |
| SITE05.WITH | SITE08 | -0.139 | 0.014 | -10.306 | 0 | -9.92857 | 0 |
| SITE05.WITH | SITE09 | -0.168 | 0.015 | -11.291 | 0 | -11.2 | 0 |
| SITE06.WITH | SITE07 | -0.156 | 0.014 | -10.897 | 0 | -11.1429 | 0 |
| SITE06.WITH | SITE08 | -0.134 | 0.013 | -10.142 | 0 | -10.3077 | 0 |
| SITE06.WITH | SITE09 | -0.162 | 0.015 | -11.076 | 0 | -10.8 | 0 |
| SITE07.WITH | SITE08 | -0.139 | 0.014 | -10.306 | 0 | -9.92857 | 0 |
| SITE07.WITH | SITE09 | -0.168 | 0.015 | -11.291 | 0 | -11.2 | 0 |
| SITE08.WITH | SITE09 | -0.145 | 0.014 | -10.457 | 0 | -10.3571 | 0 |
| Means | CHR | 2.141 | 0.135 | 15.901 | 0 | 15.85926 | 0 |
| Means | AGE | 4.786 | 0.151 | 31.628 | 0 | 31.69536 | 0 |
| Means | FEMALE | 0.92 | 0.044 | 20.664 | 0 | 20.90909 | 0 |
| Means | SITE02 | 0.307 | 0.026 | 11.63 | 0 | 11.80769 | 0 |
| Means | SITE03 | 0.254 | 0.026 | 9.885 | 0 | 9.769231 | 0 |
| Means | SITE04 | 0.307 | 0.026 | 11.63 | 0 | 11.80769 | 0 |
| Means | SITE05 | 0.403 | 0.028 | 14.371 | 0 | 14.39286 | 0 |
| Means | SITE06 | 0.387 | 0.028 | 13.958 | 0 | 13.82143 | 0 |
| Means | SITE07 | 0.403 | 0.028 | 14.371 | 0 | 14.39286 | 0 |
| Means | SITE08 | 0.346 | 0.027 | 12.819 | 0 | 12.81481 | 0 |
| Means | SITE09 | 0.418 | 0.028 | 14.761 | 0 | 14.92857 | 0 |
| Intercepts | SYCOD_CH | 1.074 | 0.324 | 3.321 | 0.001 | 3.314815 | 0.000917 |
| Intercepts | HIL_CH | -0.07 | 0.047 | -1.478 | 0.139 | -1.48936 | 0.136392 |
| Variances | CHR | 1 | 0 |  |  |  |  |
| Variances | AGE | 1 | 0 |  |  |  |  |
| Variances | FEMALE | 1 | 0 |  |  |  |  |
| Variances | SITE02 | 1 | 0 |  |  |  |  |
| Variances | SITE03 | 1 | 0 |  |  |  |  |
| Variances | SITE04 | 1 | 0 |  |  |  |  |
| Variances | SITE05 | 1 | 0 |  |  |  |  |
| Variances | SITE06 | 1 | 0 |  |  |  |  |
| Variances | SITE07 | 1 | 0 |  |  |  |  |
| Variances | SITE08 | 1 | 0 |  |  |  |  |
| Variances | SITE09 | 1 | 0 |  |  |  |  |
| Variances | LV_HIL | 1 | 0 |  |  |  |  |
| Residual.Variances | SYCOD_CH | 0.203 | 0.029 | 7.022 | 0 | 7 | 2.56E-12 |
| Residual.Variances | HIL_CH | 0.328 | 0.038 | 8.541 | 0 | 8.631579 | 0 |
| Residual.Variances | LV_SYCOD | 0.893 | 0.033 | 26.913 | 0 | 27.06061 | 0 |
| LV_HIR.BY | HIR_CH | 0.831 | 0.027 | 30.399 | 0 | 30.77778 | 0 |
| LV_SYCOD.BY | SYCOD_CH | 0.892 | 0.016 | 55.41 | 0 | 55.75 | 0 |
| LV_SYCOD.ON | LV_HIR | 0.215 | 0.18 | 1.191 | 0.234 | 1.194444 | 0.232304 |
| LV_SYCOD.ON | HIRXCHR | -0.025 | 0.082 | -0.299 | 0.765 | -0.30488 | 0.760459 |
| LV_SYCOD.ON | CHR | -0.057 | 0.049 | -1.162 | 0.245 | -1.16327 | 0.244722 |
| LV_SYCOD.ON | AGE | -0.026 | 0.052 | -0.488 | 0.626 | -0.5 | 0.617075 |
| LV_SYCOD.ON | FEMALE | -0.083 | 0.055 | -1.519 | 0.129 | -1.50909 | 0.131276 |
| LV_SYCOD.ON | SITE02 | 0.01 | 0.088 | 0.114 | 0.909 | 0.113636 | 0.909526 |
| LV_SYCOD.ON | SITE03 | 0.079 | 0.087 | 0.898 | 0.369 | 0.908046 | 0.363854 |
| LV_SYCOD.ON | SITE04 | -0.13 | 0.057 | -2.277 | 0.023 | -2.2807 | 0.022566 |
| LV_SYCOD.ON | SITE05 | 0.112 | 0.076 | 1.472 | 0.141 | 1.473684 | 0.140567 |
| LV_SYCOD.ON | SITE06 | -0.065 | 0.069 | -0.941 | 0.347 | -0.94203 | 0.346178 |
| LV_SYCOD.ON | SITE07 | -0.074 | 0.069 | -1.074 | 0.283 | -1.07246 | 0.283512 |
| LV_SYCOD.ON | SITE08 | -0.171 | 0.068 | -2.498 | 0.012 | -2.51471 | 0.011913 |
| LV_SYCOD.ON | SITE09 | 0.006 | 0.081 | 0.08 | 0.936 | 0.074074 | 0.940951 |
| LV_HIR.WITH | CHR | 0.009 | 0.045 | 0.198 | 0.843 | 0.2 | 0.841481 |
| LV_HIR.WITH | AGE | -0.022 | 0.055 | -0.405 | 0.686 | -0.4 | 0.689157 |
| LV_HIR.WITH | FEMALE | -0.009 | 0.059 | -0.161 | 0.872 | -0.15254 | 0.878759 |
| LV_HIR.WITH | SITE02 | -0.012 | 0.065 | -0.179 | 0.858 | -0.18462 | 0.853531 |
| LV_HIR.WITH | SITE03 | -0.099 | 0.074 | -1.336 | 0.182 | -1.33784 | 0.180949 |
| LV_HIR.WITH | SITE04 | -0.04 | 0.059 | -0.669 | 0.504 | -0.67797 | 0.497793 |
| LV_HIR.WITH | SITE05 | 0.005 | 0.051 | 0.098 | 0.922 | 0.098039 | 0.921901 |
| LV_HIR.WITH | SITE06 | 0.031 | 0.047 | 0.66 | 0.509 | 0.659574 | 0.509527 |
| LV_HIR.WITH | SITE07 | 0.069 | 0.069 | 0.998 | 0.318 | 1 | 0.317311 |
| LV_HIR.WITH | SITE08 | 0.034 | 0.054 | 0.628 | 0.53 | 0.62963 | 0.528937 |
| LV_HIR.WITH | SITE09 | -0.037 | 0.055 | -0.677 | 0.498 | -0.67273 | 0.501121 |
| CHR.WITH | AGE | -0.049 | 0.051 | -0.961 | 0.337 | -0.96078 | 0.336661 |
| CHR.WITH | FEMALE | -0.033 | 0.048 | -0.685 | 0.493 | -0.6875 | 0.491768 |
| CHR.WITH | SITE02 | -0.03 | 0.051 | -0.578 | 0.563 | -0.58824 | 0.556374 |
| CHR.WITH | SITE03 | -0.034 | 0.053 | -0.649 | 0.517 | -0.64151 | 0.521192 |
| CHR.WITH | SITE04 | 0.014 | 0.047 | 0.291 | 0.771 | 0.297872 | 0.765801 |
| CHR.WITH | SITE05 | -0.004 | 0.049 | -0.092 | 0.927 | -0.08163 | 0.934939 |
| CHR.WITH | SITE06 | -0.018 | 0.05 | -0.352 | 0.724 | -0.36 | 0.718847 |
| CHR.WITH | SITE07 | -0.004 | 0.049 | -0.092 | 0.927 | -0.08163 | 0.934939 |
| CHR.WITH | SITE08 | 0.024 | 0.046 | 0.533 | 0.594 | 0.521739 | 0.601852 |
| CHR.WITH | SITE09 | 0.008 | 0.048 | 0.165 | 0.869 | 0.166667 | 0.867632 |
| AGE.WITH | FEMALE | -0.11 | 0.048 | -2.305 | 0.021 | -2.29167 | 0.021925 |
| AGE.WITH | SITE02 | 0.31 | 0.052 | 5.938 | 0 | 5.961538 | 2.5E-09 |
| AGE.WITH | SITE03 | 0 | 0.045 | 0.007 | 0.995 | 0 | 1 |
| AGE.WITH | SITE04 | -0.01 | 0.039 | -0.256 | 0.798 | -0.25641 | 0.797634 |
| AGE.WITH | SITE05 | 0.151 | 0.044 | 3.466 | 0.001 | 3.431818 | 0.0006 |
| AGE.WITH | SITE06 | -0.009 | 0.041 | -0.216 | 0.829 | -0.21951 | 0.826251 |
| AGE.WITH | SITE07 | -0.143 | 0.046 | -3.122 | 0.002 | -3.1087 | 0.001879 |
| AGE.WITH | SITE08 | -0.171 | 0.049 | -3.474 | 0.001 | -3.4898 | 0.000483 |
| AGE.WITH | SITE09 | -0.03 | 0.044 | -0.67 | 0.503 | -0.68182 | 0.495354 |
| FEMALE.WITH | SITE02 | 0.034 | 0.048 | 0.704 | 0.481 | 0.708333 | 0.478738 |
| FEMALE.WITH | SITE03 | -0.018 | 0.048 | -0.373 | 0.709 | -0.375 | 0.70766 |
| FEMALE.WITH | SITE04 | 0.001 | 0.048 | 0.017 | 0.987 | 0.020833 | 0.983379 |
| FEMALE.WITH | SITE05 | 0.007 | 0.048 | 0.143 | 0.886 | 0.145833 | 0.884053 |
| FEMALE.WITH | SITE06 | -0.065 | 0.047 | -1.369 | 0.171 | -1.38298 | 0.166671 |
| FEMALE.WITH | SITE07 | 0.061 | 0.048 | 1.257 | 0.209 | 1.270833 | 0.203788 |
| FEMALE.WITH | SITE08 | -0.001 | 0.048 | -0.023 | 0.981 | -0.02083 | 0.983379 |
| FEMALE.WITH | SITE09 | 0.009 | 0.048 | 0.184 | 0.854 | 0.1875 | 0.851269 |
| SITE02.WITH | SITE03 | -0.078 | 0.01 | -7.839 | 0 | -7.8 | 6.22E-15 |
| SITE02.WITH | SITE04 | -0.094 | 0.011 | -8.641 | 0 | -8.54545 | 0 |
| SITE02.WITH | SITE05 | -0.124 | 0.013 | -9.642 | 0 | -9.53846 | 0 |
| SITE02.WITH | SITE06 | -0.119 | 0.012 | -9.507 | 0 | -9.91667 | 0 |
| SITE02.WITH | SITE07 | -0.124 | 0.013 | -9.642 | 0 | -9.53846 | 0 |
| SITE02.WITH | SITE08 | -0.106 | 0.012 | -9.108 | 0 | -8.83333 | 0 |
| SITE02.WITH | SITE09 | -0.128 | 0.013 | -9.765 | 0 | -9.84615 | 0 |
| SITE03.WITH | SITE04 | -0.078 | 0.01 | -7.839 | 0 | -7.8 | 6.22E-15 |
| SITE03.WITH | SITE05 | -0.102 | 0.012 | -8.563 | 0 | -8.5 | 0 |
| SITE03.WITH | SITE06 | -0.098 | 0.012 | -8.468 | 0 | -8.16667 | 2.22E-16 |
| SITE03.WITH | SITE07 | -0.102 | 0.012 | -8.563 | 0 | -8.5 | 0 |
| SITE03.WITH | SITE08 | -0.088 | 0.011 | -8.183 | 0 | -8 | 1.33E-15 |
| SITE03.WITH | SITE09 | -0.106 | 0.012 | -8.649 | 0 | -8.83333 | 0 |
| SITE04.WITH | SITE05 | -0.124 | 0.013 | -9.642 | 0 | -9.53846 | 0 |
| SITE04.WITH | SITE06 | -0.119 | 0.012 | -9.507 | 0 | -9.91667 | 0 |
| SITE04.WITH | SITE07 | -0.124 | 0.013 | -9.642 | 0 | -9.53846 | 0 |
| SITE04.WITH | SITE08 | -0.106 | 0.012 | -9.108 | 0 | -8.83333 | 0 |
| SITE04.WITH | SITE09 | -0.128 | 0.013 | -9.765 | 0 | -9.84615 | 0 |
| SITE05.WITH | SITE06 | -0.156 | 0.014 | -10.897 | 0 | -11.1429 | 0 |
| SITE05.WITH | SITE07 | -0.162 | 0.015 | -11.101 | 0 | -10.8 | 0 |
| SITE05.WITH | SITE08 | -0.139 | 0.014 | -10.306 | 0 | -9.92857 | 0 |
| SITE05.WITH | SITE09 | -0.168 | 0.015 | -11.291 | 0 | -11.2 | 0 |
| SITE06.WITH | SITE07 | -0.156 | 0.014 | -10.897 | 0 | -11.1429 | 0 |
| SITE06.WITH | SITE08 | -0.134 | 0.013 | -10.142 | 0 | -10.3077 | 0 |
| SITE06.WITH | SITE09 | -0.162 | 0.015 | -11.076 | 0 | -10.8 | 0 |
| SITE07.WITH | SITE08 | -0.139 | 0.014 | -10.306 | 0 | -9.92857 | 0 |
| SITE07.WITH | SITE09 | -0.168 | 0.015 | -11.291 | 0 | -11.2 | 0 |
| SITE08.WITH | SITE09 | -0.145 | 0.014 | -10.457 | 0 | -10.3571 | 0 |
| Means | CHR | 2.141 | 0.135 | 15.901 | 0 | 15.85926 | 0 |
| Means | AGE | 4.786 | 0.151 | 31.627 | 0 | 31.69536 | 0 |
| Means | FEMALE | 0.92 | 0.044 | 20.664 | 0 | 20.90909 | 0 |
| Means | SITE02 | 0.307 | 0.026 | 11.63 | 0 | 11.80769 | 0 |
| Means | SITE03 | 0.254 | 0.026 | 9.886 | 0 | 9.769231 | 0 |
| Means | SITE04 | 0.307 | 0.026 | 11.63 | 0 | 11.80769 | 0 |
| Means | SITE05 | 0.403 | 0.028 | 14.371 | 0 | 14.39286 | 0 |
| Means | SITE06 | 0.387 | 0.028 | 13.959 | 0 | 13.82143 | 0 |
| Means | SITE07 | 0.403 | 0.028 | 14.365 | 0 | 14.39286 | 0 |
| Means | SITE08 | 0.346 | 0.027 | 12.819 | 0 | 12.81481 | 0 |
| Means | SITE09 | 0.418 | 0.028 | 14.761 | 0 | 14.92857 | 0 |
| Intercepts | SYCOD_CH | 1.099 | 0.32 | 3.437 | 0.001 | 3.434375 | 0.000594 |
| Intercepts | HIR_CH | 0.01 | 0.048 | 0.209 | 0.835 | 0.208333 | 0.834969 |
| Variances | CHR | 1 | 0 |  |  |  |  |
| Variances | AGE | 1 | 0 |  |  |  |  |
| Variances | FEMALE | 1 | 0 |  |  |  |  |
| Variances | SITE02 | 1 | 0 |  |  |  |  |
| Variances | SITE03 | 1 | 0 |  |  |  |  |
| Variances | SITE04 | 1 | 0 |  |  |  |  |
| Variances | SITE05 | 1 | 0 |  |  |  |  |
| Variances | SITE06 | 1 | 0 |  |  |  |  |
| Variances | SITE07 | 1 | 0 |  |  |  |  |
| Variances | SITE08 | 1 | 0 |  |  |  |  |
| Variances | SITE09 | 1 | 0 |  |  |  |  |
| Variances | LV_HIR | 1 | 0 |  |  |  |  |
| Residual.Variances | SYCOD_CH | 0.203 | 0.029 | 7.077 | 0 | 7 | 2.56E-12 |
| Residual.Variances | HIR_CH | 0.31 | 0.045 | 6.823 | 0 | 6.888889 | 5.62E-12 |
| Residual.Variances | LV_SYCOD | 0.893 | 0.033 | 27.25 | 0 | 27.06061 | 0 |
| LV_HIH.BY | HIH_CH | 0.82 | 0.024 | 34.099 | 0 | 34.16667 | 0 |
| LV_VEMEM.BY | VEMEM_CH | 0.74 | 0.043 | 17.315 | 0 | 17.2093 | 0 |
| LV_VEMEM.ON | LV_HIH | 0.113 | 0.095 | 1.188 | 0.235 | 1.189474 | 0.234253 |
| LV_VEMEM.ON | HIHXCHR | -0.101 | 0.053 | -1.893 | 0.058 | -1.90566 | 0.056694 |
| LV_VEMEM.ON | CHR | -0.05 | 0.049 | -1.02 | 0.308 | -1.02041 | 0.307535 |
| LV_VEMEM.ON | AGE | 0.036 | 0.062 | 0.578 | 0.563 | 0.580645 | 0.56148 |
| LV_VEMEM.ON | FEMALE | -0.101 | 0.061 | -1.656 | 0.098 | -1.65574 | 0.097775 |
| LV_VEMEM.ON | SITE02 | 0.041 | 0.089 | 0.459 | 0.646 | 0.460674 | 0.645032 |
| LV_VEMEM.ON | SITE03 | 0.002 | 0.069 | 0.031 | 0.975 | 0.028986 | 0.976876 |
| LV_VEMEM.ON | SITE04 | -0.171 | 0.097 | -1.767 | 0.077 | -1.76289 | 0.07792 |
| LV_VEMEM.ON | SITE05 | 0.038 | 0.081 | 0.476 | 0.634 | 0.469136 | 0.638973 |
| LV_VEMEM.ON | SITE06 | 0.121 | 0.088 | 1.374 | 0.169 | 1.375 | 0.169131 |
| LV_VEMEM.ON | SITE07 | 0.075 | 0.077 | 0.975 | 0.33 | 0.974026 | 0.330044 |
| LV_VEMEM.ON | SITE08 | 0.057 | 0.088 | 0.648 | 0.517 | 0.647727 | 0.517161 |
| LV_VEMEM.ON | SITE09 | 0.139 | 0.092 | 1.512 | 0.13 | 1.51087 | 0.130822 |
| LV_HIH.WITH | CHR | -0.006 | 0.045 | -0.14 | 0.889 | -0.13333 | 0.89393 |
| LV_HIH.WITH | AGE | 0.038 | 0.059 | 0.644 | 0.52 | 0.644068 | 0.519531 |
| LV_HIH.WITH | FEMALE | -0.072 | 0.059 | -1.223 | 0.221 | -1.22034 | 0.222336 |
| LV_HIH.WITH | SITE02 | 0.045 | 0.066 | 0.682 | 0.495 | 0.681818 | 0.495354 |
| LV_HIH.WITH | SITE03 | -0.004 | 0.09 | -0.044 | 0.965 | -0.04444 | 0.96455 |
| LV_HIH.WITH | SITE04 | 0.072 | 0.045 | 1.613 | 0.107 | 1.6 | 0.109599 |
| LV_HIH.WITH | SITE05 | -0.057 | 0.062 | -0.917 | 0.359 | -0.91935 | 0.35791 |
| LV_HIH.WITH | SITE06 | 0.079 | 0.043 | 1.823 | 0.068 | 1.837209 | 0.066179 |
| LV_HIH.WITH | SITE07 | -0.007 | 0.058 | -0.119 | 0.905 | -0.12069 | 0.903937 |
| LV_HIH.WITH | SITE08 | -0.033 | 0.058 | -0.575 | 0.565 | -0.56897 | 0.56938 |
| LV_HIH.WITH | SITE09 | -0.045 | 0.058 | -0.77 | 0.441 | -0.77586 | 0.43783 |
| CHR.WITH | AGE | -0.049 | 0.051 | -0.954 | 0.34 | -0.96078 | 0.336661 |
| CHR.WITH | FEMALE | -0.027 | 0.048 | -0.567 | 0.57 | -0.5625 | 0.573775 |
| CHR.WITH | SITE02 | -0.045 | 0.053 | -0.849 | 0.396 | -0.84906 | 0.39585 |
| CHR.WITH | SITE03 | -0.033 | 0.052 | -0.619 | 0.536 | -0.63462 | 0.525679 |
| CHR.WITH | SITE04 | 0.015 | 0.046 | 0.331 | 0.741 | 0.326087 | 0.744359 |
| CHR.WITH | SITE05 | -0.002 | 0.048 | -0.042 | 0.967 | -0.04167 | 0.966764 |
| CHR.WITH | SITE06 | -0.015 | 0.049 | -0.305 | 0.76 | -0.30612 | 0.759511 |
| CHR.WITH | SITE07 | -0.002 | 0.048 | -0.042 | 0.967 | -0.04167 | 0.966764 |
| CHR.WITH | SITE08 | 0.026 | 0.045 | 0.579 | 0.563 | 0.577778 | 0.563414 |
| CHR.WITH | SITE09 | 0.007 | 0.048 | 0.153 | 0.878 | 0.145833 | 0.884053 |
| AGE.WITH | FEMALE | -0.111 | 0.048 | -2.328 | 0.02 | -2.3125 | 0.02075 |
| AGE.WITH | SITE02 | 0.306 | 0.052 | 5.903 | 0 | 5.884615 | 3.99E-09 |
| AGE.WITH | SITE03 | 0 | 0.045 | 0 | 1 | 0 | 1 |
| AGE.WITH | SITE04 | -0.01 | 0.039 | -0.265 | 0.791 | -0.25641 | 0.797634 |
| AGE.WITH | SITE05 | 0.151 | 0.044 | 3.456 | 0.001 | 3.431818 | 0.0006 |
| AGE.WITH | SITE06 | -0.009 | 0.041 | -0.227 | 0.82 | -0.21951 | 0.826251 |
| AGE.WITH | SITE07 | -0.144 | 0.046 | -3.132 | 0.002 | -3.13043 | 0.001745 |
| AGE.WITH | SITE08 | -0.172 | 0.049 | -3.482 | 0 | -3.5102 | 0.000448 |
| AGE.WITH | SITE09 | -0.027 | 0.045 | -0.613 | 0.54 | -0.6 | 0.548506 |
| FEMALE.WITH | SITE02 | 0.026 | 0.048 | 0.54 | 0.589 | 0.541667 | 0.588048 |
| FEMALE.WITH | SITE03 | -0.018 | 0.048 | -0.373 | 0.709 | -0.375 | 0.70766 |
| FEMALE.WITH | SITE04 | 0.001 | 0.048 | 0.017 | 0.987 | 0.020833 | 0.983379 |
| FEMALE.WITH | SITE05 | 0.007 | 0.048 | 0.143 | 0.886 | 0.145833 | 0.884053 |
| FEMALE.WITH | SITE06 | -0.065 | 0.047 | -1.369 | 0.171 | -1.38298 | 0.166671 |
| FEMALE.WITH | SITE07 | 0.061 | 0.048 | 1.257 | 0.209 | 1.270833 | 0.203788 |
| FEMALE.WITH | SITE08 | -0.001 | 0.048 | -0.023 | 0.981 | -0.02083 | 0.983379 |
| FEMALE.WITH | SITE09 | 0.015 | 0.048 | 0.311 | 0.756 | 0.3125 | 0.754661 |
| SITE02.WITH | SITE03 | -0.079 | 0.01 | -7.883 | 0 | -7.9 | 2.89E-15 |
| SITE02.WITH | SITE04 | -0.096 | 0.011 | -8.699 | 0 | -8.72727 | 0 |
| SITE02.WITH | SITE05 | -0.125 | 0.013 | -9.724 | 0 | -9.61538 | 0 |
| SITE02.WITH | SITE06 | -0.12 | 0.013 | -9.586 | 0 | -9.23077 | 0 |
| SITE02.WITH | SITE07 | -0.125 | 0.013 | -9.724 | 0 | -9.61538 | 0 |
| SITE02.WITH | SITE08 | -0.108 | 0.012 | -9.177 | 0 | -9 | 0 |
| SITE02.WITH | SITE09 | -0.129 | 0.013 | -9.819 | 0 | -9.92308 | 0 |
| SITE03.WITH | SITE04 | -0.078 | 0.01 | -7.839 | 0 | -7.8 | 6.22E-15 |
| SITE03.WITH | SITE05 | -0.102 | 0.012 | -8.563 | 0 | -8.5 | 0 |
| SITE03.WITH | SITE06 | -0.098 | 0.012 | -8.468 | 0 | -8.16667 | 2.22E-16 |
| SITE03.WITH | SITE07 | -0.102 | 0.012 | -8.563 | 0 | -8.5 | 0 |
| SITE03.WITH | SITE08 | -0.088 | 0.011 | -8.183 | 0 | -8 | 1.33E-15 |
| SITE03.WITH | SITE09 | -0.105 | 0.012 | -8.628 | 0 | -8.75 | 0 |
| SITE04.WITH | SITE05 | -0.124 | 0.013 | -9.642 | 0 | -9.53846 | 0 |
| SITE04.WITH | SITE06 | -0.119 | 0.012 | -9.507 | 0 | -9.91667 | 0 |
| SITE04.WITH | SITE07 | -0.124 | 0.013 | -9.642 | 0 | -9.53846 | 0 |
| SITE04.WITH | SITE08 | -0.106 | 0.012 | -9.108 | 0 | -8.83333 | 0 |
| SITE04.WITH | SITE09 | -0.127 | 0.013 | -9.735 | 0 | -9.76923 | 0 |
| SITE05.WITH | SITE06 | -0.156 | 0.014 | -10.897 | 0 | -11.1429 | 0 |
| SITE05.WITH | SITE07 | -0.162 | 0.015 | -11.101 | 0 | -10.8 | 0 |
| SITE05.WITH | SITE08 | -0.139 | 0.014 | -10.306 | 0 | -9.92857 | 0 |
| SITE05.WITH | SITE09 | -0.167 | 0.015 | -11.245 | 0 | -11.1333 | 0 |
| SITE06.WITH | SITE07 | -0.156 | 0.014 | -10.897 | 0 | -11.1429 | 0 |
| SITE06.WITH | SITE08 | -0.134 | 0.013 | -10.142 | 0 | -10.3077 | 0 |
| SITE06.WITH | SITE09 | -0.16 | 0.015 | -11.033 | 0 | -10.6667 | 0 |
| SITE07.WITH | SITE08 | -0.139 | 0.014 | -10.306 | 0 | -9.92857 | 0 |
| SITE07.WITH | SITE09 | -0.167 | 0.015 | -11.245 | 0 | -11.1333 | 0 |
| SITE08.WITH | SITE09 | -0.143 | 0.014 | -10.42 | 0 | -10.2143 | 0 |
| Means | CHR | 2.124 | 0.133 | 15.982 | 0 | 15.96992 | 0 |
| Means | AGE | 4.789 | 0.152 | 31.606 | 0 | 31.50658 | 0 |
| Means | FEMALE | 0.92 | 0.044 | 20.666 | 0 | 20.90909 | 0 |
| Means | SITE02 | 0.311 | 0.026 | 11.771 | 0 | 11.96154 | 0 |
| Means | SITE03 | 0.254 | 0.026 | 9.885 | 0 | 9.769231 | 0 |
| Means | SITE04 | 0.307 | 0.026 | 11.63 | 0 | 11.80769 | 0 |
| Means | SITE05 | 0.403 | 0.028 | 14.371 | 0 | 14.39286 | 0 |
| Means | SITE06 | 0.387 | 0.028 | 13.957 | 0 | 13.82143 | 0 |
| Means | SITE07 | 0.403 | 0.028 | 14.371 | 0 | 14.39286 | 0 |
| Means | SITE08 | 0.346 | 0.027 | 12.819 | 0 | 12.81481 | 0 |
| Means | SITE09 | 0.414 | 0.028 | 14.666 | 0 | 14.78571 | 0 |
| Intercepts | VEMEM_CH | 0.256 | 0.302 | 0.849 | 0.396 | 0.847682 | 0.396615 |
| Intercepts | HIH_CH | -0.059 | 0.048 | -1.233 | 0.218 | -1.22917 | 0.219009 |
| Variances | CHR | 1 | 0 |  |  |  |  |
| Variances | AGE | 1 | 0 |  |  |  |  |
| Variances | FEMALE | 1 | 0 |  |  |  |  |
| Variances | SITE02 | 1 | 0 |  |  |  |  |
| Variances | SITE03 | 1 | 0 |  |  |  |  |
| Variances | SITE04 | 1 | 0 |  |  |  |  |
| Variances | SITE05 | 1 | 0 |  |  |  |  |
| Variances | SITE06 | 1 | 0 |  |  |  |  |
| Variances | SITE07 | 1 | 0 |  |  |  |  |
| Variances | SITE08 | 1 | 0 |  |  |  |  |
| Variances | SITE09 | 1 | 0 |  |  |  |  |
| Variances | LV_HIH | 1 | 0 |  |  |  |  |
| Residual.Variances | VEMEM_CH | 0.453 | 0.063 | 7.167 | 0 | 7.190476 | 6.46E-13 |
| Residual.Variances | HIH_CH | 0.328 | 0.039 | 8.325 | 0 | 8.410256 | 0 |
| Residual.Variances | LV_VEMEM | 0.896 | 0.044 | 20.335 | 0 | 20.36364 | 0 |
| LV_HIL.BY | HIL_CH | 0.82 | 0.023 | 35.108 | 0 | 35.65217 | 0 |
| LV_VEMEM.BY | VEMEM_CH | 0.739 | 0.043 | 17.342 | 0 | 17.18605 | 0 |
| LV_VEMEM.ON | LV_HIL | -0.018 | 0.136 | -0.135 | 0.893 | -0.13235 | 0.894705 |
| LV_VEMEM.ON | HILXCHR | -0.016 | 0.07 | -0.231 | 0.818 | -0.22857 | 0.819202 |
| LV_VEMEM.ON | CHR | -0.051 | 0.048 | -1.05 | 0.294 | -1.0625 | 0.288009 |
| LV_VEMEM.ON | AGE | 0.031 | 0.062 | 0.502 | 0.616 | 0.5 | 0.617075 |
| LV_VEMEM.ON | FEMALE | -0.096 | 0.064 | -1.515 | 0.13 | -1.5 | 0.133614 |
| LV_VEMEM.ON | SITE02 | 0.039 | 0.087 | 0.452 | 0.651 | 0.448276 | 0.653954 |
| LV_VEMEM.ON | SITE03 | 0.005 | 0.075 | 0.063 | 0.949 | 0.066667 | 0.946847 |
| LV_VEMEM.ON | SITE04 | -0.172 | 0.097 | -1.77 | 0.077 | -1.7732 | 0.076196 |
| LV_VEMEM.ON | SITE05 | 0.042 | 0.079 | 0.528 | 0.597 | 0.531646 | 0.594971 |
| LV_VEMEM.ON | SITE06 | 0.114 | 0.087 | 1.316 | 0.188 | 1.310345 | 0.190079 |
| LV_VEMEM.ON | SITE07 | 0.067 | 0.078 | 0.863 | 0.388 | 0.858974 | 0.390355 |
| LV_VEMEM.ON | SITE08 | 0.063 | 0.089 | 0.71 | 0.478 | 0.707865 | 0.479029 |
| LV_VEMEM.ON | SITE09 | 0.142 | 0.093 | 1.528 | 0.127 | 1.526882 | 0.12679 |
| LV_HIL.WITH | CHR | -0.009 | 0.049 | -0.178 | 0.858 | -0.18367 | 0.85427 |
| LV_HIL.WITH | AGE | 0.014 | 0.059 | 0.233 | 0.816 | 0.237288 | 0.812433 |
| LV_HIL.WITH | FEMALE | -0.108 | 0.061 | -1.767 | 0.077 | -1.77049 | 0.076645 |
| LV_HIL.WITH | SITE02 | -0.037 | 0.062 | -0.592 | 0.554 | -0.59677 | 0.550658 |
| LV_HIL.WITH | SITE03 | 0.098 | 0.073 | 1.34 | 0.18 | 1.342466 | 0.179445 |
| LV_HIL.WITH | SITE04 | 0.104 | 0.05 | 2.086 | 0.037 | 2.08 | 0.037526 |
| LV_HIL.WITH | SITE05 | -0.086 | 0.066 | -1.318 | 0.187 | -1.30303 | 0.192564 |
| LV_HIL.WITH | SITE06 | 0.054 | 0.047 | 1.133 | 0.257 | 1.148936 | 0.250582 |
| LV_HIL.WITH | SITE07 | -0.082 | 0.06 | -1.379 | 0.168 | -1.36667 | 0.17173 |
| LV_HIL.WITH | SITE08 | 0.024 | 0.052 | 0.457 | 0.647 | 0.461538 | 0.644412 |
| LV_HIL.WITH | SITE09 | 0.003 | 0.055 | 0.052 | 0.958 | 0.054545 | 0.956501 |
| CHR.WITH | AGE | -0.049 | 0.051 | -0.954 | 0.34 | -0.96078 | 0.336661 |
| CHR.WITH | FEMALE | -0.027 | 0.048 | -0.567 | 0.57 | -0.5625 | 0.573775 |
| CHR.WITH | SITE02 | -0.045 | 0.053 | -0.849 | 0.396 | -0.84906 | 0.39585 |
| CHR.WITH | SITE03 | -0.033 | 0.052 | -0.619 | 0.536 | -0.63462 | 0.525679 |
| CHR.WITH | SITE04 | 0.015 | 0.046 | 0.331 | 0.741 | 0.326087 | 0.744359 |
| CHR.WITH | SITE05 | -0.002 | 0.048 | -0.042 | 0.967 | -0.04167 | 0.966764 |
| CHR.WITH | SITE06 | -0.015 | 0.049 | -0.305 | 0.76 | -0.30612 | 0.759511 |
| CHR.WITH | SITE07 | -0.002 | 0.048 | -0.042 | 0.967 | -0.04167 | 0.966764 |
| CHR.WITH | SITE08 | 0.026 | 0.045 | 0.579 | 0.563 | 0.577778 | 0.563414 |
| CHR.WITH | SITE09 | 0.007 | 0.048 | 0.153 | 0.878 | 0.145833 | 0.884053 |
| AGE.WITH | FEMALE | -0.111 | 0.048 | -2.328 | 0.02 | -2.3125 | 0.02075 |
| AGE.WITH | SITE02 | 0.306 | 0.052 | 5.903 | 0 | 5.884615 | 3.99E-09 |
| AGE.WITH | SITE03 | 0 | 0.045 | 0 | 1 | 0 | 1 |
| AGE.WITH | SITE04 | -0.01 | 0.039 | -0.265 | 0.791 | -0.25641 | 0.797634 |
| AGE.WITH | SITE05 | 0.151 | 0.044 | 3.456 | 0.001 | 3.431818 | 0.0006 |
| AGE.WITH | SITE06 | -0.009 | 0.041 | -0.227 | 0.82 | -0.21951 | 0.826251 |
| AGE.WITH | SITE07 | -0.144 | 0.046 | -3.132 | 0.002 | -3.13043 | 0.001745 |
| AGE.WITH | SITE08 | -0.172 | 0.049 | -3.482 | 0 | -3.5102 | 0.000448 |
| AGE.WITH | SITE09 | -0.027 | 0.045 | -0.613 | 0.54 | -0.6 | 0.548506 |
| FEMALE.WITH | SITE02 | 0.026 | 0.048 | 0.54 | 0.589 | 0.541667 | 0.588048 |
| FEMALE.WITH | SITE03 | -0.018 | 0.048 | -0.373 | 0.709 | -0.375 | 0.70766 |
| FEMALE.WITH | SITE04 | 0.001 | 0.048 | 0.017 | 0.987 | 0.020833 | 0.983379 |
| FEMALE.WITH | SITE05 | 0.007 | 0.048 | 0.143 | 0.886 | 0.145833 | 0.884053 |
| FEMALE.WITH | SITE06 | -0.065 | 0.047 | -1.369 | 0.171 | -1.38298 | 0.166671 |
| FEMALE.WITH | SITE07 | 0.061 | 0.048 | 1.257 | 0.209 | 1.270833 | 0.203788 |
| FEMALE.WITH | SITE08 | -0.001 | 0.048 | -0.023 | 0.981 | -0.02083 | 0.983379 |
| FEMALE.WITH | SITE09 | 0.015 | 0.048 | 0.311 | 0.756 | 0.3125 | 0.754661 |
| SITE02.WITH | SITE03 | -0.079 | 0.01 | -7.883 | 0 | -7.9 | 2.89E-15 |
| SITE02.WITH | SITE04 | -0.096 | 0.011 | -8.699 | 0 | -8.72727 | 0 |
| SITE02.WITH | SITE05 | -0.125 | 0.013 | -9.724 | 0 | -9.61538 | 0 |
| SITE02.WITH | SITE06 | -0.12 | 0.013 | -9.586 | 0 | -9.23077 | 0 |
| SITE02.WITH | SITE07 | -0.125 | 0.013 | -9.724 | 0 | -9.61538 | 0 |
| SITE02.WITH | SITE08 | -0.108 | 0.012 | -9.177 | 0 | -9 | 0 |
| SITE02.WITH | SITE09 | -0.129 | 0.013 | -9.819 | 0 | -9.92308 | 0 |
| SITE03.WITH | SITE04 | -0.078 | 0.01 | -7.839 | 0 | -7.8 | 6.22E-15 |
| SITE03.WITH | SITE05 | -0.102 | 0.012 | -8.563 | 0 | -8.5 | 0 |
| SITE03.WITH | SITE06 | -0.098 | 0.012 | -8.468 | 0 | -8.16667 | 2.22E-16 |
| SITE03.WITH | SITE07 | -0.102 | 0.012 | -8.563 | 0 | -8.5 | 0 |
| SITE03.WITH | SITE08 | -0.088 | 0.011 | -8.183 | 0 | -8 | 1.33E-15 |
| SITE03.WITH | SITE09 | -0.105 | 0.012 | -8.628 | 0 | -8.75 | 0 |
| SITE04.WITH | SITE05 | -0.124 | 0.013 | -9.642 | 0 | -9.53846 | 0 |
| SITE04.WITH | SITE06 | -0.119 | 0.012 | -9.507 | 0 | -9.91667 | 0 |
| SITE04.WITH | SITE07 | -0.124 | 0.013 | -9.642 | 0 | -9.53846 | 0 |
| SITE04.WITH | SITE08 | -0.106 | 0.012 | -9.108 | 0 | -8.83333 | 0 |
| SITE04.WITH | SITE09 | -0.127 | 0.013 | -9.735 | 0 | -9.76923 | 0 |
| SITE05.WITH | SITE06 | -0.156 | 0.014 | -10.897 | 0 | -11.1429 | 0 |
| SITE05.WITH | SITE07 | -0.162 | 0.015 | -11.101 | 0 | -10.8 | 0 |
| SITE05.WITH | SITE08 | -0.139 | 0.014 | -10.306 | 0 | -9.92857 | 0 |
| SITE05.WITH | SITE09 | -0.167 | 0.015 | -11.245 | 0 | -11.1333 | 0 |
| SITE06.WITH | SITE07 | -0.156 | 0.014 | -10.897 | 0 | -11.1429 | 0 |
| SITE06.WITH | SITE08 | -0.134 | 0.013 | -10.142 | 0 | -10.3077 | 0 |
| SITE06.WITH | SITE09 | -0.16 | 0.015 | -11.033 | 0 | -10.6667 | 0 |
| SITE07.WITH | SITE08 | -0.139 | 0.014 | -10.306 | 0 | -9.92857 | 0 |
| SITE07.WITH | SITE09 | -0.167 | 0.015 | -11.245 | 0 | -11.1333 | 0 |
| SITE08.WITH | SITE09 | -0.143 | 0.014 | -10.42 | 0 | -10.2143 | 0 |
| Means | CHR | 2.124 | 0.133 | 15.982 | 0 | 15.96992 | 0 |
| Means | AGE | 4.789 | 0.152 | 31.606 | 0 | 31.50658 | 0 |
| Means | FEMALE | 0.92 | 0.044 | 20.667 | 0 | 20.90909 | 0 |
| Means | SITE02 | 0.311 | 0.026 | 11.772 | 0 | 11.96154 | 0 |
| Means | SITE03 | 0.254 | 0.026 | 9.884 | 0 | 9.769231 | 0 |
| Means | SITE04 | 0.307 | 0.026 | 11.629 | 0 | 11.80769 | 0 |
| Means | SITE05 | 0.403 | 0.028 | 14.372 | 0 | 14.39286 | 0 |
| Means | SITE06 | 0.387 | 0.028 | 13.957 | 0 | 13.82143 | 0 |
| Means | SITE07 | 0.403 | 0.028 | 14.372 | 0 | 14.39286 | 0 |
| Means | SITE08 | 0.346 | 0.027 | 12.818 | 0 | 12.81481 | 0 |
| Means | SITE09 | 0.414 | 0.028 | 14.665 | 0 | 14.78571 | 0 |
| Intercepts | VEMEM_CH | 0.273 | 0.301 | 0.908 | 0.364 | 0.906977 | 0.364419 |
| Intercepts | HIL_CH | -0.073 | 0.047 | -1.548 | 0.122 | -1.55319 | 0.120377 |
| Variances | CHR | 1 | 0 |  |  |  |  |
| Variances | AGE | 1 | 0 |  |  |  |  |
| Variances | FEMALE | 1 | 0 |  |  |  |  |
| Variances | SITE02 | 1 | 0 |  |  |  |  |
| Variances | SITE03 | 1 | 0 |  |  |  |  |
| Variances | SITE04 | 1 | 0 |  |  |  |  |
| Variances | SITE05 | 1 | 0 |  |  |  |  |
| Variances | SITE06 | 1 | 0 |  |  |  |  |
| Variances | SITE07 | 1 | 0 |  |  |  |  |
| Variances | SITE08 | 1 | 0 |  |  |  |  |
| Variances | SITE09 | 1 | 0 |  |  |  |  |
| Variances | LV_HIL | 1 | 0 |  |  |  |  |
| Residual.Variances | VEMEM_CH | 0.453 | 0.063 | 7.186 | 0 | 7.190476 | 6.46E-13 |
| Residual.Variances | HIL_CH | 0.327 | 0.038 | 8.529 | 0 | 8.605263 | 0 |
| Residual.Variances | LV_VEMEM | 0.915 | 0.043 | 21.224 | 0 | 21.27907 | 0 |
| LV_HIR.BY | HIR_CH | 0.831 | 0.028 | 29.284 | 0 | 29.67857 | 0 |
| LV_VEMEM.BY | VEMEM_CH | 0.74 | 0.043 | 17.304 | 0 | 17.2093 | 0 |
| LV_VEMEM.ON | LV_HIR | 0.117 | 0.098 | 1.191 | 0.234 | 1.193878 | 0.232526 |
| LV_VEMEM.ON | HIRXCHR | -0.11 | 0.067 | -1.635 | 0.102 | -1.64179 | 0.100633 |
| LV_VEMEM.ON | CHR | -0.052 | 0.049 | -1.057 | 0.291 | -1.06122 | 0.288588 |
| LV_VEMEM.ON | AGE | 0.028 | 0.061 | 0.466 | 0.641 | 0.459016 | 0.646222 |
| LV_VEMEM.ON | FEMALE | -0.094 | 0.061 | -1.529 | 0.126 | -1.54098 | 0.123321 |
| LV_VEMEM.ON | SITE02 | 0.034 | 0.087 | 0.396 | 0.692 | 0.390805 | 0.695942 |
| LV_VEMEM.ON | SITE03 | -0.012 | 0.07 | -0.17 | 0.865 | -0.17143 | 0.863887 |
| LV_VEMEM.ON | SITE04 | -0.189 | 0.093 | -2.022 | 0.043 | -2.03226 | 0.042128 |
| LV_VEMEM.ON | SITE05 | 0.041 | 0.079 | 0.523 | 0.601 | 0.518987 | 0.60377 |
| LV_VEMEM.ON | SITE06 | 0.112 | 0.089 | 1.265 | 0.206 | 1.258427 | 0.208237 |
| LV_VEMEM.ON | SITE07 | 0.078 | 0.077 | 1.02 | 0.308 | 1.012987 | 0.311066 |
| LV_VEMEM.ON | SITE08 | 0.056 | 0.087 | 0.64 | 0.522 | 0.643678 | 0.519784 |
| LV_VEMEM.ON | SITE09 | 0.135 | 0.093 | 1.454 | 0.146 | 1.451613 | 0.146609 |
| LV_HIR.WITH | CHR | 0.012 | 0.045 | 0.269 | 0.788 | 0.266667 | 0.789726 |
| LV_HIR.WITH | AGE | -0.023 | 0.055 | -0.424 | 0.672 | -0.41818 | 0.675814 |
| LV_HIR.WITH | FEMALE | -0.004 | 0.059 | -0.069 | 0.945 | -0.0678 | 0.945948 |
| LV_HIR.WITH | SITE02 | -0.015 | 0.064 | -0.237 | 0.812 | -0.23438 | 0.814694 |
| LV_HIR.WITH | SITE03 | -0.096 | 0.073 | -1.311 | 0.19 | -1.31507 | 0.188487 |
| LV_HIR.WITH | SITE04 | -0.037 | 0.059 | -0.616 | 0.538 | -0.62712 | 0.530581 |
| LV_HIR.WITH | SITE05 | 0.005 | 0.05 | 0.095 | 0.924 | 0.1 | 0.920344 |
| LV_HIR.WITH | SITE06 | 0.031 | 0.046 | 0.674 | 0.501 | 0.673913 | 0.500367 |
| LV_HIR.WITH | SITE07 | 0.07 | 0.07 | 1.005 | 0.315 | 1 | 0.317311 |
| LV_HIR.WITH | SITE08 | 0.033 | 0.054 | 0.612 | 0.541 | 0.611111 | 0.541126 |
| LV_HIR.WITH | SITE09 | -0.039 | 0.055 | -0.713 | 0.476 | -0.70909 | 0.478268 |
| CHR.WITH | AGE | -0.049 | 0.051 | -0.954 | 0.34 | -0.96078 | 0.336661 |
| CHR.WITH | FEMALE | -0.027 | 0.048 | -0.567 | 0.57 | -0.5625 | 0.573775 |
| CHR.WITH | SITE02 | -0.045 | 0.053 | -0.849 | 0.396 | -0.84906 | 0.39585 |
| CHR.WITH | SITE03 | -0.033 | 0.052 | -0.619 | 0.536 | -0.63462 | 0.525679 |
| CHR.WITH | SITE04 | 0.015 | 0.046 | 0.331 | 0.741 | 0.326087 | 0.744359 |
| CHR.WITH | SITE05 | -0.002 | 0.048 | -0.042 | 0.967 | -0.04167 | 0.966764 |
| CHR.WITH | SITE06 | -0.015 | 0.049 | -0.305 | 0.76 | -0.30612 | 0.759511 |
| CHR.WITH | SITE07 | -0.002 | 0.048 | -0.042 | 0.967 | -0.04167 | 0.966764 |
| CHR.WITH | SITE08 | 0.026 | 0.045 | 0.579 | 0.563 | 0.577778 | 0.563414 |
| CHR.WITH | SITE09 | 0.007 | 0.048 | 0.153 | 0.878 | 0.145833 | 0.884053 |
| AGE.WITH | FEMALE | -0.111 | 0.048 | -2.328 | 0.02 | -2.3125 | 0.02075 |
| AGE.WITH | SITE02 | 0.306 | 0.052 | 5.903 | 0 | 5.884615 | 3.99E-09 |
| AGE.WITH | SITE03 | 0 | 0.045 | 0 | 1 | 0 | 1 |
| AGE.WITH | SITE04 | -0.01 | 0.039 | -0.265 | 0.791 | -0.25641 | 0.797634 |
| AGE.WITH | SITE05 | 0.151 | 0.044 | 3.456 | 0.001 | 3.431818 | 0.0006 |
| AGE.WITH | SITE06 | -0.009 | 0.041 | -0.227 | 0.82 | -0.21951 | 0.826251 |
| AGE.WITH | SITE07 | -0.144 | 0.046 | -3.132 | 0.002 | -3.13043 | 0.001745 |
| AGE.WITH | SITE08 | -0.172 | 0.049 | -3.482 | 0 | -3.5102 | 0.000448 |
| AGE.WITH | SITE09 | -0.027 | 0.045 | -0.613 | 0.54 | -0.6 | 0.548506 |
| FEMALE.WITH | SITE02 | 0.026 | 0.048 | 0.54 | 0.589 | 0.541667 | 0.588048 |
| FEMALE.WITH | SITE03 | -0.018 | 0.048 | -0.373 | 0.709 | -0.375 | 0.70766 |
| FEMALE.WITH | SITE04 | 0.001 | 0.048 | 0.017 | 0.987 | 0.020833 | 0.983379 |
| FEMALE.WITH | SITE05 | 0.007 | 0.048 | 0.143 | 0.886 | 0.145833 | 0.884053 |
| FEMALE.WITH | SITE06 | -0.065 | 0.047 | -1.369 | 0.171 | -1.38298 | 0.166671 |
| FEMALE.WITH | SITE07 | 0.061 | 0.048 | 1.257 | 0.209 | 1.270833 | 0.203788 |
| FEMALE.WITH | SITE08 | -0.001 | 0.048 | -0.023 | 0.981 | -0.02083 | 0.983379 |
| FEMALE.WITH | SITE09 | 0.015 | 0.048 | 0.311 | 0.756 | 0.3125 | 0.754661 |
| SITE02.WITH | SITE03 | -0.079 | 0.01 | -7.883 | 0 | -7.9 | 2.89E-15 |
| SITE02.WITH | SITE04 | -0.096 | 0.011 | -8.699 | 0 | -8.72727 | 0 |
| SITE02.WITH | SITE05 | -0.125 | 0.013 | -9.724 | 0 | -9.61538 | 0 |
| SITE02.WITH | SITE06 | -0.12 | 0.013 | -9.586 | 0 | -9.23077 | 0 |
| SITE02.WITH | SITE07 | -0.125 | 0.013 | -9.724 | 0 | -9.61538 | 0 |
| SITE02.WITH | SITE08 | -0.108 | 0.012 | -9.177 | 0 | -9 | 0 |
| SITE02.WITH | SITE09 | -0.129 | 0.013 | -9.819 | 0 | -9.92308 | 0 |
| SITE03.WITH | SITE04 | -0.078 | 0.01 | -7.839 | 0 | -7.8 | 6.22E-15 |
| SITE03.WITH | SITE05 | -0.102 | 0.012 | -8.563 | 0 | -8.5 | 0 |
| SITE03.WITH | SITE06 | -0.098 | 0.012 | -8.468 | 0 | -8.16667 | 2.22E-16 |
| SITE03.WITH | SITE07 | -0.102 | 0.012 | -8.563 | 0 | -8.5 | 0 |
| SITE03.WITH | SITE08 | -0.088 | 0.011 | -8.183 | 0 | -8 | 1.33E-15 |
| SITE03.WITH | SITE09 | -0.105 | 0.012 | -8.628 | 0 | -8.75 | 0 |
| SITE04.WITH | SITE05 | -0.124 | 0.013 | -9.642 | 0 | -9.53846 | 0 |
| SITE04.WITH | SITE06 | -0.119 | 0.012 | -9.507 | 0 | -9.91667 | 0 |
| SITE04.WITH | SITE07 | -0.124 | 0.013 | -9.642 | 0 | -9.53846 | 0 |
| SITE04.WITH | SITE08 | -0.106 | 0.012 | -9.108 | 0 | -8.83333 | 0 |
| SITE04.WITH | SITE09 | -0.127 | 0.013 | -9.735 | 0 | -9.76923 | 0 |
| SITE05.WITH | SITE06 | -0.156 | 0.014 | -10.897 | 0 | -11.1429 | 0 |
| SITE05.WITH | SITE07 | -0.162 | 0.015 | -11.101 | 0 | -10.8 | 0 |
| SITE05.WITH | SITE08 | -0.139 | 0.014 | -10.306 | 0 | -9.92857 | 0 |
| SITE05.WITH | SITE09 | -0.167 | 0.015 | -11.245 | 0 | -11.1333 | 0 |
| SITE06.WITH | SITE07 | -0.156 | 0.014 | -10.897 | 0 | -11.1429 | 0 |
| SITE06.WITH | SITE08 | -0.134 | 0.013 | -10.142 | 0 | -10.3077 | 0 |
| SITE06.WITH | SITE09 | -0.16 | 0.015 | -11.033 | 0 | -10.6667 | 0 |
| SITE07.WITH | SITE08 | -0.139 | 0.014 | -10.306 | 0 | -9.92857 | 0 |
| SITE07.WITH | SITE09 | -0.167 | 0.015 | -11.245 | 0 | -11.1333 | 0 |
| SITE08.WITH | SITE09 | -0.143 | 0.014 | -10.42 | 0 | -10.2143 | 0 |
| Means | CHR | 2.124 | 0.133 | 15.981 | 0 | 15.96992 | 0 |
| Means | AGE | 4.789 | 0.152 | 31.606 | 0 | 31.50658 | 0 |
| Means | FEMALE | 0.92 | 0.044 | 20.664 | 0 | 20.90909 | 0 |
| Means | SITE02 | 0.311 | 0.026 | 11.771 | 0 | 11.96154 | 0 |
| Means | SITE03 | 0.254 | 0.026 | 9.885 | 0 | 9.769231 | 0 |
| Means | SITE04 | 0.307 | 0.026 | 11.63 | 0 | 11.80769 | 0 |
| Means | SITE05 | 0.403 | 0.028 | 14.371 | 0 | 14.39286 | 0 |
| Means | SITE06 | 0.387 | 0.028 | 13.958 | 0 | 13.82143 | 0 |
| Means | SITE07 | 0.403 | 0.028 | 14.371 | 0 | 14.39286 | 0 |
| Means | SITE08 | 0.346 | 0.027 | 12.819 | 0 | 12.81481 | 0 |
| Means | SITE09 | 0.414 | 0.028 | 14.666 | 0 | 14.78571 | 0 |
| Intercepts | VEMEM_CH | 0.292 | 0.297 | 0.984 | 0.325 | 0.983165 | 0.325526 |
| Intercepts | HIR_CH | 0.008 | 0.048 | 0.174 | 0.862 | 0.166667 | 0.867632 |
| Variances | CHR | 1 | 0 |  |  |  |  |
| Variances | AGE | 1 | 0 |  |  |  |  |
| Variances | FEMALE | 1 | 0 |  |  |  |  |
| Variances | SITE02 | 1 | 0 |  |  |  |  |
| Variances | SITE03 | 1 | 0 |  |  |  |  |
| Variances | SITE04 | 1 | 0 |  |  |  |  |
| Variances | SITE05 | 1 | 0 |  |  |  |  |
| Variances | SITE06 | 1 | 0 |  |  |  |  |
| Variances | SITE07 | 1 | 0 |  |  |  |  |
| Variances | SITE08 | 1 | 0 |  |  |  |  |
| Variances | SITE09 | 1 | 0 |  |  |  |  |
| Variances | LV_HIR | 1 | 0 |  |  |  |  |
| Residual.Variances | VEMEM_CH | 0.453 | 0.063 | 7.153 | 0 | 7.190476 | 6.46E-13 |
| Residual.Variances | HIR_CH | 0.31 | 0.047 | 6.567 | 0 | 6.595745 | 4.23E-11 |
| Residual.Variances | LV_VEMEM | 0.89 | 0.05 | 17.862 | 0 | 17.8 | 0 |

Table_S7b_SensAnalysisStand_WithinHippo_Corr

| **paramHeader** | **param** | **est** | **se** | **est_se** | **pval** | **zval** | **p_exact** | **p_bonf** |
| --- | --- | --- | --- | --- | --- | --- | --- | --- |
| LV_DEPS.ON | HILXCHR | 0.008 | 0.084 | 0.095 | 0.924 | 0.095238 | 0.924126 | 1 |
| LV_GAF.ON | HILXCHR | 0.108 | 0.053 | 2.02 | 0.043 | 2.037736 | 0.041576 | 0.249458 |
| LV_NEGS.ON | HILXCHR | -0.009 | 0.074 | -0.124 | 0.902 | -0.12162 | 0.903199 | 1 |
| LV_POSS.ON | HILXCHR | 0.059 | 0.063 | 0.935 | 0.35 | 0.936508 | 0.349012 | 1 |
| LV_SYCOD.ON | HILXCHR | -0.129 | 0.086 | -1.502 | 0.133 | -1.5 | 0.133614 | 0.801686 |
| LV_VEMEM.ON | HILXCHR | -0.016 | 0.07 | -0.231 | 0.818 | -0.22857 | 0.819202 | 1 |
| LV_DEPS.ON | HIRXCHR | -0.149 | 0.048 | -3.102 | 0.002 | -3.10417 | 0.001908 | 0.011449 |
| LV_GAF.ON | HIRXCHR | 0.143 | 0.053 | 2.712 | 0.007 | 2.698113 | 0.006973 | 0.04184 |
| LV_NEGS.ON | HIRXCHR | -0.099 | 0.053 | -1.86 | 0.063 | -1.86792 | 0.061773 | 0.370636 |
| LV_POSS.ON | HIRXCHR | -0.094 | 0.043 | -2.166 | 0.03 | -2.18605 | 0.028812 | 0.172873 |
| LV_SYCOD.ON | HIRXCHR | -0.025 | 0.082 | -0.299 | 0.765 | -0.30488 | 0.760459 | 1 |
| LV_VEMEM.ON | HIRXCHR | -0.11 | 0.067 | -1.635 | 0.102 | -1.64179 | 0.100633 | 0.6038 |
| LV_DEPS.ON | HIHXCHR | -0.151 | 0.063 | -2.406 | 0.016 | -2.39683 | 0.016538 | 0.099227 |
| LV_GAF.ON | HIHXCHR | 0.139 | 0.045 | 3.084 | 0.002 | 3.088889 | 0.002009 | 0.012054 |
| LV_NEGS.ON | HIHXCHR | -0.212 | 0.057 | -3.699 | 0 | -3.7193 | 0.0002 | 0.001199 |
| LV_POSS.ON | HIHXCHR | -0.035 | 0.074 | -0.468 | 0.64 | -0.47297 | 0.636232 | 1 |
| LV_SYCOD.ON | HIHXCHR | 0.007 | 0.091 | 0.082 | 0.934 | 0.076923 | 0.938685 | 1 |
| LV_VEMEM.ON | HIHXCHR | -0.101 | 0.053 | -1.893 | 0.058 | -1.90566 | 0.056694 | 0.340166 |

Table_S7c_ SensAnalysisStand_WithinHippo_ModelSum

| **behav** | **neuro** | **estimator** | **n_obs** | **n_parameters** | **aic** | **bic** |
| --- | --- | --- | --- | --- | --- | --- |
| PosS | fchil | MLR | 435 | 105 | 3989.984 | 4417.896 |
| PosS | fchir | MLR | 435 | 105 | 4086.572 | 4514.483 |
| PosS | fchih | MLR | 435 | 105 | 3905.365 | 4333.276 |
| NegS | fchil | MLR | 434 | 105 | 4312.848 | 4740.518 |
| NegS | fchir | MLR | 434 | 105 | 4397.846 | 4825.516 |
| NegS | fchih | MLR | 434 | 105 | 4176.379 | 4604.049 |
| DepS | fchil | MLR | 435 | 105 | 4173.512 | 4601.423 |
| DepS | fchir | MLR | 435 | 105 | 4249.12 | 4677.032 |
| DepS | fchih | MLR | 435 | 105 | 4068.487 | 4496.398 |
| gaf | fchil | MLR | 434 | 105 | 4974.971 | 5402.641 |
| gaf | fchir | MLR | 434 | 105 | 5061.725 | 5489.395 |
| gaf | fchih | MLR | 434 | 105 | 4894.277 | 5321.947 |
| SyCod | fchil | MLR | 430 | 105 | 4895.242 | 5321.94 |
| SyCod | fchir | MLR | 430 | 105 | 5002.866 | 5429.563 |
| SyCod | fchih | MLR | 430 | 105 | 4822.305 | 5249.002 |
| VeMem | fchil | MLR | 430 | 105 | 4060.16 | 4486.858 |
| VeMem | fchir | MLR | 430 | 105 | 4165.286 | 4591.984 |
| VeMem | fchih | MLR | 430 | 105 | 3979.749 | 4406.447 |

Table_S8_BrainnetomeRegionToNetworkAssigments

| **Name of Region/Network** | **Brainnetome Region Names** |
| --- | --- |
| Hippocampus | rHipp_L, rHipp_R, cHipp_L, cHipp_R |
| Right Hippocampus | rHipp_R, cHipp_R |
| Left Hippocampus | rHipp_L, cHipp_L |
| Homotopic Hippocampus | RHipp_L, rHipp_R, cHipp_L, cHipp_R (only connections across hemispheres considered) |
| Amygdala | mAmyg_L, mAmyg_R, lAmyg_L, lAmyg_R |
| Thalamus | mPFtha_L, mPFtha_R, mPMtha_L, mPMtha_R, Stha_L, Stha_R, rTtha_L, rTtha_R, PPtha_L, PPtha_R, Otha_L, Otha_R, cTtha_L, cTtha_R, lPFtha_L, lPFtha_R |
| Putamen | vmPu_L, vmPu_R, dlPu_L, dlPu_R |
| Pallidum | GP_L, GP_R |
| Accumbens | NAC_L, NAC_R |
| Caudate | vCa_L, vCa_R, dCa_L, dCa_R |
| Basal Ganglia | vCa_L, vCa_R, GP_L, GP_R, NAC_L, NAC_R, vmPu_L, vmPu_R, dCa_L, dCa_R, dlPu_L, dlPu_R |
| Salience Network | vIa_L, vIa_R, dIa_L, dIa_R, vIdvIg_L, vIdvIg_R, dIg_L, dIg_R, dId_L, dId_R, A24rv_L, A24rv_R, A32p_L, A32p_R, A24cd_L, A24cd_R |
| Default-mode Network | A9m_L, A9m_R, A10m_L, A10m_R, A37dl_L, A37dl_R, A37vl_L, A37vl_R, dmPOS_L, dmPOS_R, A31_L, A31_R, A23d_L, A23d_R |
| Fronto-parietal Network | A946d_L, A946d_R, A946v_L, A946v_R, A39rd_L, A39rd_R, A40rd_L, A40rd_R, A40c_L, A40c_R, A39rv_L, A39rv_R |
| Limbic Network | A23d_L, A23d_R, A24rv_L, A24rv_R, A32p_L, A32p_R, A23v_L, A23v_R, A24cd_L, A24cd_R, A23c_L, A23c_R, A32sg_L, A32sg_R, A2834_L, A2834_R, mAmyg_L, mAmyg_R, lAmyg_L, lAmyg_R, rHipp_L, rHipp_R, cHipp_L, cHipp_R, NAC_L, NAC_R |
| Somatosensory Network | A4hf_L, A4hf_R, A6cdl_L, A6cdl_R, A4ul_L, A4ul_R, A4t_L, A4t_R, A4tl_L, A4tl_R, A6cvl_L, A6cvl_R, A123ll_L, A123ll_R, A4ll_L, A4ll_R, A123ulhf_L, A123ulhf_R, A123tonIa_L, A123tonIa_R, A2_L, A2_R, A123tru_L, A123tru_R |
| Visual Network | cLinG_L, cLinG_R, rCunG_L, rCunG_R, cCunG_L, cCunG_R, rLinG_L, rLinG_R, vmPOS_L, vmPOS_R, mOccG_L, mOccG_R, V5MT_L, V5MT_R, OPC_L, OPC_R, iOccG_L, iOccG_R, msOccG_L, msOccG_R, lsOccG_L, lsOccG_R |

Table_S9a_SensAnalysisStand_OtherNetw_Corr

| **paramHeader** | **param** | **est** | **se** | **est_se** | **pval** | **zval** | **pval_exact** | **p_bonf** |
| --- | --- | --- | --- | --- | --- | --- | --- | --- |
| LV_DEPS.ON | SALXCHR | -0.016 | 0.041 | -0.39 | 0.697 | -0.39024 | 0.696356 | 1 |
| LV_GAF.ON | SALXCHR | 0.034 | 0.041 | 0.809 | 0.419 | 0.829268 | 0.406953 | 1 |
| LV_NEGS.ON | SALXCHR | -0.022 | 0.049 | -0.454 | 0.65 | -0.44898 | 0.653446 | 1 |
| LV_POSS.ON | SALXCHR | 0.013 | 0.037 | 0.361 | 0.718 | 0.351351 | 0.725325 | 1 |
| LV_SYCOD.ON | SALXCHR | 0.168 | 0.09 | 1.872 | 0.061 | 1.866667 | 0.061948 | 0.371689 |
| LV_VEMEM.ON | SALXCHR | -0.051 | 0.066 | -0.772 | 0.44 | -0.77273 | 0.439684 | 1 |
| LV_DEPS.ON | DMNXCHR | 0.012 | 0.039 | 0.295 | 0.768 | 0.307692 | 0.758316 | 1 |
| LV_GAF.ON | DMNXCHR | 0.008 | 0.044 | 0.181 | 0.856 | 0.181818 | 0.855725 | 1 |
| LV_NEGS.ON | DMNXCHR | 0.034 | 0.042 | 0.806 | 0.42 | 0.809524 | 0.418214 | 1 |
| LV_POSS.ON | DMNXCHR | 0.027 | 0.031 | 0.86 | 0.39 | 0.870968 | 0.383772 | 1 |
| LV_SYCOD.ON | DMNXCHR | -0.051 | 0.067 | -0.762 | 0.446 | -0.76119 | 0.446541 | 1 |
| LV_VEMEM.ON | DMNXCHR | -0.052 | 0.057 | -0.909 | 0.363 | -0.91228 | 0.361621 | 1 |
| LV_DEPS.ON | FPNXCHR | -0.033 | 0.051 | -0.645 | 0.519 | -0.64706 | 0.517594 | 1 |
| LV_GAF.ON | FPNXCHR | 0.03 | 0.05 | 0.588 | 0.557 | 0.6 | 0.548506 | 1 |
| LV_NEGS.ON | FPNXCHR | 0.025 | 0.065 | 0.376 | 0.707 | 0.384615 | 0.700522 | 1 |
| LV_POSS.ON | FPNXCHR | -0.039 | 0.054 | -0.733 | 0.463 | -0.72222 | 0.470158 | 1 |
| LV_SYCOD.ON | FPNXCHR | -0.125 | 0.102 | -1.225 | 0.22 | -1.22549 | 0.220391 | 1 |
| LV_VEMEM.ON | FPNXCHR | -0.112 | 0.065 | -1.715 | 0.086 | -1.72308 | 0.084875 | 0.509248 |
| LV_DEPS.ON | LIMXCHR | 0.025 | 0.048 | 0.523 | 0.601 | 0.520833 | 0.602483 | 1 |
| LV_GAF.ON | LIMXCHR | -0.038 | 0.049 | -0.768 | 0.442 | -0.77551 | 0.438038 | 1 |
| LV_NEGS.ON | LIMXCHR | 0.09 | 0.054 | 1.665 | 0.096 | 1.666667 | 0.095581 | 0.573484 |
| LV_POSS.ON | LIMXCHR | 0.049 | 0.037 | 1.318 | 0.187 | 1.324324 | 0.185395 | 1 |
| LV_SYCOD.ON | LIMXCHR | -0.009 | 0.068 | -0.133 | 0.894 | -0.13235 | 0.894705 | 1 |
| LV_VEMEM.ON | LIMXCHR | 0.053 | 0.066 | 0.804 | 0.421 | 0.80303 | 0.421957 | 1 |
| LV_DEPS.ON | VISXCHR | 0.028 | 0.059 | 0.473 | 0.636 | 0.474576 | 0.635089 | 1 |
| LV_GAF.ON | VISXCHR | 0.022 | 0.038 | 0.579 | 0.562 | 0.578947 | 0.562625 | 1 |
| LV_NEGS.ON | VISXCHR | -0.007 | 0.055 | -0.131 | 0.896 | -0.12727 | 0.898725 | 1 |
| LV_POSS.ON | VISXCHR | 0.088 | 0.044 | 1.998 | 0.046 | 2 | 0.0455 | 0.273002 |
| LV_SYCOD.ON | VISXCHR | 0.045 | 0.058 | 0.781 | 0.435 | 0.775862 | 0.43783 | 1 |
| LV_VEMEM.ON | VISXCHR | -0.079 | 0.068 | -1.177 | 0.239 | -1.16176 | 0.245331 | 1 |
| LV_DEPS.ON | SMNXCHR | 0.021 | 0.038 | 0.558 | 0.577 | 0.552632 | 0.580516 | 1 |
| LV_GAF.ON | SMNXCHR | -0.05 | 0.057 | -0.887 | 0.375 | -0.87719 | 0.380382 | 1 |
| LV_NEGS.ON | SMNXCHR | -0.019 | 0.033 | -0.572 | 0.568 | -0.57576 | 0.564779 | 1 |
| LV_POSS.ON | SMNXCHR | -0.037 | 0.04 | -0.916 | 0.359 | -0.925 | 0.354966 | 1 |
| LV_SYCOD.ON | SMNXCHR | 0.05 | 0.058 | 0.847 | 0.397 | 0.862069 | 0.38865 | 1 |
| LV_VEMEM.ON | SMNXCHR | 0.056 | 0.057 | 0.993 | 0.321 | 0.982456 | 0.325875 | 1 |
| LV_DEPS.ON | AMYXCHR | -0.094 | 0.047 | -2.012 | 0.044 | -2 | 0.0455 | 0.273002 |
| LV_GAF.ON | AMYXCHR | 0.096 | 0.057 | 1.705 | 0.088 | 1.684211 | 0.092141 | 0.552846 |
| LV_NEGS.ON | AMYXCHR | -0.096 | 0.046 | -2.072 | 0.038 | -2.08696 | 0.036892 | 0.221352 |
| LV_POSS.ON | AMYXCHR | -0.071 | 0.046 | -1.542 | 0.123 | -1.54348 | 0.122715 | 0.736289 |
| LV_SYCOD.ON | AMYXCHR | -0.118 | 0.088 | -1.343 | 0.179 | -1.34091 | 0.17995 | 1 |
| LV_VEMEM.ON | AMYXCHR | 0.152 | 0.067 | 2.265 | 0.023 | 2.268657 | 0.023289 | 0.139735 |
| LV_DEPS.ON | BGXCHR | 0.077 | 0.048 | 1.607 | 0.108 | 1.604167 | 0.108677 | 0.652064 |
| LV_GAF.ON | BGXCHR | -0.01 | 0.052 | -0.19 | 0.85 | -0.19231 | 0.847501 | 1 |
| LV_NEGS.ON | BGXCHR | -0.058 | 0.052 | -1.12 | 0.263 | -1.11538 | 0.264686 | 1 |
| LV_POSS.ON | BGXCHR | 0.035 | 0.036 | 0.966 | 0.334 | 0.972222 | 0.33094 | 1 |
| LV_SYCOD.ON | BGXCHR | 0.054 | 0.069 | 0.782 | 0.434 | 0.782609 | 0.433857 | 1 |
| LV_VEMEM.ON | BGXCHR | 0.004 | 0.047 | 0.087 | 0.931 | 0.085106 | 0.932177 | 1 |
| LV_DEPS.ON | THAXCHR | -0.025 | 0.039 | -0.629 | 0.529 | -0.64103 | 0.521506 | 1 |
| LV_GAF.ON | THAXCHR | 0.028 | 0.044 | 0.642 | 0.521 | 0.636364 | 0.524539 | 1 |
| LV_NEGS.ON | THAXCHR | -0.029 | 0.04 | -0.704 | 0.481 | -0.725 | 0.468452 | 1 |
| LV_POSS.ON | THAXCHR | -0.017 | 0.029 | -0.593 | 0.553 | -0.58621 | 0.557736 | 1 |
| LV_SYCOD.ON | THAXCHR | -0.027 | 0.083 | -0.321 | 0.748 | -0.3253 | 0.744953 | 1 |
| LV_VEMEM.ON | THAXCHR | 0.015 | 0.066 | 0.225 | 0.822 | 0.227273 | 0.820212 | 1 |
| LV_DEPS.ON | PALXCHR | 0.037 | 0.037 | 0.999 | 0.318 | 1 | 0.317311 | 1 |
| LV_GAF.ON | PALXCHR | -0.042 | 0.057 | -0.745 | 0.456 | -0.73684 | 0.461218 | 1 |
| LV_NEGS.ON | PALXCHR | -0.014 | 0.039 | -0.368 | 0.713 | -0.35897 | 0.719614 | 1 |
| LV_POSS.ON | PALXCHR | 0.069 | 0.034 | 2.044 | 0.041 | 2.029412 | 0.042416 | 0.254498 |
| LV_SYCOD.ON | PALXCHR | -0.041 | 0.049 | -0.849 | 0.396 | -0.83673 | 0.402742 | 1 |
| LV_VEMEM.ON | PALXCHR | -0.028 | 0.064 | -0.436 | 0.663 | -0.4375 | 0.661749 | 1 |
| LV_DEPS.ON | PUTXCHR | 0.029 | 0.034 | 0.846 | 0.397 | 0.852941 | 0.393692 | 1 |
| LV_GAF.ON | PUTXCHR | 0.002 | 0.054 | 0.04 | 0.968 | 0.037037 | 0.970455 | 1 |
| LV_NEGS.ON | PUTXCHR | -0.069 | 0.039 | -1.779 | 0.075 | -1.76923 | 0.076855 | 0.461132 |
| LV_POSS.ON | PUTXCHR | 0.027 | 0.035 | 0.758 | 0.449 | 0.771429 | 0.440453 | 1 |
| LV_SYCOD.ON | PUTXCHR | 0.016 | 0.075 | 0.214 | 0.83 | 0.213333 | 0.831067 | 1 |
| LV_VEMEM.ON | PUTXCHR | -0.011 | 0.062 | -0.176 | 0.86 | -0.17742 | 0.859179 | 1 |
| LV_DEPS.ON | CAUXCHR | 0.115 | 0.055 | 2.086 | 0.037 | 2.090909 | 0.036536 | 0.219217 |
| LV_GAF.ON | CAUXCHR | -0.043 | 0.047 | -0.929 | 0.353 | -0.91489 | 0.360247 | 1 |
| LV_NEGS.ON | CAUXCHR | -0.024 | 0.057 | -0.419 | 0.675 | -0.42105 | 0.673717 | 1 |
| LV_POSS.ON | CAUXCHR | 0.034 | 0.047 | 0.723 | 0.47 | 0.723404 | 0.469432 | 1 |
| LV_SYCOD.ON | CAUXCHR | 0.077 | 0.075 | 1.033 | 0.302 | 1.026667 | 0.304577 | 1 |
| LV_VEMEM.ON | CAUXCHR | -0.002 | 0.06 | -0.034 | 0.973 | -0.03333 | 0.973409 | 1 |
| LV_DEPS.ON | ACUXCHR | -0.025 | 0.063 | -0.393 | 0.694 | -0.39683 | 0.691496 | 1 |
| LV_GAF.ON | ACUXCHR | 0.042 | 0.052 | 0.806 | 0.42 | 0.807692 | 0.419268 | 1 |
| LV_NEGS.ON | ACUXCHR | -0.064 | 0.064 | -1 | 0.317 | -1 | 0.317311 | 1 |
| LV_POSS.ON | ACUXCHR | -0.043 | 0.058 | -0.749 | 0.454 | -0.74138 | 0.458463 | 1 |
| LV_SYCOD.ON | ACUXCHR | -0.12 | 0.063 | -1.901 | 0.057 | -1.90476 | 0.056811 | 0.340866 |
| LV_VEMEM.ON | ACUXCHR | 0.052 | 0.074 | 0.699 | 0.485 | 0.702703 | 0.482241 | 1 |

Table_S9b_SensAnalysisStand_OtherNetw_ModelSum

| **behav** | **neuro** | **estimator** | **n_obs** | **n_parameters** | **aic** | **bic** |
| --- | --- | --- | --- | --- | --- | --- |
| PosS | fcsal | MLR | 435 | 105 | 3674.297 | 4102.208 |
| PosS | fcdmn | MLR | 435 | 105 | 3041.187 | 3469.098 |
| PosS | fcfpn | MLR | 435 | 105 | 3284.113 | 3712.024 |
| PosS | fclim | MLR | 435 | 105 | 3121.546 | 3549.457 |
| PosS | fcvis | MLR | 435 | 105 | 3733.054 | 4160.965 |
| PosS | fcsmn | MLR | 435 | 105 | 3588.591 | 4016.503 |
| PosS | fcamy | MLR | 435 | 105 | 3904.969 | 4332.88 |
| PosS | fcbg | MLR | 435 | 105 | 3822.299 | 4250.21 |
| PosS | fctha | MLR | 435 | 105 | 3862.13 | 4290.041 |
| PosS | fcpal | MLR | 435 | 105 | 4209.723 | 4637.634 |
| PosS | fcput | MLR | 435 | 105 | 3935.242 | 4363.153 |
| PosS | fccau | MLR | 435 | 105 | 3818.185 | 4246.096 |
| PosS | fcacu | MLR | 435 | 105 | 3973.475 | 4401.387 |
| NegS | fcsal | MLR | 434 | 105 | 3992.899 | 4420.569 |
| NegS | fcdmn | MLR | 434 | 105 | 3362.385 | 3790.055 |
| NegS | fcfpn | MLR | 434 | 105 | 3603.375 | 4031.044 |
| NegS | fclim | MLR | 434 | 105 | 3433.905 | 3861.575 |
| NegS | fcvis | MLR | 434 | 105 | 4047.923 | 4475.592 |
| NegS | fcsmn | MLR | 434 | 105 | 3905.623 | 4333.293 |
| NegS | fcamy | MLR | 434 | 105 | 4219.805 | 4647.474 |
| NegS | fcbg | MLR | 434 | 105 | 4141.573 | 4569.243 |
| NegS | fctha | MLR | 434 | 105 | 4180.245 | 4607.914 |
| NegS | fcpal | MLR | 434 | 105 | 4532.106 | 4959.776 |
| NegS | fcput | MLR | 434 | 105 | 4253.936 | 4681.605 |
| NegS | fccau | MLR | 434 | 105 | 4134.757 | 4562.427 |
| NegS | fcacu | MLR | 434 | 105 | 4294.298 | 4721.967 |
| DepS | fcsal | MLR | 435 | 105 | 3854.45 | 4282.361 |
| DepS | fcdmn | MLR | 435 | 105 | 3222.383 | 3650.294 |
| DepS | fcfpn | MLR | 435 | 105 | 3463.527 | 3891.438 |
| DepS | fclim | MLR | 435 | 105 | 3305.275 | 3733.186 |
| DepS | fcvis | MLR | 435 | 105 | 3918.419 | 4346.33 |
| DepS | fcsmn | MLR | 435 | 105 | 3771.324 | 4199.235 |
| DepS | fcamy | MLR | 435 | 105 | 4083.984 | 4511.895 |
| DepS | fcbg | MLR | 435 | 105 | 3993.484 | 4421.395 |
| DepS | fctha | MLR | 435 | 105 | 4042.094 | 4470.006 |
| DepS | fcpal | MLR | 435 | 105 | 4391.795 | 4819.706 |
| DepS | fcput | MLR | 435 | 105 | 4114.639 | 4542.551 |
| DepS | fccau | MLR | 435 | 105 | 3984.184 | 4412.095 |
| DepS | fcacu | MLR | 435 | 105 | 4160.908 | 4588.82 |
| gaf | fcsal | MLR | 434 | 105 | 4670.491 | 5098.161 |
| gaf | fcdmn | MLR | 434 | 105 | 4042.577 | 4470.247 |
| gaf | fcfpn | MLR | 434 | 105 | 4285.163 | 4712.833 |
| gaf | fclim | MLR | 434 | 105 | 4127.987 | 4555.656 |
| gaf | fcvis | MLR | 434 | 105 | 4738.544 | 5166.213 |
| gaf | fcsmn | MLR | 434 | 105 | 4586.708 | 5014.378 |
| gaf | fcamy | MLR | 434 | 105 | 4902.409 | 5330.078 |
| gaf | fcbg | MLR | 434 | 105 | 4818.481 | 5246.151 |
| gaf | fctha | MLR | 434 | 105 | 4861.8 | 5289.47 |
| gaf | fcpal | MLR | 434 | 105 | 5208.915 | 5636.585 |
| gaf | fcput | MLR | 434 | 105 | 4933.959 | 5361.629 |
| gaf | fccau | MLR | 434 | 105 | 4813.056 | 5240.726 |
| gaf | fcacu | MLR | 434 | 105 | 4978.306 | 5405.976 |
| SyCod | fcsal | MLR | 430 | 105 | 4576.989 | 5003.687 |
| SyCod | fcdmn | MLR | 430 | 105 | 3956.73 | 4383.428 |
| SyCod | fcfpn | MLR | 430 | 105 | 4192.622 | 4619.32 |
| SyCod | fclim | MLR | 430 | 105 | 4042.197 | 4468.895 |
| SyCod | fcvis | MLR | 430 | 105 | 4645.457 | 5072.155 |
| SyCod | fcsmn | MLR | 430 | 105 | 4498.774 | 4925.471 |
| SyCod | fcamy | MLR | 430 | 105 | 4801.968 | 5228.666 |
| SyCod | fcbg | MLR | 430 | 105 | 4714.774 | 5141.471 |
| SyCod | fctha | MLR | 430 | 105 | 4768.756 | 5195.453 |
| SyCod | fcpal | MLR | 430 | 105 | 5114.007 | 5540.705 |
| SyCod | fcput | MLR | 430 | 105 | 4839.844 | 5266.541 |
| SyCod | fccau | MLR | 430 | 105 | 4713.288 | 5139.986 |
| SyCod | fcacu | MLR | 430 | 105 | 4887.43 | 5314.128 |
| VeMem | fcsal | MLR | 430 | 105 | 3740.9 | 4167.598 |
| VeMem | fcdmn | MLR | 430 | 105 | 3118.477 | 3545.175 |
| VeMem | fcfpn | MLR | 430 | 105 | 3353.818 | 3780.515 |
| VeMem | fclim | MLR | 430 | 105 | 3202.796 | 3629.494 |
| VeMem | fcvis | MLR | 430 | 105 | 3807.052 | 4233.749 |
| VeMem | fcsmn | MLR | 430 | 105 | 3659.424 | 4086.122 |
| VeMem | fcamy | MLR | 430 | 105 | 3956.516 | 4383.214 |
| VeMem | fcbg | MLR | 430 | 105 | 3876.397 | 4303.094 |
| VeMem | fctha | MLR | 430 | 105 | 3924.742 | 4351.439 |
| VeMem | fcpal | MLR | 430 | 105 | 4275.093 | 4701.79 |
| VeMem | fcput | MLR | 430 | 105 | 4003.784 | 4430.482 |
| VeMem | fccau | MLR | 430 | 105 | 3875.004 | 4301.701 |
| VeMem | fcacu | MLR | 430 | 105 | 4042.814 | 4469.511 |
